# A North American Collaborative Atlas of Oncology Data Visualization with R Statistical Software

**DOI:** 10.64898/2026.03.20.26348936

**Authors:** Mohsen Soltanifar, Andrew Jay Portuguese, Yein Jeon, Jordan Gauthier, Chel Hee Lee

## Abstract

Oncology research and clinical practice in North America increasingly rely on complex endpoints, heterogeneous study designs, and high-dimensional molecular data. In this landscape, data visualization serves as a critical analytic instrument for study design communication, model diagnostics, safety reporting, and real-time clinical decision support. Despite its importance, the oncology visualization ecosystem remains fragmented across commercial platforms and bespoke scripts, lacking a unified, code-first reference that emphasizes reproducibility and auditability in the R programming environment. This paper addresses this gap by presenting a North American collaborative atlas of 62 oncology visualization templates: 24 for clinical trials, 12 for real-world evidence (RWE), and 26 common to both settings. A core innovation of this atlas is its simulation-driven approach; each plot is illustrated using transparent, reproducible data-generating mechanisms. This allows users to deterministically recreate figures and easily adapt templates to alternative endpoints, censoring patterns, and subgroup structures. The paper provides foundational notation for oncology endpoints, an operational taxonomy based on data geometry, and a consolidated review of relevant R software. We further synthesize the practical utility of these methods through four representative case studies and provide a comparative analysis of the strengths, limitations, and future challenges of oncology data visualization. A detailed tutorial on fishplot is included to demonstrate a publication-ready workflow for clonal evolution.

---

> “If the medical world is to truly understand cancer, we have to work as a team with as many collaborators as we can find. Only in this way will we dissect the many different aspects of this disease and be able to devise technologies to defeat it.”—

> Tak Wah Mak, OC, OOnt, FRS, FRSC (2023)

## 1 Introduction

### 1.1 Background

Cancer remains a defining challenge for North American public health, driving urgent innovation in prevention, diagnosis, and multi-modality treatment. The modern oncology enterprise—from early biomarker-driven detection to complex survivorship care—is increasingly data-intensive and multidisciplinary, requiring clinicians and researchers to navigate high-dimensional datasets with precision [1, 2].

The population burden in North America is substantial and persistent. In 2024 alone, the United States had projected approximately 2,001,140 new cases and 611,720 deaths [3], while Canada had estimated 247,100 new cases and 88,100 deaths [4]. Globally, cancer remains a leading cause of mortality, reinforcing the need for robust analytic tools that can effectively communicate oncology evidence to guide national policy and clinical practice [5].

In this context, data visualization serves as a critical analytic instrument rather than a mere illustration. Effective visualizations facilitate cancer prevention by tracking incidence trends, aid diagnosis through molecular profiling, and optimize treatment by making response dynamics and toxicity heterogeneity interpretable. As oncology datasets grow in complexity, visualization methods must evolve to represent longitudinal trajectories and biological structures in ways that support real-time clinical decision-making [6].

### 1.2 Motivation and Scope

Despite its importance, the oncology visualization landscape remains fragmented across commercial platforms and bespoke scripts, often lacking standardization or reproducibility. There is currently no standalone, code-first atlas that consolidates these methods into a unified workflow. This gap leaves analysts without a biblical reference for R-based visualization that emphasizes transparency and auditability in reporting [7].

This paper addresses this gap by presenting a collaborative atlas of 62 oncology visualization templates in R: 24 for clinical trials (CT), 12 for real-world evidence (RWE), and 26 common to both settings. A core innovation is our simulation-driven approach, allowing readers to reproduce figures deterministically and adapt templates to their own endpoints and censoring patterns. This controlled pedagogical strategy ensures that the underlying statistical assumptions of each plot remain transparent and adaptable [8, 9].

### 1.3 Survey Organization

The paper is organized as follows. Section 2 establishes core notation and endpoint definitions. Section 3 introduces a pragmatic taxonomy classifying plots by study setting and analytic objective. Section 4 reviews software tools and R packages relevant to oncology. Section 5 constitutes the main Atlas of Plots, providing specifications and code. Section 6 offers a detailed tutorial on fishplot visualizations for clonal evolution. Finally, Section 7 discusses the strengths, limitations, and future challenges of oncology data visualization.

## 2 Preliminaries

Readers who have studied the key topics of oncology and software programming are well-equipped with the following notations, definitions, and results. Our intent in this section is to establish a common technical language—both statistical and clinical—so that the plot specifications in later sections can be read as precise objects (data → assumptions → computation → interpretation), without repeatedly reintroducing foundational material. In keeping with the pedagogical design of this atlas, we work primarily with simulated datasets in R to illustrate each visualization under controlled, modifiable conditions [7–9].

### 2.1 Observational unit, indexing, and data schema

Unless otherwise stated, the primary observational unit is the patient. We index patients by *i* ∈ {1*,…, n*}. A generic analysis dataset can be represented as a patient-level table with:

- **Identifier:** a unique patient identifier id*_i_*.
- **Baseline covariates:** a vector *X_i_* = (*X_i_*_1_*,…, X_ip_*) capturing demographic, clinical, tumor, and treatment-history information at (or prior to) index time.
- **Exposure/treatment:** a treatment indicator *A_i_* (binary, multinomial, or continuous dose), and, when needed, time-varying treatment *A_i_*(*t*).
- **Outcome(s):** binary *Y_i_* ∈ {0, 1}, continuous *Y_i_*∈ R, count/rate *Y_i_*∈ {0, 1, 2*,…* }, ordinal *Y_i_* ∈ {1*,…, K*}, time-to-event (*T_i_,* Δ*_i_*), or longitudinal trajectories 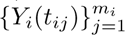.

While the patient is the default unit, certain visualizations in this atlas (e.g., meta-analysis forest plots or site-level recruitment charts) shift the indexing to the study or center level *s* ∈ {1*,…, S*}, utilizing aggregated summaries (*θ_s_, σ_s_*).

### 2.2 Endpoint families and core clinical definitions

We adopt endpoint language commonly used in oncology biostatistics [1, 2].

#### Binary endpoints

Binary outcomes typically represent response or toxicity indicators. Examples include objective response rate (ORR), disease control rate (DCR), and dose-limiting toxicity (DLT). We write:

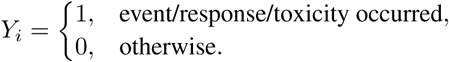

#### Continuous endpoints

Continuous endpoints include tumor burden measures (e.g., sum of lesion diameters, SLD), biomarker values, and patient-reported outcomes (PROs). We denote a continuous endpoint by *Y_i_*∈ R.

#### Counts and rates

Commonly used for safety reporting, these endpoints represent the frequency of events, such as the number of Adverse Events (AEs). For a follow-up duration *τ_i_*, the observed rate is *R_i_*= *Y_i_/τ_i_*.

#### Ordinal and composite outcomes

Ordinal outcomes capture ordered categories, such as RECIST 1.1 response: Complete Response (CR) *>* Partial Response (PR) *>* Stable Disease (SD) *>* Progressive Disease (PD). Composite outcomes may combine these states with event times to define endpoints like time to best response.

#### Time-to-event (TTE) endpoints

TTE endpoints include overall survival (OS), progression-free survival (PFS), and recurrence-free survival (RFS). Let *T_i_* ≥ 0 be the event time and *C_i_* ≥ 0 the censoring time. The observed time is:

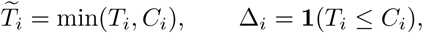

where Δ*_i_*= 1 indicates an observed event and Δ*_i_* = 0 indicates right censoring.

#### Competing risks and multi-state endpoints

When multiple event types can occur and preclude one another (e.g., relapse vs. non-relapse mortality), we denote the event type by *K_i_* ∈ {1*,…, K*} and use the *cumulative incidence function* (CIF):

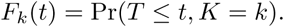

#### Longitudinal endpoints

Longitudinal outcomes (e.g., repeated tumor assessments) are observed at visit times 0 ≤ *t_i_*_1_ *<* · · · *< t_im__i_*, producing trajectories {*Y_i_*(*t_ij_*)}.

### 2.3 Phase I specific quantities

Because the atlas begins with dose-finding graphics, we define the **Maximum Tolerated Dose (MTD)** as the dose level with a true toxicity probability closest to a pre-specified **target toxicity** *θ* (typically 0.20–0.33). Performance is often visualized through the probability of escalation and the recommended dose distribution [2].

### 2.4 Effect measures and uncertainty

#### Contrasts

For two groups *g* ∈ {0, 1} with risks *p*_0_*, p*_1_, common contrasts include the risk difference (*p*_1_ − *p*_0_) and the odds ratio. For TTE, the primary contrast is the *hazard ratio* (HR), often reported as *β* = log(HR). When proportional hazards (PH) are questionable, restricted mean survival time (RMST) is preferred [10].

#### Uncertainty representation

Uncertainty is communicated through standard errors (SE), confidence intervals (CIs), or Bayesian credible intervals. We emphasize that a visually compelling plot is not an inferential guarantee; the credibility of uncertainty overlays depends on whether underlying assumptions (e.g., independent censoring, exchangeability) are plausible.

### 2.5 Observational (RWE) causal notation and diagnostics

For observational comparative effectiveness, we distinguish between the observed outcome *Y_i_* and potential outcomes (*Y_i_*(1)*, Y_i_*(0)). Key concepts include:

- **Propensity score (PS):** *e*(*X*) = Pr(*A* = 1 | *X*), used for matching or inverse probability of treatment weighting (IPTW).
- **Covariate balance:** Post-adjustment balance is assessed via standardized mean differences (SMD), which are displayed as balance plots to diagnose the adequacy of the causal design.

### 2.6 Prediction, calibration, and decision-analytic quantities

#### Discrimination and Calibration

Discrimination (AUC/ROC) summarizes ranking performance. Calibration describes the agreement between predicted risks *p^_i_* and observed event frequencies, typically visualized via calibration curves comparing binned rates [11].

#### Decision curve analysis (DCA)

DCA evaluates clinical utility by plotting *net benefit* as a function of the decision threshold probability, comparing a model-guided strategy to “treat all” and “treat none” benchmarks [12].

### 2.7 Simulation and reproducibility conventions

The atlas prioritizes *reproducible, scriptable* figure generation in R.

- **Simulation as controlled illustration.** Figures are generated from a data-generating mechanism (DGM) designed to reproduce canonical qualitative patterns (e.g., non-PH hazards) rather than specific real datasets.
- **Deterministic regeneration.** We fix random seeds and record package versions to support exact figure regeneration.
- **Modular Design.** We maintain a strict separation between the DGM layer, the data processing layer, and the plotting layer.

## 3 Taxonomy

To provide the reader with a coherent and systematically organized roadmap for the material assembled in this atlas, we adopt a pragmatic taxonomy of oncology data visualizations that is driven by (i) study setting and scientific objective,

**(1)** (ii) the geometry and provenance of the underlying data, and (iii) the inferential contract (assumptions, estimands, and uncertainty representation). This taxonomy is intentionally operational: it is designed to help analysts select an appropriate plot family for a given oncology question and to standardize plot specifications for reproducible reporting. In keeping with the pedagogical goals of this paper, we rely on simulation-based data generation to illustrate each plot under controlled and modifiable conditions [8, 9].
1. **Software (implementation layer).** We distinguish between general statistical environments and visualization-centric tooling. The atlas is implemented in R [7] and prioritizes scriptable, auditable workflows that support version control, reproducibility, and peer-review transparency.
2. **Apps (interactive layer).** We treat interactive applications (e.g., Shiny dashboards) as a complementary layer for exploratory analysis, cohort interrogation, and stakeholder communication. Nevertheless, the principal objective of the atlas is to deliver manuscript-ready figures produced directly from R code, thereby preserving provenance and enabling deterministic regeneration.
3. **Visualization object (plot-as-a-specification).** Each plot is treated as a fully specified object defined by: (i) *Purpose* (the question answered), (ii) *Required data variables* (minimum sufficient schema), (iii) *Assumptions* (statistical/structural), (iv) *Implementation* (packages/functions), and (v) *Interpretation and use cases*. This specification-first approach supports consistent comparison across plot families and study settings.
4. **Study setting (primary axis: Clinical Trials vs RWE vs Common).** We classify plots by the setting in which they most commonly arise: (i) *Clinical Trials* (design, monitoring, interventional efficacy/safety reporting), (ii) *Real World Evidence (RWE)* (observational cohorts, confounding control, treatment patterns, external comparators), and (iii) *Common* (visualizations routinely used in both settings, such as general survival summaries and evidence synthesis).
5. **Trial phase and decision point (design/monitoring axis).** Within clinical trials, we further index plots by phase (I–IV) and decision point (design-stage planning, interim monitoring, final reporting). This axis naturally separates early-phase dose-finding graphics from confirmatory efficacy displays and post-marketing safety summaries.
6. **Analytical objective (question-first indexing).** Independently of study setting, plots are indexed by their dominant objective: (i) design and operating characteristics, (ii) descriptive cohort characterization, (iii) comparative effectiveness and safety, (iv) prediction and risk stratification, (v) biomarker exploration and subgroup discovery, (vi) decision support and health economics. This objective-first view is often the most practical entry point for analysts who begin from an estimand or clinical question.
7. **Endpoint family (clinical outcome axis).** We classify plot families according to the endpoint structure: (i) binary outcomes (response, toxicity), (ii) continuous outcomes (tumor burden, biomarker values), (iii) counts and rates (AEs, healthcare utilization), (iv) time-to-event outcomes (OS/PFS/TTP), (v) competing risks and multi-state endpoints, (vi) longitudinal outcomes (QoL, repeated tumor assessments), (vii) composite/ordinal outcomes (RECIST categories, severity grades). Endpoint family determines both suitable geometry and required assumptions (e.g., censoring mechanisms).
8. **Data geometry (structure-first indexing).** We organize plots by the geometry of the underlying data: (i) distributions and uncertainty intervals, (ii) time-indexed curves, (iii) multivariate profiles and high-dimensional heatmaps, (iv) networks/flows (patient journeys, treatment sequences), (v) spatial/geographic variation, (vi) study-level aggregation (meta-analytic summaries). This structure-first index is particularly useful when the analyst inherits a dataset and must select a compatible plot class.
9. **Unit of analysis (granularity axis).** We differentiate plots whose primary unit is: (i) patient-level (waterfall, swimmer, spider), (ii) visit/episode-level (dose modifications over time, longitudinal lab trends), (iii) subgroup-level (subgroup effect displays, stratified summaries), (iv) site/center-level (heterogeneity across institutions), (v) study-level (forest plots, evidence synthesis). Granularity affects visual density, labeling strategy, and interpretability constraints.
10. **Comparison structure (single-arm vs comparative vs external control).** Plots are indexed by their comparison structure: (i) single-arm summaries (e.g., response heterogeneity), (ii) randomized arm-to-arm comparisons (KM curves, AE contrasts), (iii) non-randomized comparisons with adjustment (RWE propensity/balance graphics), (iv) external comparators (hybrid designs, historical controls). This axis helps align visualizations with the causal contrast being communicated.
11. **Time structure (cross-sectional vs longitudinal vs event-history).** We classify plots by how time enters the data: (i) cross-sectional snapshots, (ii) longitudinal trajectories (repeated measures), (iii) event-history with censoring (survival/time-to-event), (iv) cyclic dynamics (e.g., chemotherapy cycles and regrowth patterns). Time structure governs axis semantics, censoring notation, and smoothing/aggregation choices.
12. **Uncertainty representation (inferential layer).** We treat uncertainty as a first-class taxonomy criterion, distinguishing: (i) point estimates only, (ii) frequentist intervals/bands, (iii) Bayesian credible intervals, (iv) resampling-based variability (bootstrap, repeated simulation), (v) posterior predictive displays. The atlas favors explicit uncertainty depiction when a plot is used to support inference rather than description.
13. **Censoring and missingness (data integrity axis).** For time-to-event and longitudinal endpoints, we index plots by their handling of incomplete observation: (i) right censoring (with censor marks and risk tables), (ii) interval censoring or delayed entry, (iii) missing-at-random patterns in longitudinal data, (iv) informative missingness concerns. This axis is essential for preventing visually persuasive but statistically invalid narratives.
14. **Confounding control and balance (RWE diagnostic axis).** In RWE settings, we treat design diagnostics as a distinct plot family: cohort attrition/selection, covariate balance displays (pre/post adjustment), overlap/positivity checks, and sensitivity analyses. These graphics are not merely descriptive; they document whether the observational design plausibly supports the intended estimand.
15. **Treatment patterns and patient journey (care pathway axis).** We classify a class of plots devoted to real-world treatment pathways: lines of therapy, switching, discontinuation, sequencing, and class transitions. Flow-based graphics (e.g., Sankey-style transitions) are indexed under this axis because they summarize longitudinal care decisions rather than outcomes alone.
16. **Biomarkers and high-dimensional profiles (omics axis).** We separate plots that visualize high-dimensional molecular or multi-omics measurements (expression matrices, mutation landscapes, CNV profiles) from classical clinical endpoints. This axis emphasizes visualization strategies for sparsity, multiplicity, annotation density, and integrative displays linking molecular features to outcomes.
17. **Subgroups and heterogeneity (effect modification axis).** We classify plots that communicate heterogeneity of treatment effect or prognostic stratification: subgroup contrasts, interaction-focused summaries, and risk stratification displays (e.g., CART-style partitions, time-dependent prediction error curves). This axis is distinct because it requires disciplined control of multiplicity, interpretability, and pre-specification versus exploration.
18. **Model diagnostics and calibration (validation axis).** We treat diagnostics as an explicit taxonomy component, including: discrimination summaries, calibration displays, residual-style checks, and time-dependent prediction error. Diagnostic plots are indexed separately because their purpose is to validate a modeling claim rather than to present a clinical result.
19. **Evidence synthesis (aggregation axis).** We classify meta-analytic and study-aggregation plots (e.g., forest-type summaries) under evidence synthesis. This axis captures visuals where the primary variation is between studies/centers rather than between patients, and where heterogeneity is itself a key inferential object.
20. **Decision and health economics (policy axis).**We index plots that support decision-making under uncertainty, including cost-effectiveness or value-of-information style summaries. These plots are distinguished by their explicit dependence on utility, cost, or policy thresholds rather than solely clinical endpoints.
21. **Reporting readiness (publication constraints axis).**Finally, we treat publication readiness as a classification criterion: each plot must be interpretable under typical journal constraints (caption completeness, axis semantics, denominator clarity, legend economy, and color/shape accessibility). Wherever feasible, the atlas promotes plots that remain readable in grayscale print and preserve interpretability under panel compression.

The remainder of the paper instantiates this taxonomy by mapping each plot in the atlas to its study-setting category, endpoint family, objective index, and data-geometry class, accompanied by reproducible R code and simulated data-generating mechanisms that can be adapted to real oncology applications [8, 9].

## 4 Software Tools: R Packages & Shiny Apps

This section is organized in three parts. In Section 4.1, we briefly review the broader software landscape for oncology clinical trial design, analysis, and visualization. In Section 4.2, we narrow the focus to Shiny applications within the R ecosystem that support oncology data visualization and interactive exploration. In Section 4.3, we summarize the R packages used throughout this atlas to generate the figures and reproducible workflows presented in the paper. Finally, in Section 4.4 we discuss the structure of Datasets used in this Atlas.

### 4.1 General Software Landscape for Oncology Trial Design and Visualization

A diverse ecosystem of free and commercial software has been developed to support the design and analysis of oncology clinical trials, spanning sample size determination, dose-escalation and adaptive designs, interim monitoring, and endpoint-specific modeling. In routine practice, these tools are frequently complemented by general statistical platforms and programming languages that facilitate data management, modeling, and figure generation. Representative examples include the following list: nQuery [13], PASS [14], STPLAN [15], StudySize [16], CRM simulator [17], EffTox [18], BMA-CRM [19], TITE-CRM [20], ATDPH1 [21], Modified CRM v2.0 [22], JLB design [23], EWOC [24], Simon two-stage design [25], Green–Benedetti–Crowley [26], Bryant–Day design [27], predictive probability design [28], BFDesigner [29], Multc99 [30], CTD system [31], SWOG one-arm binomial [32], SWOG one-arm survival [33], SWOG two-arm survival [34], biomarker-targeted randomized design [35], biomarker-stratified randomized design [36], optimal two-stage designs for phase II trials [37], sample size for integrated phase II/III trial [38], EAST [39], PEST [40], gsDesign [41], ADDPLAN [42], S+SeqTrial [43], SWOG expected death on a study [44], adaptive randomization [45], parameter solver [46], predictive probability calculation [47], TTEDesigner [48], interaction survival [49], Biometric Research Branch of NCI [50], power and sample size (UCSF) [51], sample size (SWOG) [52], Simon’s two-stage design (UPCI) [53], MD Anderson software download [54], Johns Hopkins Comprehensive Cancer Center [55], and MGH Biostatistics Center [56]. In addition, widely used general-purpose platforms include SAS [57], S-PLUS [58], SPSS [59], Stata [60], GraphPad Prism [61], BUGS/WinBUGS/JAGS [62], Python [63–65], and R [7].

Although several of these environments provide visualization capabilities, the emphasis of the present atlas is on reproducible, code-first visualization workflows in R.

### 4.2 R Shiny applications for oncology data visualization

Within R, a substantial collection of Shiny applications has been developed to enable interactive visualization and exploratory analyses for oncology datasets, including multi-omics exploration, survival analysis dashboards, and specialized interfaces for mutation/CNV and expression workflows. In this paper, we reference the following Shiny applications as representative oncology visualization resources: TCGAbiolinksGUI [66], UCSCXenaShiny [67, 68], Sherlock-Genome [69], RaCE [70], ShinyTHOR [71], GNO-SIS [72], surviveR [73], SmulTCan [74], SurvivalGenie [75], OncoProExp [76], PCAS [77], shinyDeepDR [78], Onko_DrugCombScreen [79], TPWshiny [80], CancerClarity [81], StoryboardR [82], BodyMapR [83], shinyOP-TIK [84], SCAN360 [85], iDEP [86], ideal [87], iSEE [88], ShinyCell [89], shinyMethyl [90], SurvivalMeth [91], ShinyCNV [92], CNViz [93], mafShiny [94], CRI iAtlas [95], TRGAted [96], PHDShinyBreastCancer [97], and xOmicsShiny [98].

These tools highlight the breadth of interactive visualization capabilities available to oncology researchers; however, the primary objective of the present atlas is to provide manuscript-ready plots produced directly from R code.

### 4.3 R Packages Used in This Atlas

Across this paper, all figures are generated from simulated data within R [8, 9]. This design choice serves two goals: (i) to ensure full reproducibility of each visualization and (ii) to provide flexibility for readers to adapt the underlying code and data-generating mechanisms to their own oncology settings (e.g., endpoint definitions, effect sizes, censoring patterns, and subgroup structures). Consequently, we prioritize direct use of R packages for figure generation, rather than relying on external GUI-based software.

In total, we use 47 R packages in this atlas, including the following list:

Binom [99]; BiocManager [100]; broom [101]; circlize [102]; cmprsk [103]; ComplexHeatmap [104]; Complex-Upset [105]; cowplot [106]; DecisionCurve [107]; DiagrammeR [108]; DiagrammeRsvg [109]; dplyr [110]; fish-plot [111]; forcats [112]; ggoncoplot [113]; ggplot2 [114]; ggplotify [115]; ggrepel [116]; Hmisc [117]; htmlwidgets [118]; meta [119]; msm [120]; mstate [121]; networkD3 [122]; patchwork [123]; plotrix [124]; png [125]; precrec [126]; purrr [127]; rms [128]; rnaturalearth [129]; rnaturalearthdata [130]; rpact [131]; rpart [132]; rpart.plot [133]; rsvg [134]; scales [135]; sf [136]; stats [137]; survival [138]; survivalROC [139]; tibble [140]; tidyr [141]; tidytext [142]; tidyverse [143]; timeROC [144]; and webshot2 [145].

The complete R scripts used for data simulation, data processing, analysis, and figure generation are provided in the Supplementary Materials. In terms of the cross-classification of oncology visualization tools, the present paper focuses on the subclass of tools based on the R software platform and operating in the static mode (Figure 1).

**Figure 1:**
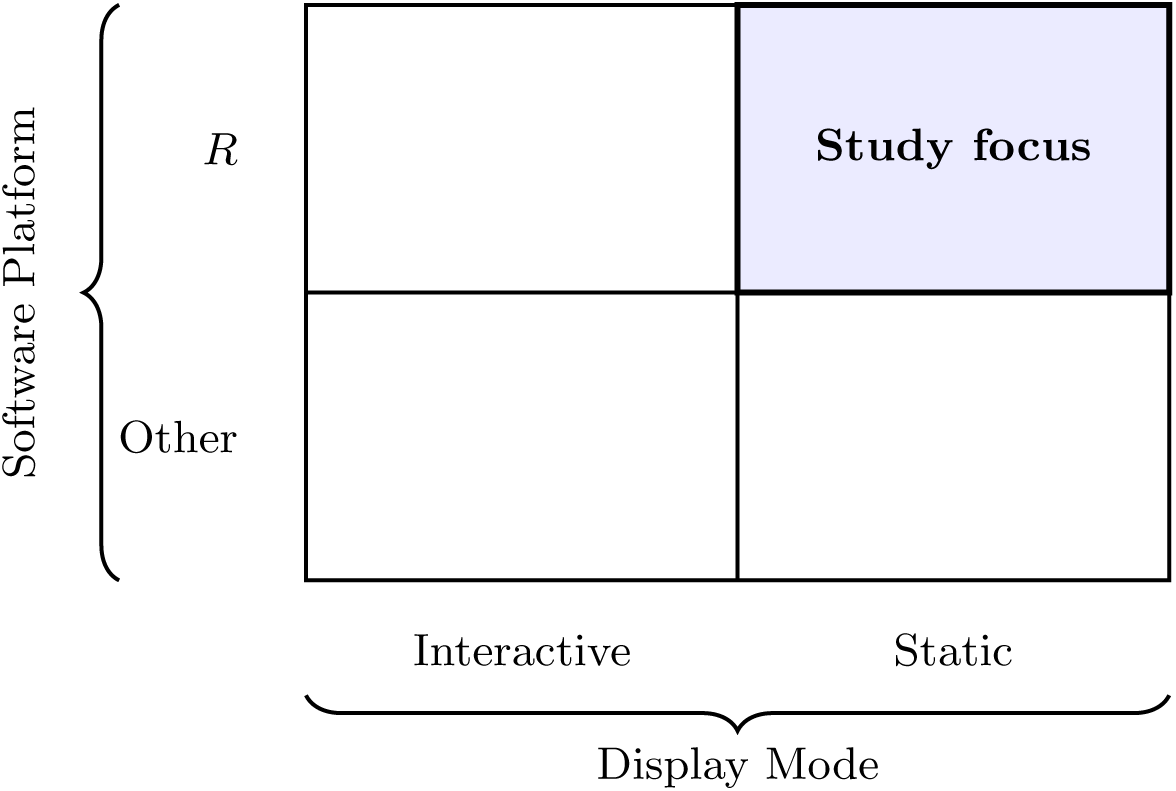
A 2 × 2 cross-classification of oncology visualization tools by software platform (*R* vs. other) and display mode (interactive vs. static). The present paper focuses on the *R*–static subclass.

### 4.4 Simulated Datasets

Across this atlas, all figures are generated from simulated datasets constructed within R using the package ecosystem summarized in Section 4.3. This simulation-first design is deliberate. First, it guarantees full computational reproducibility of the visualizations reported in Section 5. Second, it allows readers to modify the underlying data-generating mechanisms in a controlled manner, thereby adapting each template to alternative oncology settings, endpoint definitions, censoring structures, subgroup compositions, or treatment-pathway patterns. In this sense, the simulated datasets used here should be understood not as surrogates for any single empirical oncology database, but as pedagogical and methodological devices designed to expose the data geometry and inferential logic underlying each plot.

The simulations are constructed to be qualitatively faithful to canonical oncology use-cases. For example, early-phase dose-finding figures are generated from stylized toxicity mechanisms; survival figures are based on plausible time-to-event structures with censoring; safety displays arise from adverse-event count or percentage tables; and molecular displays are derived from synthetic alteration matrices and annotation tables. Throughout, the objective is not to claim empirical realism at the patient level for every numerical value, but rather to reproduce the structural features that make each visualization scientifically interpretable: ordering, grouping, censoring, uncertainty, aggregation, and longitudinal evolution. This convention is consistent with the broader pedagogical strategy of the atlas, namely, to separate the data-generating mechanism, the data-processing layer, and the plotting layer.

#### How to read the plot specifications

In all plot definitions below, the item *Data* refers to the minimum sufficient plotting schema. Unless otherwise stated, it implicitly includes the observational unit and row structure (e.g., patient, visit, study, or simulation replicate), together with the relevant data geometry (long table, wide table, matrix, or edge list). Thus, *Data* should be interpreted as a compact description of the minimal input object required to construct the plotting display.

This interpretation is vital because the atlas spans multiple units of analysis. In many figures, the natural unit is the patient; in others, it shifts to the visit, adverse-event term, or molecular feature. Consequently, the same plot family may depend on different data geometries even when displayed variable names appear similar. The reader should therefore understand each *Data* item in light of the observational-unit framework introduced earlier, rather than as a simple inventory of columns.

Moreover, variables listed under *Data* are the primary observed quantities supplied to the workflow. By contrast, many plotted quantities—such as confidence limits, Kaplan–Meier estimates, cumulative incidence functions, standardized mean differences, or net benefit values—are obtained after statistical summarization or package-mediated computation. Similarly, in flow-based visualizations, the plotting object may be an aggregated transition table computed from patient-level records. In such cases, the variables named in *Data* identify the minimal upstream inputs from which the plotting coordinates are constructed.

Accordingly, the reader should distinguish between three conceptual layers when interpreting a figure in Section 5: the *source data layer* (simulated observations), the *derived analysis layer* (transformations into estimands or summaries), and the *visualization layer* (rendering via R packages). This distinction is especially relevant for figures showing computed summaries rather than raw measurements, such as forest plots, calibration curves, and several survival-related graphics.

For this reason, when a variable appears to represent an estimate, risk score, or balancing metric, it should be read as a derived variable obtained through a standard and reproducible construction. In many cases, the transformation is implicit in the named package; in others, it follows from formulas introduced earlier. The atlas keeps figure definitions concise, but this compact notation reflects a design choice to rely on the common technical language established in the preliminaries.

Finally, for several figures, the plotting table is an aggregated, reshaped, or filtered object derived from the simulated dataset. Therefore, throughout Section 5, the term *Data* denotes the minimal schema required for valid plot construction, including the structural and computational context needed for the final display. This convention allows us to preserve a uniform specification format while maintaining clarity regarding data provenance and reproducible implementation.

## 5 Atlas of Plots

### 5.1 Category A: Clinical Trials

#### 5.1.1 Prob of DLT–Prob of Escalation Plot

- **Technical Specification**

**– Purpose:** This plot answers: How likely is the design to escalate to the next dose level as a function of the true Dose-limiting toxicity(DLT) probability at the current dose? It compares escalation aggressiveness for two rule-based designs (3+3 vs Best-of-5) [1].
**– Data:** Required variables — p (true DLT probability in [0,1]) and design (factor with levels “3+3”, “Best-of-5”).
**– Assumption:** Patient DLT outcomes at the current dose are i.i.d. Bernoulli with constant probability p; decisions follow the canonical rules with no additional stopping.
**– R Packages:** tidyverse, ggplot2, dplyr
- **Applications**

**– Use-case 1.** For pre-trial design selection in phase I oncology, to compare how aggressively different rule-based algorithms escalate across plausible toxicity rates.
**– Use-case 2.** For protocol and team communication, to explain the operating behavior of the chosen design and justify dose-escalation safeguards or modifications.
- **• Visualization**

**Figure 2:**
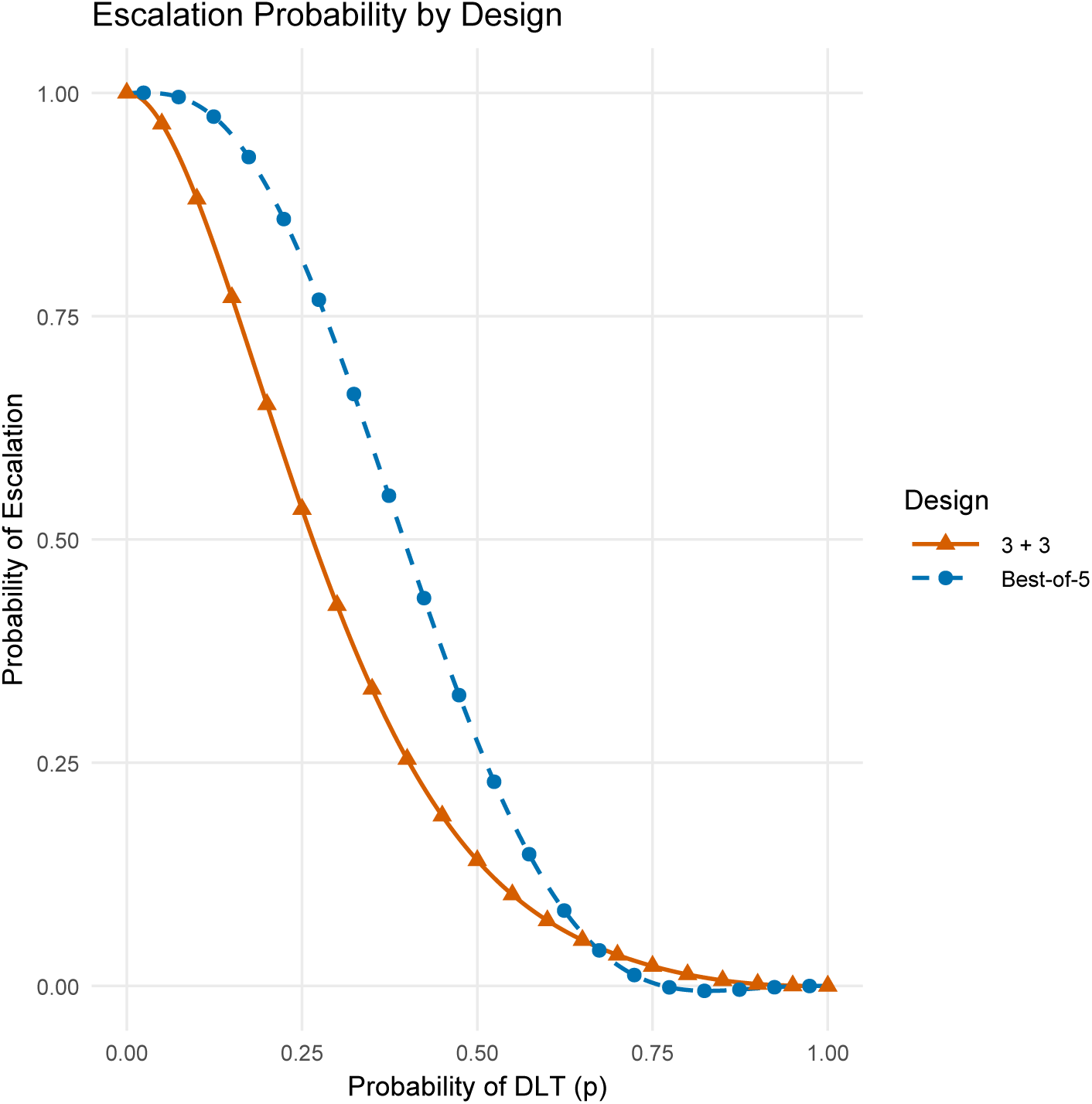
Level-wise probability of escalating to the next dose as a function of the true DLT probability under two traditional algorithms (Best-of-5 vs. 3+3).

#### 5.1.2 Prob of DLT–Mean of p(MTD) Plot

**• Technical Specification**

**– Purpose:** This plot answers: How toxic is the recommended dose on average? Specifically, it shows the mean of the true Dose-limiting toxicity (DLT) probability at the selected Maximum tolerated dose(MTD), *E*[*p*(*MTD*)], as a function of the starting-dose toxicity [1].
**– Data:** Required variables-p_start ∈ [0, 0.20] (true DLT at starting dose), design ∈ {*Two* − *stage, CRM* − *L, Best* − *of* − 5, 3 + 3}, and mean_pMTD (Monte Carlo mean of p(MTD)).
**– Assumption:** Monotone dose–toxicity; i.i.d. Bernoulli DLTs within a dose; canonical implementations of each design on a fixed dose grid and total sample size; mean computed over many replicated trials.
**– R Packages:** tidyverse, ggplot2, dplyr, tidyr.
**• Applications**

**– Use-case 1.** Target-toxicity calibration: assess whether each design centers its recommended MTD near the target toxicity (e.g., 0.30–0.33) and how this shifts when the starting dose is misspecified.
**– Use-case 2.** Design selection and protocol justification: compare expected toxicity of the chosen MTD across designs to motivate dose-grid choices, skeletons/prior settings (for CRM-L), and early stopping/safety rules.
**• Visualization**

**Figure 3:**
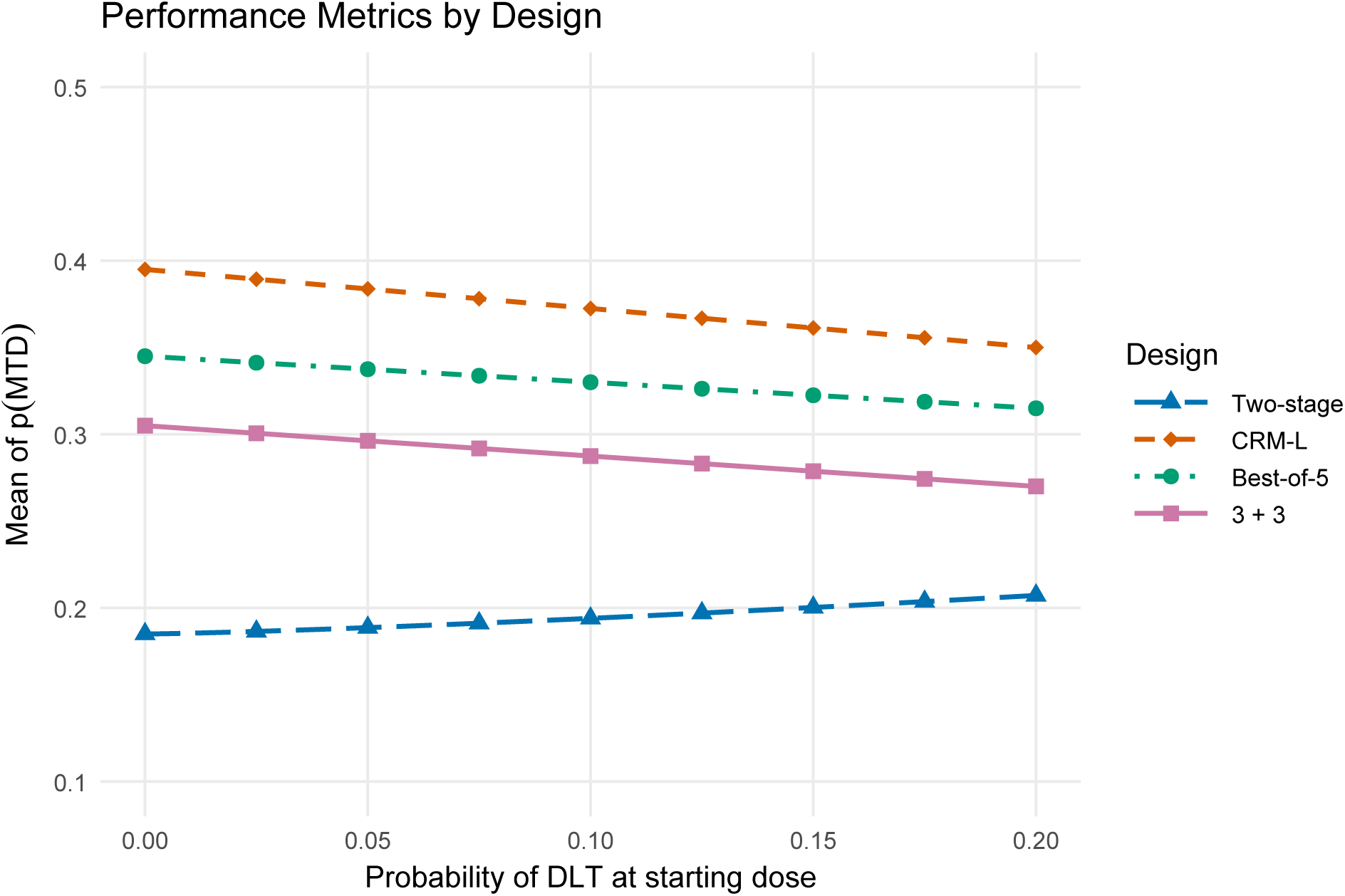
Approximated mean of the true DLT probability at the selected MTD, *E*[*p*(*MTD*)], as a function of the true DLT at the starting dose, comparing Two-stage, CRM-L, Best-of-5, and 3+3 designs. Curves are parametric approximations chosen to match the qualitative behavior of the reference panel.

#### 5.1.3 Trial History (Subject number–Dose Level)

**• Technical Specification**

**– Purpose:** This plot answers: How did dose levels evolve patient-by-patient and where did toxicities occur in each group? It visualizes the escalation/de-escalation path and DLT events over the course of a trial [1, 146, 147].
**– Data:** patient (1, · · · *, N*), group (factor: “Group1”, “Group2”), dose_level (numeric dose index), and toxicity (logical; optional tox_dose to position the marker on the y-axis).
**– Assumption:** Sequential enrollment; outcomes are i.i.d. Bernoulli at each dose; canonical rules (no dose skipping beyond protocol); two parallel groups following the same dose grid; markers denote patients with DLT.
**– R Packages:** tidyverse, ggplot2, dplyr, tidyr.
**• Applications**

**– Use-case 1.** Operational review: visualize escalation/de-escalation decisions, identify where DLTs occurred, and verify adherence to protocol rules during simulations or real-time monitoring.
**– Use-case 2.** Communication/teaching: illustrate how different patient outcomes drive the path of dose assignments across parallel groups, supporting discussions with investigators and DSMBs.
**• Visualization**

**Figure 4:**
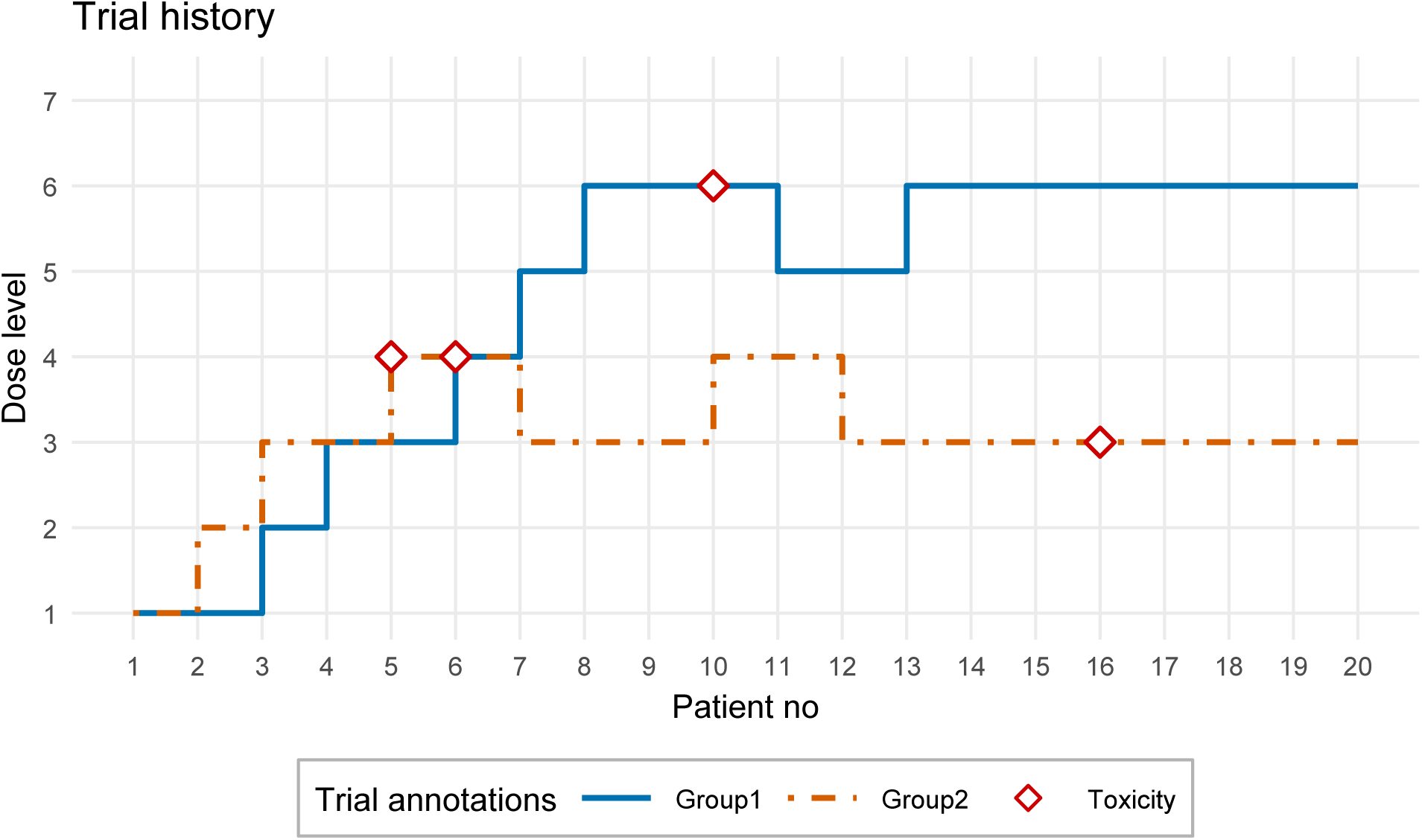
Simulated trial history for two groups. Step lines show per-patient dose levels; diamond markers indicate DLT events (“Toxicity”). Curves are parametric approximations to match the qualitative look of the reference panel.

#### 5.1.4 Dose–Efficacy Curve

**• Technical Specification**

**– Purpose:** This plot answers: How does the probability of efficacy vary with dose for different mechanistic patterns? It compares three canonical shapes observed with targeted/novel agents: linear increase, saturating Emax (monotone), and non-monotone inverted-U (loss of efficacy at very high dose) [1, 148, 149].
**– Data:** dose ∈ [0, 1], curve (factor: “Linear”, “Emax (saturating)”, “Inverted-U”), and eff (efficacy probability).
**– Assumption:** Dose is scaled to [0,1]; probabilities are on [0,1]; the three curves are stylized generative forms chosen to emulate typical behavior in the literature (not fitted to data).
**– R Packages:** tidyverse, ggplot2, dplyr, tidyr.
**• Applications**

**– Use-case 1.** Design choice and objective selection: determine whether a monotone or non-monotone working model is appropriate for phase I/II methods (e.g., MCP-Mod, BLRM/Emax vs. flexible splines) and align target-dose definitions (EDp, OBD) with plausible curve shapes.
**– Use-case 2.** Scenario building for simulations: encode mechanistic hypotheses (e.g., target saturation, off-target loss of efficacy) into simulation truth curves to evaluate the robustness of adaptive designs and endpoint-driven stopping rules.
**• Visualization**

**Figure 5:**
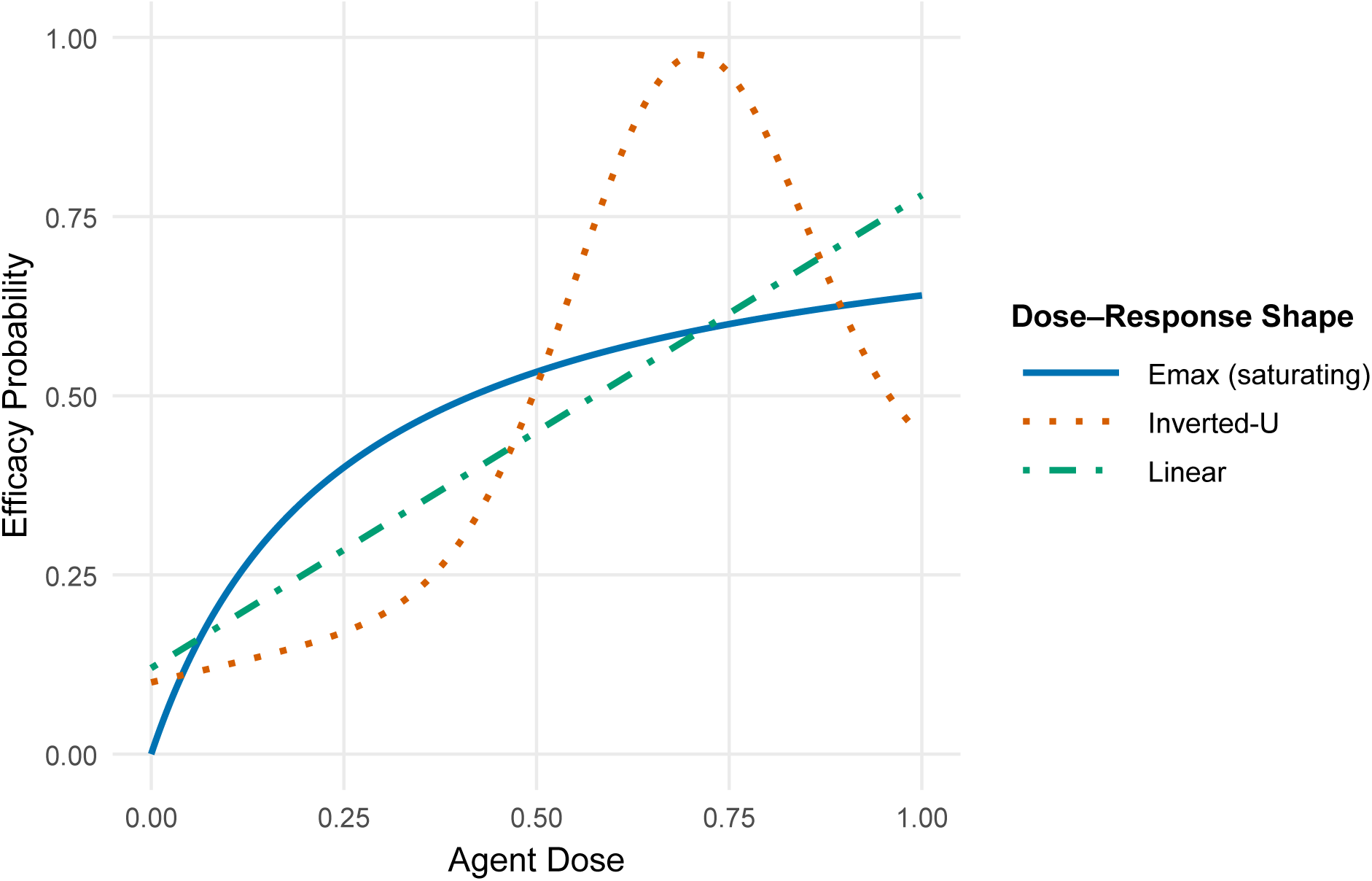
Stylized dose–efficacy relationships: linear increase (long-dash), monotone saturating Emax (solid), and non-monotone inverted-U (dotted). Parameter choices are selected to match the qualitative behavior of the reference panel; no empirical fitting was performed.

#### 5.1.5 Sample Size–Power Curve

**• Technical Specification**

**– Purpose:** This figure answers: How does required total sample size change with the noninferiority (NI) margin on the hazard ratio under exponential time-to-event(TTE) assumptions, for two target powers (80% and 90%)? [1, 150–152]
**– Data:** HR_margin (NI margin on the hazard ratio), *N* (required total sample size), Power (factor with levels 80% and 90%).
**– Assumption:** Two-arm NI design, log-rank test with proportional hazards and exponential survival; one-sided *α* fixed (implicitly), equal allocation, constant accrual over a fixed accrual window and a common follow-up window; curves shown here use illustrative numbers that match the qualitative pattern in your panel.
**– R Packages:** ggplot2, dplyr, tidyr, scales
**• Applications**

**– Use-case 1.** Design calibration and protocol justification: quantify how tightening the NI margin (e.g., HR 1.30 → 1.10 → 1.05) inflates the required N at fixed power, aiding feasibility assessments and trade-off discussions with stakeholders.
**– Use-case 2.** Sensitivity analysis: compare operating points at 80% versus 90% power to evaluate the incremental sample size required for higher power and to choose an achievable design given accrual and follow-up constraints.
**• Visualization**

**Figure 6:**
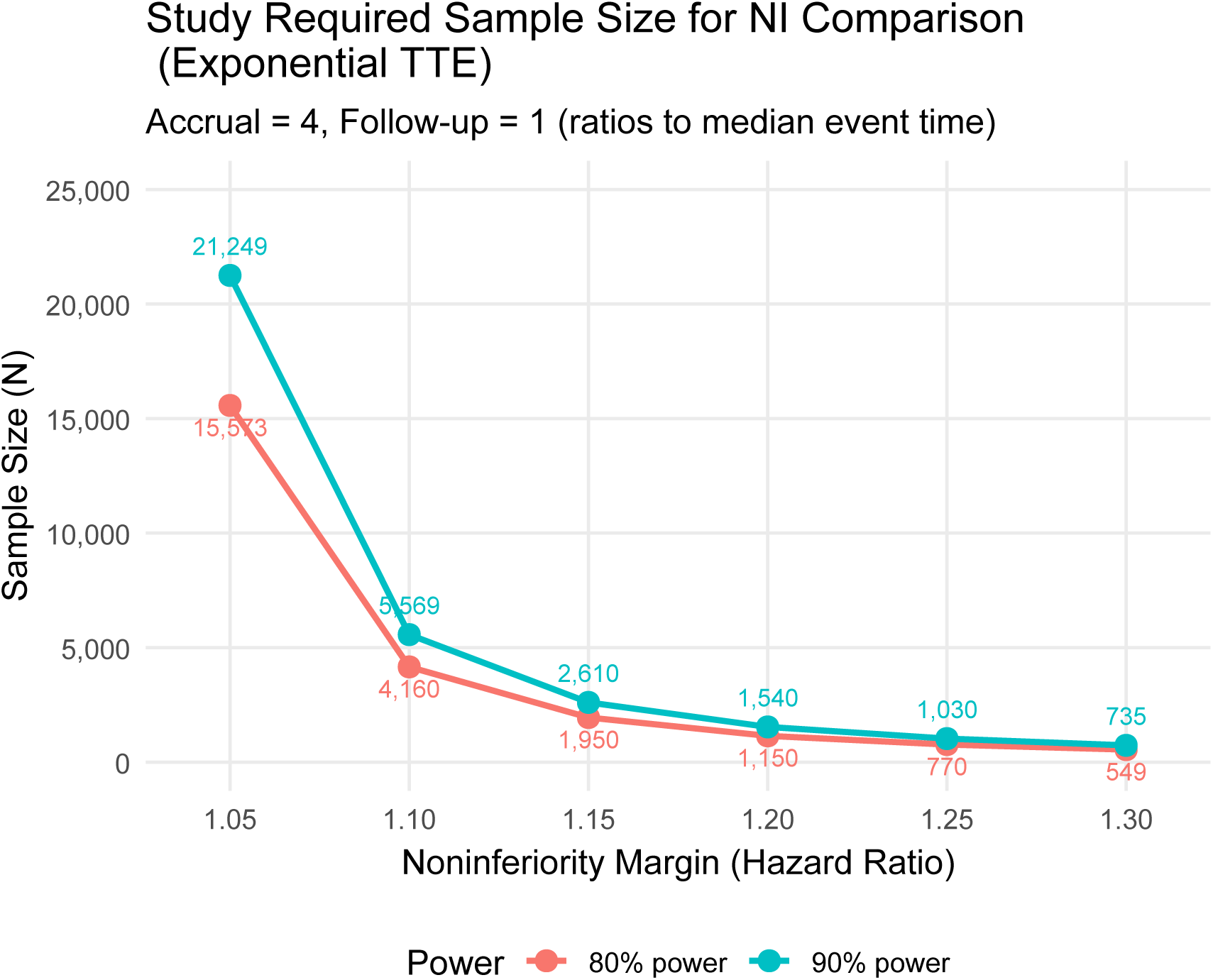
Required total sample size for an NI comparison under exponential TTE assumptions across three hazard-ratio margins for 80% and 90% power. Values are illustrative and chosen to match the qualitative pattern of the reference panel.

#### 5.1.6 Kaplan–Meier Curve

**• Technical Specification**

**– Purpose:** This figure answers: How do overall/progression-free survival (OS/PFS) profiles differ between two treatment strategies over time, including uncertainty? It displays Kaplan–Meier (KM) survival estimates with 95% confidence bands and censoring marks [1, 2, 6, 153].
**– Data:** time (years), status (1 = event; 0 = censored), arm =Chemotherapy, Hormone Therapy.
**– Assumption:**Independent right censoring; administrative censoring at 16 years; event times sampled to produce two stochastically ordered survival distributions (Chemotherapy worse than Hormone Therapy). Confidence intervals are Greenwood log–log intervals.
**– R Packages:** survival, ggplot2, dplyr, cowplot, tidyr.
**• Applications**

**– Use-case 1.** Comparative effectiveness: visualize separation of OS/PFS between two strategies with uncertainty bands to support informal assessment of treatment benefit over time.
**– Use-case 2.** Trial reporting and QC: display censoring patterns and verify that CIs extend over the analysis window, aiding checks for administrative censoring and follow-up sufficiency.
**• Visualization**

**Figure 7:**
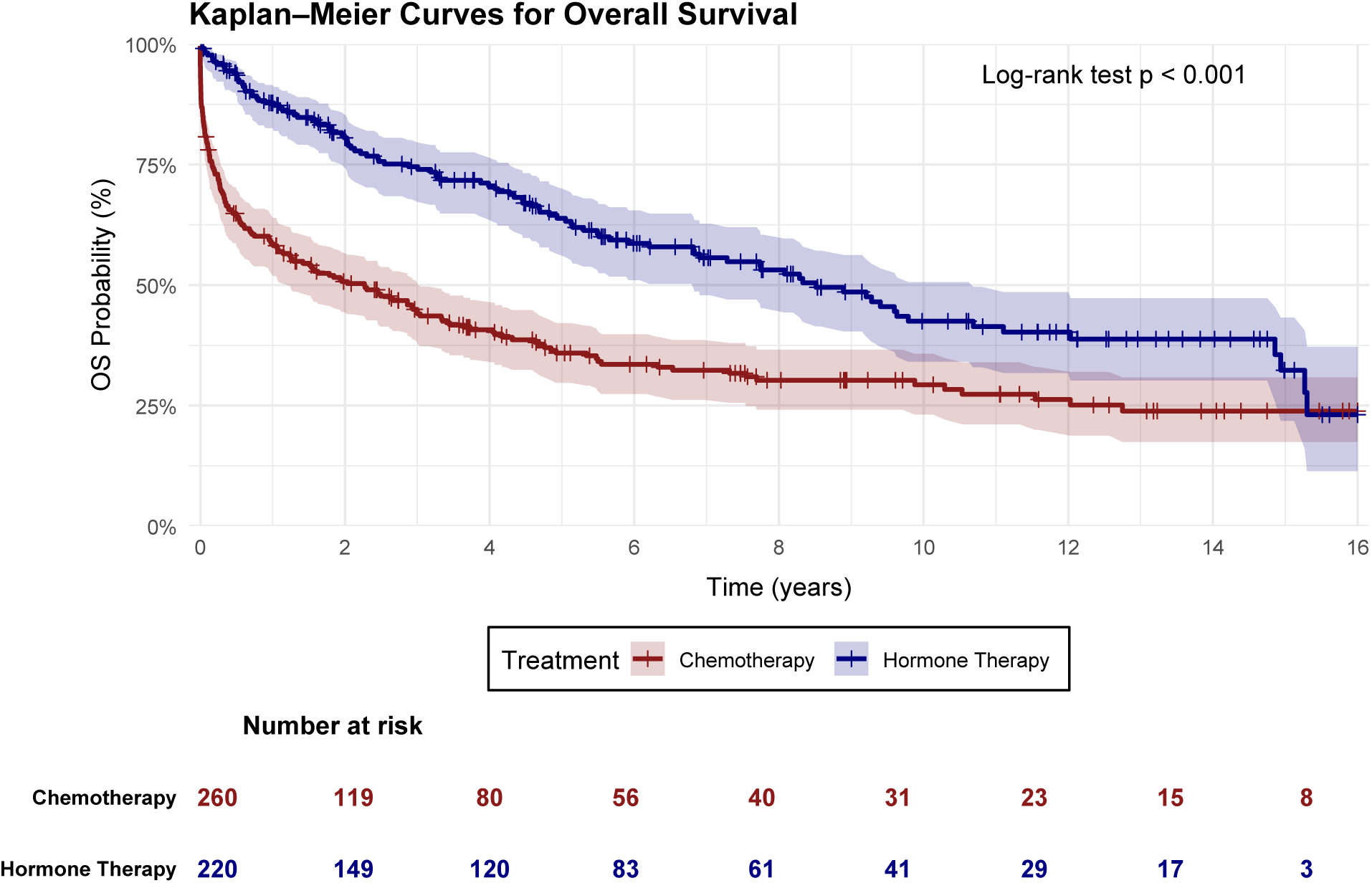
Kaplan–Meier curves for OS comparing Chemotherapy (solid) and Hormone Therapy (dotted). Shaded bands indicate Greenwood 95% CIs extending across the full 16-year horizon. Plus symbols denote censoring times. Curves and marks are generated from a plausible parametric simulation, not empirical data.

#### 5.1.7 Milestone Survival plot

**• Technical Specification**

**– Purpose:** This plot answers the question of what the survival probability is at a specific, clinically significant time point about the long-term treatment effect given time-to-event and group data [154].
**– Data:** Time-to-event (numerical), status/event indicator (binary), and treatment arm (categorical).
**– Assumption:** Independent censoring and particularly useful when the proportional hazards assumption is violated.
**– R Packages:** survival, ggplot2, dplyr, cowplot, tidyr.
**• Applications**

**– Use-case 1.** For progression-free survival in immune checkpoint inhibitor trials, to quantify the “plateau” of the survival curve that median survival fails to capture.
**– Use-case 2.** For overall survival in oncology clinical trials, to provide a cross-study comparison at a standardized intermediate endpoint (e.g., 2-year OS) when hazard ratios are non-constant.
**• Visualization**

**Figure 8:**
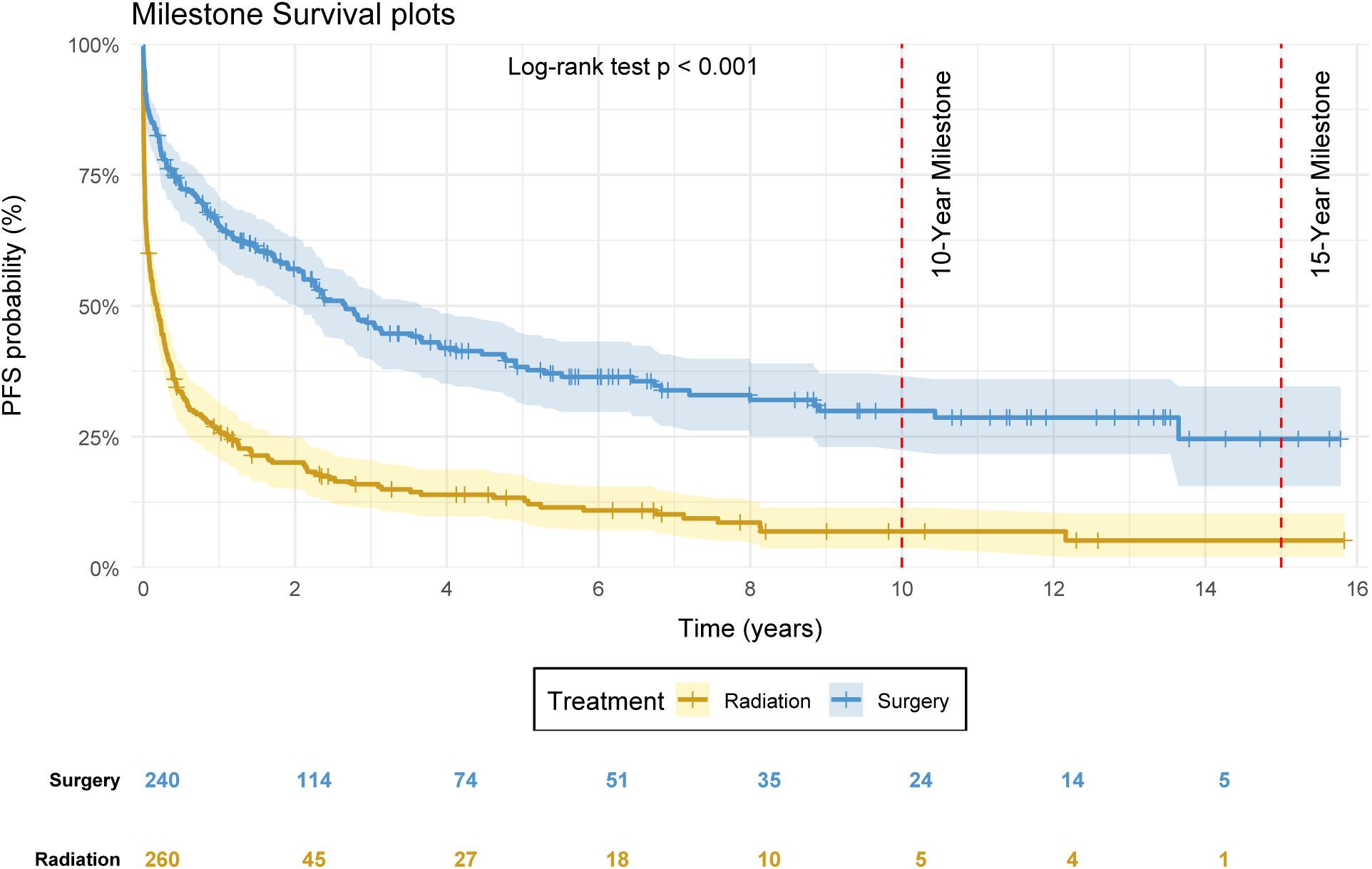
Kaplan–Meier curves for PFS (Radiation vs Surgery) with Greenwood 95% confidence ribbons with milestone marker. The dashed vertical line marks the 10-year milestone. Numbers at risk are listed along the x-axis at scheduled times.

#### 5.1.8 Waterfall chart

**• Technical Specification**

**– Purpose:** This figure answers: For each subject, what is the best tumor size change from baseline and how does it classify under RECIST? Bars above zero indicate growth (progression risk); bars below zero indicate shrinkage (response) [6].
**– Data:** id (subject identifier), best_pct (best percentage change from baseline; negative values indicate tumor shrinkage), and the derived variable recist_cat ∈ {Progression (*>* +20%), Stable (between −30% and +20%), Partial response (≤ −30%)}.
**– Assumption:** One bar per subject (best observed change); classification follows RECIST thresholds described above; subjects are ordered by *best_pct_*.
**– R Packages:** dplyr, ggplot2, forcats, scales.
**• Applications**

**– Use-case 1.** Early activity signal: visualize heterogeneity of responses and the proportion of subjects achieving RECIST partial response versus progression.
**– Use-case 2.** Dose/biomarker exploration: stratify the same plot by dose level or biomarker groups to identify subpopulations with higher shrinkage rates before confirmatory modeling.
**• Visualization**

**Figure 9:**
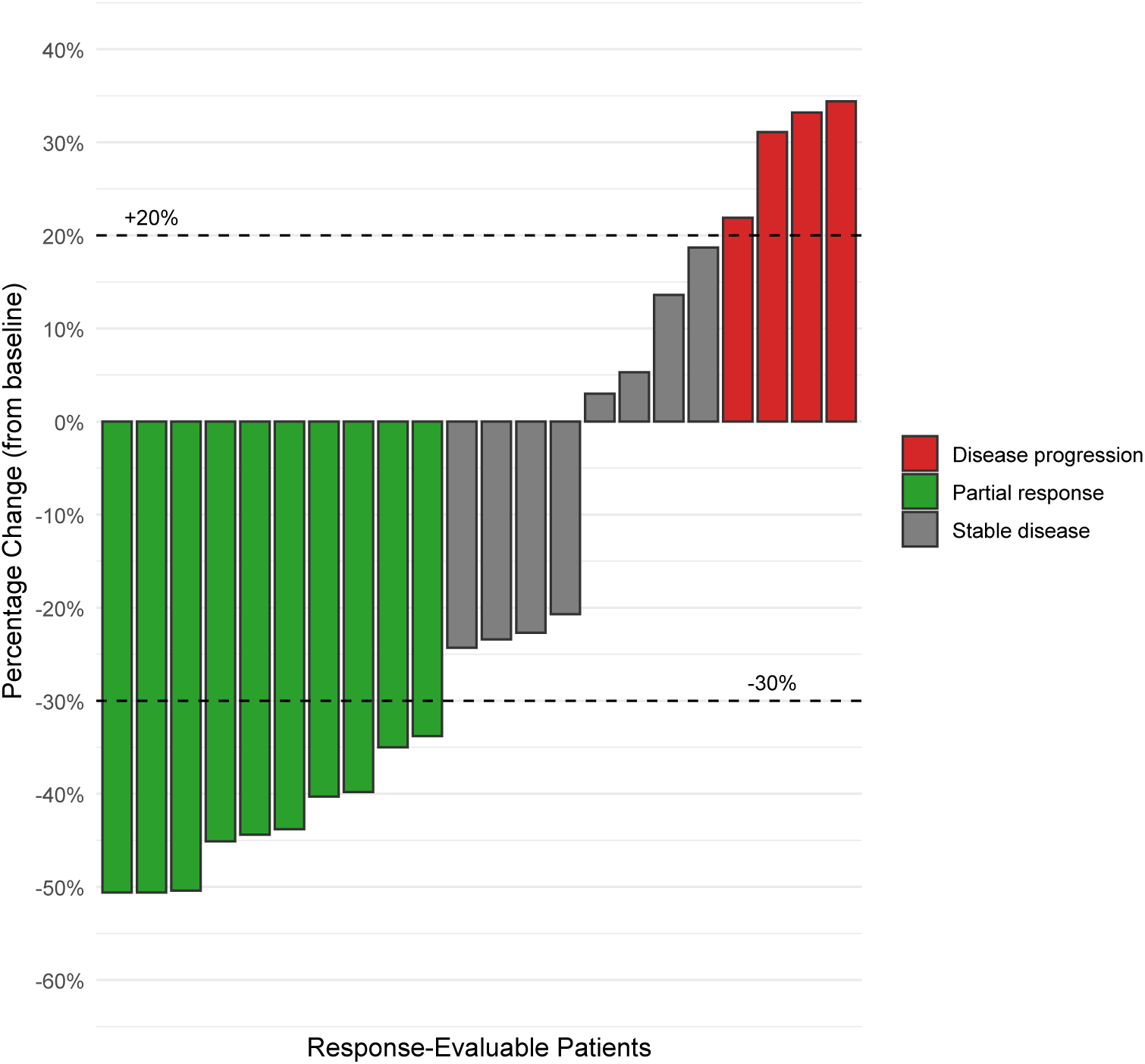
Waterfall plot of best percentage change from baseline by subject. Bars below zero indicate tumor shrinkage (green = partial response <= −30%); bars above zero indicate growth (red = progression > +20%); intermediate changes are gray (stable disease). Dashed lines mark RECIST thresholds at −30% and +20%.

#### 5.1.9 Swimmer Plot

**• Technical Specification**

**– Purpose:** This figure answers: For each subject, how long did treatment last and when did responses start, continue, and end? Which subjects were durable responders, and what was their baseline stage? [6]
**– Data:** Per subject: id, stage (1–4), tx_start, tx_end, ongoing (arrow if TRUE), resp_start (NA if none), resp_cont (NA if none), resp_end (NA if none), progression (NA if none), durable (TRUE/FALSE).
**– Assumption:** Time axis in months; one horizontal bar per subject; bars colored by baseline stage; markers encode response milestones; an arrow at the right indicates ongoing treatment at last contact.
**– R Packages:** dplyr, ggplot2, forcats
**• Applications**

**– Use-case 1.** Clinical activity overview: visualize timing and durability of responses across patients and stages to complement RECIST tables.
**– Use-case 2.** Protocol and review meetings: communicate heterogeneity in exposure and outcomes, and identify durable responders and time-to-progression patterns for subgroup exploration.
**• Visualization**

**Figure 10:**
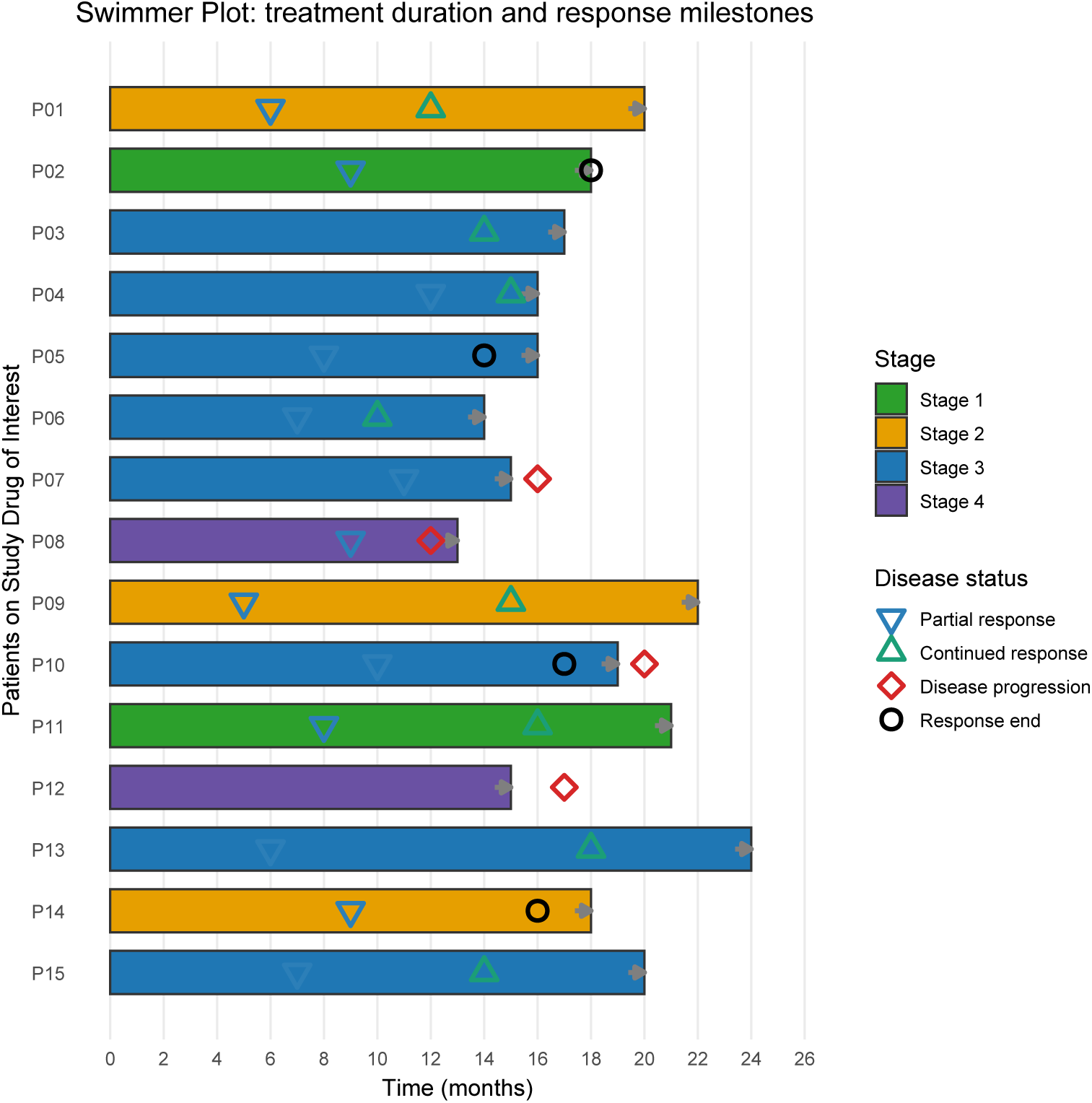
Swimmer plot demonstrating per-patient treatment duration (bar length), response onset (triangle-down), continued response (triangle-up), response end (open circle), and disease progression (diamond). Bars are colored by baseline stage (green/amber/blue/purple). Arrows indicate ongoing treatment at last contact. “Durable response” is implicitly conveyed by long bar segments between response onset/continuation and end or censoring.

#### 5.1.10 Forest plot of study–Log(HR)

**• Technical Specification**

**– Purpose:** This figure answers: Across many replicated analyses (or studies), what are the log-hazard-ratio (log[HR]) estimates and their 95% confidence intervals, and which are statistically different from 0? [1]
**– Data:** For each study *i*: study_id_i, *β_i_*(log–HR), se*_i_*, and CI limits *β_i_* ± 1.96 se*_i_*.
**– Assumption:** Independent studies; Wald CIs with normal approximation; ordering by *β* from smallest to largest; filled points indicate CIs not covering 0.
**– R Packages:** dplyr, ggplot2, purrr, tibble.
**• Applications**

**– Use-case 1.** Stability assessment across resamples or centers: quickly identify the spread of effects and which analyses show statistically meaningful departures from the null.
**– Use-case 2.** Method comparison and shrinkage diagnostics: the distribution of interval widths and significance flags helps evaluate variance estimation approaches and the potential need for shrinkage or hierarchical modeling.
**• Visualization**

**Figure 11:**
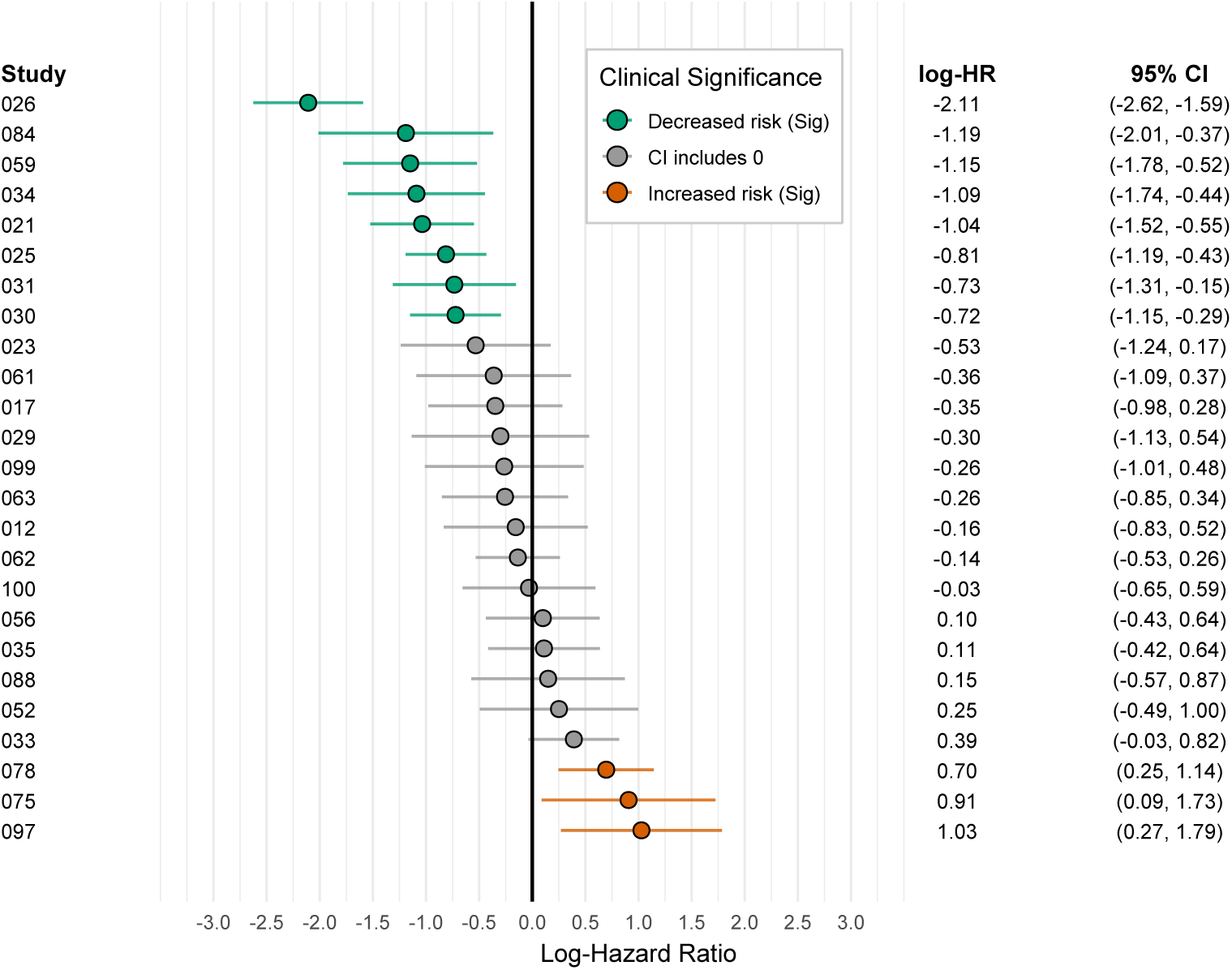
Forest plot: log-hazard-ratio estimates with 95% Wald confidence intervals across 25 studies, ordered by estimate. The vertical line marks *β* = 0. Filled points denote intervals that exclude 0.

#### 5.1.11 Oncoprint

**• Technical Specification**

**– Purpose:** This plot answers: Across a cohort, which genes are altered in which patients, and how do alteration patterns relate to basic clinical annotations (e.g., ER/PR/HER2)? [155]
**– Data:** Required variables — sample_id (string), gene (factor of target genes), alteration (categorical mutation class), and optional clinical covariates per sample (e.g., ER, PR, HER2).
**– Assumption:** Each cell indicates zero or more alterations for a gene–sample pair (simplified here to one alteration at most per cell). Alteration classes are mutually exclusive and encoded with a fixed color key.
**– R Packages:** BiocManager, ComplexHeatmap, ggoncoplot, circlize (colors), dplyr, tibble, tidyr.
**• Applications**

**– Use-case 1.** Pattern discovery and co-mutation checks: quickly identify frequently altered genes (row bars) and high-burden samples (top bars), and visually screen for mutual exclusivity or co-occurrence blocks.
**– Use-case 2.** Clinical linkage and cohort stratification: relate mutation patterns to ER/PR/HER2 status, enabling hypotheses about molecular subtypes or enrichment strategies for downstream analyses.
**• Visualization**

**Figure 12:**
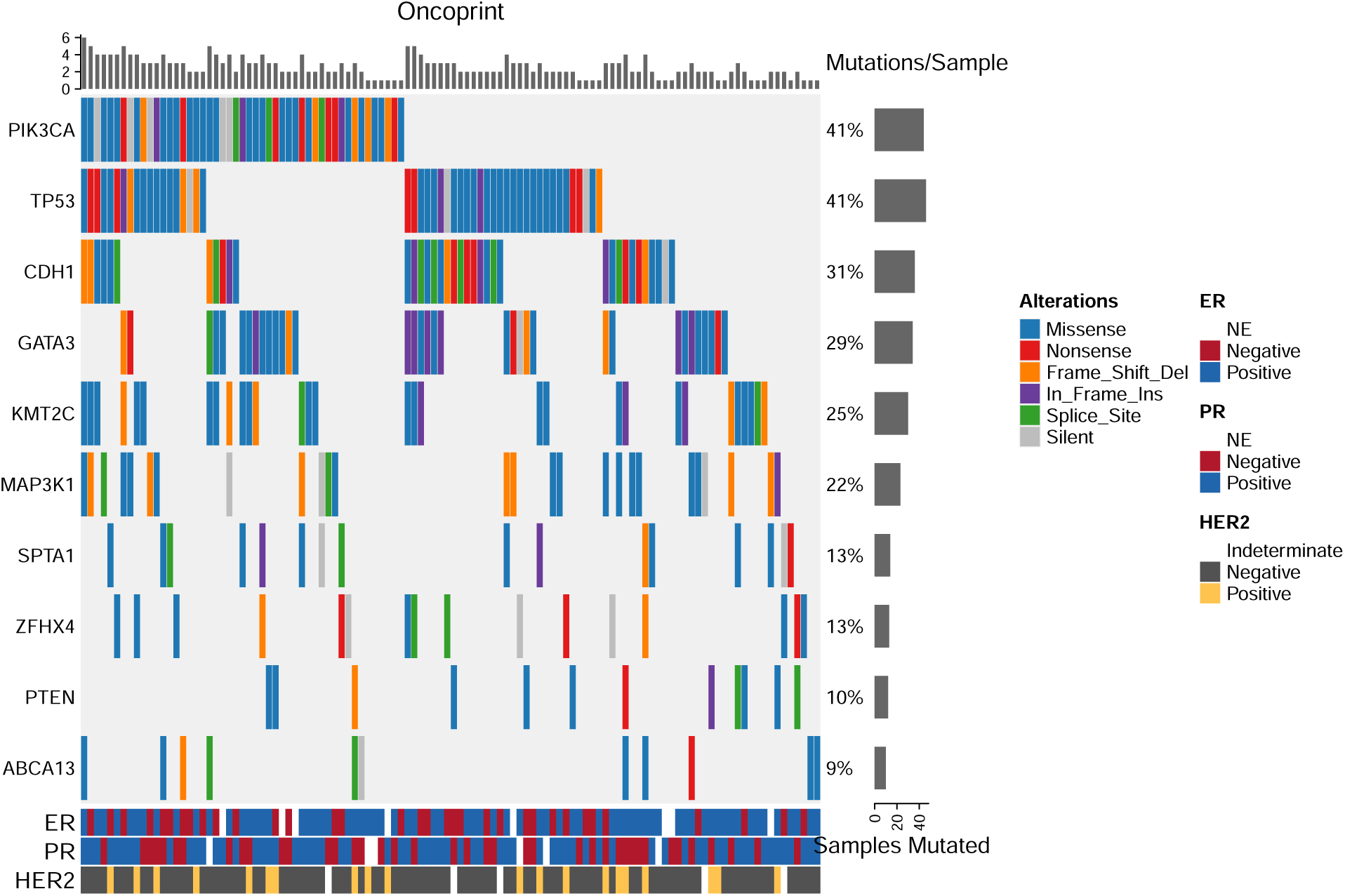
Simplified oncoprint: columns are samples and rows are target genes. Cells encode alteration type; the top bar plot shows mutations per sample, the right bar plot shows the number of samples mutated per gene, and the bottom strips annotate ER/PR/HER2 status. Colors denote alteration classes(PIK3CA: phosphatidylinositol-4,5-bisphosphate 3-kinase catalytic subunit alpha; GATA3: GATA binding protein 3; CDH1: cadherin 1; TP53: tumor protein p53; KMT2C: lysine methyltransferase 2C; MAP3K1: mitogen-activated protein kinase kinase kinase 1; SPTA1: spectrin alpha, erythrocytic 1; ZFHX4: zinc finger homeobox 4; PTEN: phosphatase and tensin homolog; ABCA13: ATP binding cassette subfamily A member 13; ER: estrogen receptor; PR: progesterone receptor; HER2: human epidermal growth factor receptor 2; NE: not evaluable).

#### 5.1.12 Adverse Events by Grade (stacked)

**• Technical Specification**

**– Purpose:** This plot answers: Which grade 3–5 adverse events are most frequent, and how do rates differ between two study arms (ASC alone vs ASC+FOLFOX), including the subset deemed chemotherapy-related [156, 157]
**– Data:** Required variables — ae (event term), arm (ASC alone, ASC+FOLFOX), pct (percentage of patients), and type (All grade 3–5; Chemo-related grade 3–5).
**– Assumption:**Percentages are simulated and represent intent-to-treat incidence; each AE is mutually exclusive for labeling but not for occurrence; events are displayed from most to least frequent by the combination arm within each panel.
**– R Packages:** dplyr, tidyr, ggplot2, scales, tidytext.
**• Applications**

**– Use-case 1.** Safety profile communication: quickly compare the burden and pattern of serious AEs between treatment strategies and highlight chemotherapy-related toxicities.
**– Use-case 2.** Protocol and label discussions: identify events exceeding thresholds (e.g., >=5%) to prioritize risk-mitigation strategies, patient-information content, or safety-monitoring plans.
**• Visualization**

**Figure 13:**
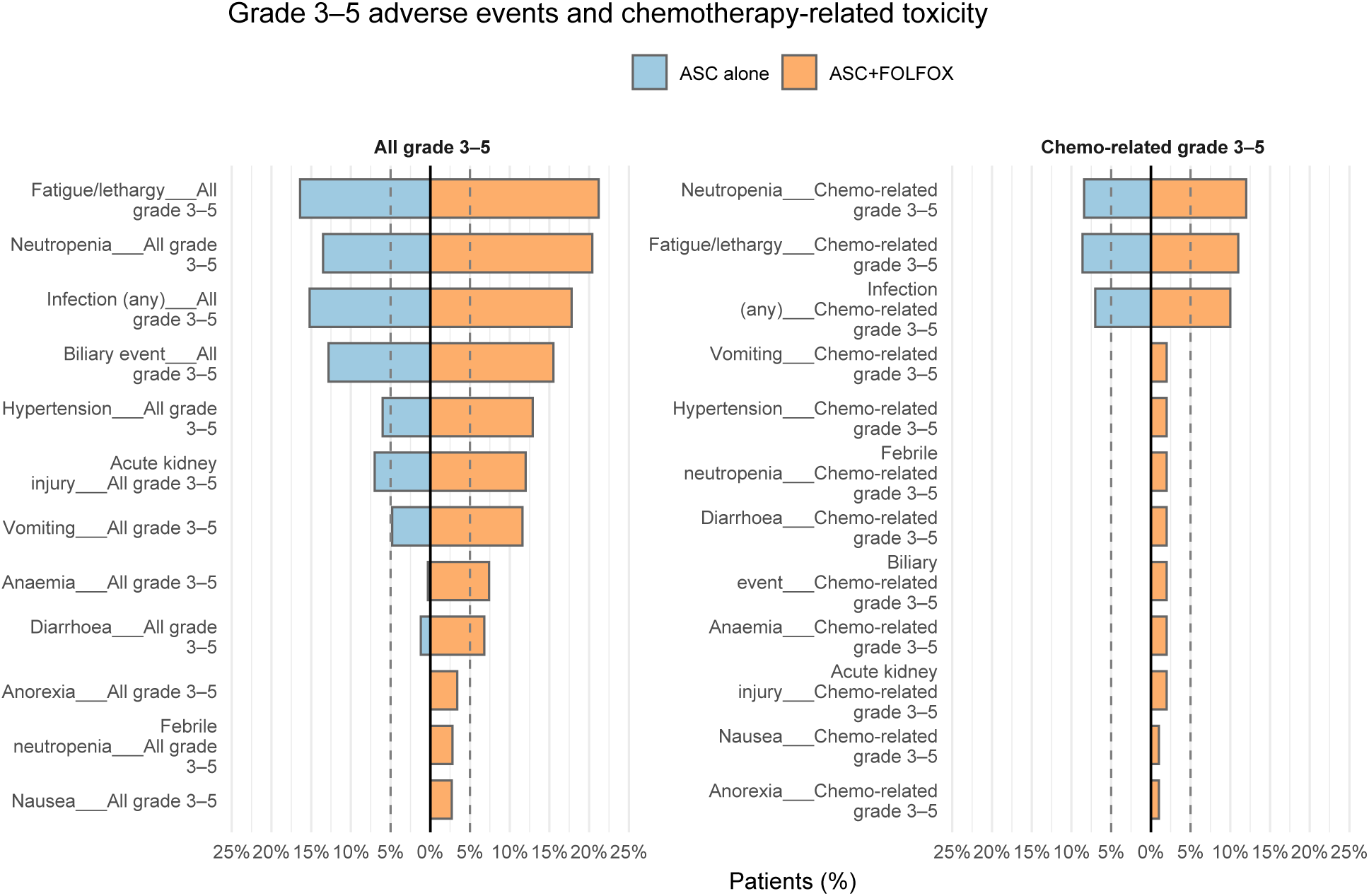
Mirrored bar charts of grade 3–5 adverse event rates comparing ASC alone (left, blue) and ASC+FOLFOX (right, orange). The dashed lines mark 5% reference levels. A second panel shows chemotherapy-related grade 3–5 events.

#### 5.1.13 Adverse Events Heatmap (term by grade/frequency)

**• Technical Specification**

**– Purpose:** This plot answers: Which oncology AEs occur most often and how do their frequencies compare across treatments [158, 159]
**– Data:** Required variables — term (preferred AE term), treatment (e.g., A/B/C/D), n (event count).
**– Assumption:** Counts are intent-to-treat tallies (multiple events per patient allowed); no grade stratification in this simplified version; color encodes frequency.
**– R Packages:** dplyr, ggplot2, tidyr, scales.
**• Applications**

**– Use-case 1.** Safety signal detection: quickly identify high-frequency AEs and treatments associated with increased counts.
**– Use-case 2.** Cross-arm comparison and protocol review: compare toxicity profiles across regimens to prioritize monitoring or mitigation strategies.
**• Visualization**

**Figure 14:**
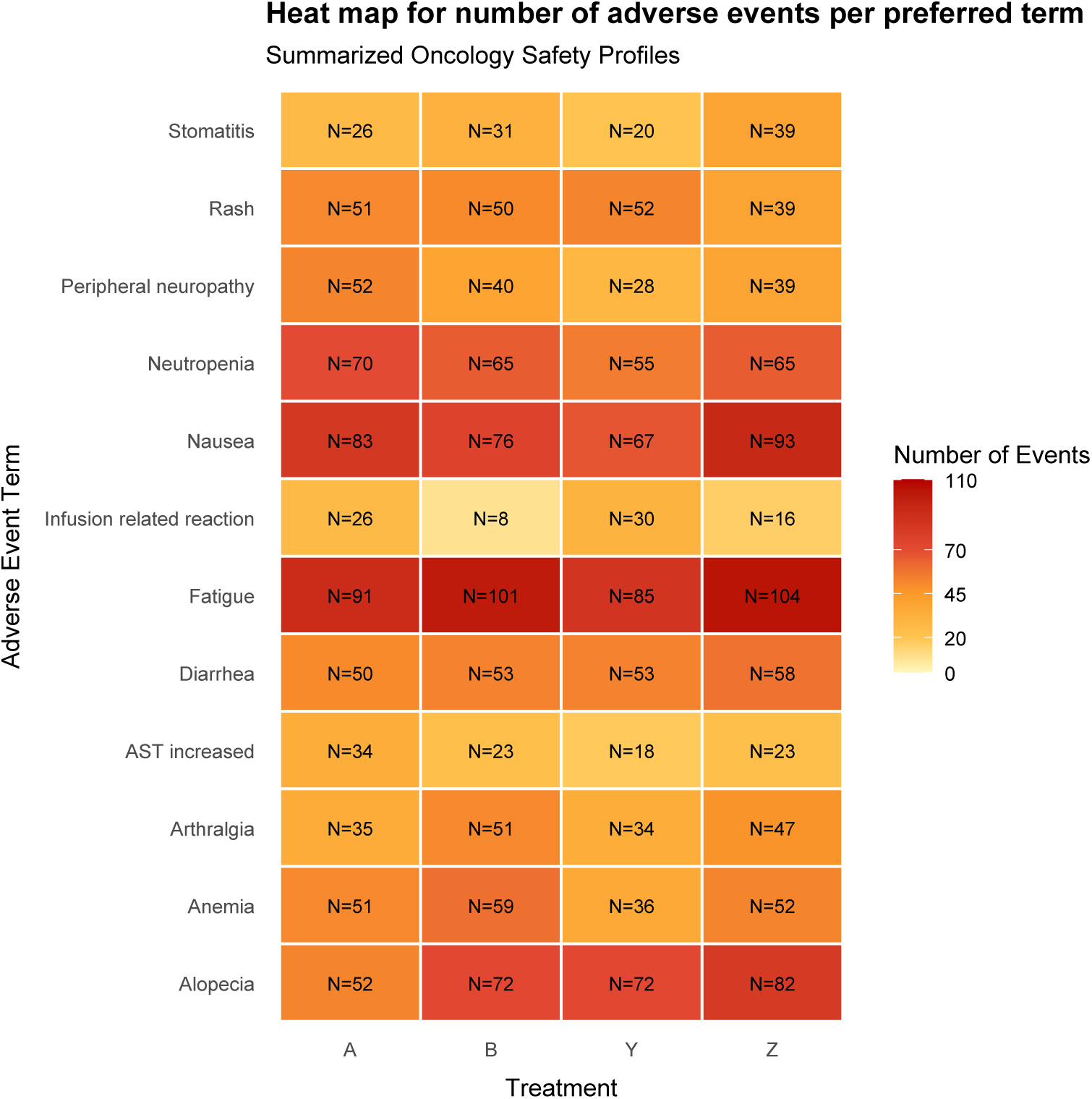
Heat map of AE counts by preferred term (rows) and treatment (columns). Warmer colors indicate higher frequencies; cell labels display counts.

#### 5.1.14 Dose intensity / modification over time

**• Technical Specification**

**– Purpose:** This plot answers: How does the administered dose level change over study time for each patient, and when do dose reductions/escalations occur [160]
**– Data:** Required variables — *patient_id_* (factor), day (study day), dose (planned/administered level), optionally offset (small y-shift to avoid overplotting when lines coincide).
**– Assumption:** Dosing is piecewise constant between change points; changes occur only at visit days; no imputation across gaps; plotted value is the actually administered dose level (not relative intensity).
**– R Packages:** dplyr, tidyr, ggplot2 (step geometry via *geom_step_/geom_segment_*).
**• Applications**

**– Use-case 1.** Safety/PK review meetings: visualize per-patient dose holds/reductions and recovery escalations, aiding attribution of AEs relative to exposure.
**– Use-case 2.** Operational oversight: check protocol adherence (e.g., maximum allowed escalations, mandatory reductions) and identify patterns that motivate schedule adjustments or supportive-care guidelines.
**• Visualization**

**Figure 15:**
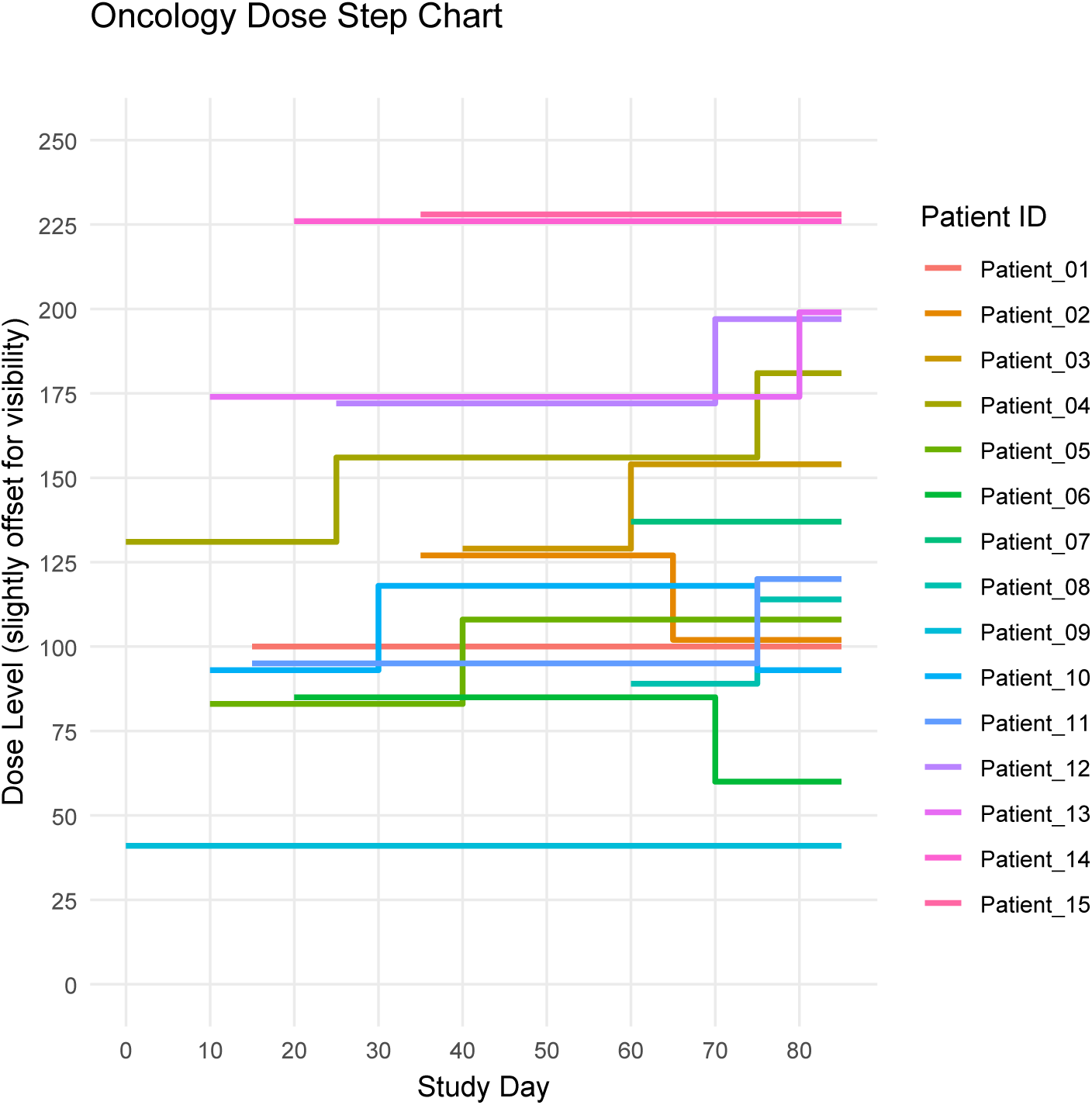
Step chart of dose level versus study day for 15 patients. Each line shows piecewise-constant dosing with modifications (reductions/escalations) at visit days. Small vertical offsets are applied solely to reduce overlap and do not represent dose differences.

#### 5.1.15 Calibration plot

**• Technical Specification**

**– Purpose:** Visualize how well a multinomial risk model’s predicted class probabilities agree with observed class frequencies (“calibration”) across the probability range for each outcome class. For perfect calibration, the curve for each class lies on the 45 degrees line [11].
**– Data:** Required variables: predicted probabilities for each class (e.g., p_benign, p_borderline, p_invasive summing to 1) and the observed class label for each individual.
**– Assumption:** Non-parametric calibration curves (LOESS/vector splines) adequately summarize the mean observed proportion as a smooth function of the predicted probability for each class. (Multinomial calibration can be built from a nominal logistic recalibration framework; non-parametric “vector spline” smoothers are a recommended option.)
**– R Packages:** dplyr, tidyr, ggplot2, stats
**• Applications**

**– Use-case 1.** For a three-class diagnostic model (benign/borderline/invasive) in ovarian tumor assessment, to evaluate whether predicted class probabilities are clinically trustworthy before deployment at a new hospital site; recalibrate if systematic miscalibration is observed.
**– Use-case 2.** For head-to-head comparison of competing multinomial models (e.g., MLR vs. machine-learning), to judge which model maintains better “weak calibration” on external validation when curves are compared to the 45 degrees line across the full probability range.
**• Visualization**

**Figure 16:**
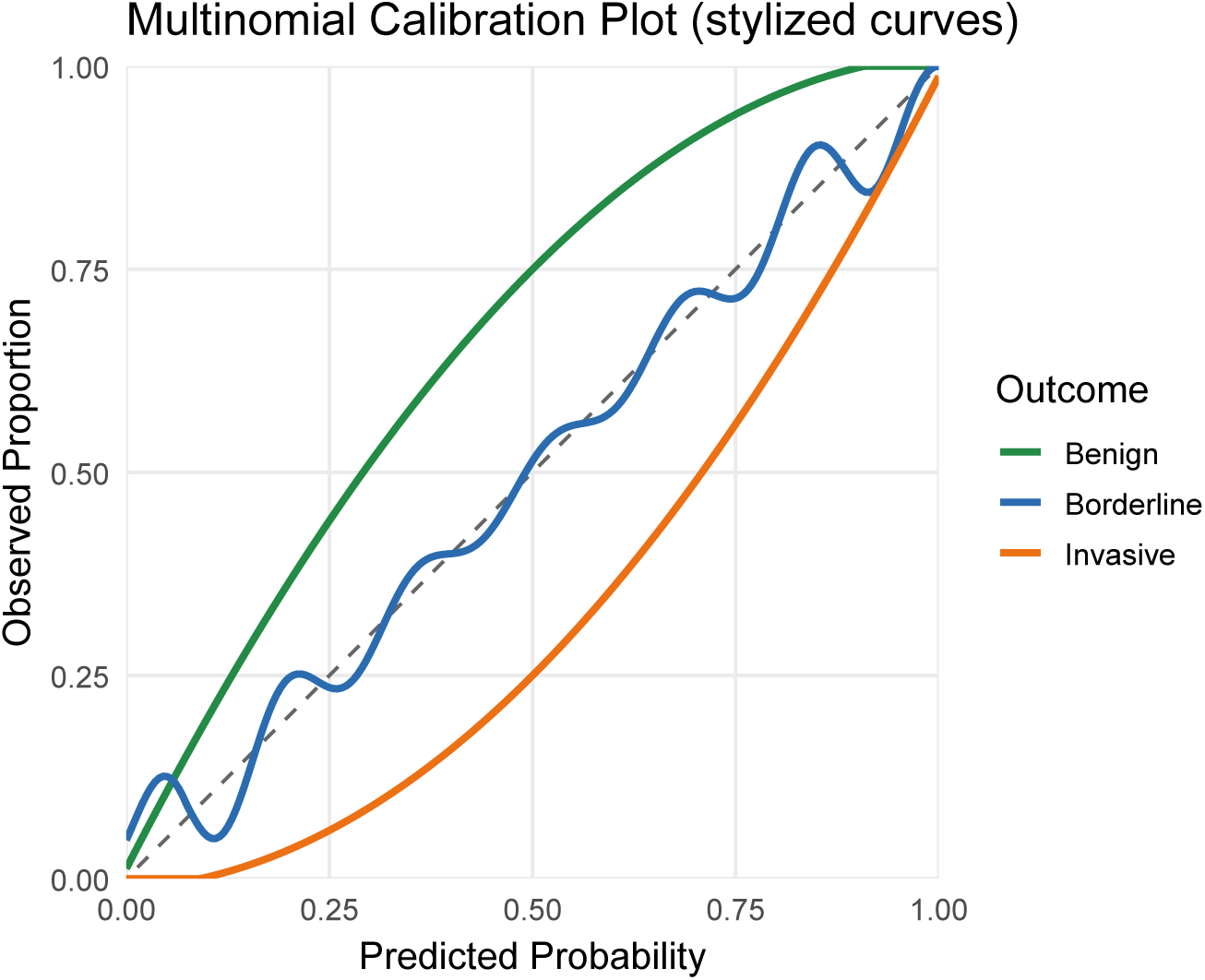
Multinomial calibration plot with smooth (LOESS) curves for benign (green), borderline (orange), and invasive (blue); the dashed 45 degrees line indicates perfect calibration. Curves above/below the diagonal indicate under-/over-prediction, respectively.

#### 5.1.16 Time-dependent ROC / AUC

**• Technical Specification**

**– Purpose:** Quantify how well a baseline biomarker predicts a time-to-event outcome at clinically relevant horizons by plotting Receiver operating characteristic *ROC*(*t*) and Area under the curve *AUC*(*t*) that explicitly account for censoring and the dynamic nature of events. In oncology, this is commonly used to evaluate whether a marker discriminates patients who will develop toxicity (or progress) by a fixed time t versus those event-free at t [161].
**– Data:** Required variables: patient-level marker at baseline (e.g., RILA %), event time, event indicator (1=event, 0=censored), and the prediction horizon(s) t (e.g., 12, 24, 36, 50 months).
**– Assumption:** Uses time-dependent ROC estimators for censored data (nearest-neighbor / KM, incident–or cumulative–dynamic formulations) so that cases/controls are defined with respect to time t and right-censoring is accommodated.
**– R Packages:** survivalROC or timeROC for ROC(t) and AUC(t); optional Cox model to build composite risk scores.
**• Applications**

**– Use-case 1.** For late RT fibrosis in early-stage breast cancer, estimate ROC(t) at 12/24/36/50 months and report AUC(t), optimal thresholds, sensitivity, specificity, PPV/NPV to guide thresholding decisions for dose adaptation.
**– Use-case 2.** For toxicity risk scores combining a biomarker with clinical covariates, compare AUC(t) of the biomarker alone versus a Cox-derived composite score across multiple horizons.
**• Visualization**

**Figure 17:**
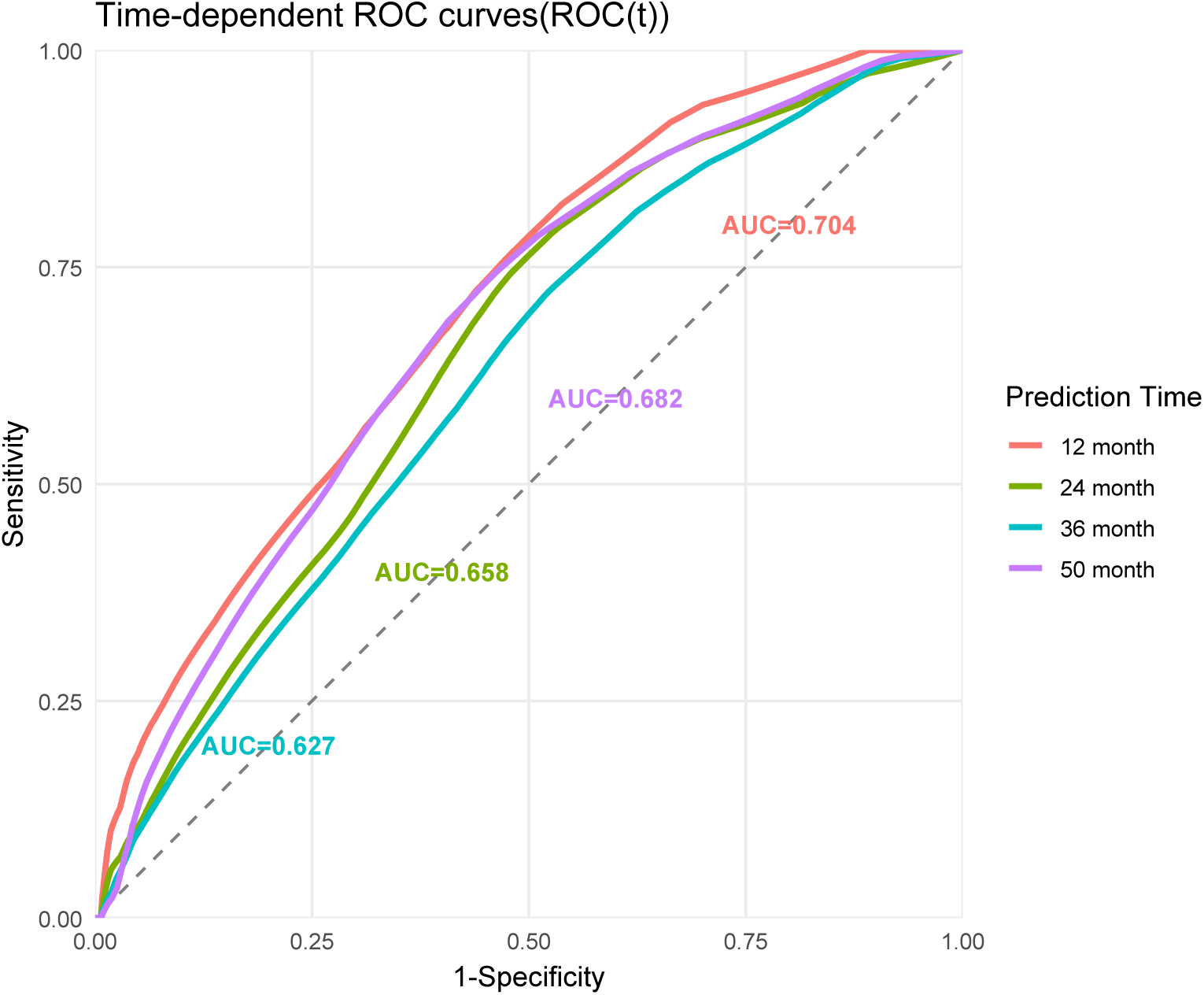
Time-dependent ROC curves, ROC(t), for a baseline biomarker predicting the event by 12, 24, 36, and 50 months; smooth ROC(t) estimates are shown for each horizon, and the grey dashed 45 degree line denotes no discrimination (random guess).

#### 5.1.17 Precision–Recall curve

**• Technical Specification**

**– Purpose:** Evaluate discriminative performance under class imbalance, where positive cases are rare—typical in oncology for outcomes like severe toxicity or a specific mutation. Precision Recall(PR) curves focus on the trade-off between Recall (Sensitivity/TPR) and Precision/Positive Predictive Value(PPV). They are often more informative than ROC curves when negatives far outnumber positives [162–164].
**– Data:** Required variables: binary outcome y (1=event, 0=non-event) and at least one continuous prediction score per model (higher → more likely positive).
**– Assumption:** Event prevalence is low; thresholds sweep from 0 → 1 to compute precision and recall without adding point glyphs.
**– R Packages:** precrec (PR/ROC computation and AUPRC), dplyr, ggplot2.
**• Applications**

**– Use-case 1.** Biomarker triage under rarity: when the endpoint is rare (e.g., severe immune-related AE), PR curves and Area under the precision–recall curve(AUPRC) provide a more faithful view of useful precision at clinically acceptable sensitivity than ROC/AUC.
**– Use-case 2.** Model selection and thresholding: compare candidate models (e.g., logistic vs. tree-based) and choose operating points that balance sensitivity with acceptable PPV for confirmatory testing or clinical alerting.
**• Visualization**

**Figure 18:**
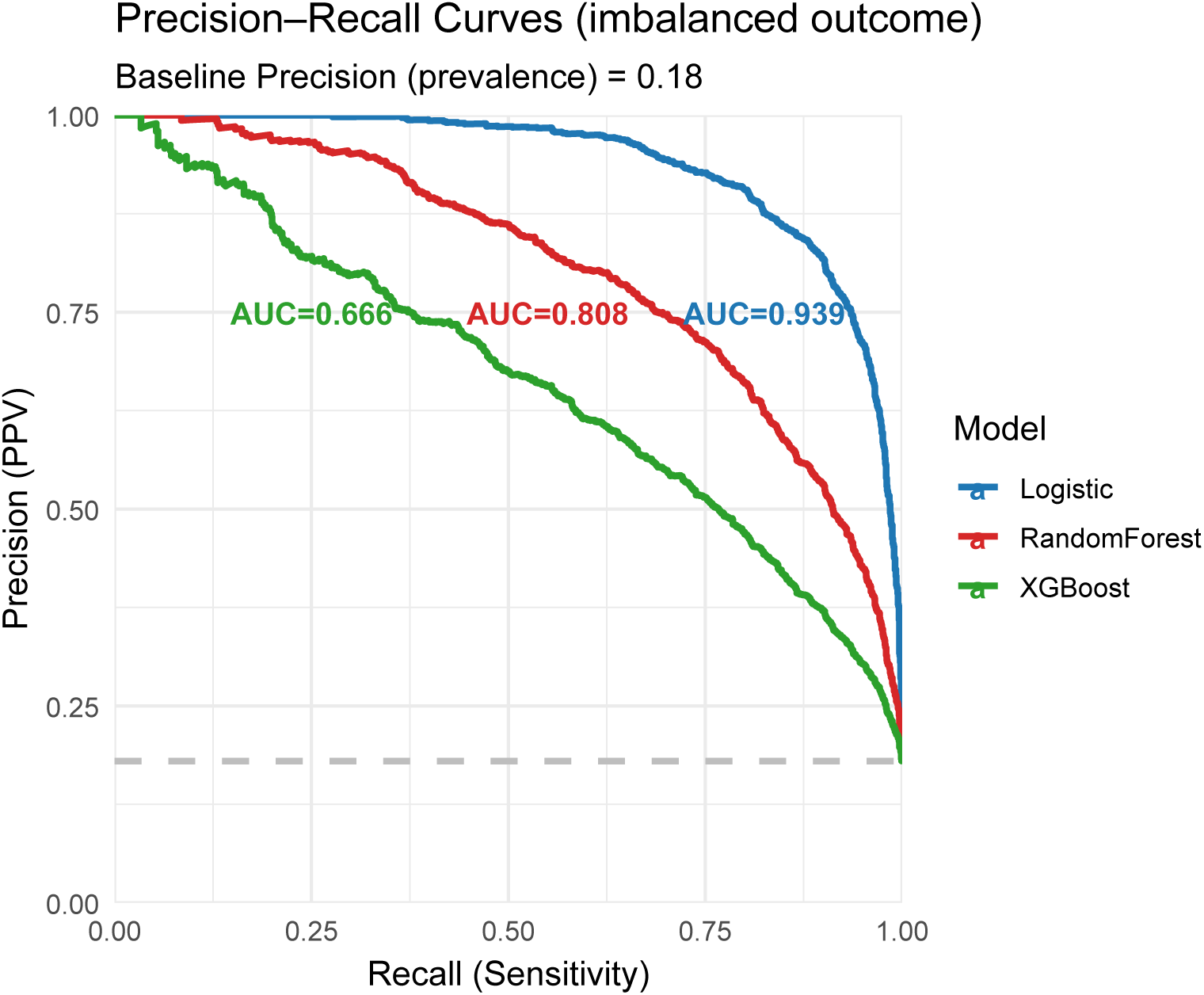
Precision–Recall curves for three predictive models under an imbalanced outcome (event prevalence =18%; dashed horizontal line). Curves are shown as smooth lines only; area under the PR curve (AUPRC) can be reported per model to compare performance.

#### 5.1.18 Decision Curve Analysis

**• Technical Specification**

**– Purpose:** Quantify the clinical utility of a risk model by plotting Net Benefit (NB) across threshold probabilities used to trigger an intervention (e.g., biopsy, adjuvant therapy). DCA compares a model against “treat none” and “treat all” [12, 165].
**– Data:** Binary outcome *y* (event/no event), a continuous predicted risk *p*^ per subject, and a clinically reasonable **threshold range** (e.g., 0–0.3).
**– Assumption:** Thresholds reflect a rational trade-off between benefit and harm; specifically, the decision-analytic identity *R* = *C/*(*B* + *C*) links threshold *R* to cost:benefit, and NB is NB(*R*) = TPR(*R*)*P* – 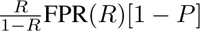 FPR(*R*)[1 − *P*].
**– R Packages:** Base R + dplyr, ggplot2 (custom NB implementation); or the authors’ package **Decision-Curve** for ready-made DCA utilities.
**• Applications**

**– Use-case 1.** Clinical policy selection. When there is no consensus threshold, inspect the DCA to find ranges of R where the model dominates both “treat all” and “treat none,” indicating positive clinical utility for adopting the model-guided strategy.
**– Use-case 2.** Subgroup strategy. If different patient strata merit different thresholds (e.g., frailty vs. fit), stratify the DCA and compare curves within each subpopulation rather than reading a single population curve at multiple thresholds.
**• Visualization**

**Figure 19:**
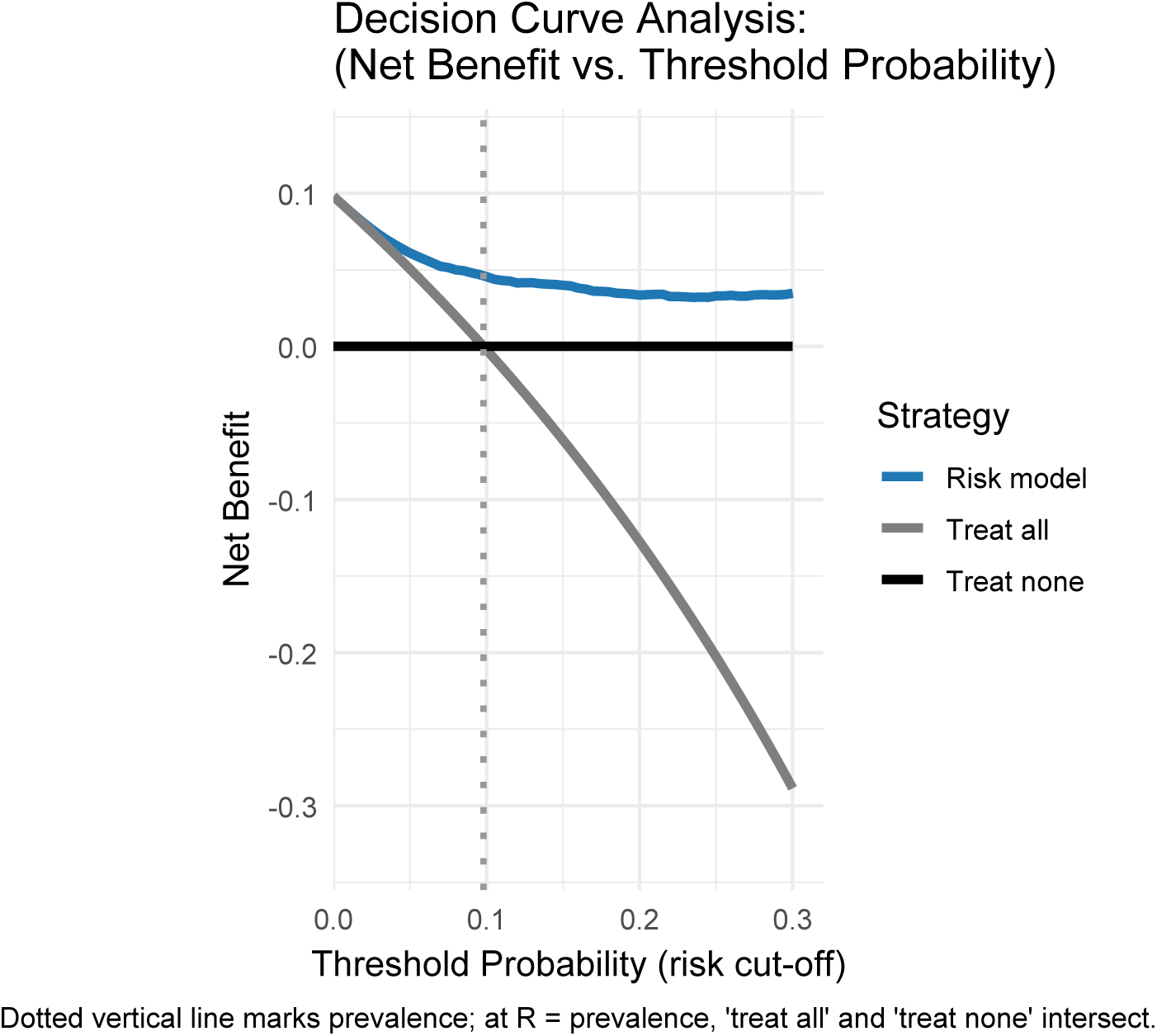
Decision curve analysis comparing a risk-based strategy with “treat all” and “treat none.” Net Benefit is plotted over thresholds 0–0.30; the dotted vertical line marks the event prevalence, where “treat all” intersects “treat none.”

#### 5.1.19 Nomogram

**• Technical Specification**

**– Purpose:** Provide an individualized, point-based prediction of 5-year recurrence-free survival (RFS) and show how well predictions agree with outcomes(calibration) [166, 167].
**– Data:** Patient-level predictors (e.g., tumor size in cm, mitotic index high/low, primary site) and a binary 5-year RFS outcome.
**– Assumption:** A logistic (or Cox) model summarizes the relationship between predictors and 5-year RFS; points are proportional to regression effects; calibration is assessed by grouping predictions and comparing observed vs predicted probabilities.
**– R Packages:** tidyverse, survival (optional), ggplot2, broom, binom.
**• Applications**

**– Use-case 1.** Pre-operative counseling: Convert patient characteristics to points and an RFS probability to inform shared decisions (e.g., surgery type, adjuvant therapy).
**– Use-case 2.** Trial eligibility/risk stratification: Use total points to stratify patients into risk groups for randomization or subgroup analyses.
**• Visualization**

**Figure 20:**
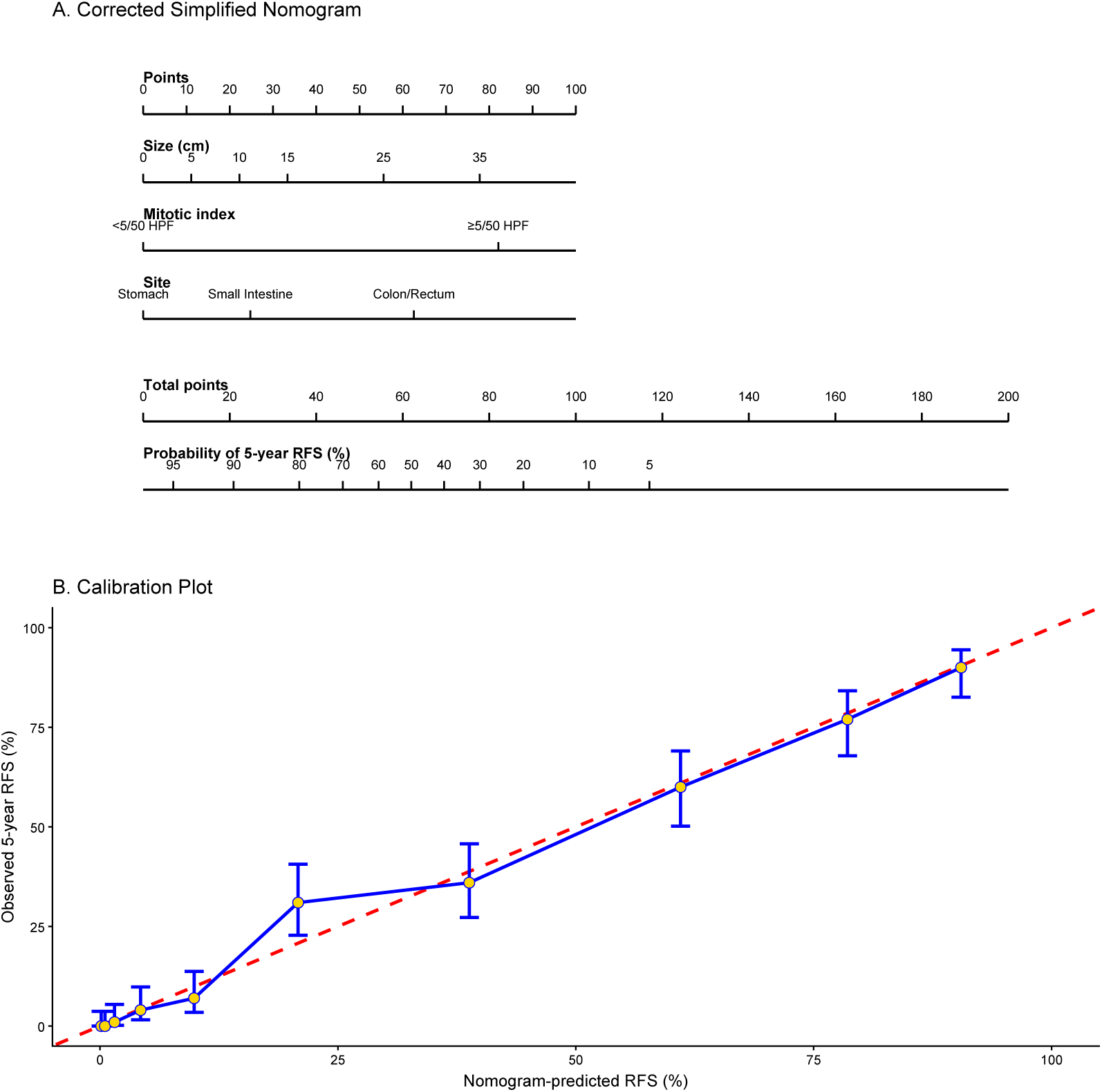
A Nomogram (A) and its calibration (B) for 5-year recurrence-free survival (RFS): the dashed 45 degree line denotes perfect calibration.

#### 5.1.20 Accrual over Time

**• Technical Specification**

**– Purpose:** This plot answers: “How is cumulative patient enrollment progressing over calendar time, and—given accrual to date—what is the expected future accrual with uncertainty?” [168–170]
**– Data:** Required variables: patient_id (unique subject); enroll_time (time since trial activation; e.g., days/weeks/months) Optional: site / region (if stratifying), screen_fail indicator (if tracking screening separately)
**– Assumption:** Accrual is generated by a counting process; for forecasting we assume a Poisson–Gamma (PG) recruitment model with a constant underlying accrual rate and a prespecified census time (time of interim review).
**– R Packages:** ggplot2, stats, dplyr.
**• Applications**

**– Use-case 1.** Recruitment monitoring. For an oncology trial, to detect whether observed enrollment is tracking below expectations and trigger mitigation (activate additional sites, broaden eligibility, extend recruitment).
**– Use-case 2.** Forecasting operational timelines. For a DMC/steering committee update, to estimate the expected time to reach the target sample size and quantify uncertainty under a PG recruitment model.
**• Visualization**

**Figure 21:**
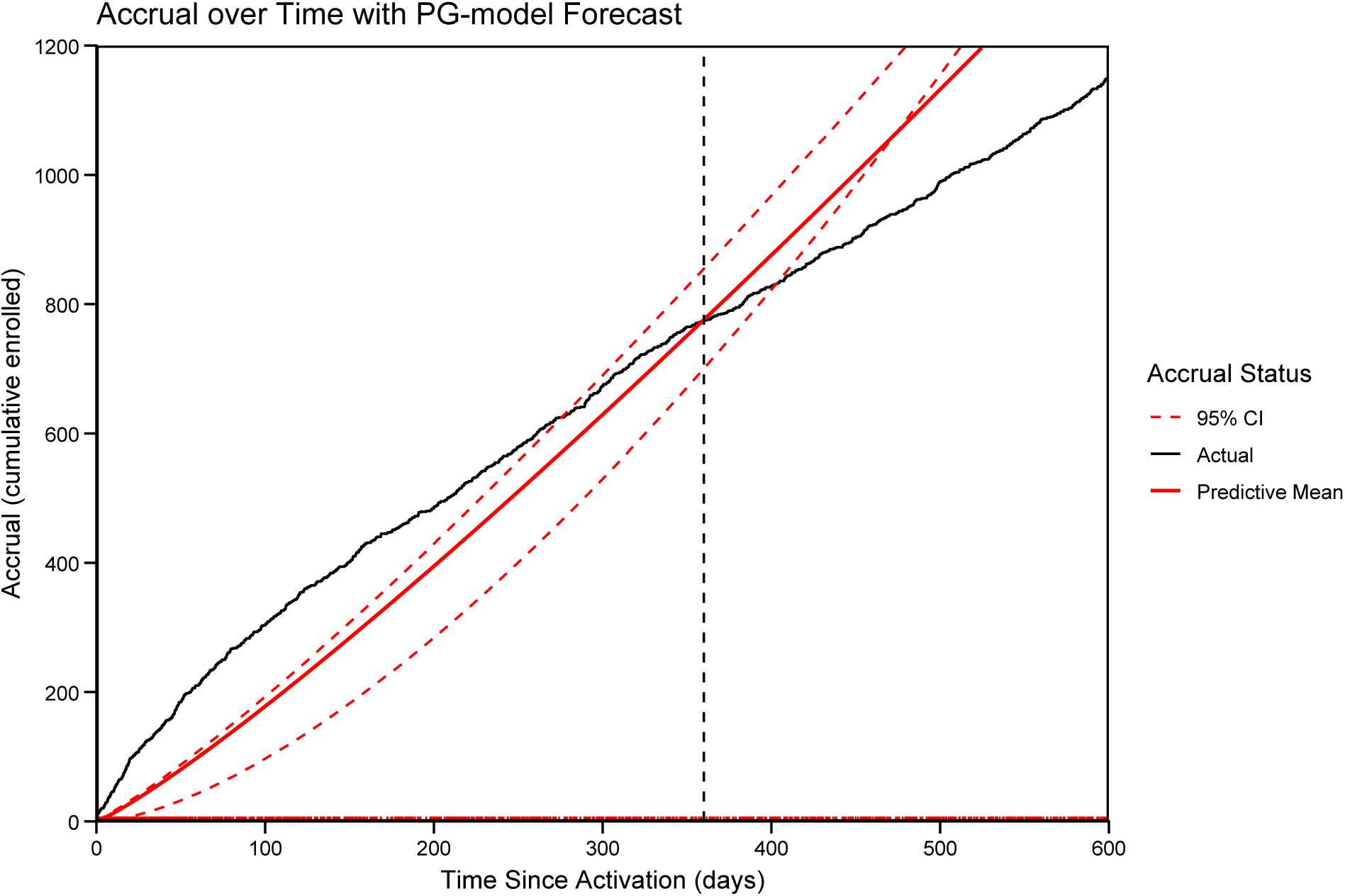
Accrual (black, solid; smoothed) together with the Poisson–Gamma (PG) model predictive mean (red, solid) and 95% prediction bands (red, dashed). The census time (interim look) is indicated by the vertical dashed line.

#### 5.1.21 Group-Sequential Boundaries / Alpha Spending

**• Technical Specification**

**– Purpose:** This plot answers how interim stopping thresholds (Z-boundaries) and cumulative Type-I error spent (*α*-spending) evolve as information accrues under different group-sequential designs [171–173].
**– Data:** info_frac (information fraction, 0–1), design (e.g., O’Brien–Fleming / Pocock / Haybittle–Peto), and either z_crit (critical Z) and/or alpha_cum (cumulative alpha spent).
**– Assumption:** Pre-specified interim looks with known information fractions; one-sided efficacy testing with total *α* allocated via a chosen spending rule (or an approximation to it).
**– R Packages:** rpact, ggplot2, patchwork.
**• Applications**

**– Use-case 1.** For a time-to-event oncology RCT with interim OS/PFS analyses, to choose an interim monitoring approach (e.g., O’Brien–Fleming when you want strong early conservatism but near-final ≈ 0.05 behavior).
**– Use-case 2.** For a DMC charter / SAP, to communicate operating rules (how hard it is to stop early for efficacy, and how *α* is preserved for the final analysis) in a single, audit-friendly graphic.
**• Visualization**

**Figure 22:**
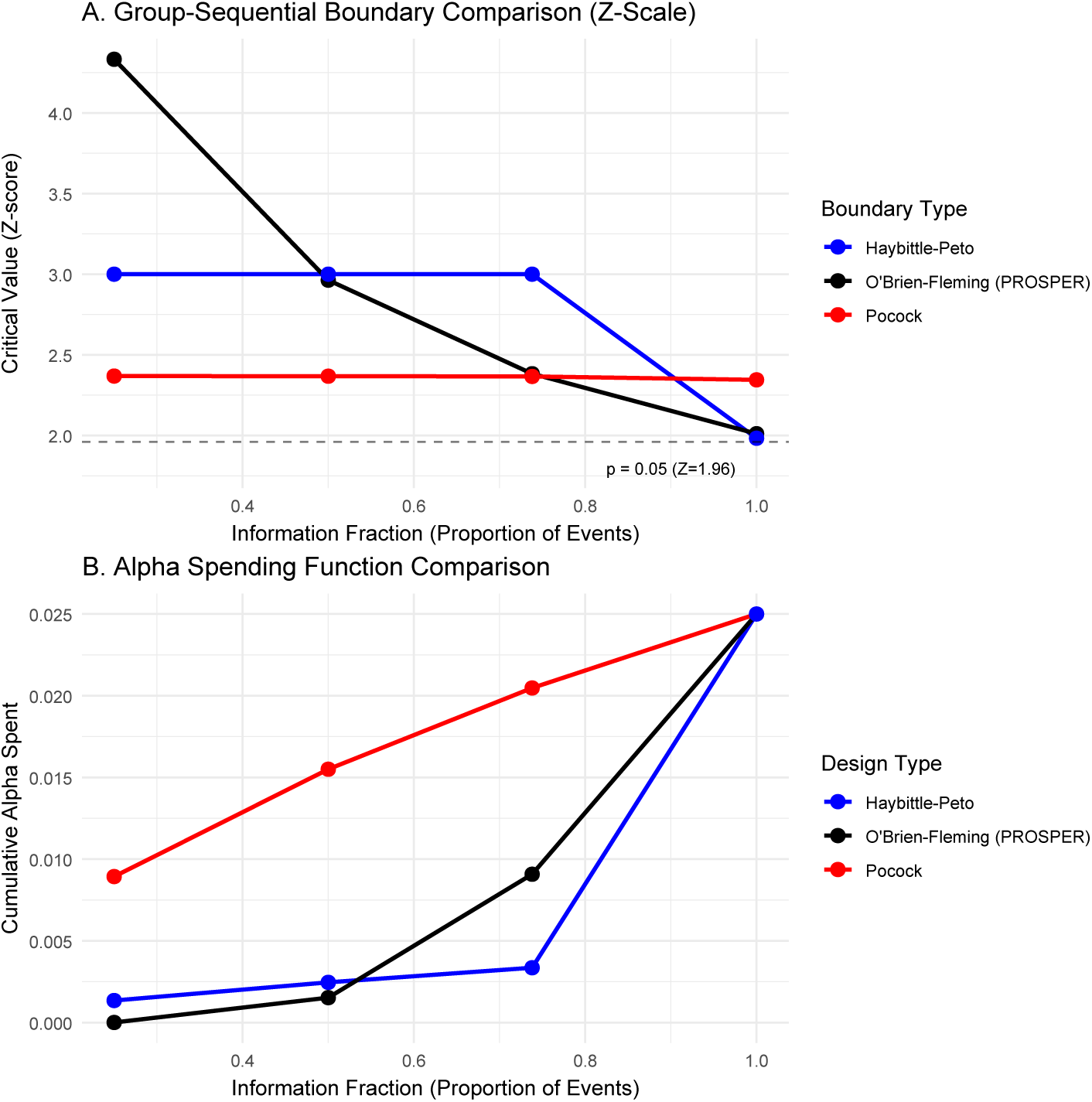
Group-sequential boundaries and alpha-spending profiles across interim looks. Panel A compares one-sided efficacy Z-boundaries over information fraction for three common designs (Haybittle–Peto, O’Brien–Fleming, Pocock) with total *α*= 0.025. Panel B displays the corresponding cumulative *α* spent over time using a nominal-alpha proxy implied by the interim critical values; conservative early-stopping designs spend less *α* early and reserve more for later analyses.

#### 5.1.22 Bayesian Predictive Probability Plot

**• Technical Specification**

**– Purpose:** Visualize how the predictive probability of final trial success evolves over accruing sample size to support interim go/no-go decisions (futility vs efficacy) [174, 175].
**– Data:** n (interim sample size / information time), PP (predictive probability of success at final), optional x (cumulative responders or interim effect summary), and decision thresholds (e.g., PP_futility, PP_efficacy).
**– Assumption:** Bayesian predictive probability is computed under a pre-specified Bayesian model (likelihood + prior) and a pre-defined success criterion at the planned final analysis.
**– R Packages:** ggplot2, patchwork, dplyr, scales.
**• Applications**

**– Use-case 1.** For an adaptive Phase II oncology trial, to support interim futility stopping when PP drops below a pre-specified threshold (e.g., 0.10).
**– Use-case 2.** For a platform trial decision workflow (I-SPY2-style), to support graduation/efficacy when PP exceeds a high threshold (e.g., 0.90), prioritizing arms for confirmatory testing.
**• Visualization**

**Figure 23:**
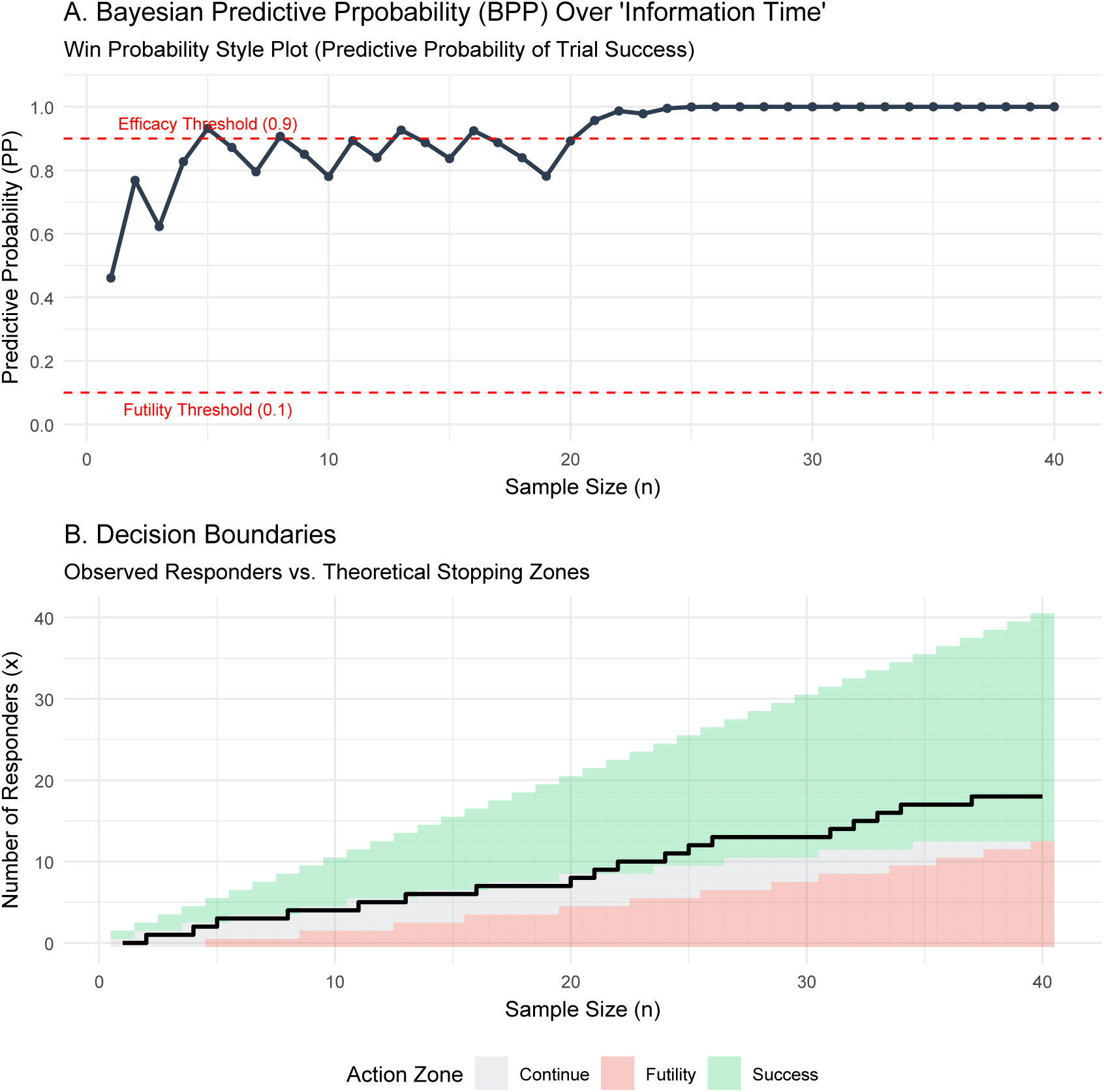
Bayesian predictive probability (PP) monitoring plot for an adaptive oncology trial.

#### 5.1.23 Operating Characteristics Curves beyond Power

**• Technical Specification**

**– Purpose:** Visualize and compare dose-finding accuracy across multiple true-toxicity scenarios by plotting the percentage of correct Maximum tolerated dose(MTD) selection for competing Phase I designs (e.g., 3+3 vs model-assisted designs) [176–178].
**– Data:** scenario (1–16), target_dlt (e.g., 0.25, 0.30), design (3+3, mTPI, BOIN), pct_correct (0–100).
**– Assumption:** Percentages come from Monte Carlo operating-characteristics simulations under prespecified scenario toxicity curves and fixed design parameters (target DLT, cohort rules, stopping rules).
**– R Packages:** ggplot2.
**• Applications**

**– Use-case 1.** In Phase I dose-finding design selection, compare candidate designs’ MTD selection accuracy under sponsor-defined scenario libraries (optimistic, pessimistic, and misspecified toxicity shapes).
**– Use-case 2.** In protocol design discussions, justify choosing a model-assisted design (e.g., BOIN/mTPI) over 3+3 by summarizing accuracy trade-offs at the trial’s target DLT rate(s).
**• Visualization**

**Figure 24:**
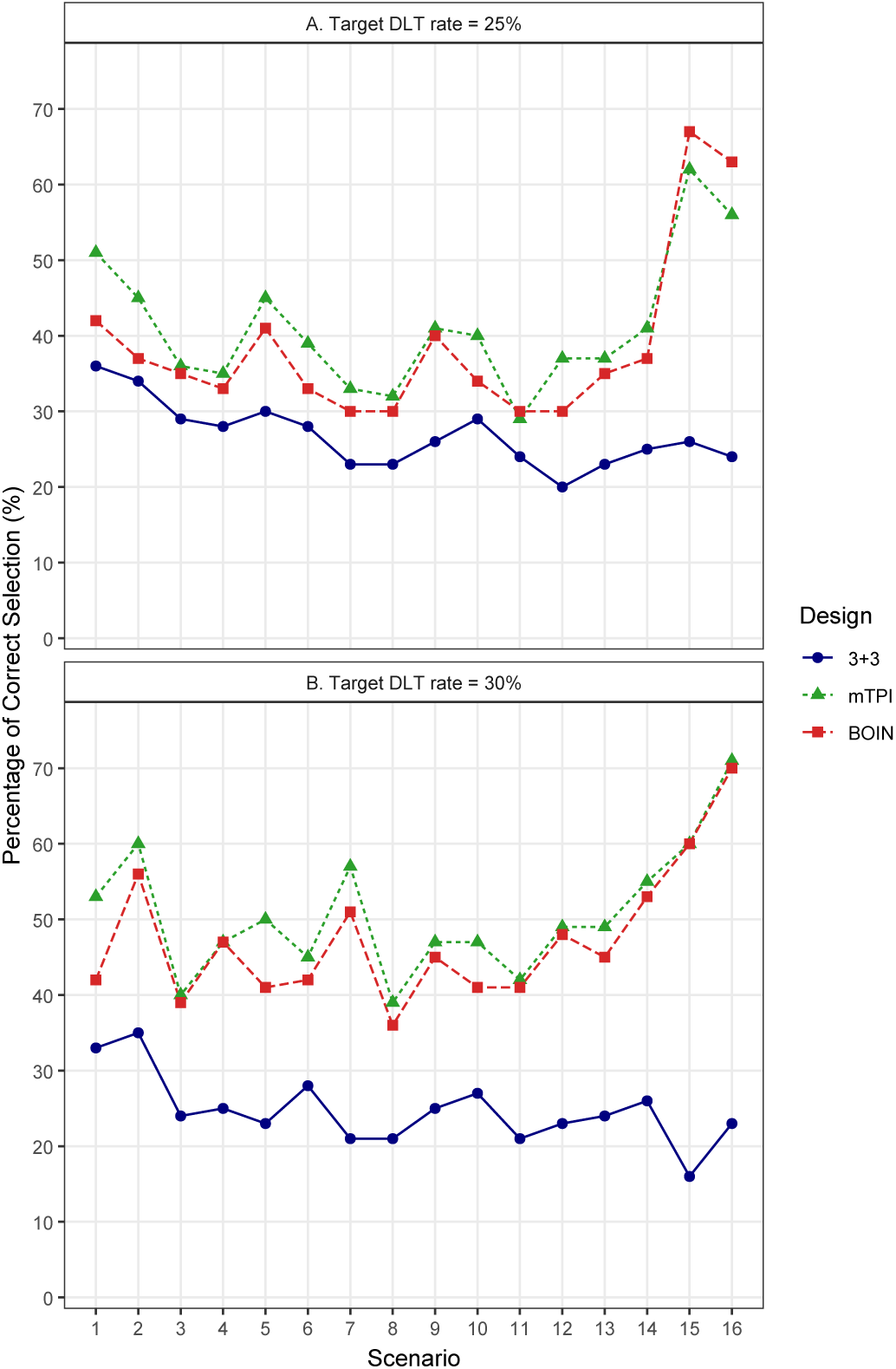
Percentage of correct selection of the MTD under the 3+3, mTPI and BOIN designs across 16 scenarios, separately for target DLT rates of 25% and 30%. Higher is better.

#### 5.1.24 Time-Varying Covariate (TVC) Survival Curves

**• Technical Specification**

**– Purpose:** Visualize how survival differs between groups defined by a time-varying covariate (TVC), and show how naïve KM and landmark KM can differ from time-dependent Cox–based curves (Smith–Zee / Extended KM) when the covariate changes after baseline [179, 180].
**– Data:** id, time, status (event indicator), xt_change_time (time TVC switches 0 → 1; NA if never), plus a chosen landmark_time.
**– Assumption:** Independent censoring; TVC change times are measured without error; TD Cox assumes proportional hazards given the TVC process; landmark analysis is conditional on being event-free at the landmark and depends on landmark choice.
**– R Packages:** [1] Comprehensive List built in Shiny app: https://samplen.shinyapps.io/TVCurve/ [2] Partial List: survival, ggplot2, patchwork.
**• Applications**

**– Use-case 1.** In CAR-T studies evaluating post-infusion interventions (e.g., HCT as a TVC), compare naïve/landmark curves versus TD Cox–based curves to detect and communicate immortal-time bias and landmark sensitivity.
**– Use-case 2.** In oncology RWE where treatment switching or rescue therapy occurs after baseline, use Smith–Zee / Extended KM to present clinically interpretable survival differences while respecting time-varying exposure definitions.
**• Visualization**

**Figure 25:**
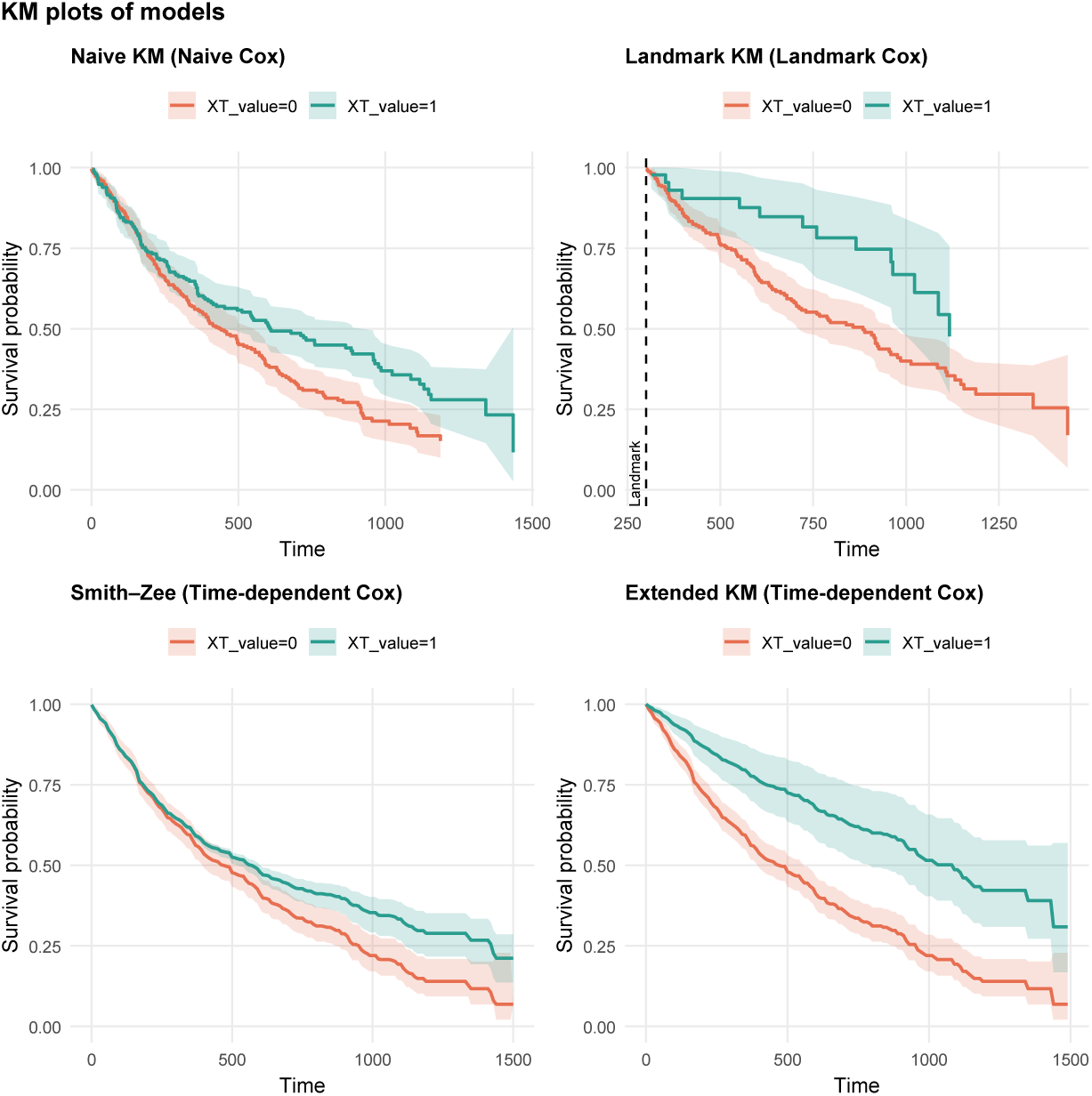
Time-varying covariate survival curves comparing Naïve KM, Landmark KM, and two time-dependent Cox model visualizations (Smith–Zee and Extended KM) for XT_value (0 vs 1). Naïve and landmark approaches may be biased or sensitive to the landmark choice when covariate status changes after baseline, whereas TD Cox–based curves accommodate covariate changes over time and provide standardized visualizations

### 5.2 Category B: Real World Evidence (RWE)

#### 5.2.1 Love plot (covariate balance plot after PSM / IPTW)

**• Technical Specification**

**– Purpose:** Diagnose covariate balance between treatment groups before vs after adjustment (typically Propensity score matching(PSM) or Inverse probability of treatment weighting(IPTW)) using absolute standardized mean differences (ASMD/SMD) [181–183].
**– Data:** Covariate name (covariate), Balance metric pre-adjustment (asmd_unadjusted), Balance metric post-adjustment (asmd_adjusted): (Optional) cohort/arm indicator (e.g., Arm 2: neoadjuvant systemic therapy cohort)
**– Assumption:** After PSM/IPTW, ASMD *<* 0.10 is commonly interpreted as adequate balance for that covariate.
**– R Packages:** ggplot2 (core), optionally dplyr for ordering.
**• Applications**

**– Use-case 1.** RWE comparative effectiveness (oncology): Show that RT vs no-RT cohorts are comparable on measured confounders after PSM/IPTW before estimating OS/RFS effects.
**– Use-case 2.** Model diagnostics: Identify which covariates remain imbalanced (ASMD ≥ 0.10) and require re-specifying the PS model, adding interactions/splines, trimming, or switching to weighting.
**• Visualization**

**Figure 26:**
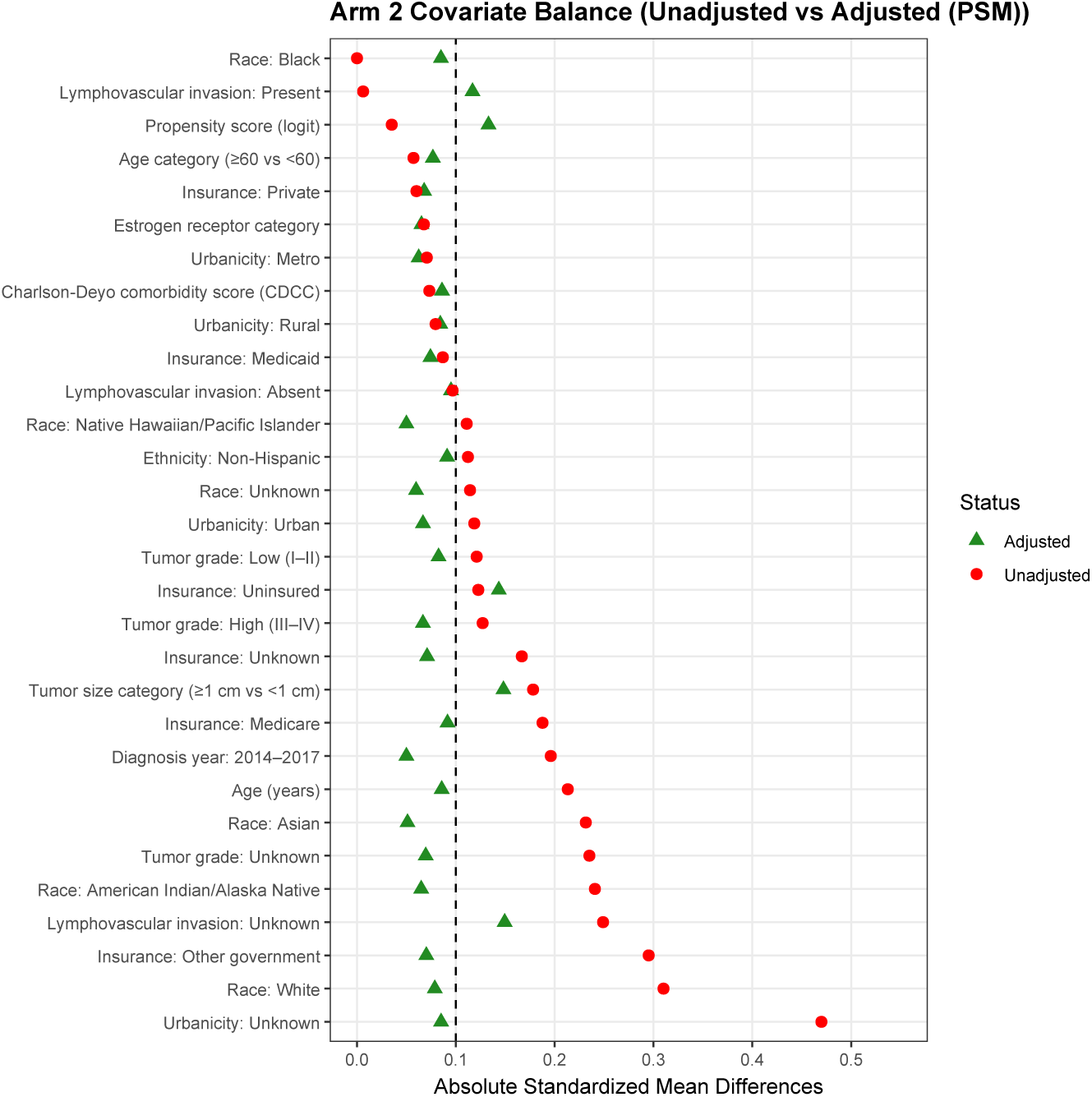
Covariate balance using propensity score matching. Love plot depicting absolute standardized mean differences (ASMDs) for baseline covariates comparing the radiation therapy (RT) and no-RT groups before adjustment (unadjusted) and after propensity score matching (adjusted). The vertical dashed line indicates the balance threshold (ASMD = 0.10); values below this threshold are generally interpreted as indicating adequate post-matching balance between treatment groups.

#### 5.2.2 Subgroup Analysis

**• Technical Specification**

**– Purpose:** Display treatment-effect heterogeneity across clinically relevant subgroups by plotting hazard ratios (HRs) (with 95% CIs) from a Cox model for overall survival (OS) [184, 185]
**– Data:** subgroup, level, events, patients, HR, LCL, UCL (and optionally an ordering variable).
**– Assumption:** The subgroup HRs are interpretable under the Cox proportional hazards framework; subgroup estimates are typically exploratory (multiplicity and power constraints).
**– R Packages:** ggplot2, dplyr
**• Applications**

**– Use-case 1.** Confirm consistency of treatment benefit across clinically important strata (age, ECOG, PD-L1 TPS, brain metastases).
**– Use-case 2.** Pre-specify effect-modifier hypotheses (e.g., PD-L1 strata) and visually check whether subgroup CIs plausibly overlap.
**• Visualization**

**Figure 27:**
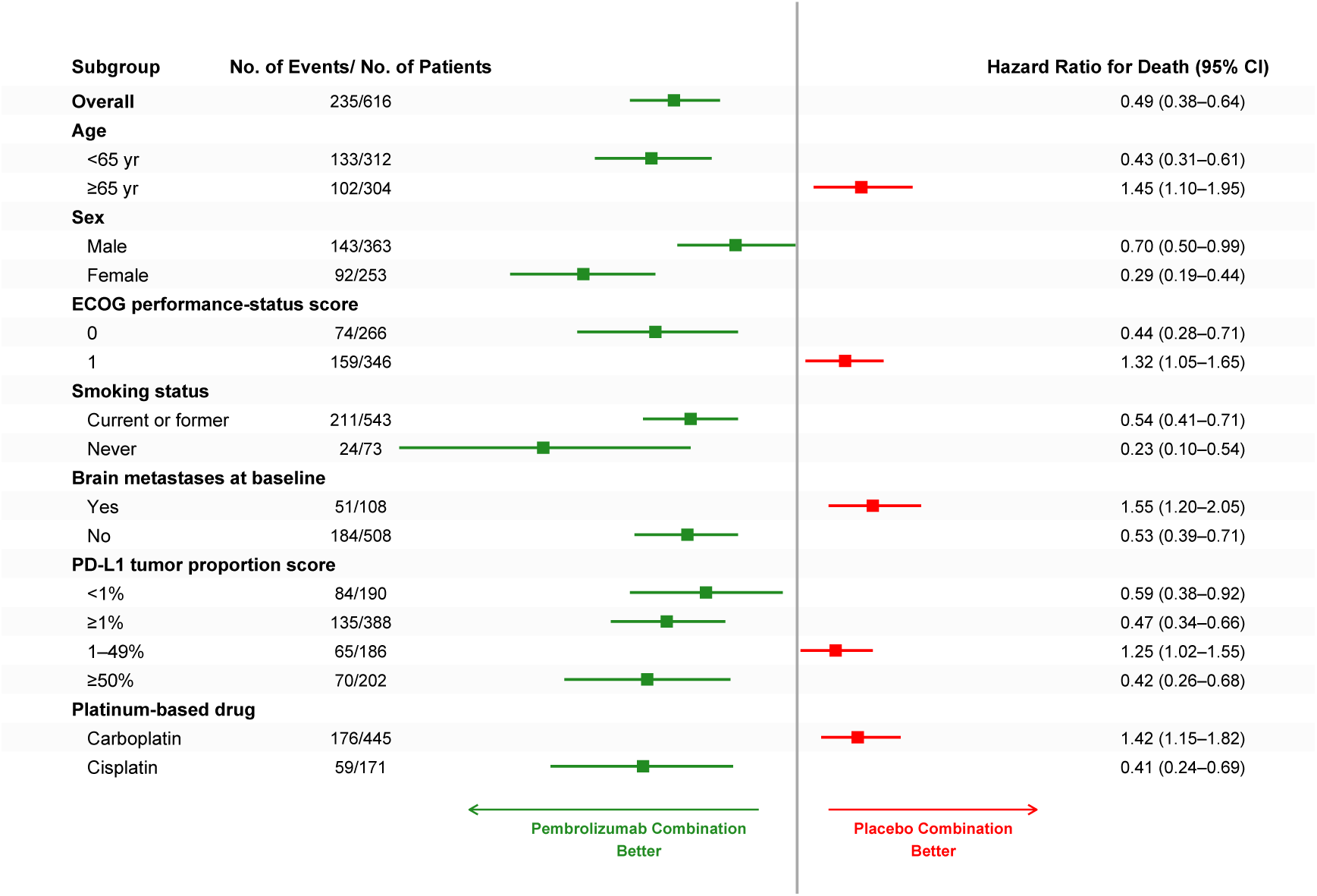
Subgroup analysis of overall survival (OS) in metastatic nonsquamous NSCLC (KEYNOTE-189). Squares denote subgroup hazard ratios (HRs) and horizontal lines denote 95% confidence intervals from stratified Cox models comparing pembrolizumab + pemetrexed + platinum vs placebo + pemetrexed + platinum. The vertical dashed line marks *HR* = 1.0; values *<* 1 favor pembrolizumab-combination therapy. Event counts are shown as events/patients for each subgroup.

#### 5.2.3 Value of Operation–Prob of Cost Efficacy

**• Technical Specification**

**– Purpose:** Visualizes, across willingness-to-pay (WTP) thresholds, the probability that each strategy is cost-effective (i.e., yields the highest net monetary benefit, NMB) [186, 187].
**– Data:** Probabilistic sensitivity analysis (PSA) draws with cost and Quality-adjusted life year(QALY) per strategy (per simulation), plus a WTP grid (e.g., $0–$200,000 per QALY).
**– Assumption:** Decision rule = choose strategy with max NMB at each WTP, under the PSA joint uncertainty.
**– R Packages:** dplyr, tidyr; ggplot2; base stats::splinefun for smooth curves.
**• Applications**

**– Use-case 1.** In oncology Health economics and outcomes research(HEOR) submissions, report decision uncertainty by showing how the preferred strategy changes as WTP varies (payer-specific thresholds).
**– Use-case 2.** Compare alternative regimens (e.g., chemo vs IO-combo) under PSA to quantify probability of being cost-effective rather than only Incremental cost-effectiveness ratio(ICER) point estimates.
**• Visualization**

**Figure 28:**
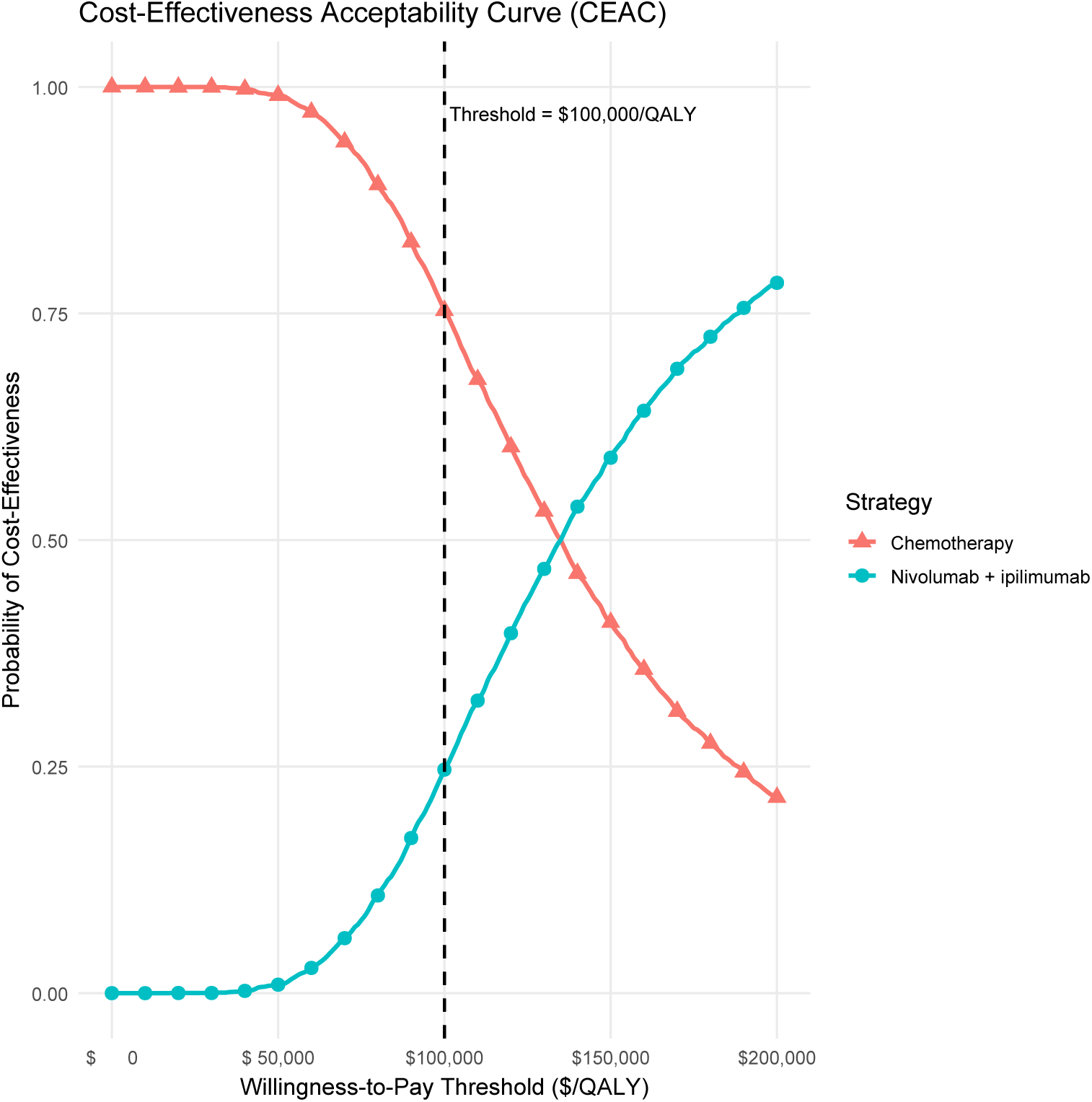
Cost-effectivenes acceptability curves (CEACs)”, “comparing nivolumab plus ipilimumab versus chemotherapy. The curves show,”, “across willingness-to-pay thresholds, the probability that each strategy is”, “cost-effective (highest net monetary benefit) under probabilistic sensitivity analysis.”, “The vertical dashed line marks the reference threshold of $100,000 per QALY.”

#### 5.2.4 Tornado Diagram

**• Technical Specification**

**– Purpose:** Visualize which single input parameters drive the largest swing in a base-case economic result (typically ICER or Net Monetary Benefit) in an oncology HEOR/RWE analysis [186, 187].
**– Data:** parameter, min_impact, max_impact (impact on outcome vs base-case, often as % change or absolute change), plus optionally base_case (usually 0 after centering).
**– Assumption:** One-way sensitivity—vary one parameter at a time while holding all other parameters fixed at base-case values.
**– R Packages:** ggplot2, dplyr, tidyr(optional).
**• Applications**

**– Use-case 1.** In an oncology cost-effectiveness model, identify whether drug cost, utilities, or transition hazards dominate uncertainty in ICER/NMB.
**– Use-case 2.** In RWE-informed HEOR, justify scenario refinement (e.g., better AE costing) by showing which real-world inputs most change the conclusion at a given WTP threshold.
**• Visualization**

**Figure 29:**
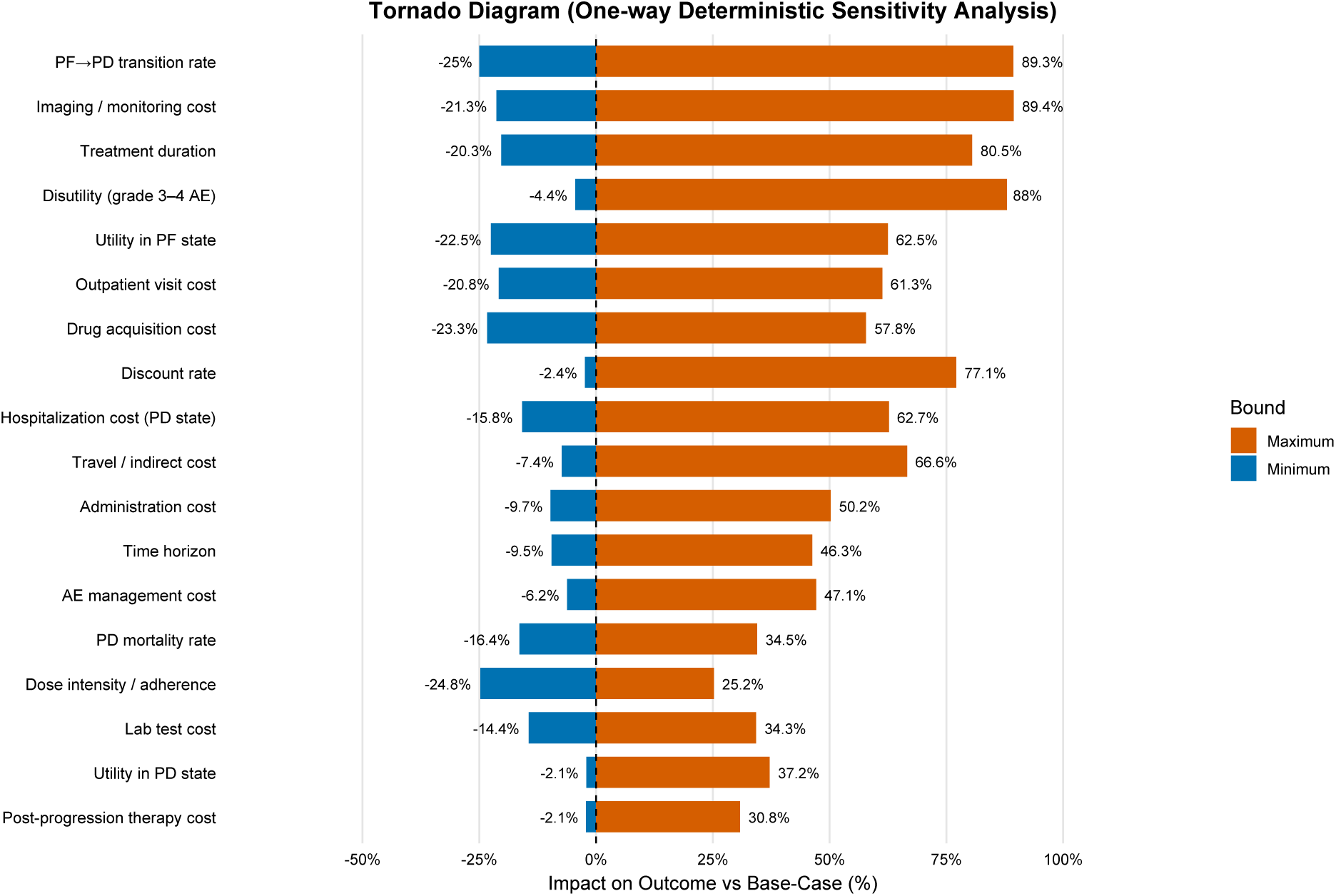
One-way deterministic sensitivity analysis (tornado diagram). Horizontal bars show the change in the model outcome (e.g., ICER) from the base-case (0, dashed line) when each parameter is varied individually to its minimum and maximum values, holding all other inputs fixed. Parameters are ordered by total swing (|min| + |max|), highlighting the most influential drivers.

#### 5.2.5 Sankey/Alluvial Diagram

**• Technical Specification**

**– Purpose:** This plot answers “How do patients flow through sequential lines of therapy (1*L* → 2*L* → 3*L*) and terminal outcomes (death/censored)?” [188, 189]
**– Data:** patient_id, lot (1L/2L/3L), regimen (per LOT), terminal_status (Death/Censored), plus optional followup_start, followup_end.
**– Assumption:** Each patient occupies exactly one state per LOT, with monotone progression (no backtracking).
**– R Packages:** networkD3, dplyr, tidyr htmlwidgets, webshot2.
**• Applications**

**– Use-case 1.** In oncology RWE, summarize treatment sequencing (1*L* → 2*L* → 3*L*+) and quantify where attrition occurs (death/censoring) to contextualize survival comparisons.
**– Use-case 2.** Identify dominant and rare pathways to motivate subgroup definitions (e.g., “pembro-combo in 1L then chemo in 2L”) for downstream effectiveness/safety analyses.
**• Visualization**

**Figure 30:**
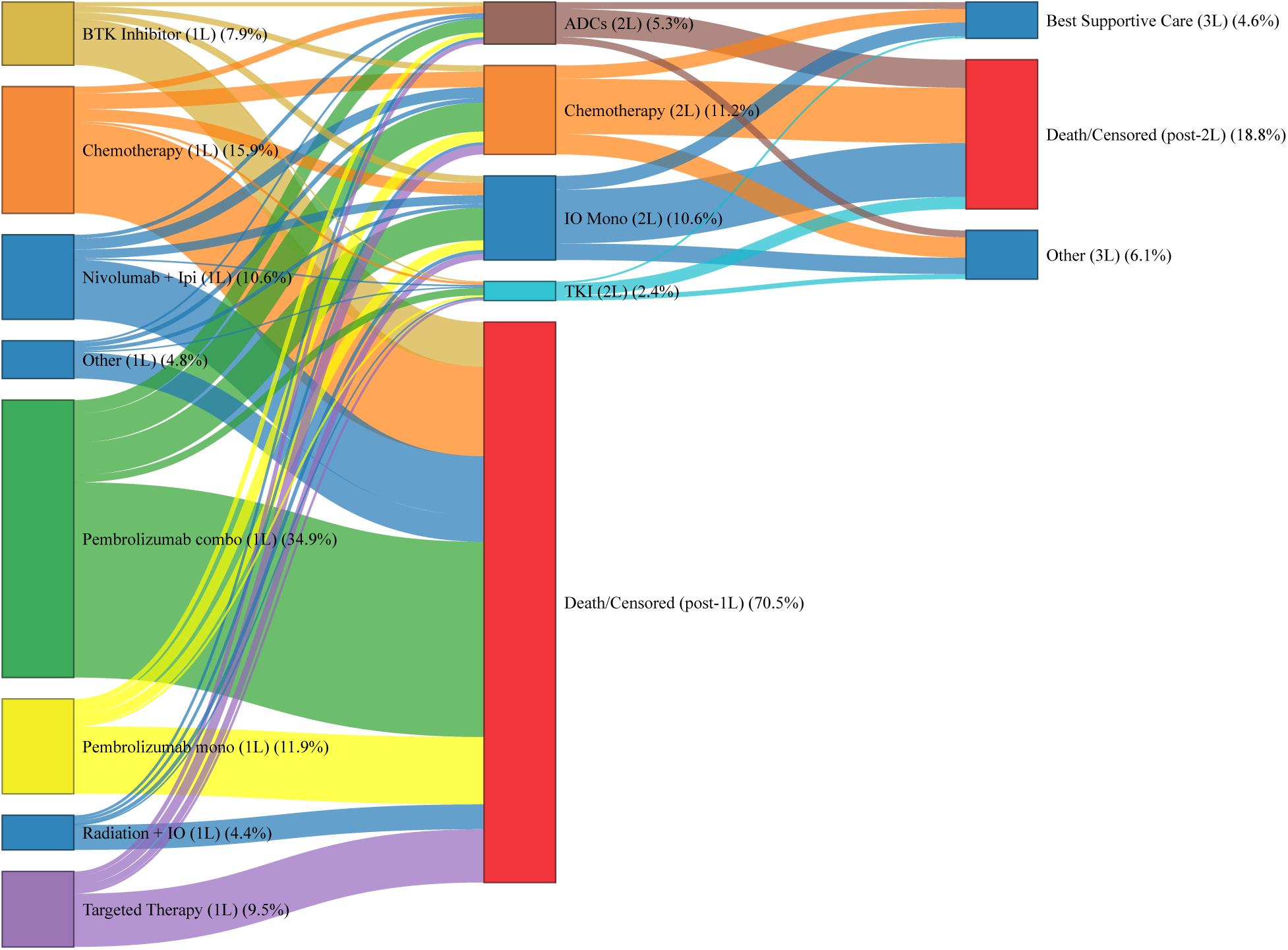
Sankey diagram of sequential treatment patterns during follow-up (simulated example). Node widths represent the number (or %) of patients receiving each line of therapy and terminal outcomes (death or censoring). Link widths are proportional to the number of patients transitioning between states, enabling rapid identification of dominant treatment pathways and attrition across lines of therapy.

#### 5.2.6 Chord Diagram

**• Technical Specification**

**– Purpose:** Visualize switching / transitions between categorical oncology states (e.g., lines of therapy or regimens) by encoding the volume of flow between categories as chord widths [190, 191].
**– Data:** Long-format transitions or a square transition matrix with variables such as from, to, n (counts) or M[from,to] = n.
**– Assumption:** Transitions are correctly defined (one row per switch) and categories are mutually exclusive at each transition step.
**– R Packages:** circlize::chordDiagram() (+ optional dplyr for wrangling).
**• Applications**

**– Use-case 1.**Treatment sequencing & switching (RWE): Summarize how patients move from 1L to 2L/3L regimens and identify dominant pathways (e.g., chemo → BTKi/venetoclax-like classes).
**– Use-case 2.** Care pathways / healthcare setting flows: Visualize transitions between treatment settings (community → academic center) or states (on-treatment → off-treatment → censor/death if included as nodes).
**• Visualization**

**Figure 31:**
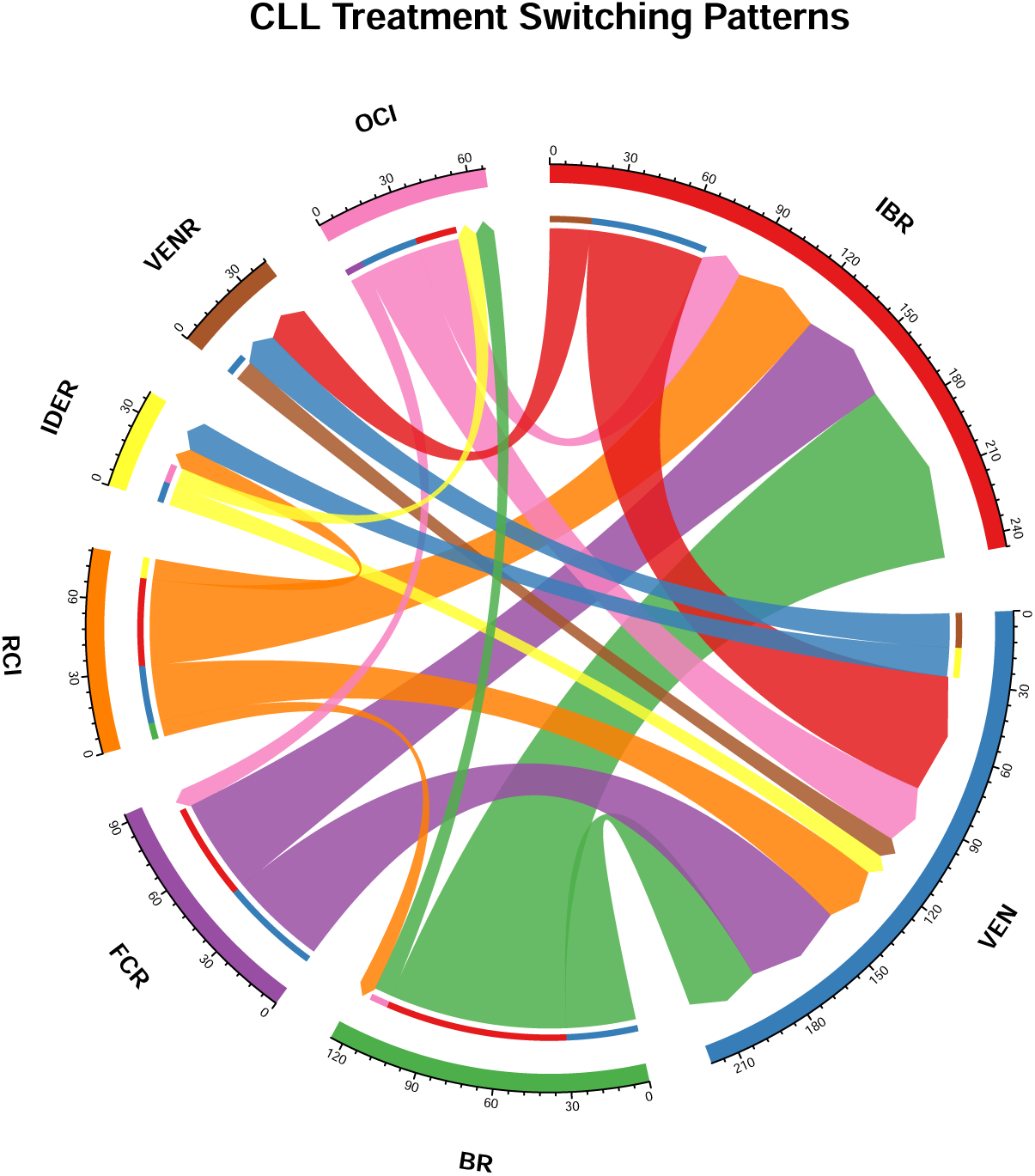
Chord diagram of treatment switches in a real-world oncology cohort. Each perimeter segment denotes a treatment regimen; ribbon width is proportional to the number of patients switching from the source regimen to the destination regimen(IBR = Ibrutinib; VEN = Venetoclax; BR = Bendamustine–Rituximab; FCR = Fludarabine–Cyclophosphamide–Rituximab; RCI = Rituximab–Chlorambucil–Idelalisib; IDER = Idelalisib–Dexamethasone–Rituximab; VENR = Venetoclax–Rituximab; OCI = Obinutuzumab–Chlorambucil–Ibrutinib.).

#### 5.2.7 Spaghetti Plot

**• Technical Specification**

**– Purpose:** Visualize individual-patient longitudinal trajectories of tumor burden (e.g., SLD) over follow-up time to assess heterogeneity in response patterns [192].
**– Data:** patient_id, week (or visit time), sld_mm (Sum of Lesion Diameters(SLD), mm).
**– Assumption:** Repeated measures per patient are time-ordered and comparable (same measurement definition across visits).
**– R Packages:** ggplot2.
**• Applications**

**– Use-case 1.** Baseline heterogeneity: Shows how patients start from very different absolute tumor burdens (important in RWE and heterogeneous trial populations).
**– Use-case 2.** Growth/shrinkage dynamics: Supports qualitative assessment of response patterns (shrinkage, regrowth, stability) before formal modeling.
**• Visualization**

**Figure 32:**
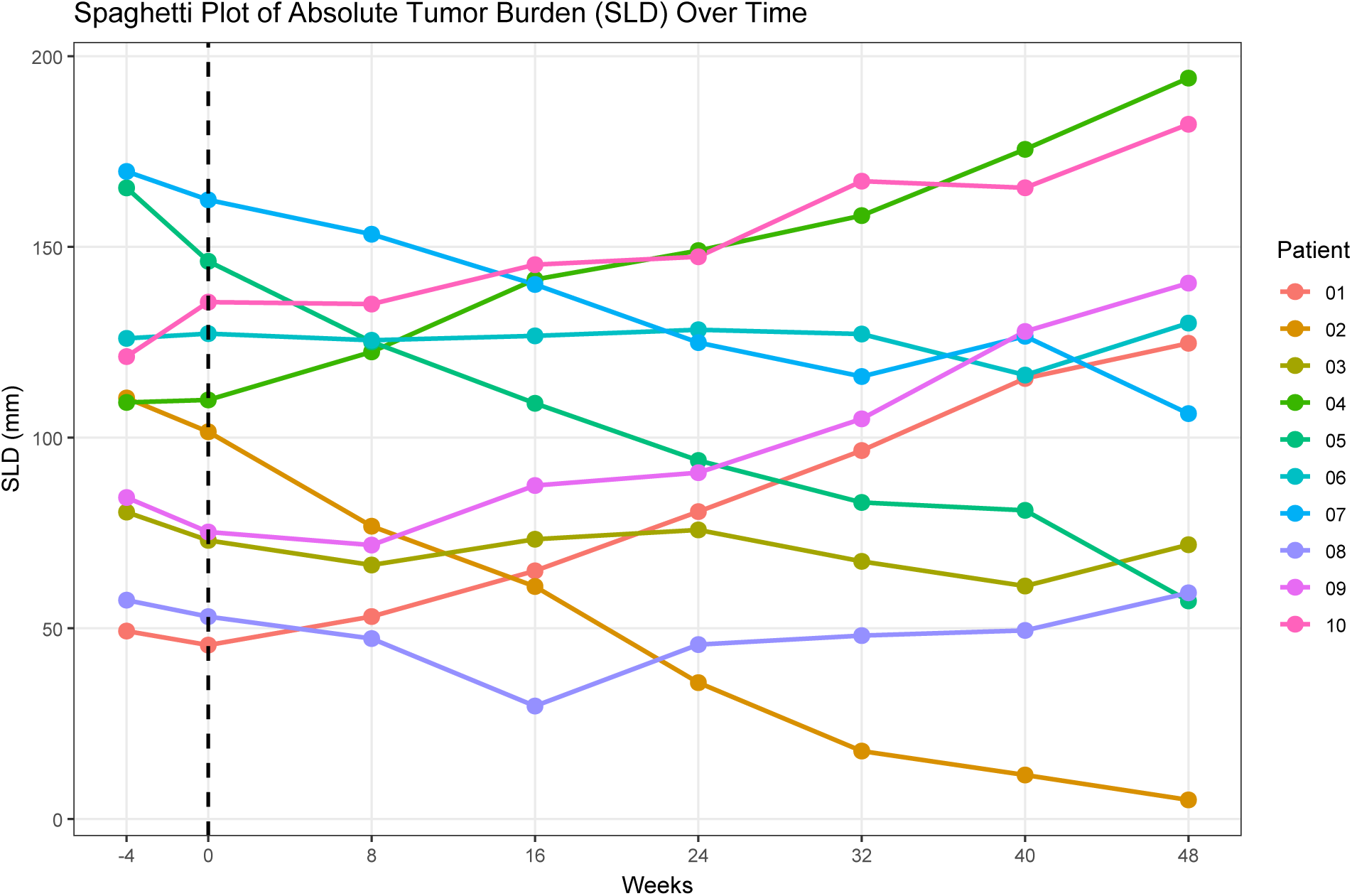
Spaghetti plot of absolute tumor burden (SLD, mm) over follow-up time (weeks) for 10 simulated patients. Each colored line represents one patient’s longitudinal SLD trajectory. The vertical dashed line marks baseline (week 0).

#### 5.2.8 Patient-Journey Timeline (Gantt-style)

**• Technical Specification**

**– Purpose:** Visualize within-patient treatment sequences over calendar time (start/stop segments) to make treatment switching, interruptions, and heterogeneity in real-world trajectories immediately visible [193].
**– Data:** patient_id, day_start, day_end, line_tx (treatment line/category per interval); optional index_day (vertical reference day).
**– Assumption:** Patient-level intervals are non-overlapping (or pre-resolved) within each patient timeline.
**– R Packages:** ggplot2, dplyr.
**• Applications**

**– Use-case 1.** RWE treatment sequencing: Summarize switching patterns across lines of therapy (LOT), including early discontinuation and rapid cycling.
**– Use-case 2.** Operational “messiness” audit: Identify gaps, overlaps, and heterogeneous on-treatment durations that impact time-to-event endpoints and causal estimands.
**• Visualization**

**Figure 33:**
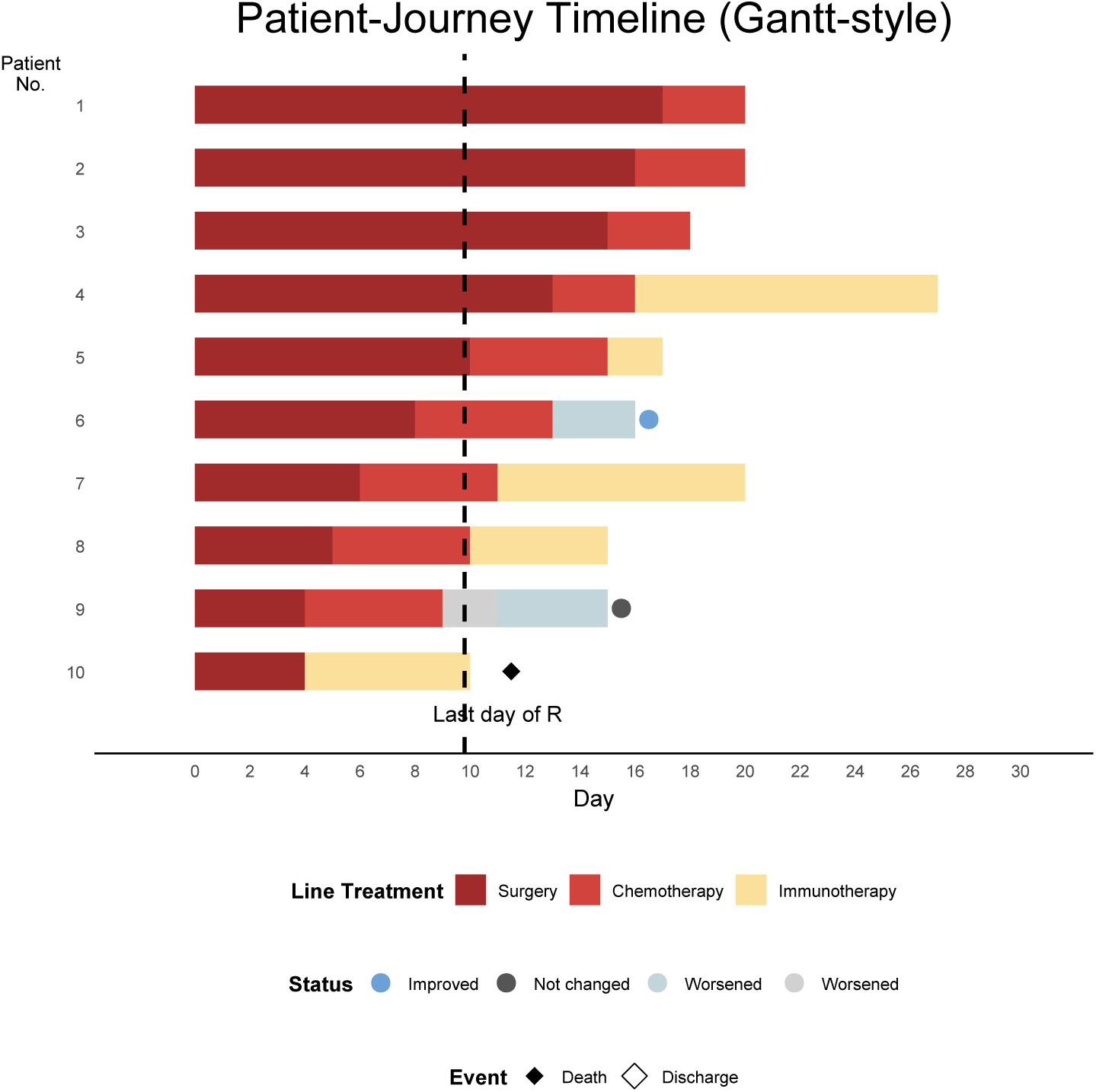
Patient-journey timeline (Gantt-style) for 10 patients showing sequential treatment intervals across days 0–30. Each horizontal bar represents a patient; contiguous segments indicate treatment intervals over time. The dashed vertical line marks a reference day (e.g., end of an index regimen or landmark time; R=Radiation therapy).

#### 5.2.9 Cost-Effectiveness Plane (CEP)(ΔCost vs ΔEffect)

**• Technical Specification**

**– Purpose:** Visualize the joint uncertainty in **incremental effectiveness** (Δ*E*) and **incremental cost** (Δ*C*) for a treatment comparison, and interpret results relative to a **willingness-to-pay (WTP)** threshold line [1, 194, 195].
**– Data:** id (simulation/replicate), dE_qaly (Δ*E* in QALYs), dC_cost (Δ*C* in $), and wtp (e.g., 50,000$/QALY).
**– Assumption:** Points arise from a coherent uncertainty procedure (e.g., probabilistic sensitivity analysis, bootstrap, or IPW-based resampling) so that (Δ*E,* Δ*C*) are comparable across replicates.
– **R Packages:** ggplot2
**• Applications**

**– Use-case 1.** HEOR / RWE oncology CE: Summarize PSA uncertainty around an ICER and show whether draws lie mostly below the WTP line (more likely cost-effective) versus above it.
**– Use-case 2.** Comparator communication: Quickly convey whether the new strategy is typically more effective & more costly (NE quadrant) versus potentially dominant (SE) or dominated (NW).
**• Visualization**

**Figure 34:**
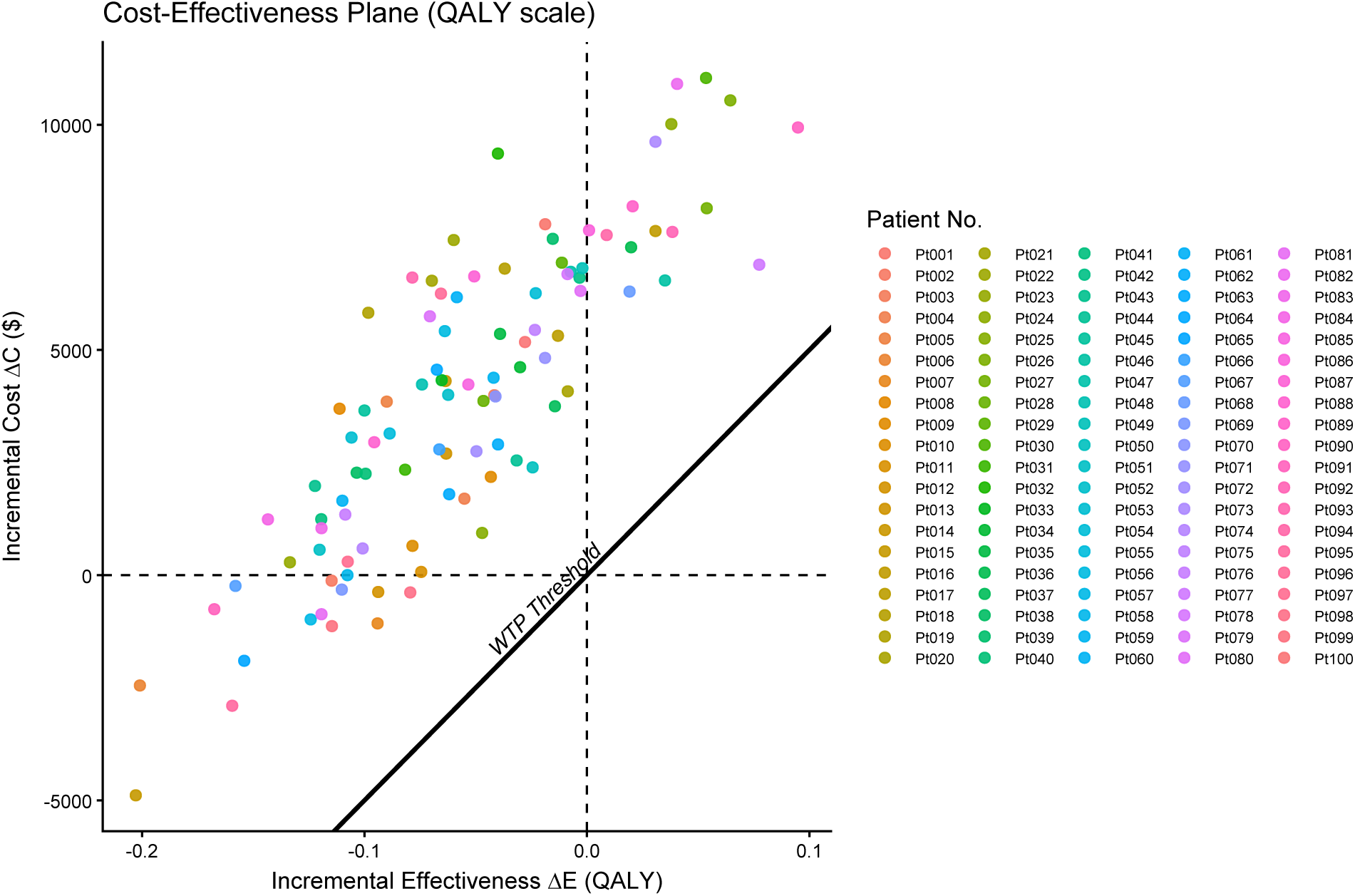
Incremental cost (Δ*C*) versus incremental effectiveness (Δ*E*) in Quality-Adjusted Life Years (QALYs) for an IPTW cohort; the diagonal line represents the WTP threshold of $50,000 per QALY. Each point is one replicate/simulation draw of (Δ*E,* Δ*C*).

#### 5.2.10 Cost-Effectiveness Acceptability Curve (CEAC)

**• Technical Specification**

**– Purpose:** Visualize (over a range of willingness-to-pay, WTP) the probability that each treatment strategy is cost-effective, given joint uncertainty in incremental costs and effects (typically from probabilistic sensitivity analysis / Monte Carlo simulation) [1, 196, 197].
**– Data:** strategy (treatment), lambda (WTP, e.g., $/QALY), p_ce (Pr[cost-effective]), plus underlying PSA draws of incremental ΔC (cost) and ΔE (QALY).
**– Assumption:** Cost-effectiveness defined via Net Monetary Benefit: *NMB*(*λ*) = *λ*Δ*E* −Δ*C* (cost-effective if NMB>0).
**– R Packages:** dplyr, tidyr, ggplot2 (CEAC computed by indicator averaging across PSA draws; smooth curve via geom_smooth()).
Applications

**– Use-case 1.** HEOR decision support: Provide payers/HTA bodies the probability each strategy is cost-effective at jurisdiction-specific WTP thresholds (e.g., 50k–150k $/QALY).
**– Use-case 2.** Uncertainty communication: Summarize probabilistic sensitivity analysis results without relying on a single ICER point estimate (especially when Δ*E* is small/variable).
**• Visualization**

**Figure 35:**
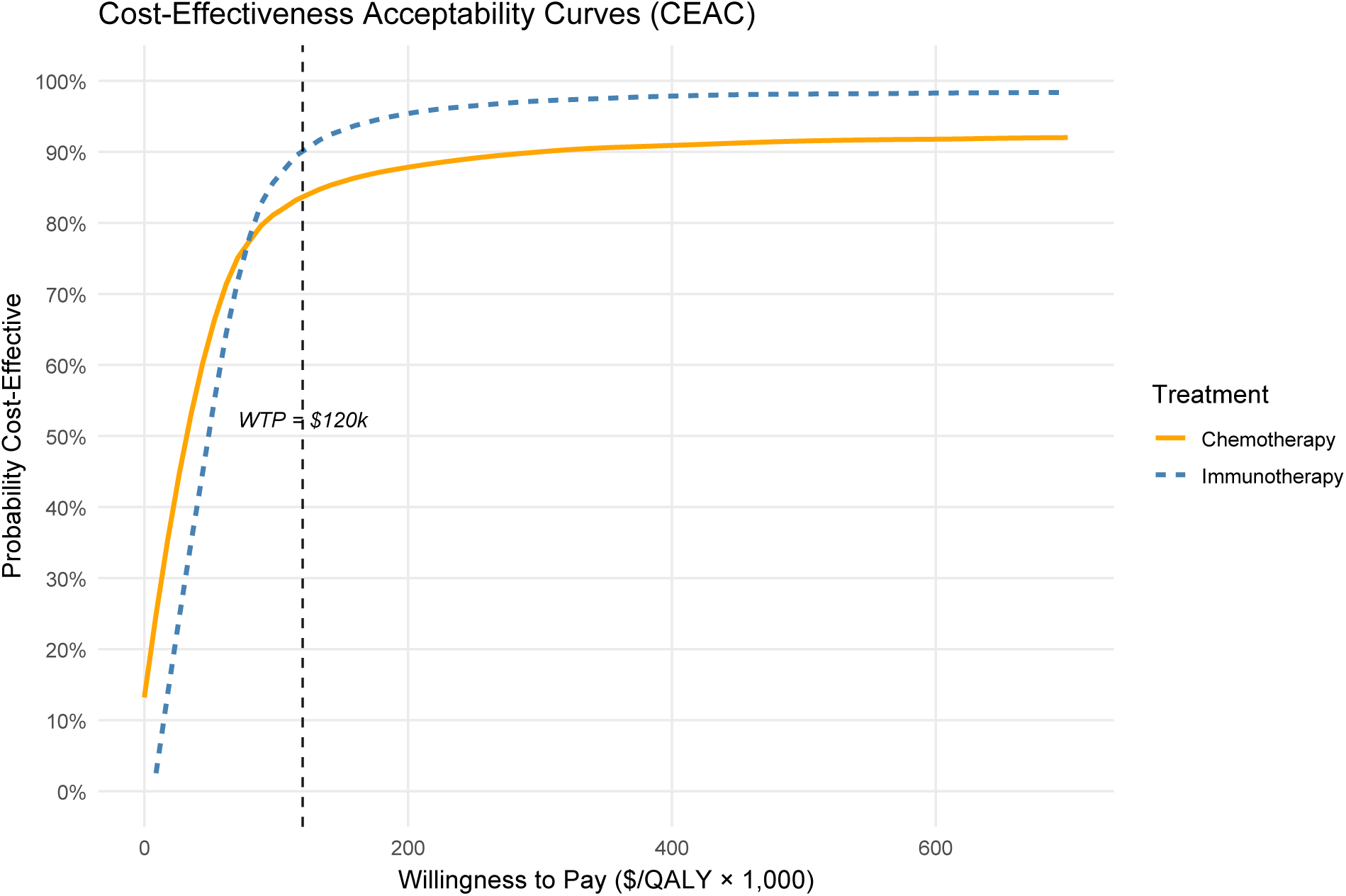
Cost-effectiveness acceptability curves (CEACs) comparing two oncology strategies (e.g., Immunotherapy vs Chemotherapy). The y-axis shows the probability each strategy is cost-effective, defined by positive net monetary benefit *λ*Δ*E* − Δ*C >* 0, across willingness-to-pay thresholds (x-axis; $/QALY × 1,000).

#### 5.2.11 Propensity Score Overlap / Common Support Plot (PS density by treatment; pre/post weighting or matching)

**• Technical Specification**

**– Purpose:** Diagnose positivity / common support and visually assess whether matching (or weighting) improves overlap of the propensity-score distributions between treatment and control [198].
**– Data:** id, trt (0/1), ps (estimated propensity score), and an indicator for stage (e.g., Before matching, After matching).
**– Assumption:** Positivity—both groups have nonzero probability of receiving either treatment within the analyzed covariate region.
**– R Packages:** ggplot2.
**• Applications**

**– Use-case 1.** PSM/IPTW diagnostics (oncology RWE): Verify whether balancing achieved overlap sufficient for causal effect estimation (OS/PFS, treatment utilization, toxicity endpoints).
**– Use-case 2.** Trimming decision support: Identify score regions with near-zero density for one group (violated positivity) → consider trimming, re-matching, or restricting estimand.
**• Visualization**

**Figure 36:**
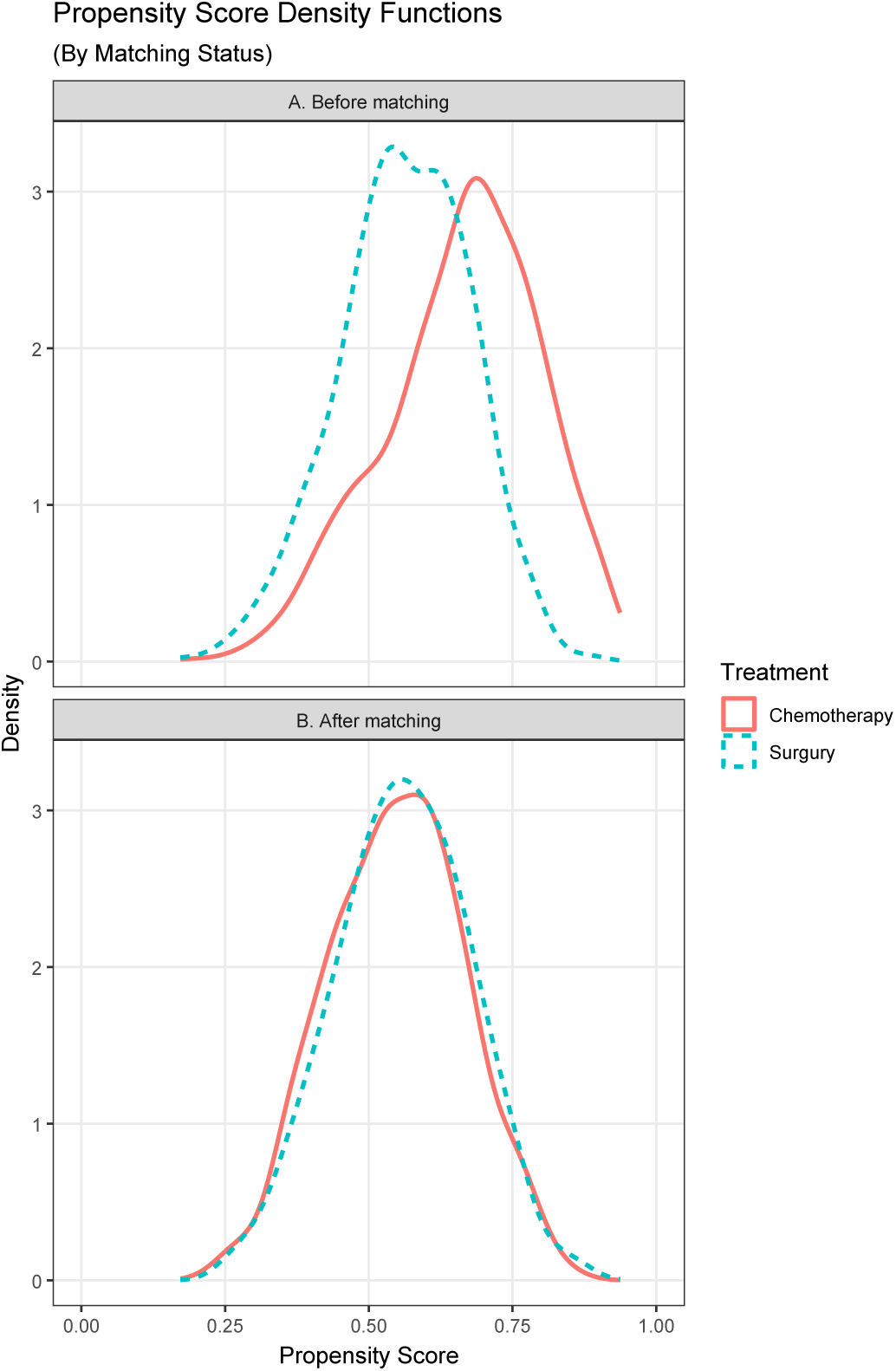
Propensity score density functions before and after matching. The top panel shows the pre-matching propensity score distributions for the treatment and control cohorts, highlighting potential lack of common support. The bottom panel shows increased overlap after matching, consistent with improved comparability under the positivity assumption.

#### 5.2.12 Geographic Variation Choropleth Map

**• Technical Specification**

**– Purpose:** Visualize geographic heterogeneity (“hotspots”) in an oncology metric (here: age-adjusted incidence) across predefined regions (e.g., Canadian Provinces, U.S. states) [199, 200].
**– Data:** region_id (province/state), value (incidence rate), optional year/period, optional data_available flag.
**– Assumption:** Rates are age-adjusted to a standard population and comparable across states for the specified period.
**– R Packages:** ggplot2, dplyr, sf, rnaturalearth, rnaturalearthdata [*Note: This set of required packages may change depending to the region and scale.]
**• Applications**

**– Use-case 1.** Access & equity: identify “postcode lottery” patterns in incidence, screening uptake, or treatment access by geography.
**– Use-case 2.** Resource planning: prioritize regions for outreach clinics, referral pathways, or diagnostic capacity.
**• Visualization**

**Figure 37:**
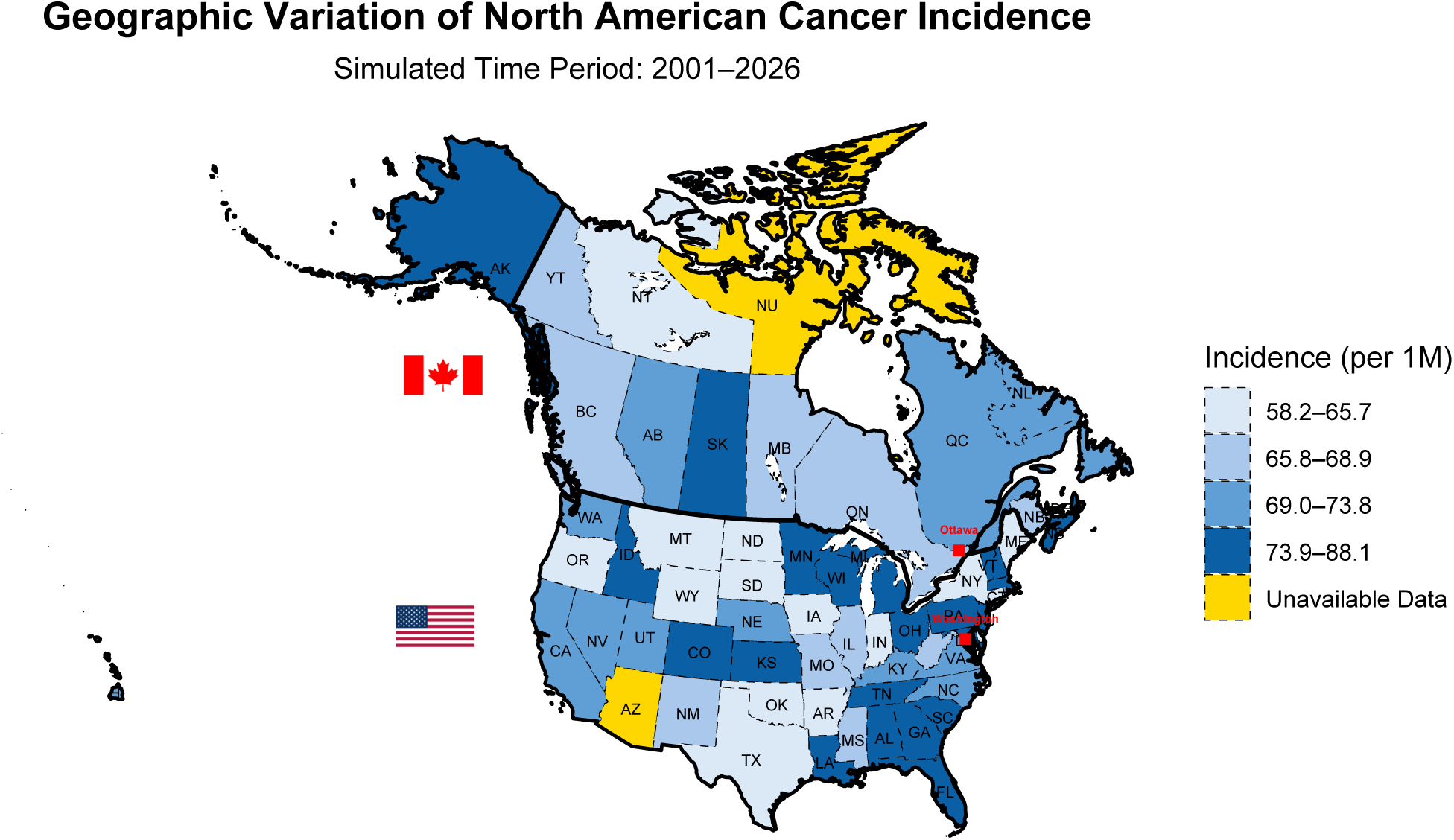
Geographic variation choropleth map: simulated age-adjusted incidence rates per 1,000,000 for Canadian provinces and U.S. states, grouped into the same four legend bins shown in the reference figure; a small subset is labeled ‘Unavailable Data’.

### 5.3 Category C: Common in Clinical Trials and RWE

#### 5.3.1 Meta Analysis Plots

**• Technical Specification**

**– Purpose:** Forest Plot summarizes 12 studies’ effect estimates (e.g., proportion / ORR) and quantifies the pooled random-effects estimate; Funnel Plot assess small-study effects / publication bias [201, 202].
**– Data:** study_id, events, total (or an effect size + SE directly).
**– Assumption:** True effects vary across studies; observed effects differ from study-specific true effects due to sampling error and between-study heterogeneity (*τ* ^2^).
**– R Packages:** meta, ggplot2, patchwork, dplyr, cowplot, ggplotify.
**• Applications**

**– Use-case 1.**Systematic reviews of plausible outcomes (e.g., ORR/DCR) in single-arm oncology evidence (e.g., early-phase immunotherapy signals), where pooled outcomes (e.g., proportions) are the primary endpoint.
**– Use-case 2.** Comparative effectiveness meta-analyses (then forest is typically HR/OR/RR), with funnel plots used to evaluate small-study effects when ≥ 10 studies exist.
**• Visualization**

**Figure 38:**
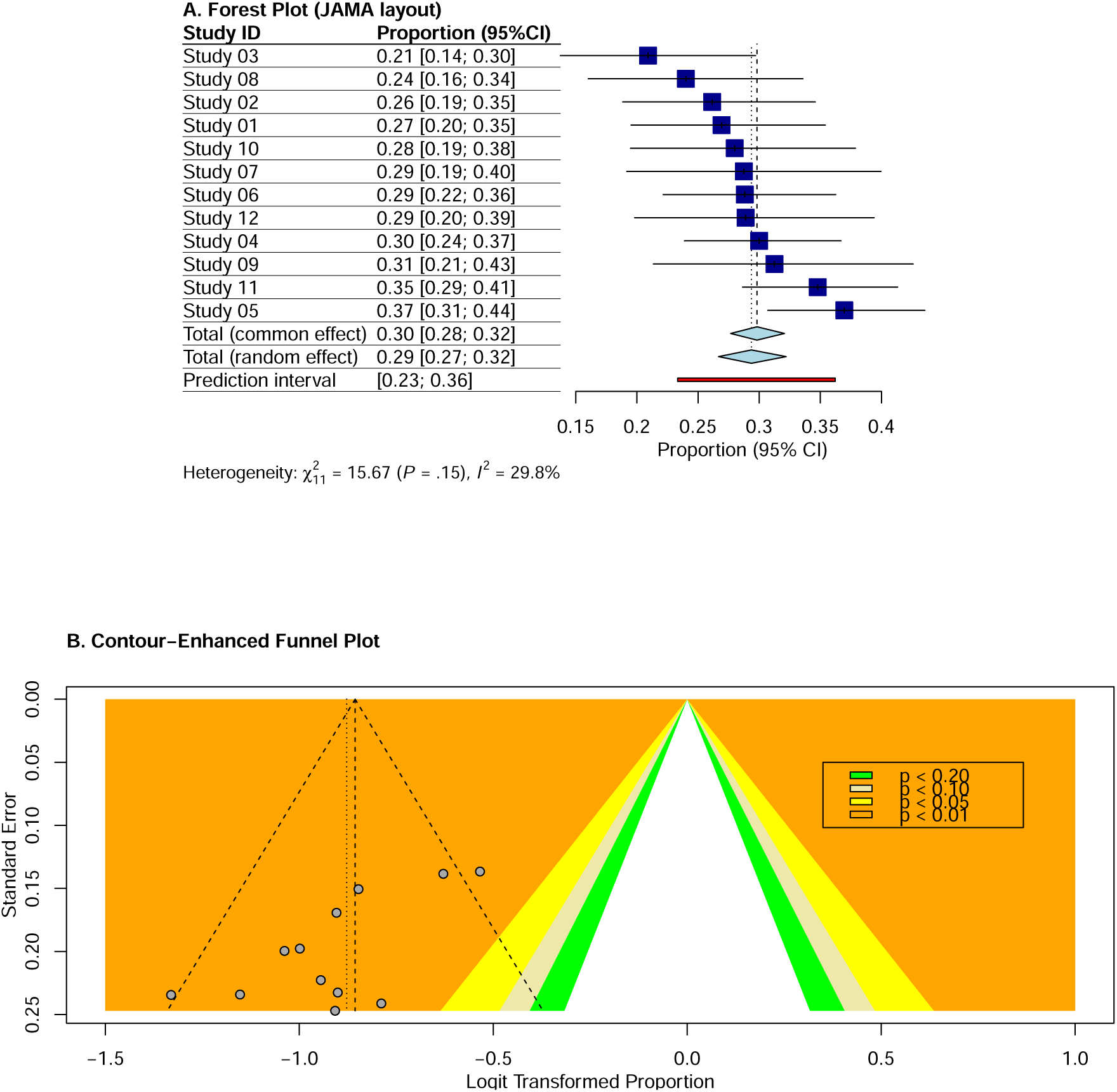
Meta-Analysis Plots: Panel A. a forest plot of 12 studies; Panel B. a contour-enhanced funnel plot.

#### 5.3.2 CONSORT/STROBE participant-flow diagram

**• Technical Specification**

**– Purpose:** Visualize cohort attrition and participant disposition from screening/data pull → exclusions → allocation/follow-up → analysis (CONSORT for RCT; STROBE-style for RWE) [203, 204].
**– Data:** stage labels, node counts, reasons for exclusion, arm labels (if RCT), and analysis set definitions (e.g., ITT).
**– Assumption:** counts are mutually exclusive and sum consistently across branches (screened = excluded + randomized/eligible; randomized = arm1 + arm2).
**– R Packages:** DiagrammeR, DiagrammeRsvg, rsvg(export to file, no on-screen rendering).
**• Applications**

**– Use-case 1.** [i] RCT transparency: Quickly communicates screening pressure, protocol attrition, and the analysis population definition (e.g., ITT). [ii] RWE credibility checks: Makes data-loss mechanisms explicit (missingness, eligibility rules), supporting interpretability and transportability.
**– Use-case 2.** Reviewer-facing QA: Helps reviewers verify denominator consistency and whether exclusions could induce selection bias.
**• Visualization**

**Figure 39:**
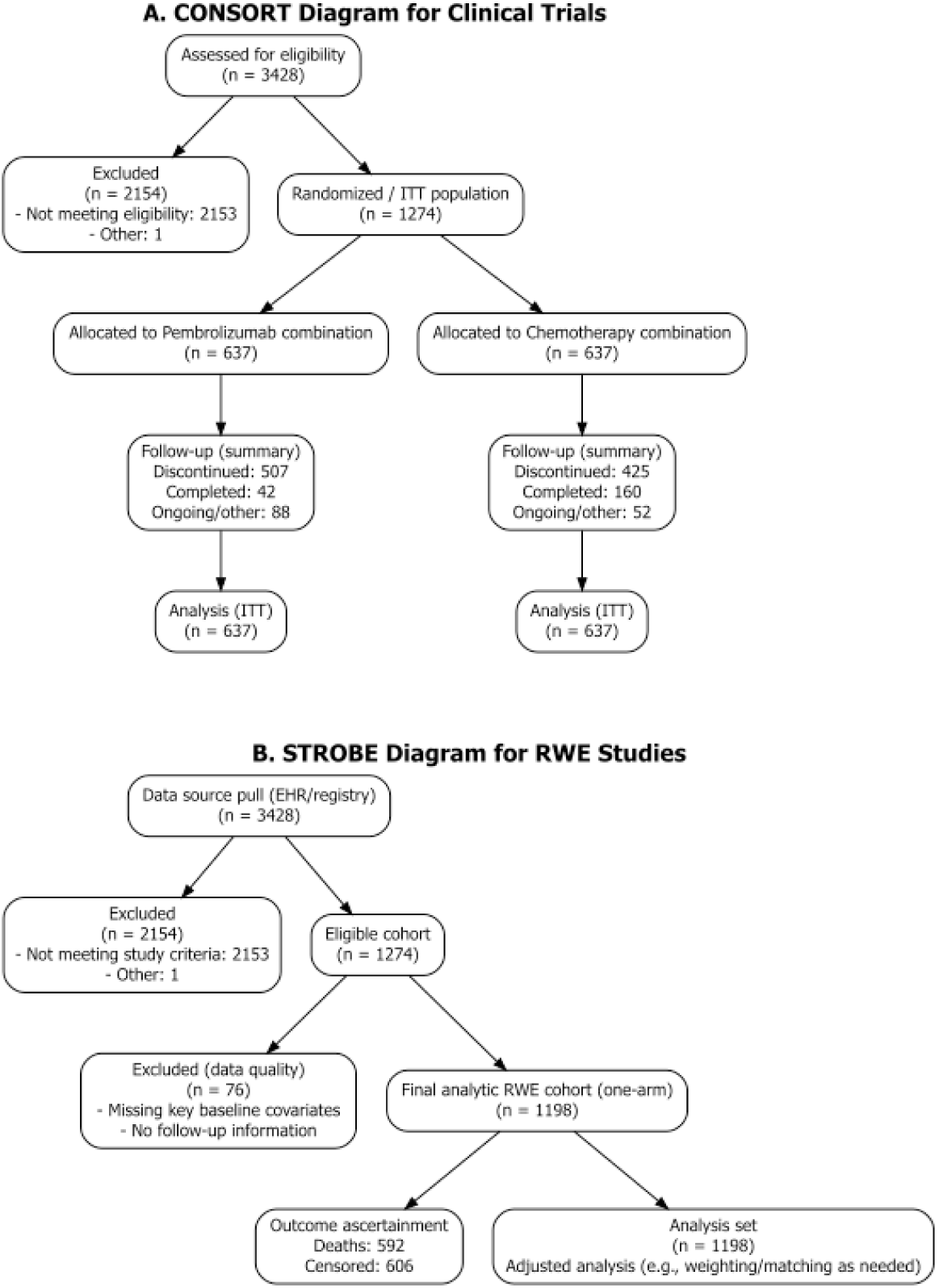
Participant-flow diagrams. (A) CONSORT-style disposition for a simplified two-arm randomized trial: 3428 assessed, 2154 excluded, 1274 randomized (637 per arm) and analyzed by intention-to-treat (ITT). (B) STROBE-style cohort derivation for a simplified one-arm real-world evidence (RWE) study: starting from 3428 in the data pull, exclusions yield a final analytic cohort of 1198 with outcomes summarized as deaths and censoring.

#### 5.3.3 Marker value–Response Distribution Plot

**• Technical Specification**

**– Purpose:** Visualize how the **response probability** varies as a function of a (log-transformed) **continuous biomarker** for two treatments (T1 vs T2), and how a biomarker cut-off *c* induces marker-positive *M* ^+^ = {*X > c*} vs marker-negative *M* ^−^ = {*X* ≤ *c*} subgroups [1].
**– Data:** patient-level marker *X* (continuous), treatment indicator *T* ∈ {T1, T2}, and binary response *Y* ∈ {0, 1}.
**– Assumption:** The response is modeled with a **logit** link: Pr(*Y* = 1 | *X* = *x, T* = *j*) = 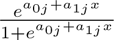, *j* ∈ {1, 2}, and the marker distribution can be taken as **normal** *X* ∼ *N* (*µ, σ*^2^) when simulating scenarios.
**– R Packages:** ggplot2.
**• Applications**

**– Use-case 1.** Biomarker cut-off justification: visually supports why a specific *c* is chosen (e.g., separating a region where T2 meaningfully outperforms T1).
**– Use-case 2.** Trial-design comparison: supports power/operating-characteristic work by linking marker prevalence *v_M_*+ (*c*) and subgroup response rates to design performance.
**• Visualization**

**Figure 40:**
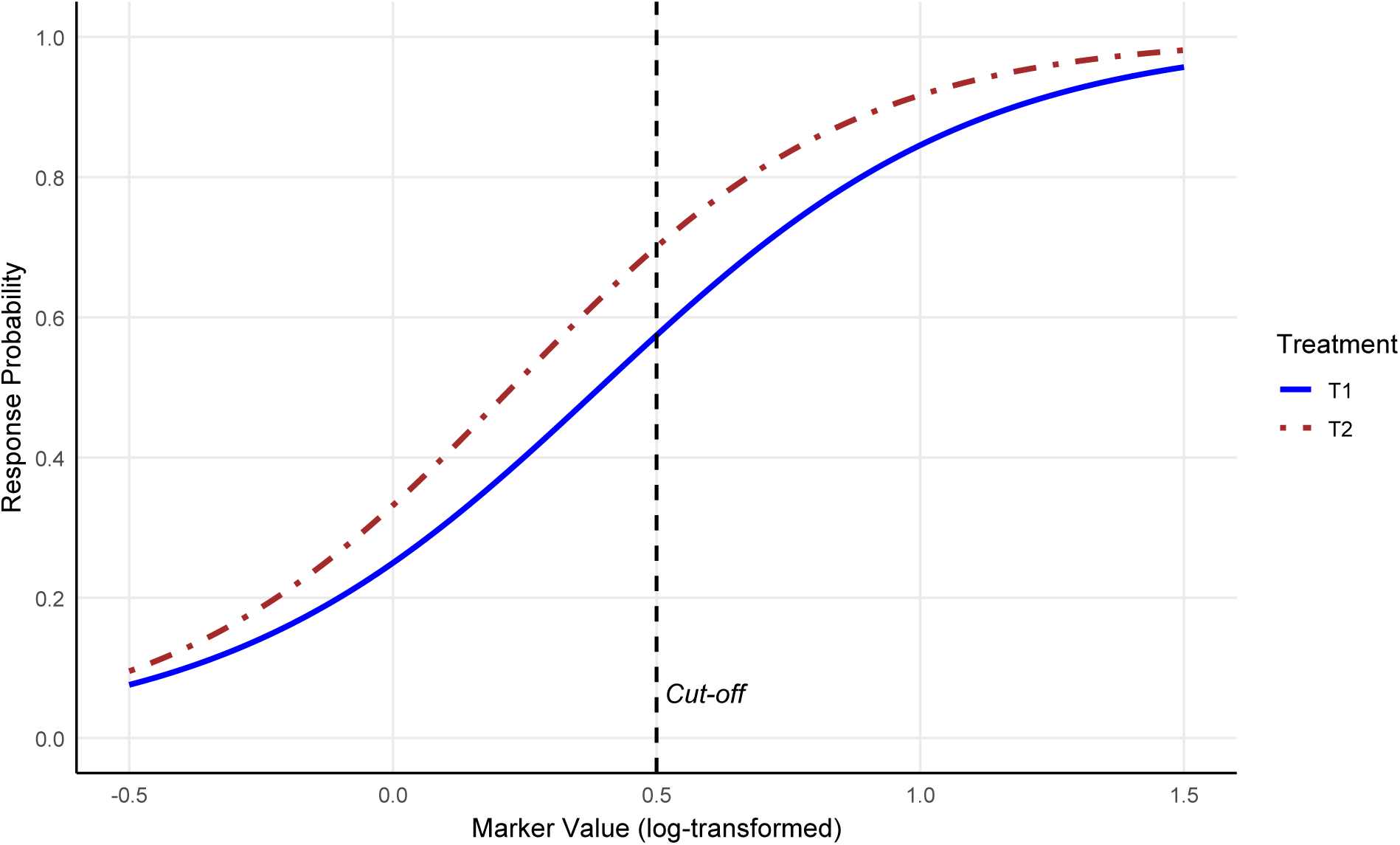
Marker value–response distribution plot. Response probability is plotted versus the (log-transformed) marker value *x* for two treatments (T1, T2) under a logistic response model. The vertical line denotes the biomarker cut-off *c* defining marker-positive *M* ^+^ = {*X > c*} versus marker-negative *M* ^−^ = {*X* ≤ *c*} patients; curves illustrate the expected response profiles used for scenario-based design evaluation.

#### 5.3.4 Recurrence Score–Log(HR) Curve

**• Technical Specification**

**– Purpose:** Visualize the modeled relationship between Recurrence Score (RS) and the Cox regression log hazard ratio (log HR) under two treatment strategies (e.g., Chemotherapy vs No chemotherapy), highlighting the RS “equivalence” point where treatments are predicted to have equal effect [1].
**– Data:** *RS* (continuous, e.g., 0–30), Treatment indicator *Z* ∈ {Chemo, No chemo} Time-to-event *T*, event indicator *δ* (for Cox model fit), Optional: model-predicted log *HR*(*RS, Z*) and CI for the equivalence point.
**– Assumption:** A Cox proportional hazards model (possibly with RS×treatment interaction) is used to obtain a modeled log(*HR*) as a function of RS and therapy; the equivalence RS is where the two modeled curves intersect.
**– R Packages:** ggplot2, dplyr.
**• Applications**

**– Use-case 1.** Predictive marker signal: Shows whether treatment effect varies with RS (interaction-like behavior).
**– Use-case 2.** Clinical decision thresholding: The equivalence RS and its CI support defining “low RS” vs “high RS” regions where chemotherapy is predicted inferior vs superior.
**• Visualization**

**Figure 41:**
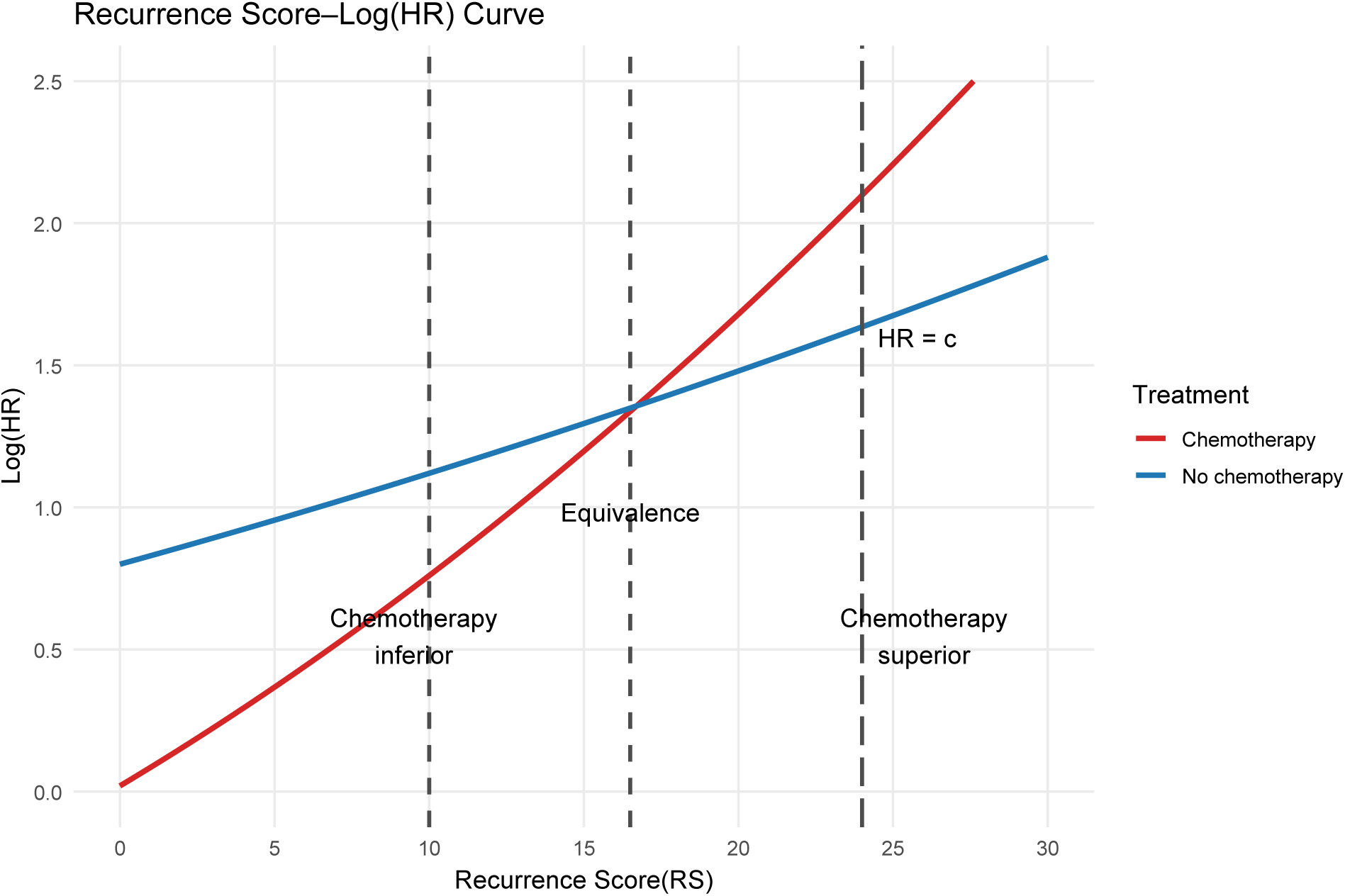
Modeled relationship of the Cox regression log HR relating survival to recurrence score (RS) and therapy. The middle dashed vertical line indicates the RS value where treatments are equivalent (intersection of modeled curves). The side dashed vertical lines indicate the lower/upper bound of the 95% confidence interval for the point of equivalence.

#### 5.3.5 Time–Quality of Life Curve

**• Technical Specification**

**– Purpose:** Visualize and compare health-state utility trajectories over time for two treatments; the area under each curve approximates total Quality Adjusted Life Years(QALYs), and the area between curves approximates QALYs gained by the better treatment [1].
**– Data:** time (e.g., months), utility (0=death, 1=perfect health; can allow “worse than death” if your instrument permits), treatment (Trt 1 vs Trt 2). Optionally death_time per arm.
**– Assumption:** Utility is measured/assigned at discrete times and is interpolated (often stepwise/linear) between assessments to approximate the integral.
**– R Packages:** ggplot2, dplyr, tidyr.
**• Applications**

**– Use-case 1.** HTA/HEOR reporting: Convert longitudinal HRQoL into QALYs for incremental cost-effectiveness (ICER, NMB).
**– Use-case 2.** Trial vs RWE comparison: Show how real-world discontinuations/toxicity patterns change the utility trajectory and cumulative QALYs.
**• Visualization**

**Figure 42:**
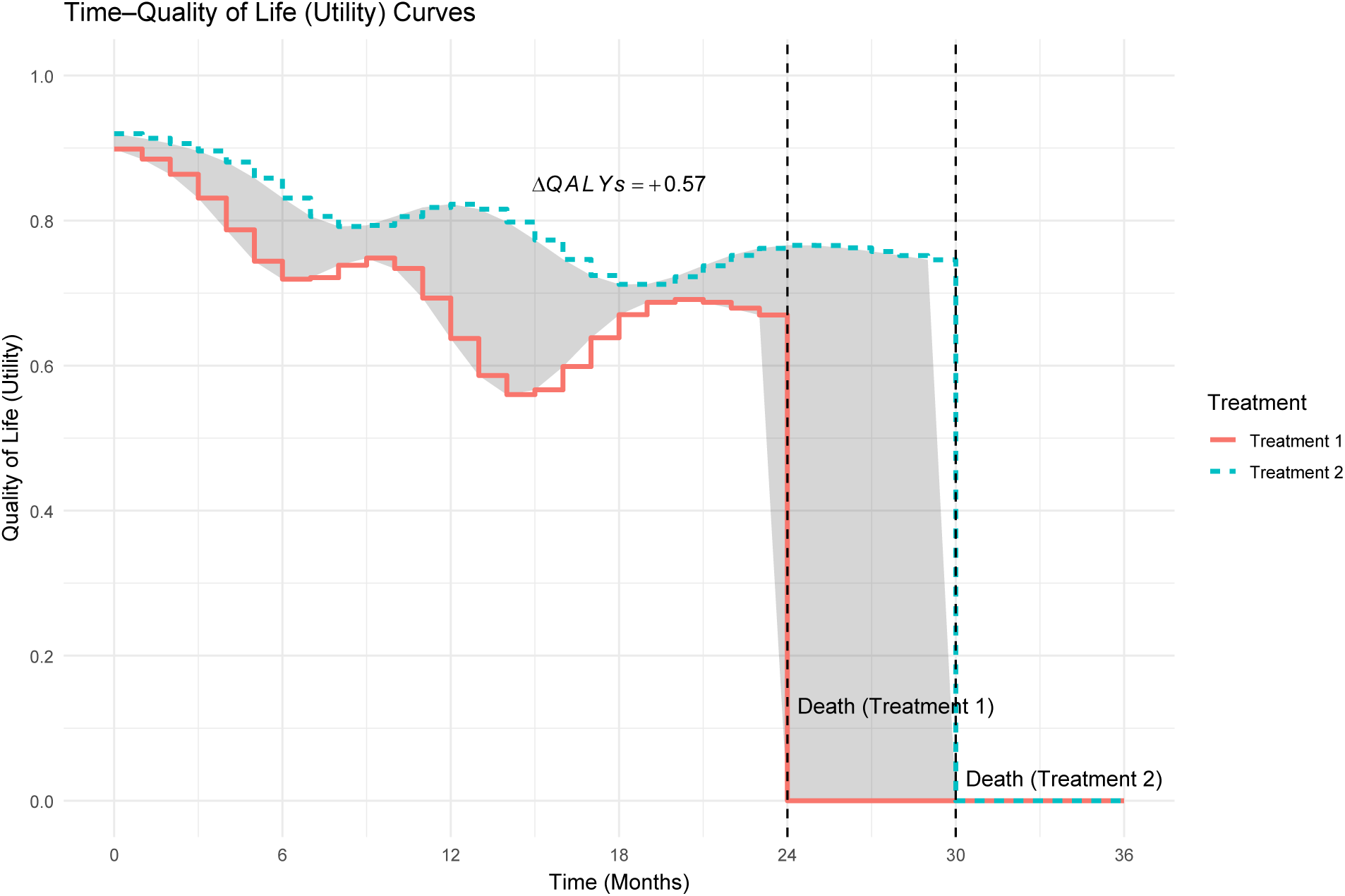
Time–quality of life (utility) curves for two treatments, where utility is anchored at 1 (perfect health) and 0 (death) and the area under the curve represents accumulated quality-adjusted survival (QALYs). The dashed vertical lines indicate the time of death under Treatment 1 and Treatment 2; the shaded region between curves represents QALYs gained by the treatment with higher utility over time.

#### 5.3.6 Time - Cancer Cells Counts(Size)

**• Technical Specification**

**– Purpose:** Visualize tumor burden dynamics over time under chemotherapy cycles, emphasizing fractional (log-kill) reductions during dosing and regrowth between cycles (sawtooth pattern), versus an untreated growth trajectory [205, 206].
**– Data:** time, tumor_cells (or tumor_size), arm (treatment strategy label), and optionally cycle_window (for chemo-on shading).
**– Assumption:** Each chemotherapy cycle removes a constant fraction of tumor cells; between cycles, residual cells regrow exponentially (with possible “wound-healing” acceleration).
**– R Packages:** ggplot2, dplyr, tidyr.
**• Applications**

**– Use-case 1.** Mechanistic oncology modeling: Demonstrate log-kill behavior, resistance/regrowth, and the impact of cycle timing/intensity.
**– Use-case 2.** Regimen comparison: Contrast cycle spacing, kill fraction, and accelerated regrowth (e.g., post-treatment inflammatory/ wound-healing effects).
**• Visualization**

**Figure 43:**
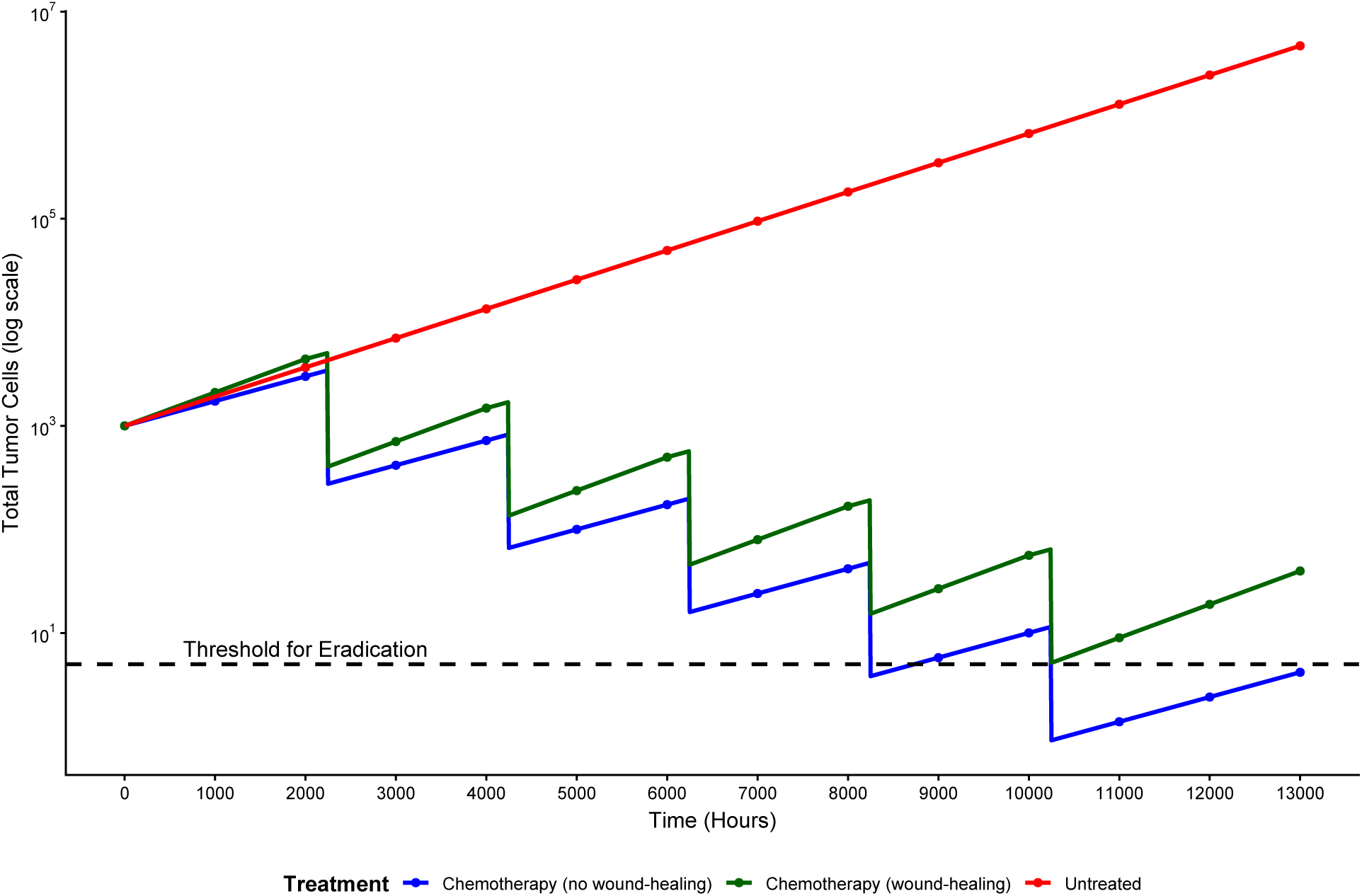
Simulated tumor-burden trajectories under an untreated course versus cyclic chemotherapy under a fractional-kill (log-kill) mechanism, shown on a semi-log scale. Vertical shaded bands indicate chemotherapy cycle windows; chemotherapy arms exhibit stepwise reductions followed by inter-cycle regrowth, with accelerated regrowth under a wound-healing scenario.

#### 5.3.7 Classification and Regression Trees(CART)

**• Technical Specification**

**– Purpose:** Visualize a recursive partitioning model (CART) that splits a cohort into prognostic subgroups using covariate cutpoints [1].
**– Data:** y (outcome; e.g., event yes/no), and candidate predictors (e.g., nodes, progrez, tsize, age, estrez).
**– Assumption:** The tree is an interpretable approximation to the underlying risk surface; splits are chosen to optimize an impurity criterion (e.g., Gini) and then optionally pruned.
**– R Packages:** rpart, rpart.plot.
**• Applications**

**– Use-case 1.** Clinical trials (prognostic enrichment): Identify cutpoints (e.g., node count, receptor levels, tumor size) that define risk strata for subgroup summaries or hypothesis generation.
**– Use-case 2.** RWE (risk stratification / confounding diagnostics): Build interpretable risk groups for outcome modeling, then compare baseline balance or treatment effects within leaves.
**• Visualization**

**Figure 44:**
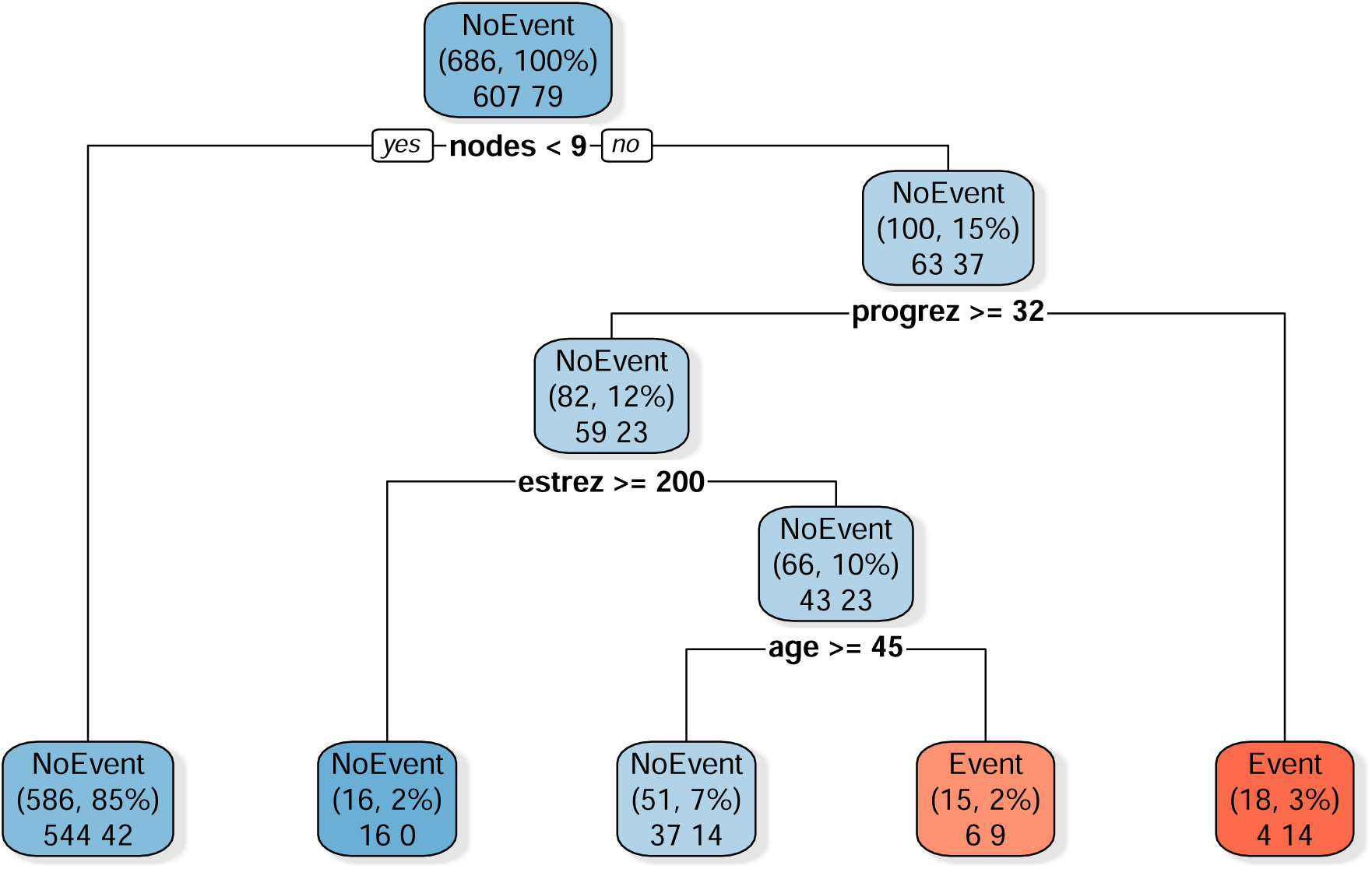
Classification and regression tree (CART) illustrating recursive partitioning into prognostic subgroups using common clinical covariates (e.g., lymph nodes, receptor measures, tumor size, age). The displayed tree is a simplified, simulated example in the style of the GBSG-2 study figure, with node boxes showing subgroup size and predicted event risk.

#### 5.3.8 Time–Prediction Error Curve

**• Technical Specification**

**– Purpose:** Visualize and compare time-dependent prediction error of multiple prognostic classification schemes (lower curve = better predictive performance over time) [1].
**– Data:** time (continuous; e.g., years from 0 to 6), method (factor; e.g., Kaplan–Meier, Cox(PI), NPI, CART), pred_error (numeric; typically the Brier-score–based prediction error at each time)
**– Assumption:** Prediction error is computed as a **time-indexed Brier-type loss** comparing observed event status at time *t* to estimated event-free probability *S*^^^(*t* | *Z*) (with censoring handled via standard reweighting if needed).
**– R Packages:** ggplot2::ggplot, ggplot2, dplyr, tidyr (optional).
**• Applications**

**– Use-case 1.** Compare prognostic schemes (e.g., Cox prognostic index vs. NPI vs. CART) on the same cohort across clinically relevant horizons.
**– Use-case 2.** Model selection with time horizon: pick the method minimizing prediction error around a prespecified decision time (e.g., 3 or 5 years).
**• Visualization**

**Figure 45:**
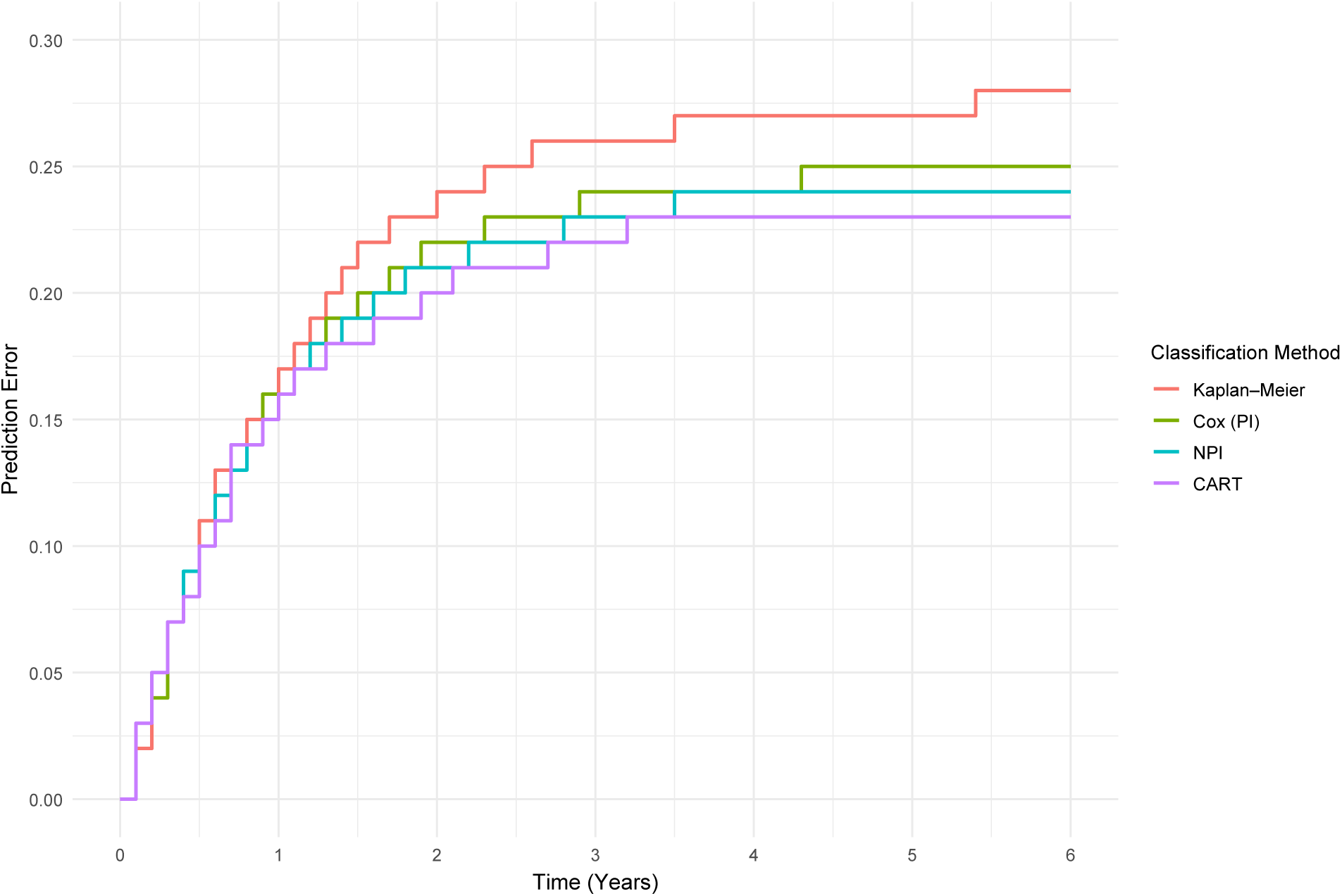
Estimated time-dependent prediction error curves (Brier-score–based) for four prognostic classification schemes (Kaplan–Meier, Cox prognostic index, NPI, and CART) over 0–6 years. The prediction error at time *t* summarizes the discrepancy between the observed event indicator by *t* and the estimated event-free probability; lower curves indicate better predictive performance over time.

#### 5.3.9 Spider Plot

- Technical Specification

**– Purpose:** Display within-patient longitudinal percent change in target lesion size from baseline to visualize tumor shrinkage/growth patterns over time (often aligned with best overall response) [207].
**– Data:** patient_id (factor/character; 10 patients),time_wk (numeric; e.g., 0–54 weeks),pct_change (nu­meric; % change from baseline target-lesion sum), disease_status (factor; e.g., Complete response, Partial response, Stable disease, Progression, New lesion).
**– Assumption:** Percent change is computed relative to baseline for each patient and followed on a fixed visit grid.
**– R Packages:** ggplot2, dplyr.
- **• Applications**

**– Use-case 1.** Clinical trials: Visualize individual tumor-burden trajectories to contextualize best overall response categories (CR/PR/SD/PD).
**– Use-case 2.** RWE cohorts: Compare response dynamics across regimens or biomarker strata (e.g., responders vs non-responders) using the same longitudinal endpoint definition.
- **• Visualization**

**Figure 46:**
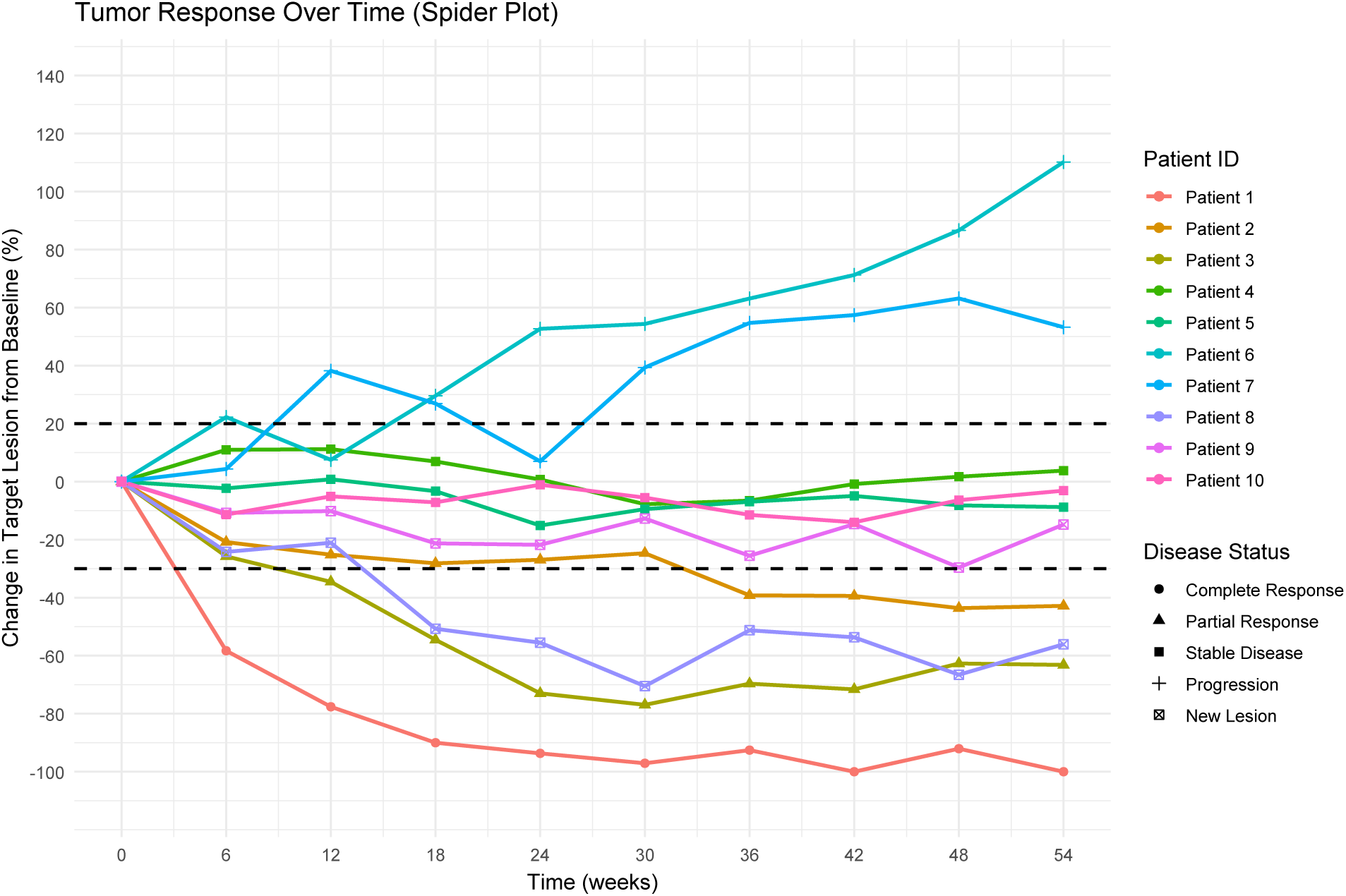
Hypothetical spider plot of longitudinal percent change in target lesion size from baseline for 10 patients followed over 0–54 weeks. Lines are color-coded by patient and styled by disease status (complete response, partial response, stable disease, progression, and new lesion).

#### 5.3.10 Cumulative Incidence Function (CIF) Plot

**• Technical Specification**

**– Purpose:** Visualize the event-specific cumulative incidence function (CIF) over time when competing risks are present (e.g., cancer death vs cardiovascular death), so probabilities are not inflated as they can be under Kaplan–Meier [208–210].
**– Data:** time (follow-up time), status (0=censored, 1=event of interest, 2=competing event), optional group (if stratified).
**– Assumption:** Competing events are real alternatives (once one occurs, the other cannot).
**– R Packages:** cmprsk, ggplot2.
**• Applications**

**– Use-case 1.** Oncology RWE: Cancer-specific mortality in the presence of substantial non-cancer death (cardiovascular, infection, other causes).
**– Use-case 2.** HTA / outcomes reporting: Cause-specific absolute risks over time for decision-making (e.g., 5-year cancer death CIF vs other-cause death CIF).
**• Visualization**

**Figure 47:**
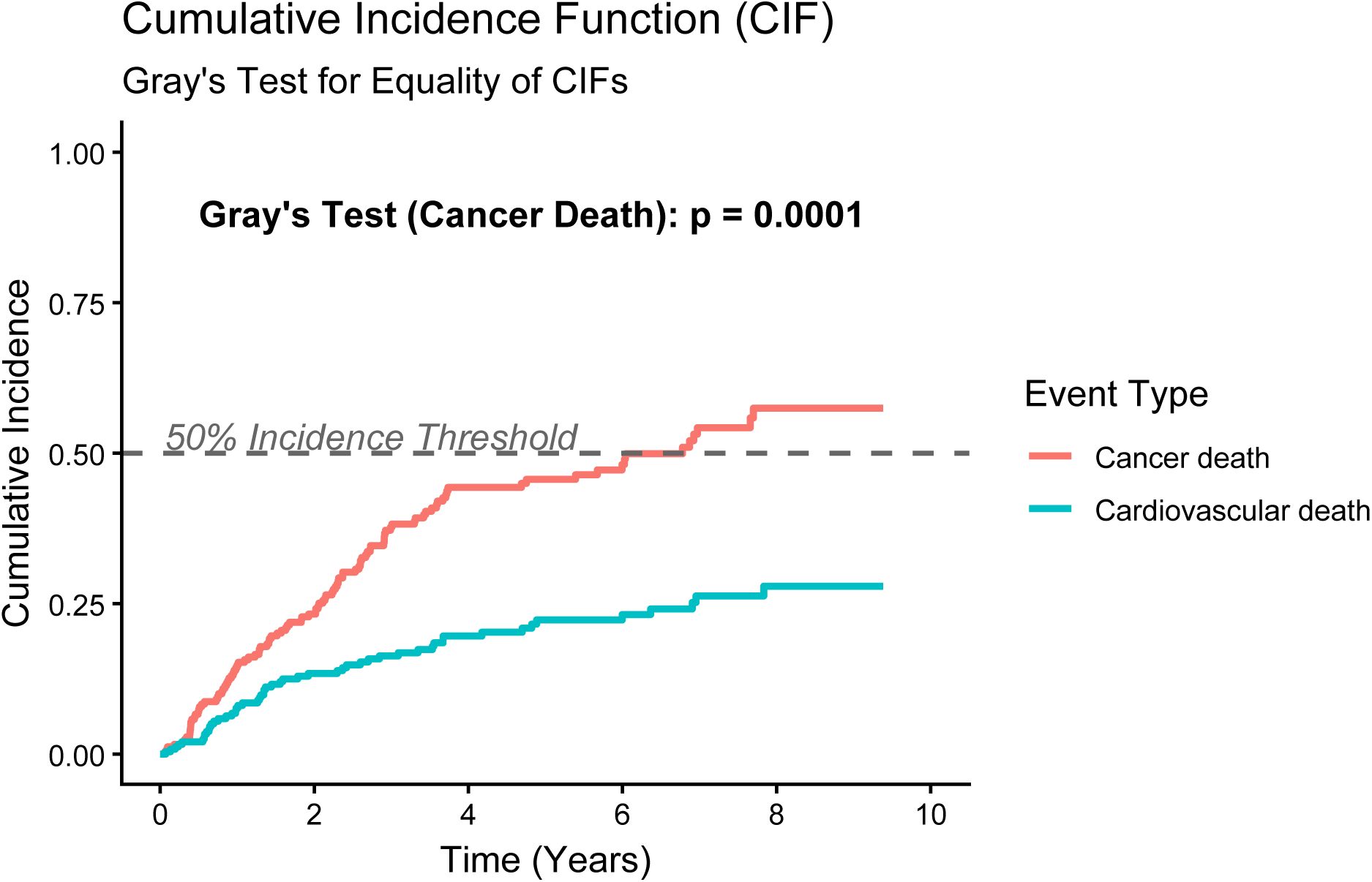
Cumulative incidence curves for competing events. Event-specific CIFs for Cancer death and Cardiovascular death are plotted over 10 years(CIF estimates the probability of each cause occurring over time while explicitly accounting for the competing risk).

#### 5.3.11 Stacked Cumulative Incidence Plot

**• Technical Specification**

**– Purpose:** Visualize how multiple competing causes of death accumulate over time by stacking cause-specific cumulative incidence functions (CIFs); the top of the stack is the total cumulative incidence (≤ 1) [211, 212].
**– Data:** time (years since diagnosis), cause (e.g., Breast Cancer / Disease of Heart / Other Cancers / Other Causes), cif (cause-specific cumulative incidence in [0, 1]).
**– Assumption:** CIFs are nondecreasing in time and the stacked sum across causes does not exceed 1.
**– R Packages:** ggplot2, dplyr, tidyr (optional).
**• Applications**

**– Use-case 1.** Competing risks reporting: Show how cause-specific mortality contributes to overall mortality over follow-up (e.g., cancer vs cardiovascular vs other).
**– Use-case 2.** Clinical interpretation: In advanced-stage older populations, stacked CIFs highlight whether outcomes are primarily driven by cancer death versus non-cancer causes.
**• Visualization**

**Figure 48:**
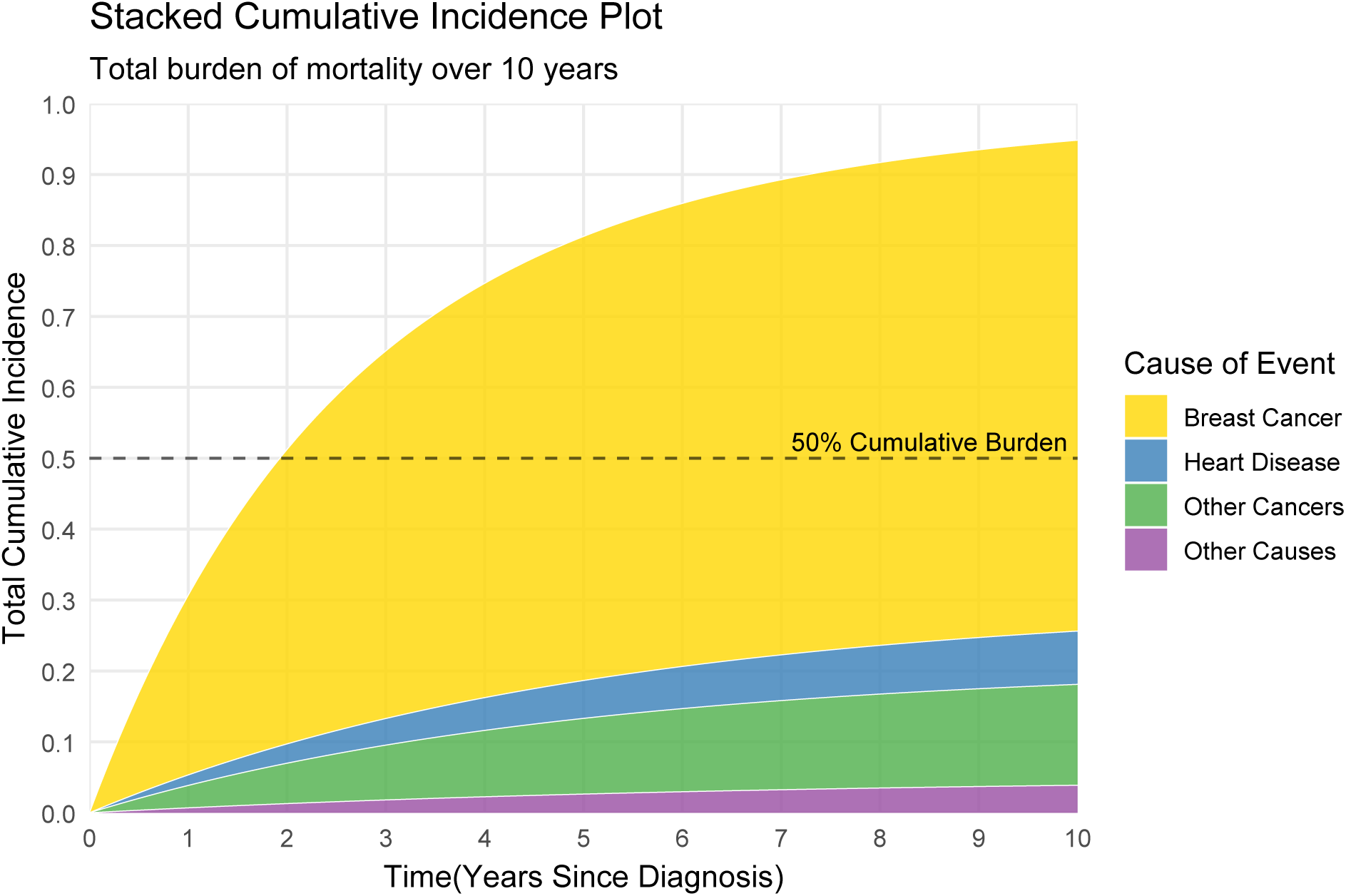
Stacked cumulative incidence function plot for competing causes of death in an older cohort (Age 80+) with distant-stage disease. Areas represent cause-specific cumulative incidence over time; the stacked total shows the overall probability of death from any listed cause by each time point.

#### 5.3.12 Hazard Ratio over time HR(t) Plot

**• Technical Specification**

**– Purpose:** Visualize non-proportional hazards by plotting cause-specific hazard functions over time for Treatment vs Control; crossing hazards indicate the treatment effect changes direction over follow-up [10, 213].
**– Data:** time (continuous follow-up time; e.g., years), hazard (instantaneous hazard rate at each time), arm (Treatment vs Control)
**– Assumption:** Hazards are smooth functions of time and not required to be proportional.
**– R Packages:** ggplot2, dplyr, tidyr.
Applications

**– Use-case 1.** Diagnose PH violations: A crossing hazard pattern [a] Supports using time-varying effects (e.g., Cox model with time-by-treatment interaction) rather than a single constant HR; [b] Helps pinpoint when treatment risk switches from unfavorable to favorable (or vice versa).
**– Use-case 2.** Method selection: Motivates alternatives such as Restricted mean survival time(RMST), weighted log-rank, or piecewise hazard/PH modeling when effects are not constant.
**• Visualization**

**Figure 49:**
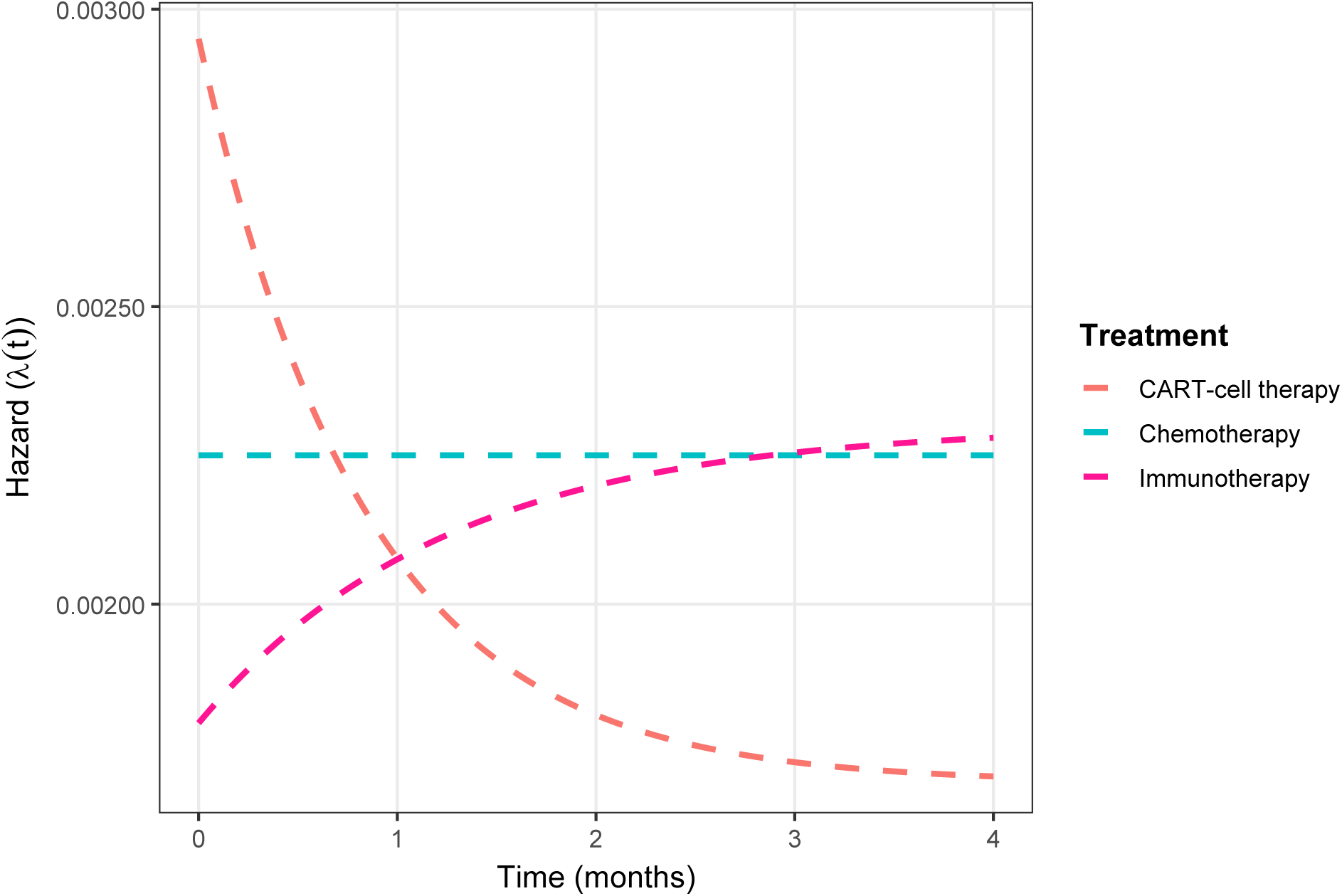
Hazard curves over time illustrating non-proportional hazards via a crossing pattern.

#### 5.3.13 Log(–log) Survival Plot

**• Technical Specification**

**– Purpose:** A log(− log(*S*(*t*))) plot (often against log(time)) is a proportional-hazards (PH) diagnostic: if hazards are approximately proportional across groups (e.g., Scarff–Bloom–Richardson(SBR) grade 1/2/3), then the transformed curves are roughly parallel over time [214].
**– Data:** time: follow-up time (e.g., years since diagnosis), status: event indicator (1=event, 0=censored), grade: subgroup factor (e.g., SBR grade 1/2/3).
**– Assumption:** Cox PH is plausible so there are parallel log(− log(*S*(*t*))) curves.
**– R Packages:** survival, ggplot2, dplyr, tidyr for data shaping.
**• Applications**

**– Use-case 1.** Cox PH assumption check before reporting a single HR for prognostic groups (e.g., grade, stage, biomarker strata).
**– Use-case 2.** Model choice guidance: [a] if curves are not parallel, consider time-varying effects; [b] Covariate screening in multivariable survival modeling to identify variables driving non-PH.
**• Visualization**

**Figure 50:**
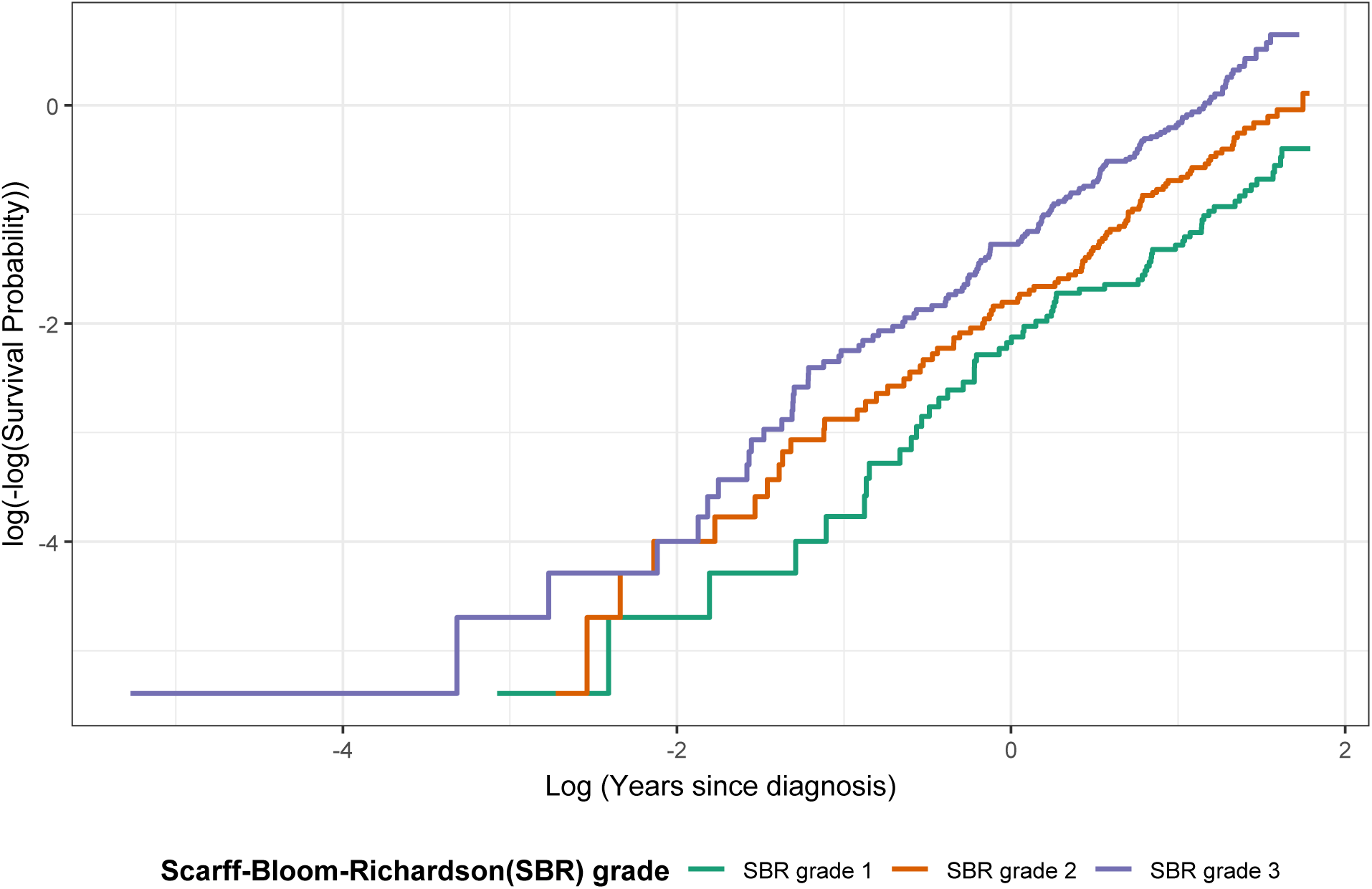
Log(− log(*S*(*t*))) curves as a function of time (log scale) by SBR grade. The plot displays Kaplan–Meier–based log(− log(survival)) transformations for three tumor-grade strata to visually assess proportional hazards; approximate parallelism supports the PH assumption, while crossing/ divergence suggests time-dependent effects.

#### 5.3.14 Schoenfeld Residual Plot

**• Technical Specification**

**– Purpose:** Diagnose violations of the Cox proportional hazards (PH) assumption by checking whether scaled Schoenfeld residuals exhibit a systematic trend over time [214].
**– Data:** time (follow-up), status (event indicator), and at least one covariate (here: grade with a contrast SBR grade 3 vs grade 1).
**– Assumption:** Under PH, the covariate effect is time-constant so the residual trend should be approximately horizontal around 0 (Systematic departure of the smoothed residual trend from 0 suggests violation of the proportional hazards assumption for the given covariate).
**– R Packages:** survival, ggplot2.
**• Applications**

**– Use-case 1.** Model checking in oncology RCTs/RWE: Validate whether a single Cox HR is interpretable for a key covariate (e.g., grade, biomarker, treatment
**– Use-case 2.** Non-PH follow-up: If violated, motivate time-varying Cox terms, landmark analyses, or alternative estimands (e.g., RMST).
**• Visualization**

**Figure 51:**
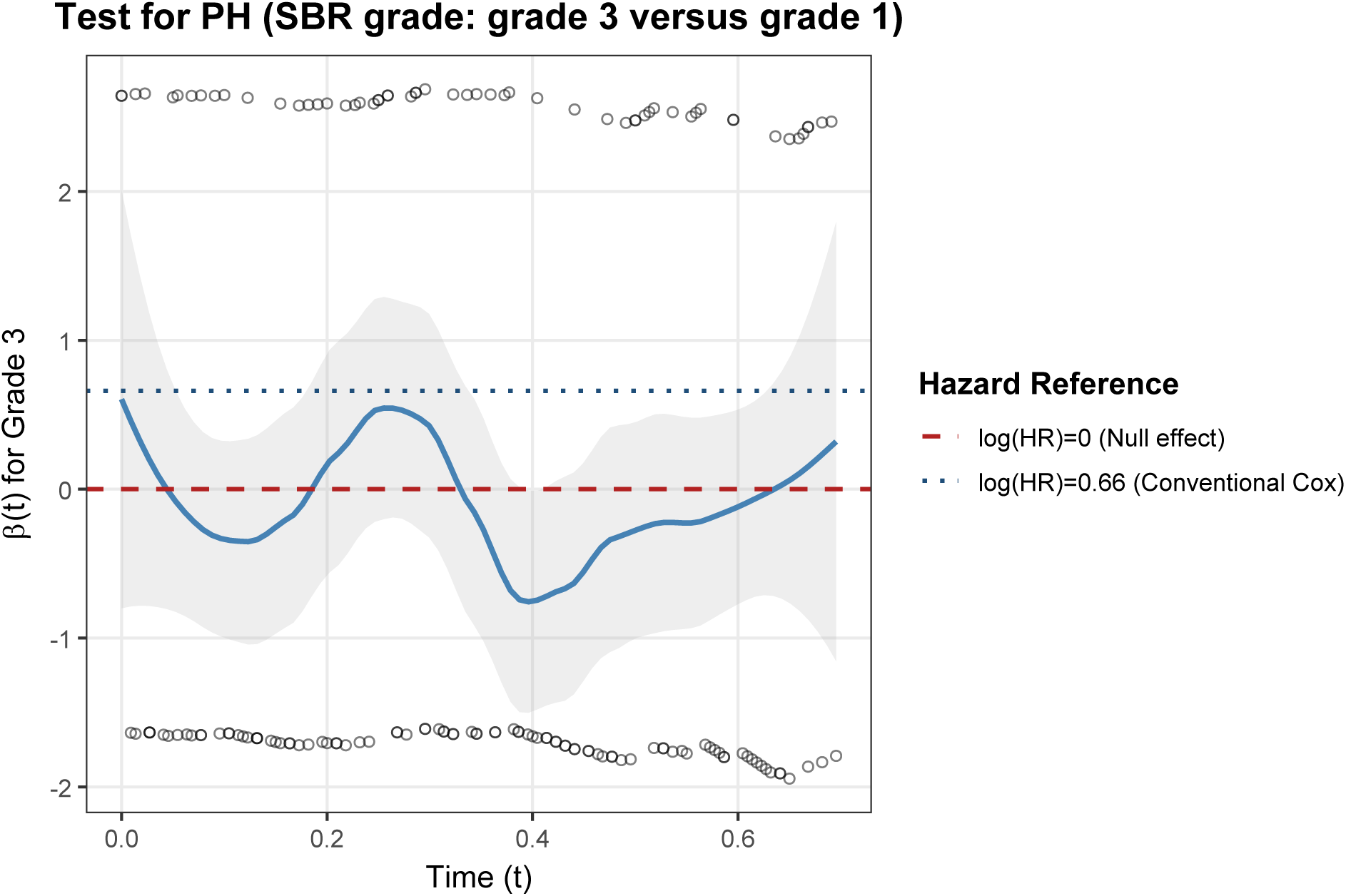
Scaled Schoenfeld residuals over time for SBR grade (grade 3 vs grade 1) with a smoothed trend and 95% confidence band.

#### 5.3.15 Martingale / Deviance Residual Plot

**• Technical Specification**

**– Purpose:** Diagnose the functional form of a continuous covariate in a Cox PH model by plotting cumulative martingale residuals against the covariate (non-random structure suggests nonlinearity / misspecification) [215, 216].
**– Data:** time (follow-up), status (event indicator 0/1), continuous covariate (log_bili or log(bilirubin)); optional additional covariates.
**– Assumption:** Cox proportional hazards framework with correctly specified covariate functional form (this plot checks that piece).
**– R Packages:** survival, stats, ggplot2.
**• Applications**

**– Use-case 1.** Functional-form check (primary): If the observed cumulative residual curve shows systematic curvature vs continuous covariate (log_bili or log(bilirubin)), consider restricted cubic splines or piecewise-linear effects.
**– Use-case 2.** Model refinement: Use this diagnostic to justify transforming continuous prognostic factors (e.g., tumor burden markers) before final Cox modeling.
**• Visualization**

**Figure 52:**
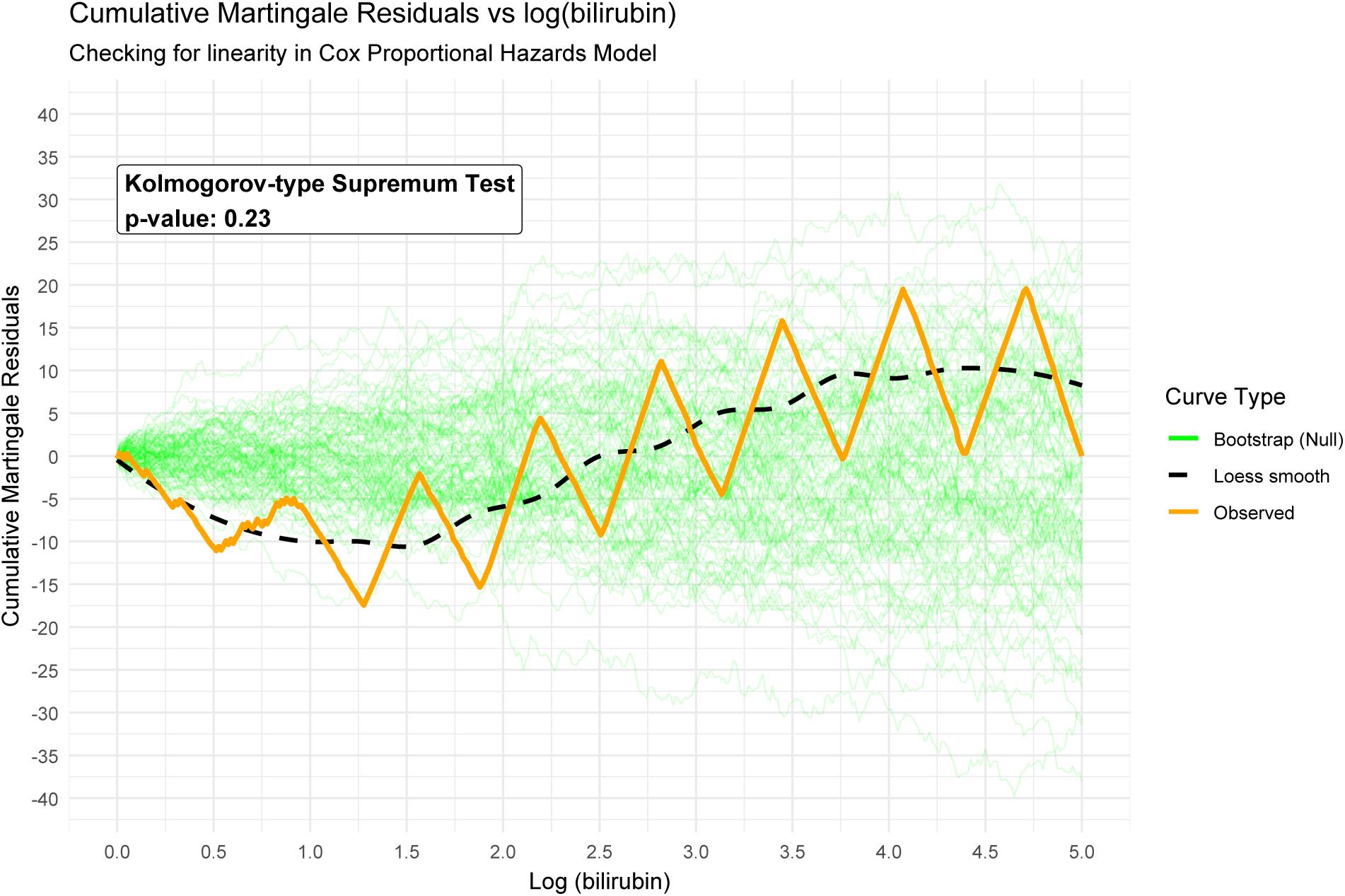
Cumulative martingale residuals plotted against log(bilirubin) from a Cox proportional hazards model (simulated example). A largely structureless, mean-zero wandering pattern is consistent with an adequate linear covariate specification; systematic curvature or sustained drift suggests the covariate functional form may require transformation (e.g., spline, log/threshold) or additional terms.

#### 5.3.16 RMST Difference vs Time

**• Technical Specification**

**– Purpose:** Visualize how the restricted mean survival time difference ΔRMST(*τ*) = RMST_Intervention_(*τ*) − RMST_Control_(*τ*) evolves as the restriction time *τ* varies [217, 218].
**– Data:** time (follow-up time), status (1=event, 0=censored), arm (Intervention vs Control), and a grid of restriction times *τ*.
**– Assumption:** Independent (non-informative) right-censoring and *τ* lies within the observed time window for stable estimation.
**– R Packages:** survival, ggplot2, dplyr, tidyr, broom.
**• Applications**

**– Use-case 1.** Non-PH settings: Summarize treatment benefit when hazards cross or vary over time (immunotherapy delayed effects).
**– Use-case 2.** Clinical interpretation: Identify the time horizons *τ* where benefit emerges or dissipates.
**• Visualization**

**Figure 53:**
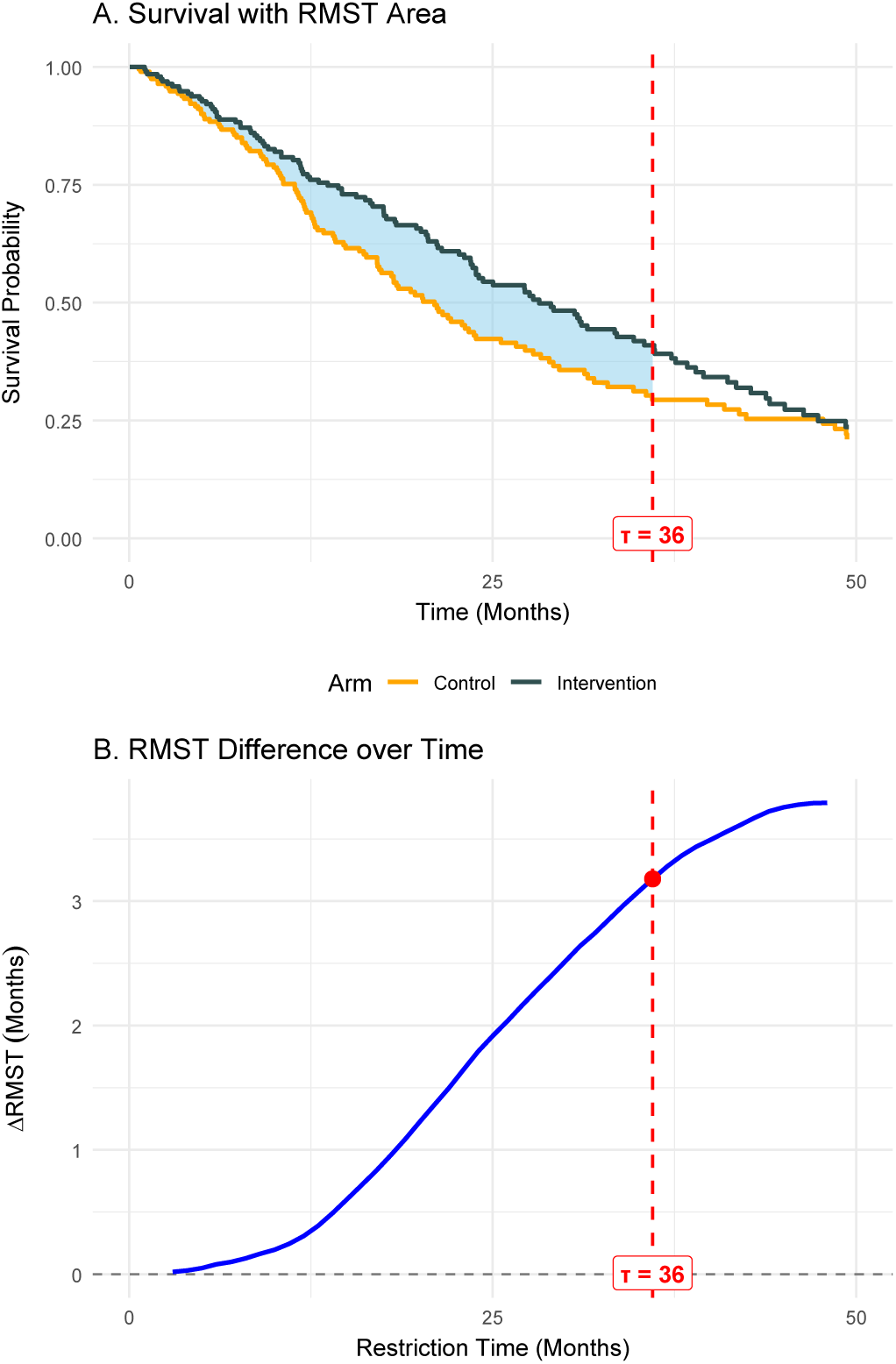
The RMST difference ΔRMST(*τ*) (Intervention – Control) versus restriction time *τ*; positive values indicate average survival benefit up to the time *τ*.

#### 5.3.17 Multi-state Stacked Probability Plot

**• Technical Specification**

**– Purpose:** Visualize current state probabilities over follow-up time for multiple clinical states (e.g., CR/Relapse/Death) and compare trajectories between treatment arms within a stratum (here: ITD-Low) [219, 220].
**– Data:** time (months), arm (e.g., Midostaurin, Placebo), state (e.g., CR, Relapse, Death), p_state (state occupancy probability), stratum (e.g., ITD-Low).
**– Assumption:** State-occupancy probabilities are estimated from a multi-state model and interpreted marginally over the cohort at each time *t*.
**– R Packages:** ggplot2, dplyr, tidyr, mstate, msm.
**• Applications**

**– Use-case 1.** Trial reporting (multi-state endpoints): Communicate how treatment shifts the cohort through remission, relapse, and death over follow-up (beyond a single Kaplan–Meier curve).
**– Use-case 2.**Health economics / state-based modeling: Provide direct inputs/validation visuals for Markov or partitioned-survival cost-effectiveness structures where time-in-state drives costs/utilities.
**• Visualization**

**Figure 54:**
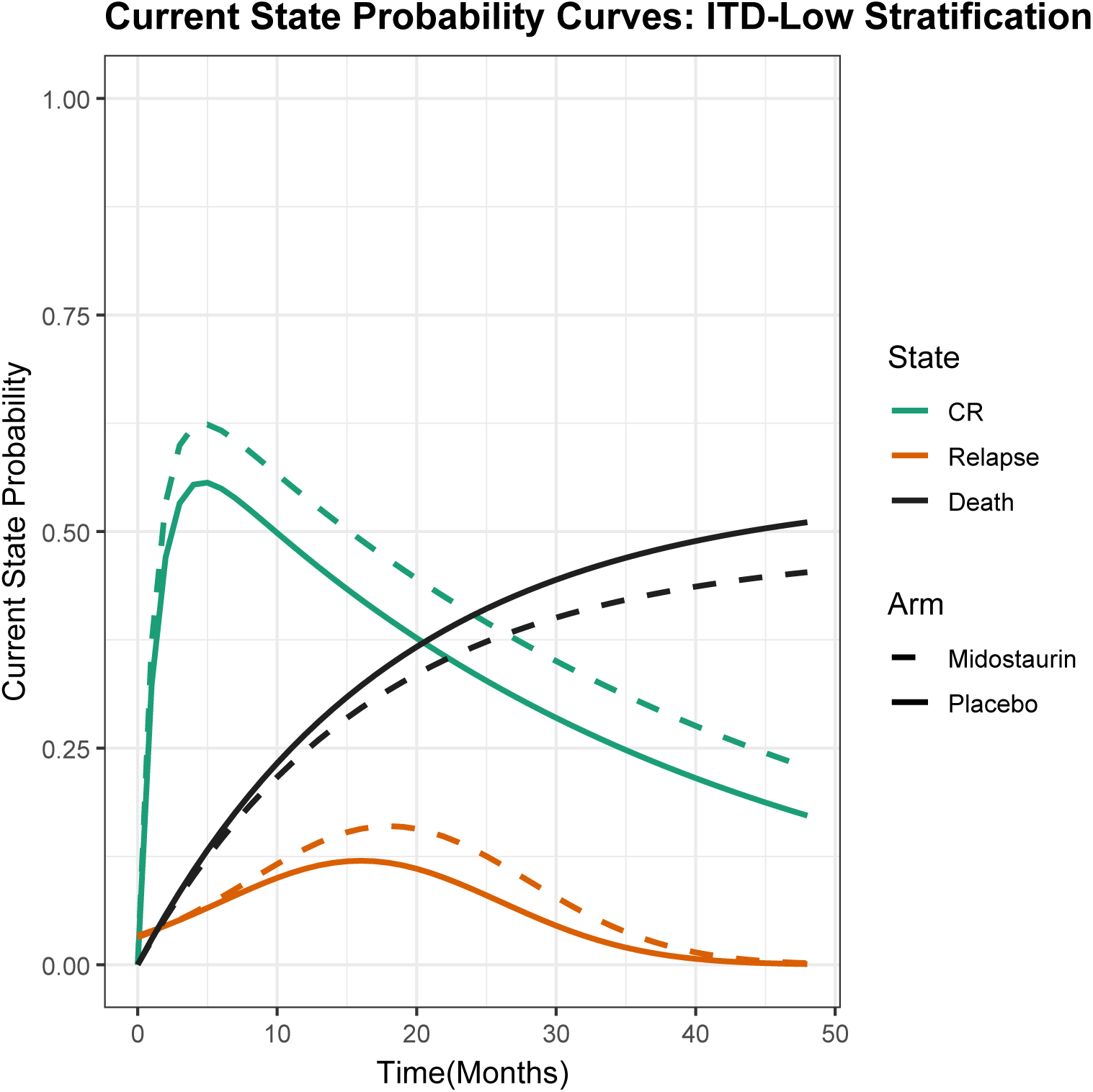
Multi-state current state probability curves (ITD-Low stratification). The panel displays simulated current state occupancy probabilities over 0–48 months for three clinically interpretable states—CR (complete remission), Relapse, and Death—shown separately for Midostaurin (dashed) versus Placebo (solid), (CR = complete remission; ITD = internal tandem duplication.).

#### 5.3.18 Best Overall Response (BOR) Distribution Plot

**• Technical Specification**

**– Purpose:** Summarize and compare best overall response (BOR) categories between treatment arms (RECIST-style endpoints) [221, 222].
**– Data:** arm, response_cat (CR/PR/SD/PD/NE), n (counts) or pct (percentages), and N_arm (arm totals).
**– Assumption:** BOR categories are mutually exclusive and exhaustive within each arm (each patient contributes exactly one BOR).
**– R Packages:** ggplot2, dplyr, tidyr, patchwork, scales.
**• Applications**

**– Use-case 1.** Study Reporting: [a] Efficacy summary: Quick between-arm comparison of categorical tumor response endpoints (RECIST-style); [b] Derived endpoints: Compute and report ORR = CR + PR and DCR = CR + PR + SD by arm/subgroup.
**– Use-case 2.** Safety/interpretation context: Pair with duration-based plots (e.g., swimmer) to distinguish short-lived PR/SD from durable benefit.
**• Visualization**

**Figure 55:**
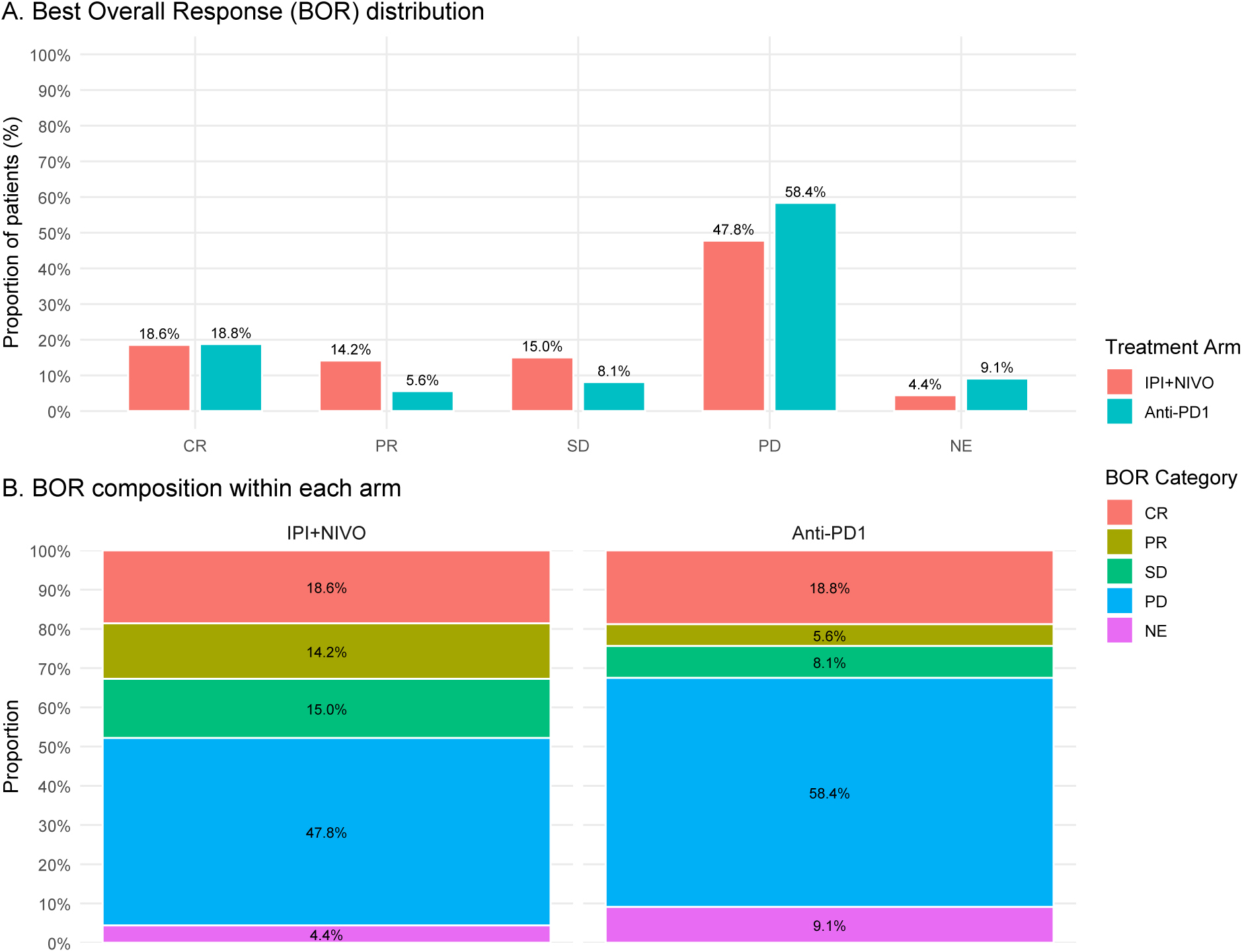
Distribution of best overall response (BOR) in patients treated with ipilimumab plus nivolumab (IPI+NIVO; n = 33) versus anti–PD-1 monotherapy (n = 22). BOR categories include complete response (CR), partial response (PR), stable disease (SD), progressive disease (PD), and not evaluable (NE). Panel A shows the percentage of patients in each BOR category by arm; Panel B shows the within-arm BOR composition derived from the same counts.

#### 5.3.19 Lollipop Mutation Map

**• Technical Specification**

**– Purpose:** Visualize mutation hotspots along a linear gene/protein coordinate and compare how hotspot mutations distribute across tumor types [223, 224].
**– Data:** [a] Lollipop panel: position (bp or aa index), n_patients (count), optionally mutation_label and is_hotspot; [b] Hotspot composition panel: hotspot (e.g., −124/-146/-138/Other), cancer_type, percent.
**– Assumption:** Positions are measured on a common reference coordinate system (e.g., bp relative to transcription start site), and counts represent unique patients/events per position.
**– R Packages:** ggplot2, dplyr, tidyr, patchwork, scales.
**• Applications**

**– Use-case 1.** Reporting Study: [a] Hotspot discovery and reporting: Identify and communicate recurrent somatic mutation positions and their relative patient burden; [b] Biology/annotation linkage: Overlay functional regions (promoter motifs/domains) to interpret whether hotspots cluster in biologically meaningful elements.
**– Use-case 2.** Clinical/RWE stratification: Compare hotspot prevalence and tumor-type mix across cohorts (trial vs RWE) to understand enrichment and generalizability.
**• Visualization**

**Figure 56:**
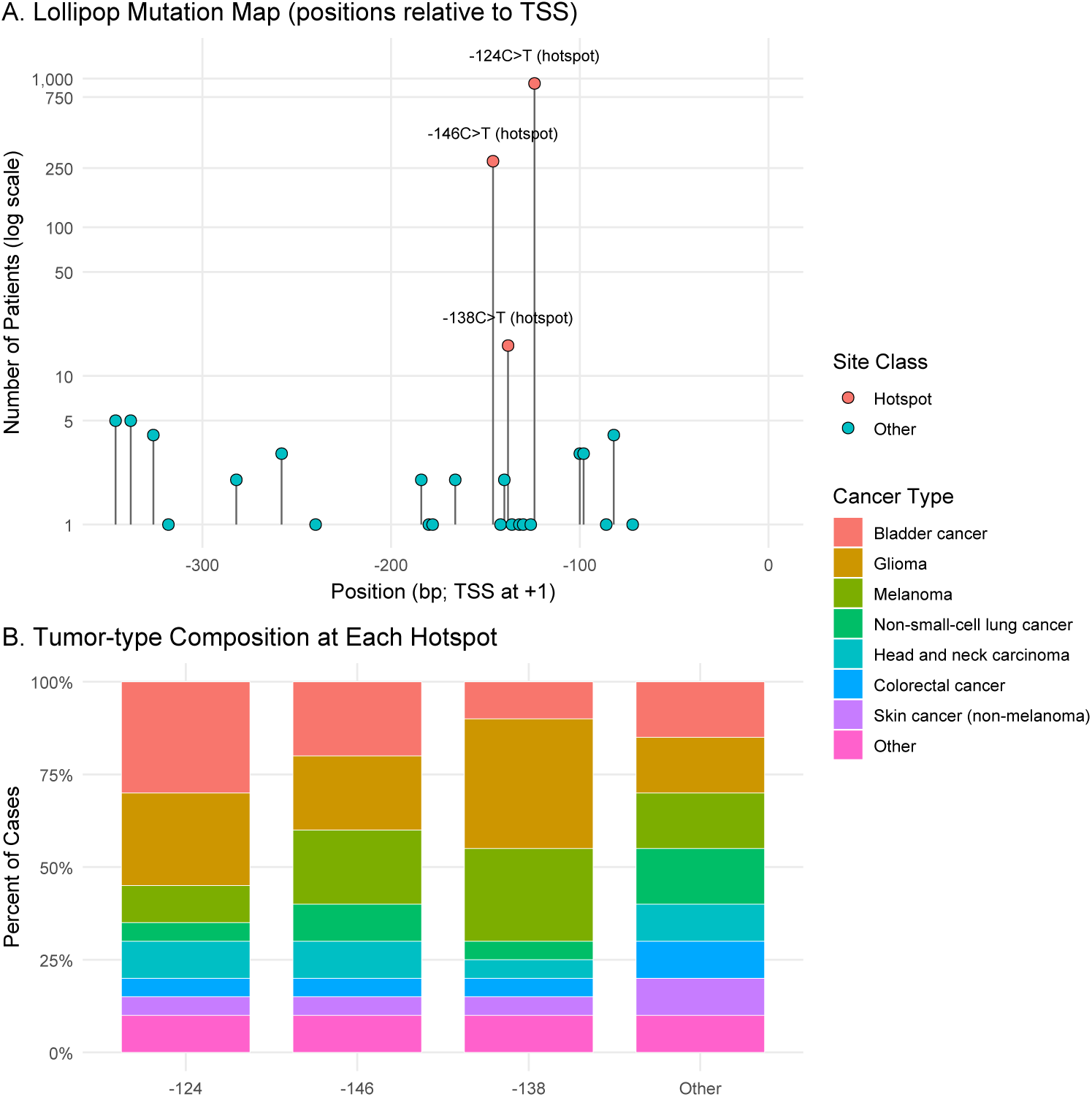
Lollipop mutation map showing the location and frequency of promoter mutations along a linear coordinate system (positions shown relative to the transcription start site, +1). The height of each lollipop indicates the number of patients with a mutation at that position (log scale), highlighting common hotspot sites (e.g., −124, −146, −138). The right panel summarizes the tumor-type composition (percent of cases) for each hotspot category.

#### 5.3.20 Volcano Plot

**• Technical Specification**

**– Purpose:** This plot answers which genes show the largest differential expression (log2 fold-change) with the strongest statistical evidence (-log10 p-value) between two groups (e.g., CPR vs non-CPR) [225, 226].
**– Data:** gene_id, log2FC, p_value (or padj), and a derived group indicator for üp in CPR” vs üp in non-CPR”.
**– Assumption:** Differential expression testing is valid (e.g., properly normalized expression; multiplicity addressed if using padj).
**– R Packages:** ggplot2, ggrepel, dplyr.
**• Applications**

**– Use-case 1.** Biomarker Discovery: Identify candidate transcriptomic biomarkers that distinguish responders (CPR) vs non-responders, then prioritize genes with large | log_2_(*FC*)| and strong significance for downstream validation (qPCR/IHC, independent cohort).
**– Use-case 2.** Mechanism-of-action / Pathway hypothesis: Use the top CPR-upregulated vs non-CPR-upregulated genes as inputs to pathway enrichment (e.g., IFN-*γ* signaling, cytotoxic T-cell programs) to generate mechanistic hypotheses about why response differs.
**• Visualization**

**Figure 57:**
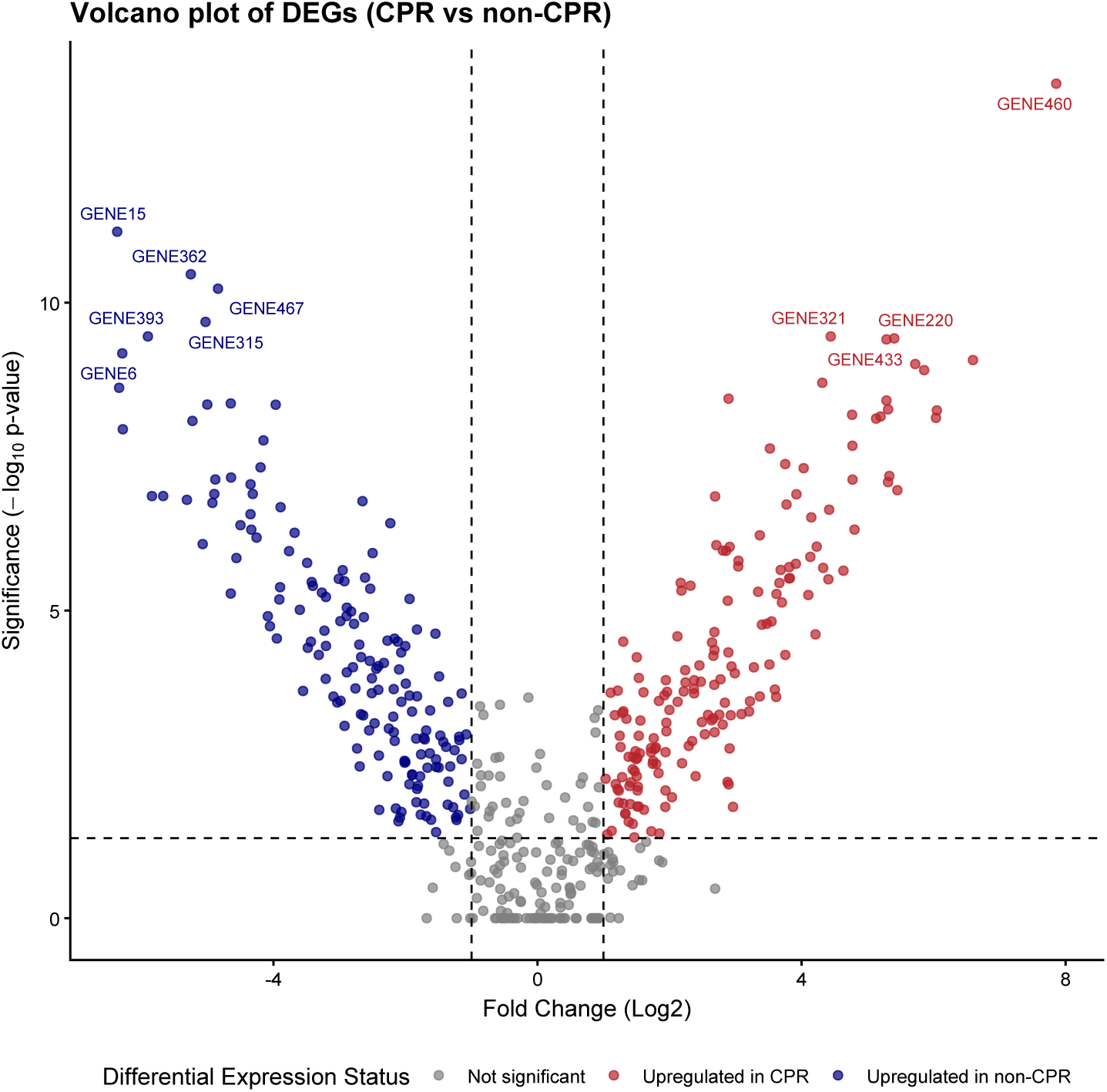
Volcano plot showing differentially expressed genes (DEGs) between CPR and non-CPR pretreatment tumors. The x-axis shows log2 fold change and the y-axis shows -log10(p-value). Genes meeting the significance and effect-size thresholds are highlighted as upregulated in CPR (red) or upregulated in non-CPR (blue); non-significant genes are shown in grey. Dashed lines indicate the pre-specified fold-change and p-value cutoffs.

#### 5.3.21 Longitudinal Patient Reported Outcome(PRO) Plot

**• Technical Specification**

**– Purpose:** Visualize and compare mean change from baseline in a PRO (e.g., PRO-CTCAE symptom severity) over follow-up between treatment arms, with uncertainty via 95% CIs [227, 228].
**– Data:** id, arm (dose group), week (visit time), pro_change (change-from-baseline score; can be symptom severity, interference, etc.).
**– Assumption:** Missingness is approximately ignorable for the chosen summary (e.g., MMRM/estimand-based mean); CIs reflect sampling uncertainty at each visit.
**– R Packages:** ggplot2, dplyr, tidyr (optional reshaping).
**• Applications**

**– Use-case 1.** Tolerability profiling: Compare symptom trajectories (e.g., fatigue, constipation, neuropathy) between doses/arms to quantify net tolerability beyond CTCAE.
**– Use-case 2.** Dose selection / regimen optimization: Identify sustained divergence in PRO burden (clinically meaningful worsening) that may favor a lower dose despite similar efficacy.
**• Visualization**

**Figure 58:**
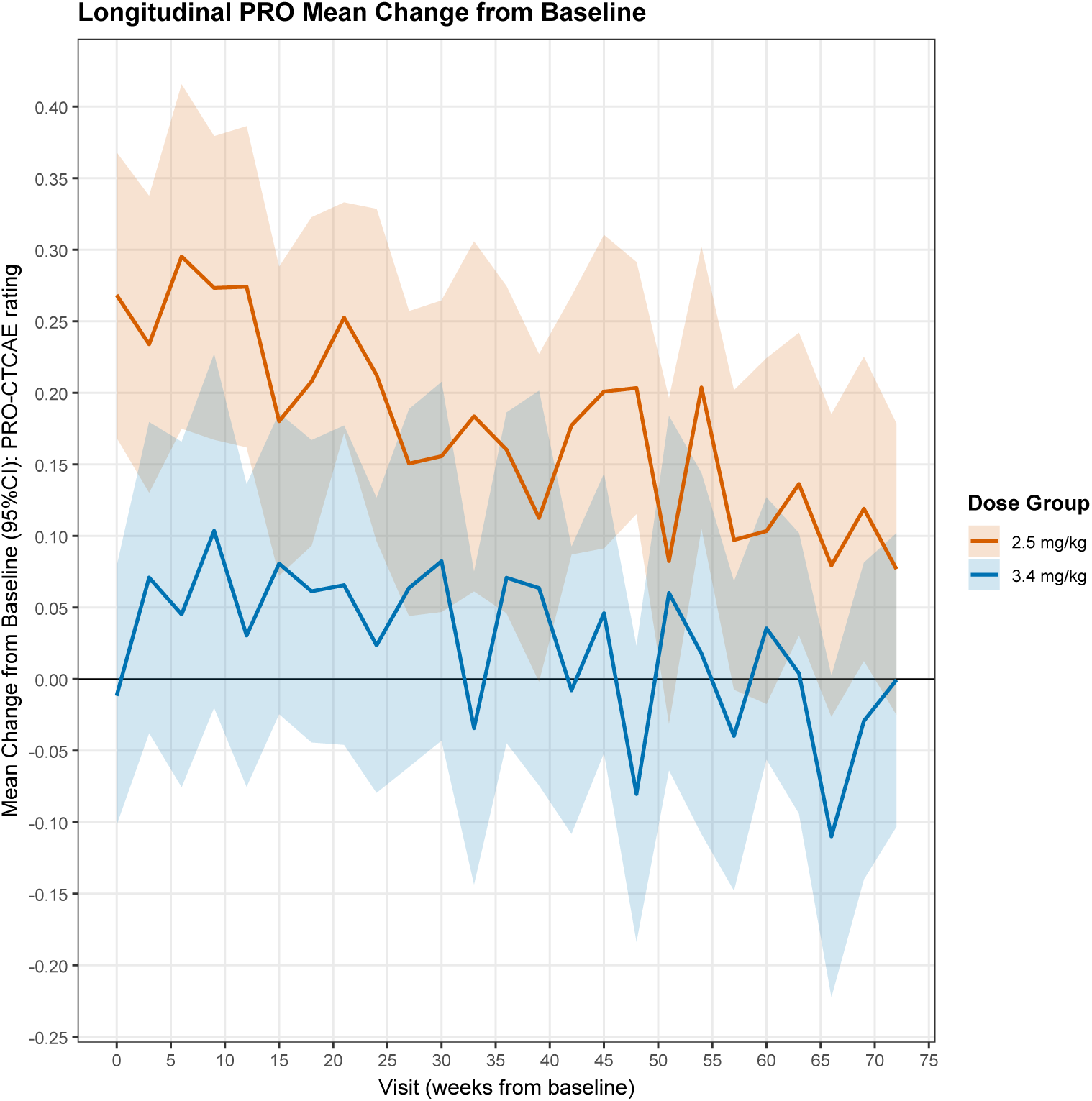
Longitudinal patient-reported outcome (PRO) mean change from baseline over follow-up by dose group, with 95% confidence bands. Solid lines show arm-specific mean change from baseline at each visit; shaded ribbons indicate pointwise 95% CIs. Values above 0 indicate worse symptom burden relative to baseline (direction can be flipped depending on instrument coding).

#### 5.3.22 Cumulative Threshold Response Curve

**• Technical Specification**

**– Purpose:** Visualize, for each treatment arm, the cumulative % of patients achieving at least a given improvement (or change) on a continuous PRO scale across all possible thresholds (not one responder cut) [228, 229].
**– Data:** arm, change from baseline, time (continuous), optional clinically meaningful thresholds (vertical reference lines).
**– Assumption:** PRO change is measured on a comparable continuous scale across arms; the CDF meaningfully summarizes heterogeneity of response.
**– R Packages:** ggplot2.
**• Applications**

**– Use-case 1.** Responder analysis without an arbitrary cut: Report what fraction of patients meet any clinically relevant threshold (e.g., ≤ −2.5) while still showing the full distribution.
**– Use-case 2.** Treatment differentiation across the entire outcome scale: Detect whether an effect is driven by a minority of “super-responders” vs. a broad shift of the whole distribution.
**• Visualization**

**Figure 59:**
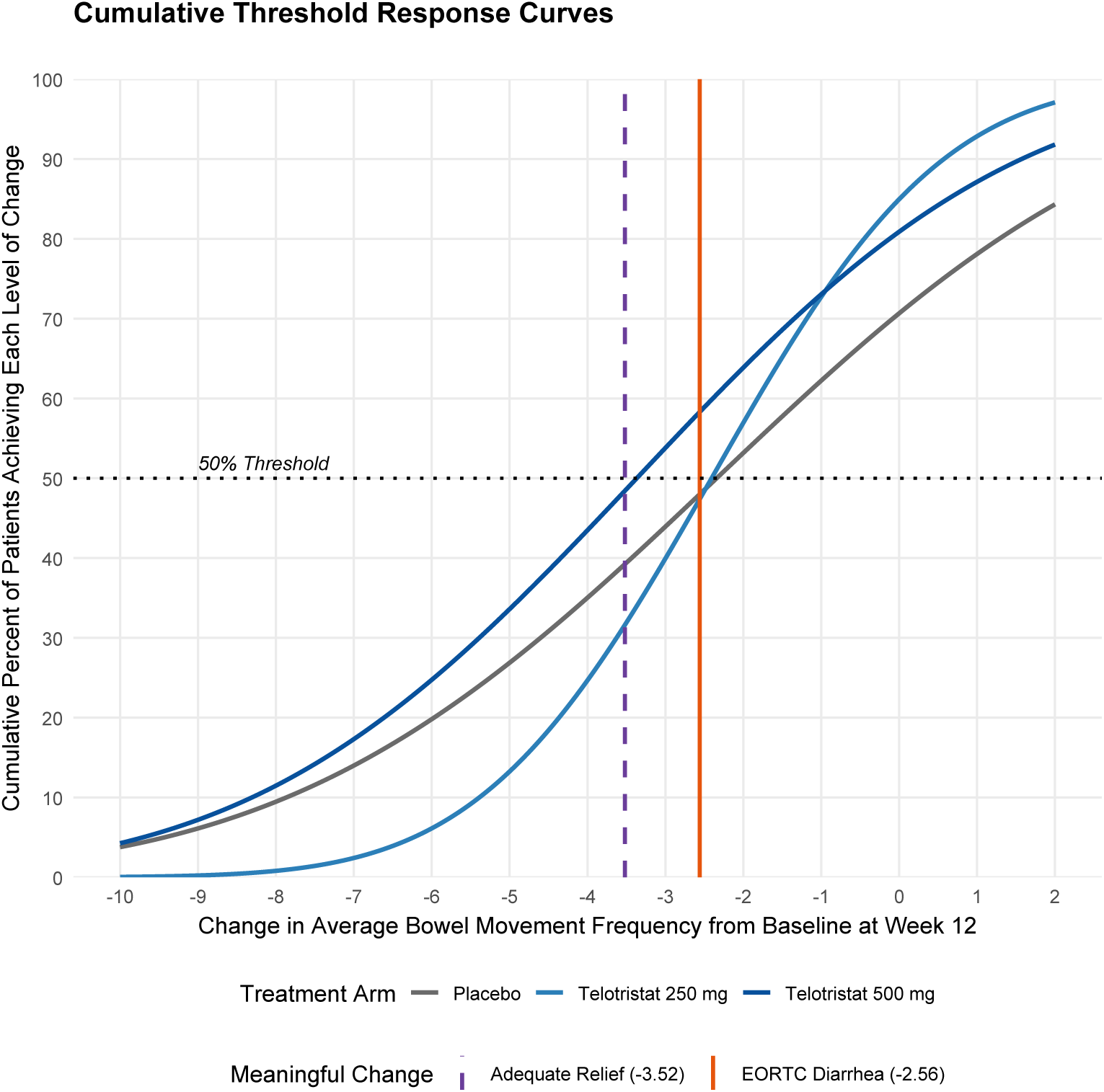
Cumulative threshold response (CDF) of change in average bowel-movement frequency from baseline at Week 12 by treatment arm. Curves show the cumulative proportion of patients achieving each level of change across the full threshold range. Vertical reference lines indicate example clinically meaningful change thresholds (Adequate Relief and EORTC Diarrhea anchors).

#### 5.3.23 Time to Deterioration(TtD) in Quality of Life (QoL) Score

**• Technical Specification**

**– Purpose:** Visualize and compare how long patients maintain baseline QoL (i.e., time until clinically meaningful deterioration) between treatment arms [230, 231].
**– Data:** time (months), event (1 = deterioration, 0 = censored), arm (group), optional visit_schedule / MCID definition.
**– Assumption:** Independent (non-informative) censoring; deterioration is defined a priori (e.g., ≥ 10-point worsening on an EORTC scale).
**– R Packages:** survival, ggplot2, ggplot2.
**• Applications**

**– Use-case 1.** Compare QoL durability between treatment arms when OS/PFS are similar, to quantify “net clinical benefit” beyond survival.
**– Use-case 2.** Support tolerability labeling by estimating medians/landmarks for time-to-worsening in key domains (fatigue, pain, bowel function).
**• Visualization**

**Figure 60:**
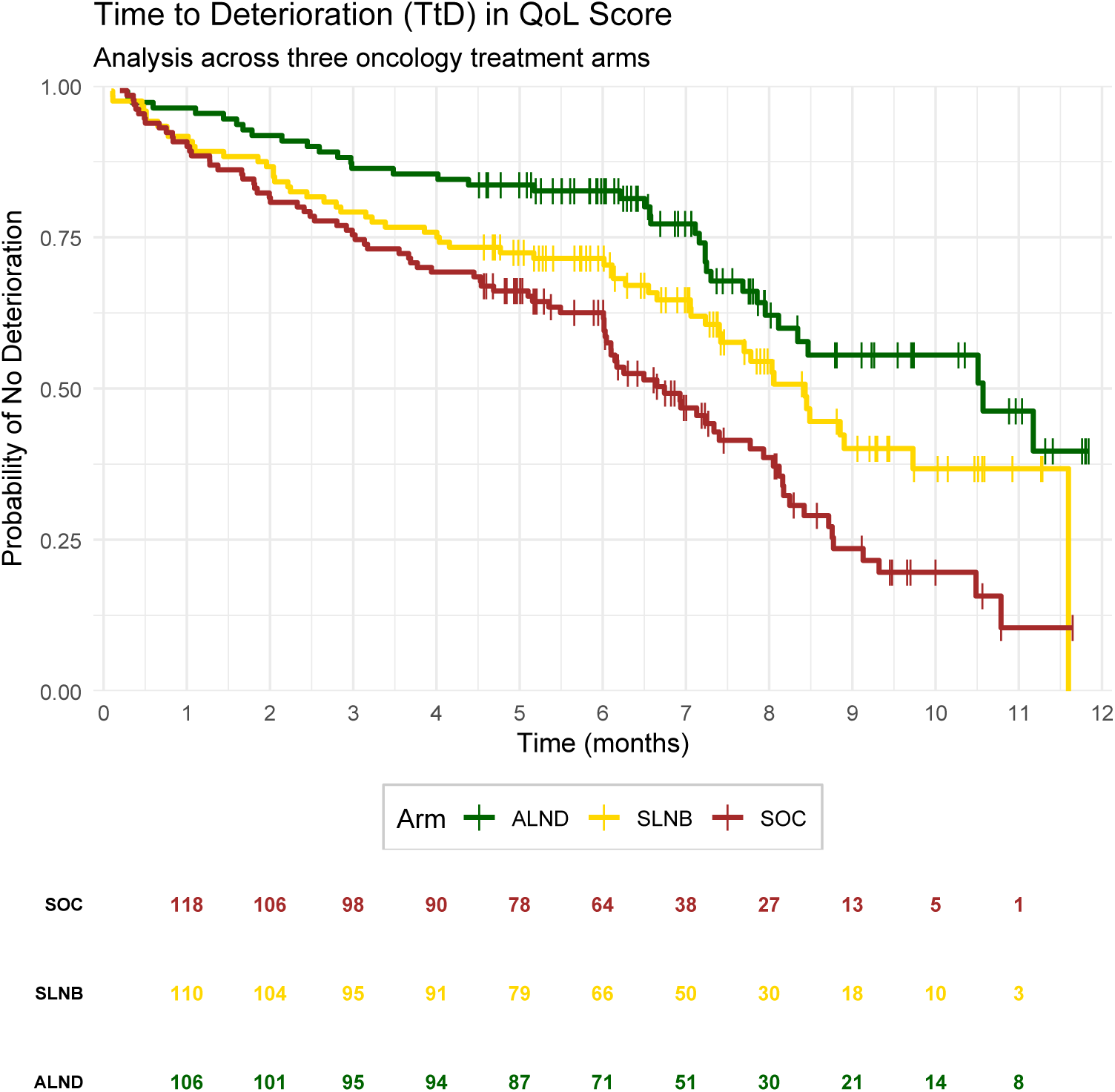
Time to clinically meaningful deterioration in quality-of-life score (TtD). Kaplan–Meier curves show the probability of remaining free of deterioration over time for two treatment arms (example: ALND vs SLNB). Tick marks indicate censoring; numbers at risk are shown at 3, 6, 9, and 12 months. Deterioration is defined a priori using a clinically meaningful threshold (e.g., ≥ 10-point worsening on a validated PRO scale).

#### 5.3.24 Restricted Cubic Spline (RCS) effect plot

**• Technical Specification**

**– Purpose:** Visualize a nonlinear effect of a continuous predictor *x* on an outcome (often hazard ratio in a Cox model) without forcing linearity [232].
**– Data:** time (follow-up time), status (event indicator: 1=event, 0=censored), continuous predictor x (e.g., biomarker, lab value, age)
**– Assumption:** Cox model structure with smooth, piecewise-cubic effect in *x* and linear tails beyond boundary knots (restricted cubic spline), optional adjusters *z*1*, z*2, · · ·.
**– R Packages:** ggplot2, survival, rms.
**• Applications**

**– Use-case 1.** Biomarker risk modeling (oncology): Model continuous MRD, cytokines, glucose, etc., against relapse/NRM/OS without arbitrary dichotomization (and test whether the effect is nonlinear).
**– Use-case 2.**Clinical cut-point support: If a threshold is needed operationally, use the RCS curve to justify a candidate region, rather than picking a cut-point blindly (with caution about information loss).
**• Visualization**

**Figure 61:**
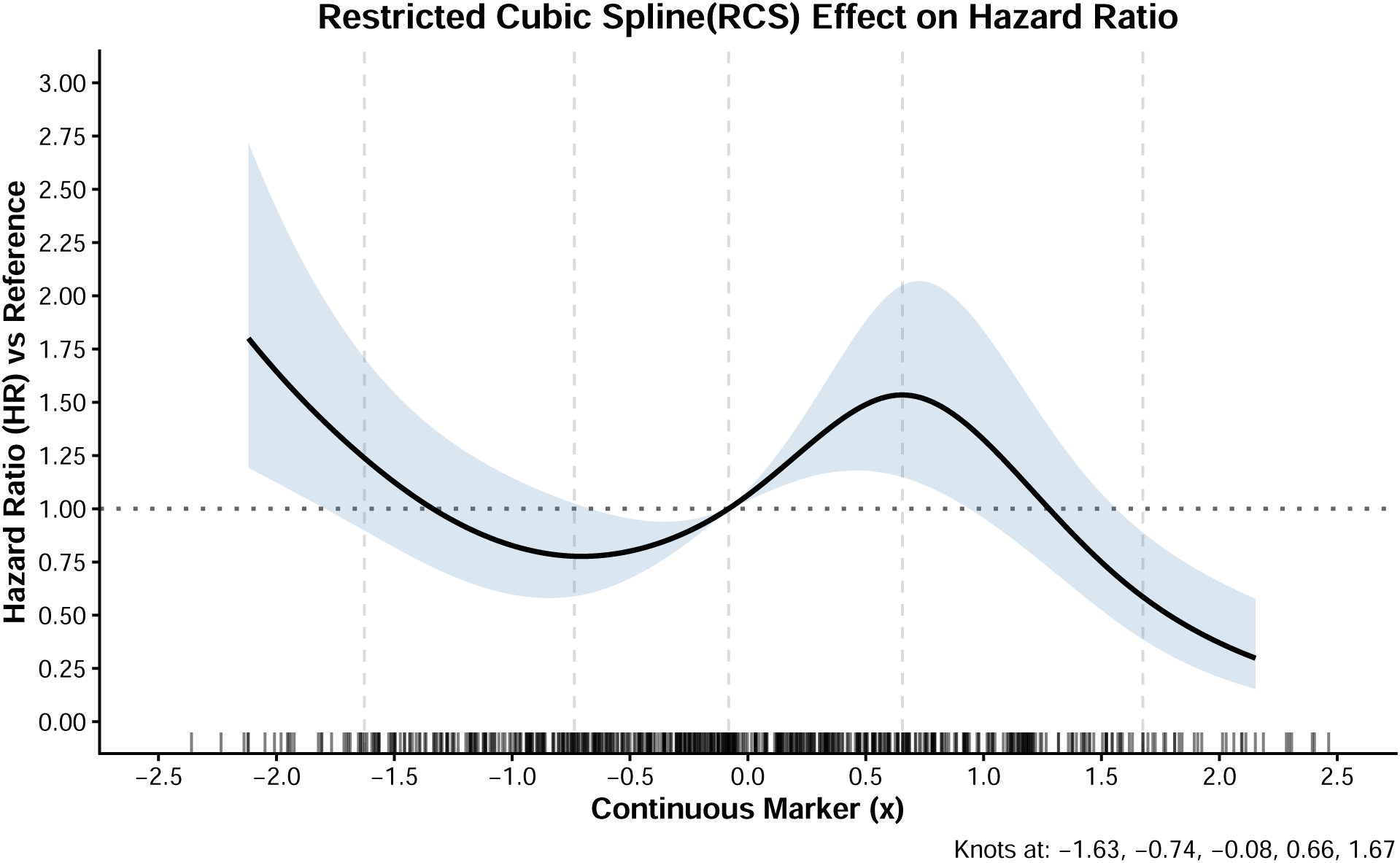
Restricted cubic spline (RCS) effect plot for a continuous marker. The solid curve shows the estimated hazard ratio *HR*(*x*) from a Cox model with an RCS term for *x*, referenced to *x*_0_ (median). Dashed curves indicate the 95% confidence interval. Vertical dashed lines mark spline knot locations (5th, 25th, 50th, 75th, 95th percentiles), with linear tails beyond the boundary knots.

#### 5.3.25 Upset Plot

**• Technical Specification**

**– Purpose:** Visualize the intersection structure of multiple binary subgroup-defining covariates (e.g., pf/bm/stage/hx) and compare intersection sizes between treatment arms (Chemotherapy vs. Surgery) [233, 234].
**– Data:** id, several 0/1 set-membership columns (e.g., pf, bm, stage, hx), plus arm (Chemotherapy/Surgery).
**– Assumption:** Sets are well-defined binary indicators (each subject either belongs to a set or not).
**– R Packages:** ComplexUpset, ggplot2, patchwork, dplyr.
**• Applications**

**– Use-case 1.** Subgroup feasibility in trials: Identify which combinations of eligibility/clinical features have sufficient sample size to support credible subgroup analyses.
**– Use-case 2.** Subgroup feasibility in trials: Identify which combinations of eligibility/clinical features have sufficient sample size to support credible subgroup analyses.
**• Visualization**

**Figure 62:**
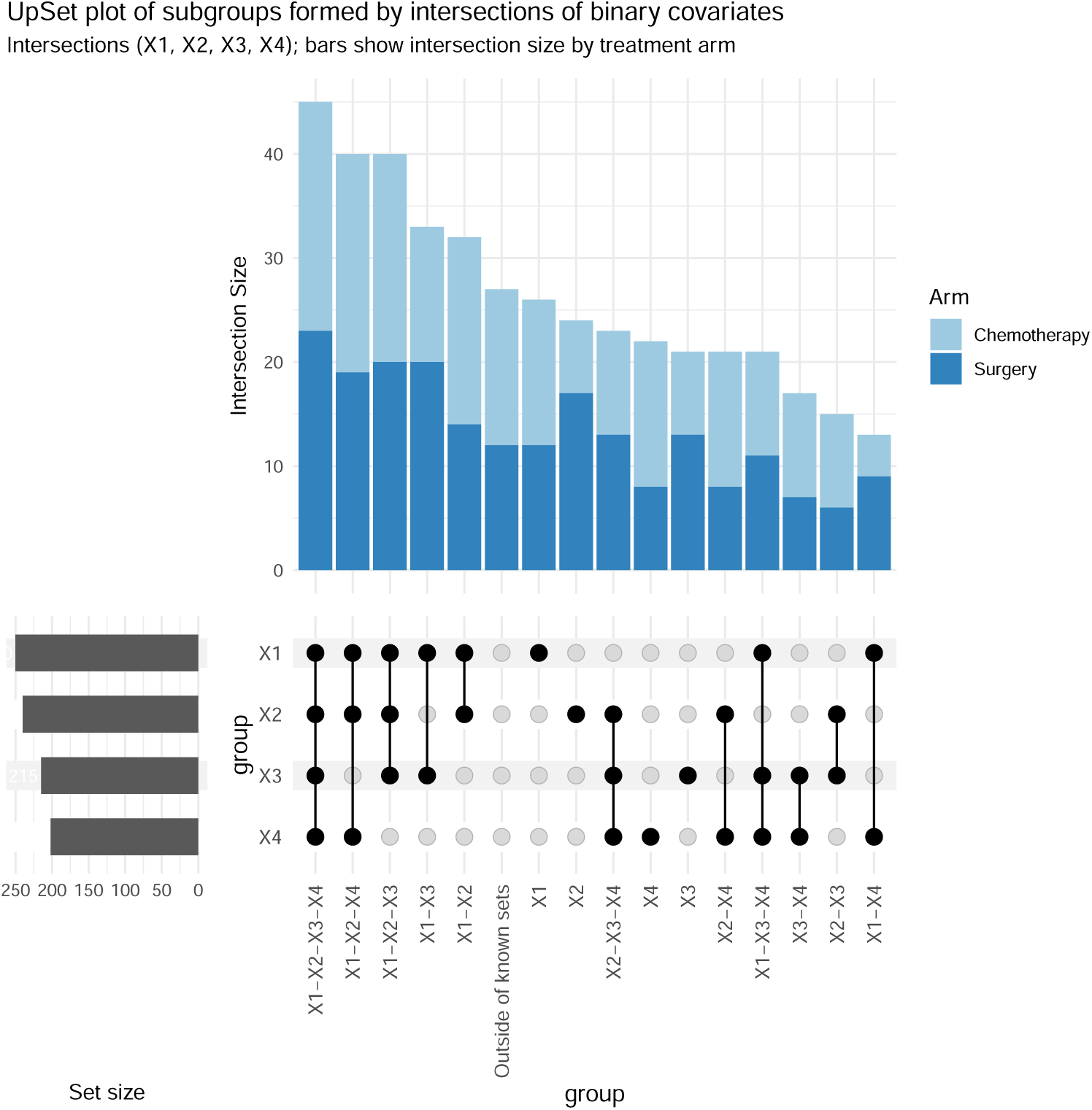
UpSet plot displaying the subgroups formed by intersections of binary subgroup-defining covariates (pf, bm, stage, hx). The vertical bars show intersection sizes, stacked by treatment arm (Control vs Experimental), and the horizontal bars show marginal set sizes for each covariate.

## 6 Tutorial: fishplot for Tumor Evolution

### 6.1 What is a fishplot?

A *fishplot*-common in both clinical trials and real world evidence studies-visualizes tumor clonal evolution over time by drawing each subclone as a polygon whose height represents its prevalence; polygons are arranged to respect parent–child lineage and the chronological order of sampling [235, 236]. The intent is a publication-ready depiction of clonal remodeling (e.g., a therapy-induced bottleneck followed by relapse expansion).

### 6.2 Interpretation

At each timepoint, the vertical thickness of a clone is proportional to its estimated fraction (or an interpretable scale). Visual cues typically used in oncology narratives include (i) extinction (a clone shrinks to zero), (ii) selective sweep (a resistant lineage expands post-therapy), and (iii) branching (multiple descendants expanding in parallel).

**Figure 63:**
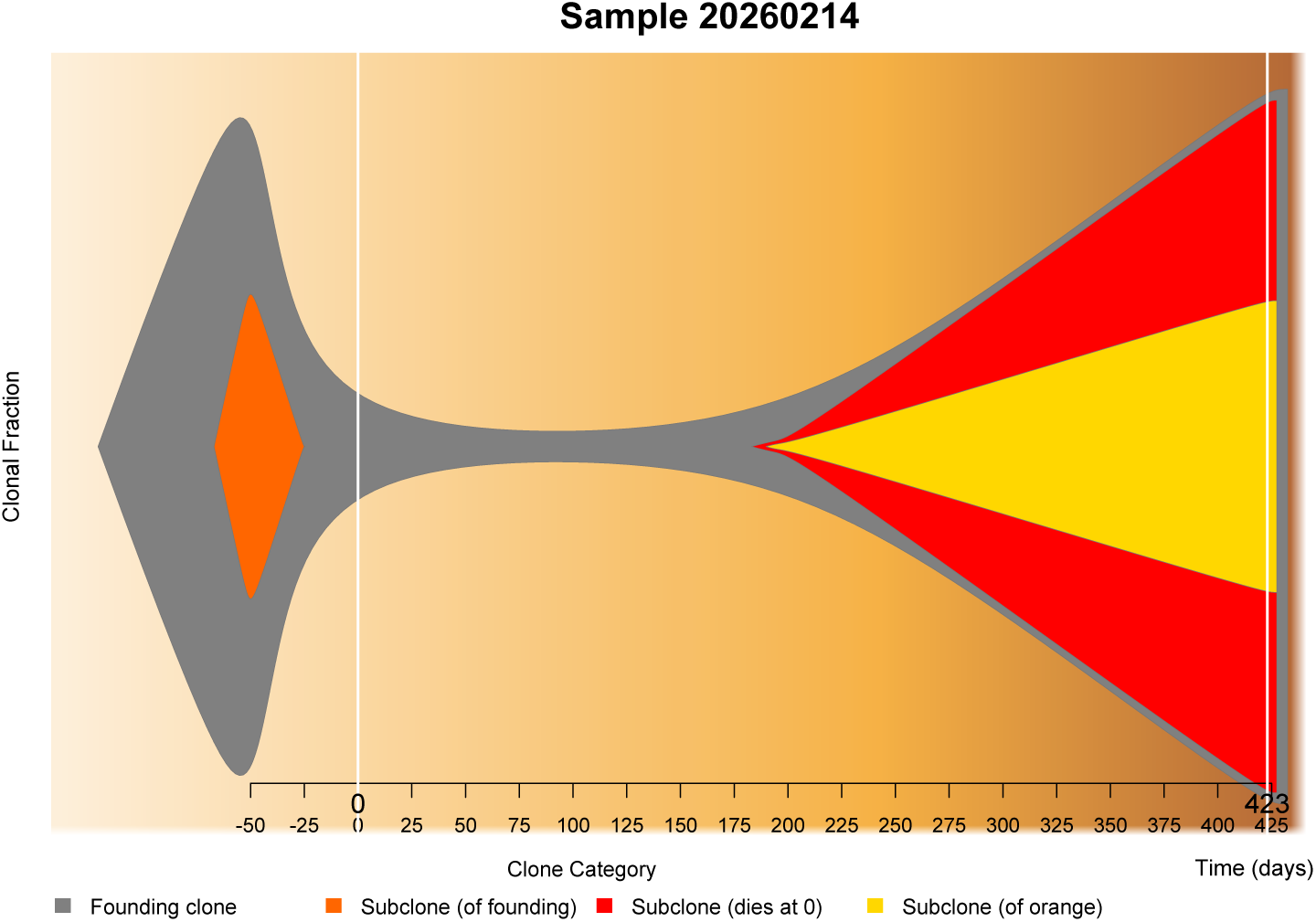
Visualizing tumor evolution with a fishplot package example (simulated). The fishplot displays subclonal fractions across longitudinal sampling timepoints, with polygons representing subclones and parent–child relationships encoding tumor phylogeny.

### 6.3 Plotting ‘fishplot’ in R Software

#### 6.3.1 Required Data Objects

At minimum, the programmers need the following data columns:

**(1) Timepoints** (e.g., days since diagnosis/therapy start).
**(2) Clonal fractions** (prevalence of each clone at each timepoint).
**(3) Parental relationships** (a lineage vector encoding descent).

#### 6.3.2 Core Assumption(s)

Fractions are *comparable across timepoints* (same unit scale; often each timepoint is normalized so fractions sum to 1 or 100).

#### 6.3.3 Tools

The provided implementation in R software uses fishplot [111] with supporting packages Hmisc [117], plotrix [124], and png [125]. For broader empirical data associated context on clonal evolution visualization techniques, see [235]; and, for an oncology example where clonal evolution is a central analytic story, see [236].

#### 6.3.4 Workflow Overview

The canonical workflow is:

CreateFishObject();→; layoutClones();→; fishPlot().

#### 6.3.5 Minimal Reproducible R Code

The following code is a clean, end-to-end example (including axis ticks, annotations, and export to high-resolution PNG + vector PDF):

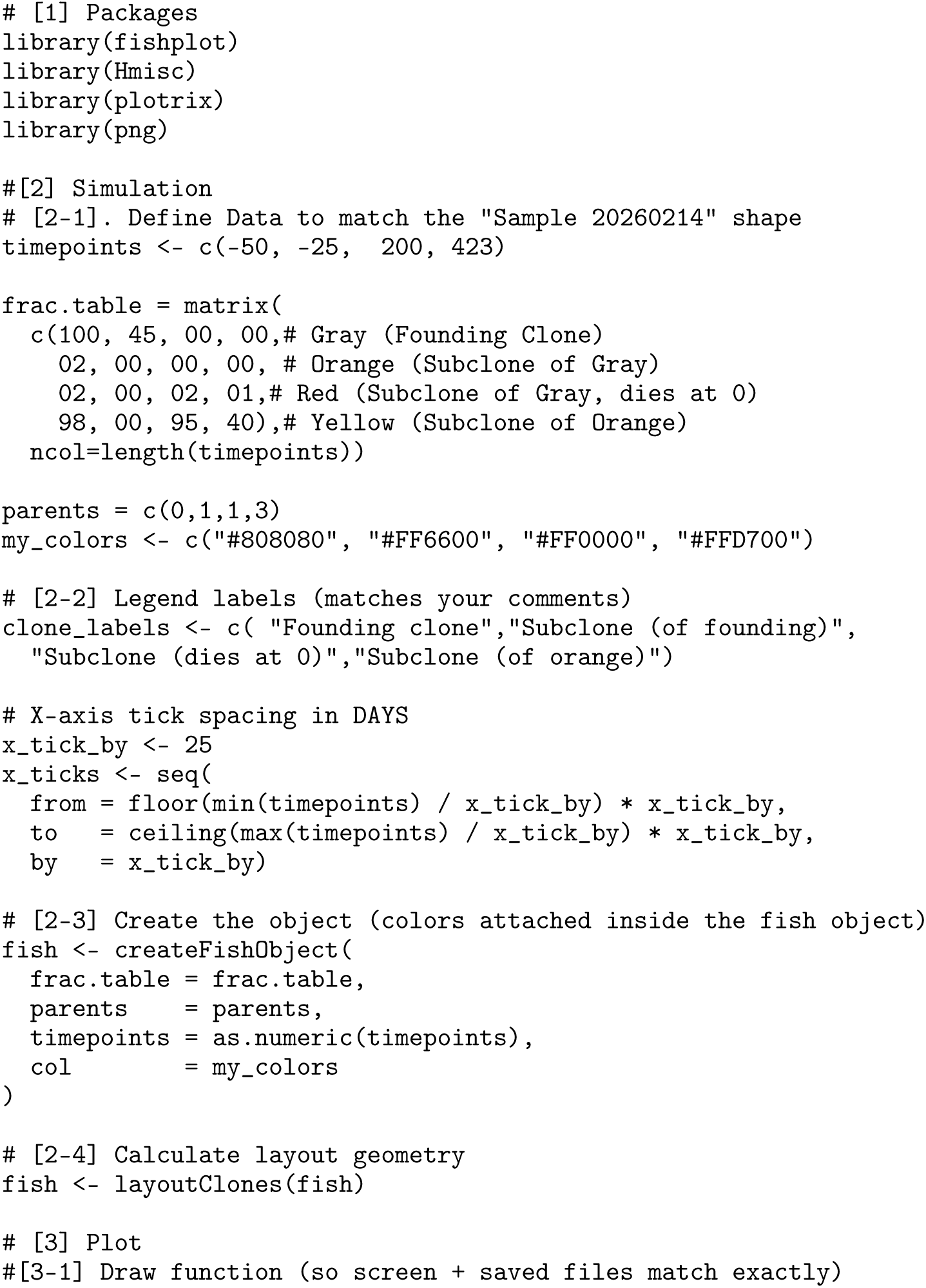
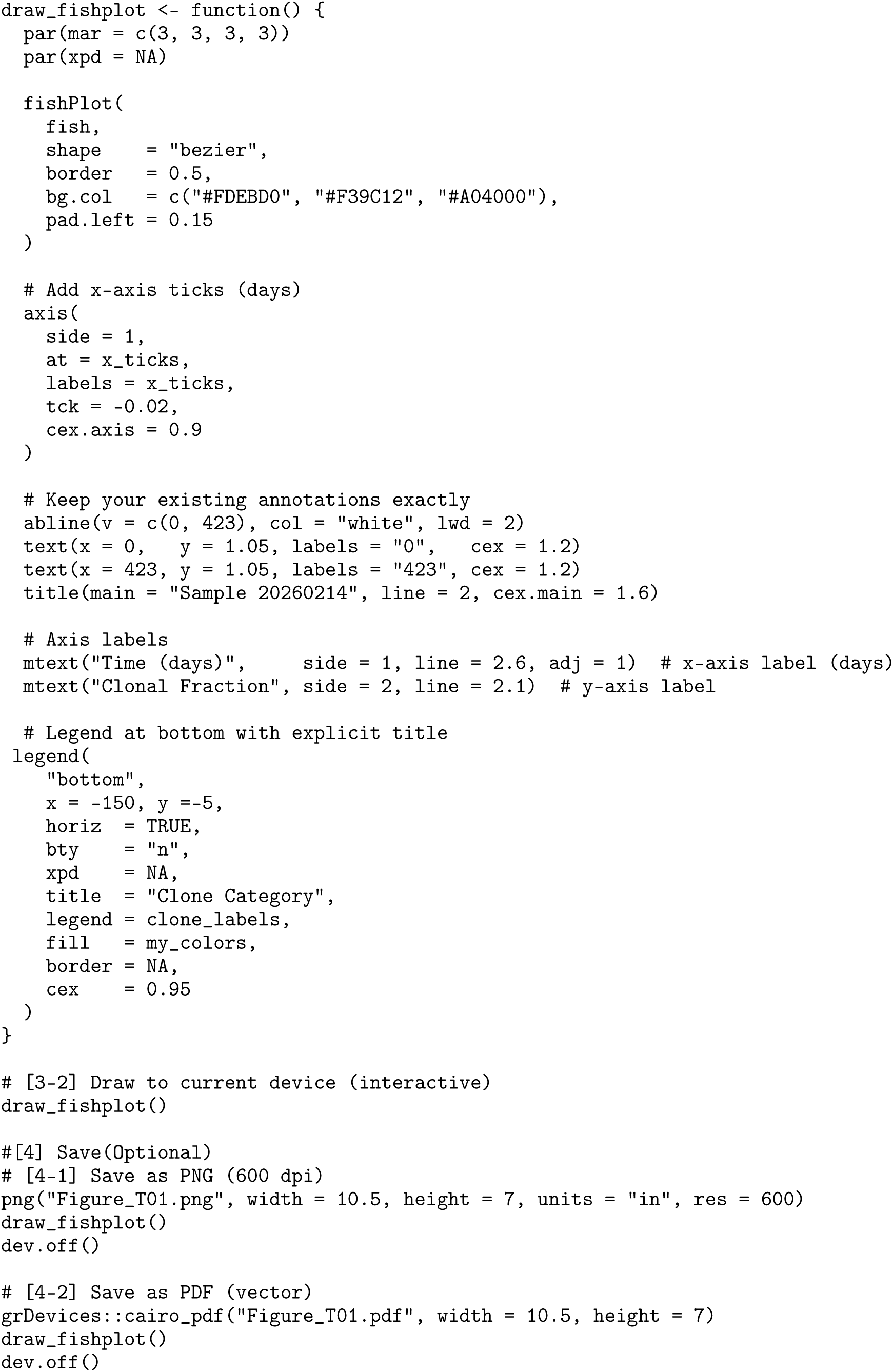

#### 6.3.6 General Comments and Guidance

1. **Fractions must be on a stable scale across samples.** If timepoints come from different assays/batches, reconcile normalization *before* plotting; otherwise the fishplot can look plausible while being quantitatively misleading.
2. **Lineage is a model input, not an inference.** The parents vector encodes your inferred phylogeny; uncertainty in phylogeny should be handled upstream (sensitivity analyses, alternative trees), not hidden inside the plot.
3. **Be explicit about time origin.** Negative times (as in the example) can be useful to show pre-therapy history, but label the origin (e.g., day 0 = treatment start) and mark key events with vertical lines/annotations.
4. **Separate storytelling from estimation.** The fishplot is the *communication layer*; document how clonal fractions were obtained and what constraints were applied (sum-to-1, purity adjustment, etc.).

#### 6.3.7 Best Practices

1. **QC checklist at each timepoint.** Verify (i) no negative fractions, (ii) fractions are comparable across timepoints, and (iii) the parent–child relationships are consistent with observed appearances/disappearances.
2. **Use consistent clone labels and a stable palette.** Keep labels short but biologically interpretable (driver mutation, resistance mechanism, or clonal ID). The example demonstrates a clear legend placed below the plot for publication layouts.
3. **Export both vector and raster.** Use a vector PDF for manuscripts and a high-resolution PNG (e.g., 600 dpi) for slides or journal submission systems that rasterize figures.
4. **Align plot narrative to oncology use-cases.** Fishplots are particularly effective for (i) therapy resistance narratives and relapse dynamics, and (ii) downstream visualization from subclonal calling + phylogeny pipelines.

#### 6.3.8 Pitfalls and Common Failure Modes

1. **Over-interpreting smooth geometry.** Smooth boundaries (e.g., shape=“bezier”) improve readability but do *not* imply continuous-time measurement; the only measured values are at the sampled timepoints.
2. **Inconsistent normalization across time.** If one timepoint is normalized to tumor-only cellularity and another to tumor+normal, visual changes can be artifacts.
3. **Ambiguous “clone death”.** A clone shrinking to zero might reflect detection limits rather than true extinction. If relevant, annotate detection thresholds or state them in the caption/methods.
4. **Legend/annotation clipping.** When placing legends outside the main panel, enable drawing outside the plotting region (e.g., par(xpd=NA)) and adjust margins; otherwise legends can overlap labels or be cut off.

## 7 Discussion

This paper developed a code-first atlas of oncology data visualizations implemented in R, with each figure framed as a specification linking *data requirements* to *assumptions*, *computation*, and *interpretation*. A core design choice throughout the atlas is the systematic use of simulation: by generating stylized datasets under controlled mechanisms, we aim to isolate what each plot conveys, when it is reliable, and how it can be adapted to the reader’s own oncology setting [7–9]. In this Discussion, we (i) compare plot families in terms of strengths, limitations, and appropriate scope, (ii) illustrate how multiple plots can be combined into coherent real-world analytic narratives, and (iii) highlight open challenges and future directions for oncology visualization practice.

### 7.1 Comparative Analysis

A practical message of the atlas is that oncology visualizations should be selected *backwards* from the scientific claim one wants to support, rather than *forwards* from what is easy to plot. In particular, the same dataset can support multiple competing visual narratives (e.g., treatment benefit summarized by a single hazard ratio versus a time-varying effect depiction), and the burden is on the analyst to choose the visualization that matches the estimand, assumptions, and intended audience [1, 2, 10].

**Table 1:** Comparative strengths and limitations of major plot families in the atlas. The “recommended use” column emphasizes when a plot is most defensible and informative, given typical oncology data structures.

| Plot family | Key strengths | Limitations / common pitfalls (and recommended use) |
| --- | --- | --- |
| Design & operating-characteristic plots (Phase I/II/III) | Directly communicate algorithm behavior (e.g., escalation aggressiveness, selection accuracy) and facilitate protocol justification under scenario libraries [1, 2]. | Can be misleading if scenarios do not reflect plausible clinical truth; performance metrics depend on tuning parameters and stopping rules. Recommended for design selection and team communication, not as post-hoc proof of superiority. |
| Patient-level response plots (waterfall, spider, swimmer) | Make heterogeneity visible; distinguish durable benefit from transient change; support clinical storytelling with explicit within-patient trajectories. | Overplotting and ordering choices can alter perception; censoring and assessment schedules may be hidden. Recommended as complements to tabular RECIST summaries and model-based estimates, not replacements. |
| Time-to-event summaries (KM, milestone, hazard curves, RMST-style displays) | Provide intuitive time-indexed treatment comparisons with uncertainty bands; milestone views can be clinically interpretable when medians are insufficient. | A single constant HR can be hard to interpret under non-proportional hazards; diagnostics and alternative summaries should be considered [10]. Recommended to pair (i) curves + risk tables with (ii) at least one diagnostic/robust summary when PH is doubtful. |
| Model diagnostics (log-log survival, Schoenfeld/martingale-style plots; calibration) | Convert assumptions into visible checks; support defensible model reporting and transparent refinement. | Diagnostics are often underused or overinterpreted; smoothing can create false patterns. Recommended for internal QC and reviewer-facing justification (especially for Cox PH and risk models) [11]. |
| RWE design diagnostics (Love plots, PS overlap/common support, flow diagrams) | Document whether an observational comparison is credible; make positivity and balance (pre/post adjustment) auditable; clarify attrition/selection pressure. | Can be gamed by selective covariate inclusion; good balance does not eliminate unmeasured confounding. Recommended as mandatory companions to comparative effectiveness figures in RWE, not optional add-ons. |
| Treatment pathways and transitions (Sankey/alluvial, chord, Gantt-style timelines) | Summarize real-world sequencing and attrition across lines of therapy; quickly surface dominant and rare pathways; support cohort-definition transparency. | Requires careful state definitions (one state per time/line) and consistent denominators; long tails can clutter. Recommended for RWE treatment-pattern characterization prior to outcome modeling. |
| High-dimensional omics visuals (oncoprint, volcano, lollipop/hotspot maps) | Compress high-dimensional molecular evidence into interpretable patterns; facilitate subtype annotation and hypothesis generation. | Multiple testing, annotation density, and cohort comparability can be undercommunicated; risk of narrative overreach. Recommended to couple with clear thresholds, external validation plans, and succinct annotation conventions. |
| Decision/HEOR visuals (CE plane, CEAC, tornado) | Represent joint uncertainty and decision thresholds; communicate sensitivity to key drivers and payer-specific willingness-to-pay. | Interpretation depends on model structure and uncertainty propagation; one-way sensitivity omits interactions. Recommended for decision support in HEOR with explicit statement of assumptions and threshold rationale. |
| Evolution narratives (fish-plot) | Encodes clonal remodeling (extinction, selective sweeps, branching) in a publication-ready geometry; effective for resistance/relapse stories [111]. | Risk of over-interpreting smooth boundaries; normalization and phylogeny uncertainty must be documented. Recommended as a communication layer downstream of validated clonal calling and phylogeny inference. |

A second comparative axis is *code-first packages versus interactive applications*. Interactive Shiny dashboards can be valuable for exploration and stakeholder engagement, but they are not inherently reproducible unless they are paired with versioned code, deterministic data pipelines, and exportable figure specifications. For a journal-facing atlas, we therefore prioritize scriptable workflows and treat apps as supportive interfaces rather than primary deliverables.

**Table 2:** Code-first versus app-first workflows for oncology visualization.

| Workflow | Advantages | Trade-offs / recommended use |
| --- | --- | --- |
| Code-first (R packages) | Deterministic regeneration; easy auditing under peer review; integrates naturally with simulation studies and literate programming; supports automated testing of figures [7, 9]. | Higher initial learning curve; requires disciplined coding standards. Recommended for manuscripts, regulatory-facing deliverables, and reusable templates. |
| App-first (Shiny dashboards) | Rapid exploratory analysis; accessible to non-programmers; supports interactive subgroup drilling and presentation-ready exploration. | Risk of hidden assumptions and “click-path” irreproducibility without logged parameters; export defaults can be inconsistent across sessions. Recommended for exploration and communication when paired with code snapshots and fixed export pipelines. |

### 7.2 Applications

We outline four representative case-study patterns to illustrate how multiple plots from the atlas can be assembled into an analysis narrative. These case studies are intended as transferable templates rather than claims about any single real dataset.

#### Case Study 1: Early-phase dose-finding design selection and communication

A typical Phase I workflow begins with operating-characteristic plots comparing candidate escalation rules across plausible toxicity scenarios, followed by a trial-history visualization to communicate how patient-level DLT outcomes drive escalation/de-escalation decisions [1, 2]. When efficacy is also of interest (Phase I/II settings), dose–efficacy curves (stylized or model-based) help align the dose recommendation with the intended clinical objective (e.g., MTD vs OBD). In practice, this bundle of plots functions as a protocol-facing explanation: (i) what the algorithm tends to do, (ii) where it is conservative/aggressive, and (iii) what safety guardrails are implied.

#### Case Study 2: RWE comparative effectiveness with transparent design diagnostics

A defensible RWE analysis typically begins with cohort derivation (participant-flow visualization) and proceeds to explicit balance and overlap diagnostics (Love plot and propensity-score density overlap) before displaying outcome contrasts (e.g., weighted survival curves, subgroup summaries). The diagnostic layer documents whether the observational comparison plausibly targets the intended contrast and whether positivity is sufficiently supported. This workflow also clarifies how much the analysis depends on modeling choices (propensity-score specification, trimming, matching ratio), which can then be summarized transparently in sensitivity plots [9].

#### Case Study 3: Therapy resistance and relapse narratives via tumor evolution visualization

When longitudinal sampling supports clonal reconstruction, fishplots provide an interpretable depiction of clonal remodeling over time (extinction, selective sweep, and branching) [111]. Importantly, the fishplot should be treated as the *communication layer*: analysts must explicitly document normalization conventions, uncertainty in phylogeny, and detection-limit considerations. In other words, the plot is most persuasive when paired with a clear statement of what was estimated upstream and what was assumed (e.g., fractions summing to 1 at each time point; lineage treated as given).

#### Case Study 4: Decision support in oncology HEOR

For health economic evaluations, cost-effectiveness planes summarize joint uncertainty in incremental cost and incremental effectiveness, while CEACs quantify the probability of cost-effectiveness across willingness-to-pay thresholds. Tornado diagrams then provide a deterministic complement, highlighting the most influential input parameters. In combination, these plots support both technical validation (are results stable to plausible parameter ranges?) and stakeholder interpretation (does the preferred strategy change across plausible thresholds?) [9].

### 7.3 Open Challenges & Future Directions

Despite a mature ecosystem of oncology visualizations, several methodological and practical challenges remain.

1. **Estimand alignment and the risk of visually convenient summaries.** A central challenge is ensuring that the visualization matches the estimand and does not implicitly smuggle in assumptions. The classic example is time-to-event analysis: when hazards are not proportional, a single constant HR can become both unstable and difficult to interpret; visual alternatives and diagnostics should be elevated from optional to standard practice [10].
2. **Reproducibility at scale (beyond “one-off” scripts).** Atlas-style figure generation benefits from modular data-generating mechanisms, standardized plotting APIs, pinned package versions, and automated regression tests for visual outputs. Simulation studies are particularly sensitive to silent code drift, which motivates stronger engineering practices in statistical figure pipelines [7, 9].
3. **From exploratory dashboards to publication-ready evidence.** Interactive apps lower the barrier to exploration, but they also increase the risk of untracked analytic decisions. A practical direction is to integrate dashboard interactions with code export (parameter logging, session metadata, and deterministic figure regeneration), so that exploratory workflows can transition cleanly into manuscript-grade reporting.
4. **RWE credibility: communicating bias mechanisms, not only outcomes.** In observational oncology, visualization should not end at outcome curves. Attrition, missingness, overlap, and balance are part of the scientific claim. A future direction is to further standardize “design-first” figure bundles that accompany every RWE comparative analysis (derivation + overlap + balance + sensitivity), thereby making bias mechanisms visible to readers.
5. **High-dimensional molecular visualization with controlled narrative risk.** Oncoprints, volcano plots, and hotspot maps are powerful but can amplify selective reporting. Future work should emphasize principled thresholding, multiplicity-aware summaries, and validation-oriented visuals that distinguish discovery from confirmation.
6. **Prediction and clinical utility: calibrating what “good” means.** Predictive models are increasingly reported with ROC/PR metrics, calibration curves, and decision-curve analysis. However, these visuals are often misinterpreted unless the target population, prevalence, and decision thresholds are clearly stated. Broader adoption of calibration-focused reporting [11] and disciplined decision-curve interpretation [12, 165] remains an important direction.
7. **Accessibility and robustness under publication constraints.** Color accessibility, grayscale legibility, annotation density, and export to both vector and high-resolution raster formats are recurring practical constraints. These constraints are not cosmetic: they directly affect interpretability, error rates, and the ability of reviewers to verify claims.
8. **Required Packages Maintenance.** A practical but recurrent challenge in atlas-style visualization is the long-term maintenance of the package dependencies required to reproduce each figure. In R, packages evolve continuously: versions are incremented, function signatures may change, defaults and graphical themes may be modified, and in some cases packages are archived or removed (e.g., due to deprecation, licensing, or trademark issues). Such dependency drift can interrupt previously stable plotting pipelines, particularly when figures rely on non-exported functionality, implicit defaults, or transitive dependencies that change outside the analyst’s control. To mitigate these risks, future atlas implementations should adopt explicit dependency management: (i) record package versions and session information for every figure, (ii) pin environments using lockfiles or project-level reproducibility tools, (iii) modularize code so that data simulation, data processing, and plotting layers can be updated independently, and (iv) implement lightweight regression checks (including visual comparisons) to detect unintended figure changes after package updates. These practices reduce the probability that routine package maintenance will compromise the reproducibility and visual fidelity of the atlas over time [7, 9].

In summary, the atlas is intended to function as a practical reference for epidemiologists, health economists, statisticians and programmers working in oncology. By framing each plot as a specification (data, assumptions, computation, interpretation) and by anchoring the exposition in reproducible R-based simulation workflows, we aim to support transparent visualization practice across clinical trials, real-world evidence, and hybrid analytic settings [7–9].

## Conclusions

Oncology research increasingly depends on high-dimensional data sources, complex endpoints, and heterogeneous study designs. In this setting, visualization is not merely a reporting accessory; rather, it is a technical interface that connects clinical questions to statistical evidence in a form that can be inspected, critiqued, and communicated. In this paper, we assembled a code-driven atlas of 62 oncology visualizations in R, spanning clinical trials (24 plots), real-world evidence (12 plots), and plot families that are routinely utilized in both settings (26 plots).

By establishing a common technical language—integrating statistical notation, software implementation, and clinical context—this atlas contributes a consolidated set of figure templates covering a broad range of oncology objectives. These range from early-phase dose-finding and operating-characteristic displays to survival modeling, treatment-pathway summaries, and decision-analytic graphics. Our emphasis throughout is pragmatic: each visualization is paired with a **minimum sufficient data schema**, a clear set of assumptions, and a reproducible implementation. This approach ensures that the figures can be adapted to alternate endpoints, censoring patterns, and cohort structures without redesigning the visualization from first principles.

Several directions follow naturally from this work. First, future versions can expand the atlas by incorporating additional plot families, such as multi-omic integration or health-equity metrics, and by strengthening cross-links between plot choice and the underlying estimand. Second, the long-term utility of the atlas depends on robust reproducibility infrastructure, including version-pinned environments, to ensure figures remain regenerable as the R ecosystem evolves. Finally, we view this atlas as a living technical resource: its greatest value will be realized when it is stress-tested and extended by the oncology biostatistics and data-science community in both applied trial and real-world settings.

## Abbreviations

The following abbreviations are used in this manuscript:

1L: First line (of therapy); 2L: Second line (of therapy); 3+3: Rule-based phase I dose-escalation design (3+3 design); 3L: Third line (of therapy); ABC-06: Advanced Biliary Tract Cancer trial 06; ABCA13: ATP-binding cassette subfamily A member 13; ADC: Antibody–drug conjugate; AE: Adverse event(s); ALND: Axillary lymph node dissection; ASC: Active symptom control; ASMD: Absolute standardized mean difference; AUC: Area under the curve; AUPRC: Area under the precision–recall curve; BLRM: Bayesian logistic regression model; BOIN: Bayesian optimal interval (design); BOR: Best overall response; BR: Bendamustine–rituximab; BTK: Bruton tyrosine kinase; BTKi: Bruton tyrosine kinase inhibitor; CAR-T: Chimeric antigen receptor T-cell (therapy); CART: Classification and regression tree(s); CCC19: COVID-19 and Cancer Consortium; CDF: Cumulative distribution function; CDH1: Cadherin 1; CEAC: Cost-effectiveness acceptability curve; CEP: Cost-effectiveness plane; CI: Confidence interval; CIF: Cumulative incidence function; CONSORT: Consolidated Standards of Reporting Trials; CPR: Complete pathological response; CR: Complete response / complete remission; CRM-L: Continual reassessment method (logistic model variant); DCA: Decision curve analysis; DCR: Disease control rate; DEGs: Differentially expressed genes; DLT: Dose-limiting toxicity; DMC: Data monitoring committee; DSMB: Data and Safety Monitoring Board; ECOG: Eastern Cooperative Oncology Group (performance status); EDp: Effective dose achieving p% response; Emax: Maximum effect (Emax) dose–response model; EORTC: European Organisation for Research and Treatment of Cancer; ER: Estrogen receptor; FCR: Fludarabine–cyclophosphamide–rituximab; FOLFIRINOX: Folinic acid (leucovorin) + fluorouracil + irinotecan + oxaliplatin; FOLFOX: Folinic acid (leucovorin) + fluorouracil + oxaliplatin; FPR: False positive rate; GATA3: GATA binding protein 3; HCT: Hematopoietic cell transplantation; HEOR: Health economics and outcomes research; HER2: Human epidermal growth factor receptor 2; HR: Hazard ratio; HRQoL: Health-related quality of life; IBR: Ibrutinib; ICER: Incremental cost-effectiveness ratio; IDER: Idelalisib–dexamethasone–rituximab; IFN-*γ*: Interferon gamma; IHC: Immunohistochemistry; IO: Immuno-oncology / immunotherapy; IPI: Ipilimumab; IPTW: Inverse probability of treatment weighting; ITD: Internal tandem duplication; ITD-Low: Low internal tandem duplication subgroup; ITT: Intention-to-treat; KM: Kaplan–Meier; KMT2C: Lysine methyltransferase 2C; LOESS: Locally estimated scatterplot smoothing; LOT: Line(s) of therapy; MAP3K1: Mitogen-activated protein kinase kinase kinase 1; MCP-Mod: Multiple comparison procedures and modeling; MMRM: Mixed model for repeated measures; MRD: Measurable (minimal) residual disease; mTPI: Modified toxicity probability interval (design); MTD: Maximum tolerated dose; NB: Net benefit; NE: Not evaluable; NI: Noninferiority; NIVO: Nivolumab; NMB: Net monetary benefit; NPI: Nottingham prognostic index; NPV: Negative predictive value; NRG-BR008: NRG-BR008 Breast Cancer trial 008 (HERO); NRM: Non-relapse mortality; NSCLC: Non-small-cell lung cancer; OBD: Optimal biological dose; OCI: Obinutuzumab–chlorambucil–ibrutinib; ORR: Objective response rate; OS: Overall survival; PD: Progressive disease / progression (e.g., PD state); PD-1: Programmed cell death protein 1; PD-L1: Programmed death-ligand 1; PFS: Progression-free survival; PI: Prognostic index; PIK3CA: Phosphatidylinositol-4,5-bisphosphate 3-kinase catalytic subunit alpha; PK: Pharmacokinetics; PP: Predictive probability; PPV: Positive predictive value; PR: Partial response; progesterone receptor (context-dependent); PRO: Patient-reported outcome; PRO-CTCAE: Patient-Reported Outcomes version of the Common Terminology Criteria for Adverse Events; PS: Propensity score; PSA: Probabilistic sensitivity analysis; PSM: Propensity score matching; PTEN: Phosphatase and tensin homolog; QALY: Quality-adjusted life year; QoL: Quality of life; qPCR: Quantitative polymerase chain reaction; RCI: Rituximab–chlorambucil–idelalisib; RCS: Restricted cubic spline; RCT: Randomized controlled trial; RECIST: Response Evaluation Criteria in Solid Tumors; RFS: Recurrence-free survival; RILA: Radiation-induced lymphocyte apoptosis; RMST: Restricted mean survival time; ROC: Receiver operating characteristic (curve); RS: Recurrence score; RT: Radiation therapy / radiotherapy; RWE: Real-world evidence; SAP: Statistical analysis plan; SBR: Scarff–Bloom–Richardson (tumor grade); SD: Stable disease; SE: Standard error; SLD: Sum of lesion diameters; SLNB: Sentinel lymph node biopsy; SMD: Standardized mean difference; SPTA1: Spectrin alpha, erythrocytic 1; STROBE: Strengthening the Reporting of Observational Studies in Epidemiology; TD: Time-dependent / time-dependent Cox; TKI: Tyrosine kinase inhibitor; TP53: Tumor protein p53; TPR: True positive rate; TSS: Transcription start site; TTE: Time-to-event; TtD: Time to deterioration; TVC: Time-varying covariate; VEN: Venetoclax; VENR: Venetoclax–rituximab; ZFHX4: Zinc finger homeobox 4.

## Supplementary

The complete R scripts for reproducing the simulated datasets and figures in this paper can be downloaded freely at:

[a] Archive(ResearchGate): https://www.researchgate.net/profile/Mohsen-Soltanifar;
[b] Repository(GitHub): https://imstatsbee.github.io/vizOnc/.

## Author Contributions

Conceptualization, M.S.; methodology, M.S.; software, M.S. and C.H.L.; validation, M.S., A.J.P., Y.J., J.G. and C.H.L.; formal analysis, M.S. and C.H.L; investigation, M.S., A.J.P., Y.J., J.G. and C.H.L; resources, M.S.; data curation, M.S.; writing—original draft, M.S., A.J.P., Y.J., J.G. and C.H.L; writing—review and editing, M.S., A.J.P., Y.J., J.G. and C.H.L.; visualization, M.S., A.J.P., Y.J., J.G. and C.H.L.; supervision, M.S. and C.H.L.; project administration, M.S. and C.H.L.; funding acquisition, M.S., A.J.P., Y.J., J.G. and C.H.L. All authors have read and agreed to the published version of the manuscript.

## Data Availability Statement

All datasets in this paper were simulated and their codes are available in supplementary materials.

## Ethics Statement

This study used exclusively simulated data generated for the purpose of demonstrating oncology data visualization techniques. No real human participant data were used; therefore, institutional review board (IRB) approval and informed consent were not required.

## Supporting information

https://www.researchgate.net/profile/Mohsen-Soltanifar

## Acknowledgments

This work is dedicated to the millions of families across the world who have faced the cancer diagnosis of a beloved family member, and particularly to those who have lost someone dear to the disease. Their courage and journeys are the silent drivers of this collaborative effort. It is our hope that by refining how we visualize and interpret oncology data, we may contribute to a future of clearer insights, more effective treatments, and ultimately, a world where fewer families are touched by these losses.

