## Supplementary material for "A North American Collaborative Atlas of Oncology Data Visualization with R Statistical Software": https://www.researchgate.net/profile/Mohsen-Soltanifar

#### R Codes for Oncology Data Visualization

**Citation:** Soltanifar, M., Portuguese, A.J., Jeon, Y., Gauthier, J., & Lee, C.H. (2026). *A North American Collaborative Atlas of Oncology Data Visualization with R Statistical Software.* medRxiv preprint.
https://doi.org/xx.xxxxx/xxxx.xx.xx.xxxxxxxx
**Contact:** mohsen[dot]soltanifar[at]alumni[dot]utoronto[dot]ca
**Version:** 1.0.0.
**Archive(ResearchGate):** https://www.researchgate.net/profile/Mohsen-Soltanifar
**Repository(GitHub):** https://imstatsbee.github.io/vizOnc/
**Licence (Paper):** CC BY 4.0.
**Licence (Code):** MIT.
**Date:** 2026-03-20

**Scope of the supplementary material.**
This supplementary document provides the R code used to reproduce the 62 data visualizations presented in the paper. The collection is organized to reflect the major analytic settings covered in the manuscript, namely 24 plots for clinical trials (CT), 12 plots for real-world evidence (RWE), and 26 plots that are common to both CT and RWE applications. This structure is intended to help readers navigate the code base efficiently and to locate examples that correspond directly to the conceptual taxonomy developed in the main article.

**Purpose and practical use.**
The aim of this supplementary file is to offer a transparent, reproducible, and practically useful coding companion to the paper. Beyond documenting the graphical outputs themselves, the material is designed to support statisticians, data scientists, clinical researchers, and applied programmers who wish to understand, adapt, or extend these visualization templates in their own oncology workflows. In this sense, the supplement serves not only as technical documentation, but also as a working reference for implementation in applied research settings.

**Reproducibility and computing environment.**
All code in this supplement is intended to be reproduced in R statistical software under R version 4.5.2 (2025-10-31 ucrt), with platform specification x86_64-w64-mingw32/x64. Recording the computing environment in this way is important for computational reproducibility, package compatibility, and long-term reuse of the scripts. Readers attempting to rerun or adapt the code are encouraged to use the same or a closely compatible R installation in order to minimize version-related discrepancies.

**Reader guidance and interpretation.**
Taken together, these materials are meant to function as a concise technical atlas of oncology data-visualization code in R. Each script is presented as a reproducible example that readers may run directly, inspect for methodological structure, and modify for their own data sources or scientific questions. We hope that this supplement makes the paper more accessible in practice by bridging the gap between conceptual discussion and executable implementation, thereby supporting both methodological learning and publication-quality figure generation.

### Section A. Clinical Trials(CT)

**Figure 5.1.1.**

```
#[1] Packages
library(ggplot2)
library(dplyr)

#[2] Escalation probability functions (level-wise)
p_escalate_3plus3 <- function(p) (1 - p)^3 + 3 * p * (1 - p)^5 # 0/3 OR 1/3 then 0/3
p_escalate_best5  <- function(p) (1 - p)^5 + 5 * p * (1 - p)^4 # =< 1/5 DLT among 5

# c > 1 stretches the gap; try c = 1.4, 1.5, etc.
p_escalate_3plus3b <- function(p, c = 5) {
  f1  <- p_escalate_3plus3(p)
  f2  <- p_escalate_best5(p)
  ((1 +  c) * f1 + (1 -  c) * f2) / 2
}

p_escalate_best5b <- function(p, c = 12) {
  f1  <- p_escalate_3plus3(p)
  f2  <- p_escalate_best5(p)
  ((1 - c) * f1 + (1 + c) * f2) / 2
}

#[3] Grid of true DLT probabilities
grid <- tibble::tibble(
  p = seq(0, 1, length.out = 501)
) |>
  mutate(
    `3 + 3`      = p_escalate_3plus3b(p),
    `Best-of-5`  = p_escalate_best5b(p)
  ) |>
  tidyr::pivot_longer(cols = c(`3 + 3`, `Best-of-5`),
                      names_to = "Design", values_to = "Pr_escalate")

#[4] Plot
# [4-1]. Ensure levels are clean
grid$Design <- factor(grid$Design, levels = c("3 + 3", "Best-of-5"))

# [4-2]. Define a subset for points (every 25th point for clarity)
grid_points <- grid[seq(1, nrow(grid), by = 25), ]

Figure_A01 <- ggplot(grid, aes(x = p, y = Pr_escalate, color = Design,
                               linetype = Design, shape = Design)) +
  # Draw the smooth lines for all data
  geom_line(linewidth = 1) +
  # Add sparse points using the subsetted data
  # We specify the aesthetic here to ensure it maps to the global Design factor
  geom_point(data = grid_points, size = 3) +
  # Sync all scales: Names must be IDENTICAL to merge into one legend
  scale_shape_manual(name = "Design",
                     values = c("3 + 3" = 17, "Best-of-5" = 16)) +
```

```r
  scale_color_manual(name = "Design",
                     values = c("3 + 3" = "#D55E00", "Best-of-5" = "#0072B2")) +
  scale_linetype_manual(name = "Design",
                        values = c("3 + 3" = "solid", "Best-of-5" = "dashed")) +
  labs(
    x = "Probability of DLT (p)",
    y = "Probability of Escalation",
    title = "Escalation Probability by Design"
  ) +
  coord_cartesian(xlim = c(0, 1), ylim = c(0, 1)) +
  theme_minimal(base_size = 14) +
  theme(
    legend.position = "right",
    panel.grid.minor = element_blank(),
    legend.key.width = unit(1.5, "cm") # Makes the dashed line easier to see in legend
  )

Figure_A01

#[5] Save (Optional)
ggsave("Figure_A01.png", Figure_A01, width = 8, height = 8, dpi = 300)
ggsave("Figure_A01.pdf", Figure_A01, width=8, height=8, device = grDevices::cairo_pdf)
```

**Citation:** Soltanifar, M., Portuguese, A.J., Jeon, Y., Gauthier, J., & Lee, C.H. (2026). A North American Collaborative Atlas of Oncology Data Visualization with R Statistical Software. medRxiv preprint. https://doi.org/xx.xxxxx/xxxx.xx.xx.xxxxxxxx, ©2026 Soltanifar, M. et al. (CC BY 4.0/MIT)

**Figure 5.1.2.**

```r
#[1] Packages
library(ggplot2)
library(dplyr)
library(tidyr)
library(forcats)

#[2] Grid of starting-dose toxicities
p_grid <- tibble(p_start = seq(0, 0.20, by = 0.0025))

#[3] Parametric approximations
approx_curves <- p_grid |>
  mutate(
    `Two-stage` = pmax(0.10, pmin(0.50, 0.185 + 0.18 * (p_start^1.3))),
    `CRM-L`     = pmax(0.10, pmin(0.50, 0.395 - 0.225 * p_start)),
    `Best-of-5` = pmax(0.10, pmin(0.50, 0.345 - 0.150 * p_start)),
    `3 + 3`     = pmax(0.10, pmin(0.50, 0.305 - 0.175 * p_start))
  ) |>
  pivot_longer(cols = -p_start, names_to = "Design", values_to = "mean_pMTD") |>
  mutate(
    Design = fct_relevel(Design, "Two-stage", "CRM-L", "Best-of-5", "3 + 3")
  )

#[4] Define Professional Manual Scales
lt <- c("Two-stage" = "longdash",
        "CRM-L"     = "dashed",
        "Best-of-5" = "dotdash",
        "3 + 3"     = "solid")

cols <- c("Two-stage" = "#0072B2",
          "CRM-L"     = "#D55E00",
          "Best-of-5" = "#009E73",
          "3 + 3"     = "#CC79A7")

shapes <- c("Two-stage" = 17, # Triangle
            "CRM-L"     = 18, # Diamond
            "Best-of-5" = 16, # Circle
            "3 + 3"     = 15) # Square

#[5] Plot
# Create sparse subset for points to keep lines "thin"
approx_points <- approx_curves %>%
  group_by(Design) %>%
  slice(seq(1, n(), by = 10)) %>%
  ungroup()

Figure_A02 <- ggplot(approx_curves, aes(x = p_start, y = mean_pMTD,
                                        linetype = Design,
                                        color = Design,
                                        shape = Design)) +
  geom_line(linewidth = 1.1) +
  geom_point(data = approx_points, size = 3) +
```

```r
  # ALL manual scales must share the SAME 'name' to merge the legends
  scale_linetype_manual(name = "Design", values = lt) +
  scale_color_manual(name = "Design", values = cols) +
  scale_shape_manual(name = "Design", values = shapes) +
  coord_cartesian(xlim = c(0, 0.20), ylim = c(0.10, 0.50)) +
  labs(
    x = "Probability of DLT at starting dose",
    y = expression("Mean of " * p(MTD)),
    title = "Performance Metrics by Design"
  ) +
  theme_minimal(base_size = 14) +
  theme(
    legend.position = "right",
    legend.key.width = unit(1.5, "cm"), # Essential for dash visibility
    panel.grid.minor = element_blank()
  )

Figure_A02

#[6] Save(Optional)
ggsave("Figure_A02.png", Figure_A02, width = 9, height = 6, dpi = 300)
ggsave("Figure_A02.pdf", Figure_A02, width=9, height=6, device = grDevices::cairo_pdf)
```

**Figure 5.1.3.**

```
#[1] Packages
library(ggplot2)
library(dplyr)
library(tidyr)
library(forcats)

# [2] Trial size and dose grid
n_pat <- 20
doses <- 1:7

# [3] Dose paths
g1_path <- tibble(
  patient    = 1:n_pat,
  group      = "Group1",
  dose_level = c(1, 1, 2, 3, 3, 4, 5, 6, 6, 6, 5, 5, 6, 6, 6, 6, 6, 6, 6, 6)
)

g2_path <- tibble(
  patient    = 1:n_pat,
  group      = "Group2",
  dose_level = c(1, 2, 3, 3, 4, 4, 3, 3, 3, 4, 4, 3, 3, 3, 3, 3, 3, 3, 3, 3)
)

dose_hist <- bind_rows(g1_path, g2_path) |>
  mutate(group = fct_relevel(group, "Group1", "Group2"))

# [4] Toxicity events
tox <- tribble(
  ~patient, ~group,   ~tox_dose,
  6,        "Group1", 4,
  10,       "Group1", 6,
  5,        "Group2", 4,
  16,       "Group2", 3
)

# ---- Define Professional Colors ----
# Blue for Group 1, Orange for Group 2, Red for Toxicity
plot_cols <- c(
  "Group1"   = "#0072B2",
  "Group2"   = "#D55E00",
  "Toxicity" = "#CC0000"
)

#[5] Plot

Figure_A03 <- ggplot() +
  # ---- MAIN PLOT LAYERS ----
geom_step(
  data = dose_hist,
  aes(x = patient, y = dose_level, linetype = group, color = group),
  linewidth = 1.1,
```

```
  show.legend = FALSE
) +
  geom_point(
    data = tox,
    aes(x = patient, y = tox_dose),
    shape = 23, size = 3.5, stroke = 1.2, fill = "white", color = plot_cols["Toxicity"],
    show.legend = FALSE
  ) +

  # ---- LEGEND-ONLY LAYERS (keeping your structure) ----
geom_segment(
  data = tibble(
    legend_key = c("Group1", "Group2"),
    lty = c("solid", "dotdash")
  ),
  aes(x = 0, xend = 1, y = 0, yend = 0,
      colour = legend_key, linetype = legend_key),
  linewidth = 1,
  inherit.aes = FALSE
) +
  geom_point(
    data = tibble(legend_key = "Toxicity"),
    aes(x = 0.5, y = 0, colour = legend_key),
    shape = 23, size = 3, stroke = 1, fill = "white",
    inherit.aes = FALSE
  ) +

  # ---- SCALES ----
scale_linetype_manual(
  values = c("Group1" = "solid", "Group2" = "dotdash"),
  guide = "none"
) +
  scale_colour_manual(
    values = plot_cols,
    breaks = c("Group1", "Group2", "Toxicity"),
    name = "Trial annotations"
  ) +
  guides(
    colour = guide_legend(
      override.aes = list(
        linetype = c("solid", "dotdash", "blank"),
        shape    = c(NA, NA, 23),
        fill     = c(NA, NA, "white"),
        linewidth = c(1.1, 1.1, 0)
      )
    )
  ) +

  coord_cartesian(xlim = c(1, n_pat), ylim = c(1, 7.2)) +
  scale_x_continuous(breaks = seq(1, n_pat, by = 1)) +
  scale_y_continuous(breaks = doses) +
  labs(
    title = "Trial history",
```

```r
    x = "Patient no",
    y = "Dose level"
  ) +
  theme_minimal(base_size = 13) +
  theme(
    legend.position   = "bottom",
    legend.background = element_rect(fill = "white", colour = "grey70"),
    legend.key        = element_rect(fill = "white", colour = NA),
    legend.key.width  = unit(1.2, "cm"),
    panel.grid.minor  = element_blank() # Cleans up the background
  )

Figure_A03

# [6]  Save(optional)
ggsave("Figure_A03.png", Figure_A03, width = 8, height = 4.8, dpi = 300)
ggsave("Figure_A03.pdf", Figure_A03, width=8, height=4.8, device = grDevices::cairo_pdf)
```

**Figure 5.1.4.**

```r
#[1] Packages
library(ggplot2)
library(dplyr)
library(tidyr)
library(forcats)

#[2] Dose grid & Functions
dose <- tibble(x = seq(0, 1, length.out = 501))

lin_fun  <- function(x) pmin(1, pmax(0, 0.12 + 0.66 * x))
emax_fun <- function(x) 0.80 * (x / (0.25 + x))
invU_fun <- function(x) {
  peak <- 0.70
  mu   <- 0.70
  sig  <- 0.15
  y    <- peak * exp(-((x - mu)^2) / (2 * sig^2))
  y    <- pmin(1, y + 0.1 + 0.25 * x)
  pmin(1, pmax(0, y))
}

curves <- dose |>
  mutate(
    `Linear`               = lin_fun(x),
    `Emax (saturating)`    = emax_fun(x),
    `Inverted-U`           = invU_fun(x)
  ) |>
  pivot_longer(-x, names_to = "Curve", values_to = "eff") |>
  mutate(Curve = fct_relevel(Curve, "Emax (saturating)", "Inverted-U", "Linear"))

#[3] Define Professional Manual Scales
lt <- c("Emax (saturating)" = "solid",
        "Inverted-U"        = "dotted",
        "Linear"            = "dotdash")

# Professional Colorblind-Friendly Palette (Okabe-Ito)
cols <- c("Emax (saturating)" = "#0072B2", # Professional Blue
          "Inverted-U"        = "#D55E00", # Professional Orange/Red
          "Linear"            = "#009E73") # Professional Green

#[4] Plot
Figure_A04 <- ggplot(curves, aes(x = x, y = eff, linetype = Curve, color = Curve)) +
  geom_line(linewidth = 1.1) +
  scale_linetype_manual(
    values = lt,
    name   = "Dose-Response Shape" # Fixed to match exactly
  ) +
  scale_color_manual(
    values = cols,
    name   = "Dose-Response Shape" # Fixed to match exactly
  ) +
  coord_cartesian(xlim = c(0, 1), ylim = c(0, 1)) +
```

```r
  labs(
    x = "Agent Dose",
    y = "Efficacy Probability",
    title = ""
  ) +
  theme_minimal(base_size = 12) +
  theme(
    legend.position  = "right",
    legend.direction = "vertical",
    legend.title     = element_text(size = 11, face = "bold"),
    legend.text      = element_text(size = 10),
    legend.key.width = unit(1.5, "cm"),
    panel.grid.minor = element_blank()
  )

Figure_A04

#[5] Save(Optional)
ggsave("Figure_A04.png", Figure_A04, width = 7, height = 4.8, dpi = 300)
ggsave("Figure_A04.pdf", Figure_A04, width=7, height=4.8, device = grDevices::cairo_pdf)
```

**Figure 5.1.5.**

```r
#[1]Packages
library(ggplot2)
library(dplyr)
library(tidyr)
library(scales)

# [2] Data
dat <- tribble(
  ~HR_margin, ~Power,        ~N,
  1.05,  "80% power", 15573,
  1.10,  "80% power",  4160,
  1.15,  "80% power",  1950, # Added
  1.20,  "80% power",  1150, # Added
  1.25,  "80% power",   770, # Added
  1.30,  "80% power",   549,

  1.05,  "90% power", 21249,
  1.10,  "90% power",  5569,
  1.15,  "90% power",  2610, # Added
  1.20,  "90% power",  1540, # Added
  1.25,  "90% power",  1030, # Added
  1.30,  "90% power",   735
)

# [3] Plot - keeping all your original styling
Figure_A05 <- ggplot(dat, aes(x = HR_margin, y = N, color = Power, group = Power)) +
  geom_line(linewidth = 1) +
  geom_point(size = 3) +
  # labels: 80% below, 90% above
  geom_text(
    data = filter(dat, Power == "80% power"),
    aes(label = comma(N)),
    vjust = 1.5, size = 3, show.legend = FALSE
  ) +
  geom_text(
    data = filter(dat, Power == "90% power"),
    aes(label = comma(N)),
    vjust = -1, size = 3, show.legend = FALSE
  ) +
  scale_x_continuous(
    breaks = seq(1.05, 1.30, by = 0.05),
    limits = c(1.04, 1.31) # Slightly widened for label clearance
  ) +
  scale_y_continuous(labels = comma, limits = c(0, 25000)) +
  labs(
    title = "Study Required Sample Size for NI Comparison \n (Exponential TTE)",
    subtitle = "Accrual = 4, Follow-up = 1 (ratios to median event time)",
    x = "Noninferiority Margin (Hazard Ratio)",
    y = "Sample Size (N)",
    color = "Power"
  ) +
```

```r
  theme_minimal(base_size = 12) +
  theme(
    legend.position = "bottom",
    panel.grid.minor = element_blank()
  )

Figure_A05

#[4] Save (Optional)
ggsave("Figure_A05.png", Figure_A05, width = 6, height = 5, dpi = 300)
ggsave("Figure_A05.pdf", Figure_A05, width=6, height=5, device = grDevices::cairo_pdf)
```

**Citation:** Soltanifar, M., Portuguese, A.J., Jeon, Y., Gauthier, J., & Lee, C.H. (2026). A North American Collaborative Atlas of Oncology Data Visualization with R Statistical Software. medRxiv preprint. https://doi.org/xx.xxxxx/xxxx.xx.xx.xxxxxxxx, ©2026 Soltanifar, M. et al. (CC BY 4.0/MIT)

**Figure 5.1.6.**

```r
# [1] Packages
library(survival)
library(dplyr)
library(tidyr)
library(ggplot2)
library(cowplot)
set.seed(31701)

# [2] Simulate two-arm OS with wide separation
n1 <- 260; n2 <- 220
t_admin <- 16

# Target probabilities at Year 16
p1_target <- 0.25 # Chemotherapy
p2_target <- 0.30 # Hormone Therapy

# Shape parameters to induce different curve shapes
shape_chemo   <- 0.35
shape_hormone <- 0.85

# Recalculate scales to hit targets at t = 16
scale_chemo   <- t_admin / (-log(p1_target))^(1 / shape_chemo)
scale_hormone <- t_admin / (-log(p2_target))^(1 / shape_hormone)

rweib <- function(n, shape, scale) {
  scale * (-log(runif(n)))^(1 / shape)
}

tt1 <- tibble(
  arm   = "Chemotherapy",
  t_evt = rweib(n1, shape = shape_chemo, scale = scale_chemo),
  cenz  = runif(n1, 0, t_admin + 2),
  time  = pmin(t_evt, cenz, t_admin),
  status= as.integer(t_evt <= pmin(cenz, t_admin))
)

tt2 <- tibble(
  arm   = "Hormone Therapy",
  t_evt = rweib(n2, shape = shape_hormone, scale = scale_hormone),
  cenz  = runif(n2, 0, t_admin + 2),
  time  = pmin(t_evt, cenz, t_admin),
  status= as.integer(t_evt <= pmin(cenz, t_admin))
)

dat <- bind_rows(tt1, tt2)

# [3] Fit KM
fit <- survfit(Surv(time, status) ~ arm, data = dat, conf.type = "log-log")

# [4] Log-rank test
lr_test <- survdiff(Surv(time, status) ~ arm, data = dat)
```

```r
p_value <- 1 - pchisq(lr_test$chisq, df = length(lr_test$n) - 1)
p_label <- if (p_value < 0.001) {
  "Log-rank test p < 0.001"
} else {
  paste0("Log-rank p = ", formatC(p_value, format = "f", digits = 3))
}

# [5] Tidy KM data
km_df <- do.call(rbind, lapply(seq_along(fit$strata), function(i) {
  idx <- if (length(fit$strata) == 1) TRUE else rep(FALSE, length(fit$time))
  if (length(fit$strata) > 1) {
    k <- cumsum(fit$strata)
    lo <- ifelse(i == 1, 1, k[i - 1] + 1)
    hi <- k[i]
    idx[lo:hi] <- TRUE
  }
  tibble(
    arm   = names(fit$strata)[i],
    time  = fit$time[idx],
    surv  = fit$surv[idx],
    lower = fit$lower[idx],
    upper = fit$upper[idx],
    n.cen = fit$n.censor[idx]
  )
})) |>
  mutate(arm = sub("^arm=", "", arm))

cen_pts <- km_df |>
  filter(n.cen > 0) |>
  mutate(y = surv)

# [6] Aesthetics
lt <- c("Chemotherapy" = "solid", "Hormone Therapy" = "solid")
col_map <- c("Chemotherapy" = "firebrick4", "Hormone Therapy" = "navyblue")
fill_map <- c("Chemotherapy" = "firebrick4", "Hormone Therapy" = "navyblue")

# [7] Main KM plot
gp6_pre <- ggplot(km_df, aes(x = time, y = surv, linetype = arm)) +
  geom_ribbon(aes(ymin = lower, ymax = upper, fill = arm), alpha = 0.20, colour = NA) +
  geom_step(aes(color = arm), linewidth = 1.05) +
  geom_point(data = cen_pts, aes(y = y, color = arm), shape = 3, size = 2) +
  annotate("text", x = 10.8, y = 0.94, label = p_label, hjust = 0, size = 4.2) +
  scale_x_continuous(
    limits = c(0, 16),
    breaks = seq(0, 16, 2),
    expand = c(0.01, 0)
  ) +
  scale_y_continuous(
    limits = c(0, 1),
    breaks = seq(0, 1, 0.25),
    labels = function(x) paste0(round(100 * x), "%"),
    expand = c(0, 0)
  ) +
```

```r
  scale_linetype_manual(values = lt, name = "Treatment") +
  scale_color_manual(values = col_map, name = "Treatment") +
  scale_fill_manual(values = fill_map, name = "Treatment") +
  labs(
    x = "Time (years)",
    y = "OS Probability (%)",
    title = "Kaplan-Meier Curves for Overall Survival"
  ) +
  theme_minimal(base_size = 12) +
  theme(
    legend.position = "bottom",
    legend.background = element_rect(fill = "white", colour = "black"),
    axis.title.x = element_text(margin = margin(t = 8)),
    axis.text.x = element_text(margin = margin(t = 2)),
    plot.title = element_text(face = "bold"),
    plot.margin = margin(5, 5, 2, 5)
  )

# [8] Risk table data
times <- seq(0, 16, by = 2)
risk_summary <- summary(fit, times = times)

risk_df <- tibble(
  time   = risk_summary$time+0.25,
  n.risk = risk_summary$n.risk,
  arm    = sub("^arm=", "", risk_summary$strata)
)

risk_df$arm <- factor(risk_df$arm, levels = c("Hormone Therapy", "Chemotherapy"))

# [9] Risk table plot
risk_table <- ggplot(risk_df, aes(x = time, y = arm, label = n.risk)) +
  geom_text(aes(color = arm), size = 4, fontface = "bold") +
  scale_x_continuous(
    limits = c(0, 17),
    breaks = seq(0, 16, 2),
    expand = c(0.01, 0)
  ) +
  scale_color_manual(values = col_map) +
  labs(y = NULL, x = NULL) +
  theme_minimal(base_size = 12) +
  theme(
    panel.grid.major = element_blank(),
    panel.grid.minor = element_blank(),
    axis.text.x = element_blank(),
    axis.title.x = element_blank(),
    axis.ticks.x = element_blank(),
    axis.text.y = element_text(face = "bold", color = "black"),
    legend.position = "none",
    plot.margin = margin(0, 5, 5, 5)
  )

# Optional "Number at risk" label above table
```

```r
risk_title <- ggplot() +
  annotate("text", x = 0, y = 0.5, label = "Number at risk", hjust = 0,
           fontface = "bold", size = 4.2) +
  theme_void() +
  xlim(0, 1) + ylim(0, 1)

# [10] Combine plots
Figure_A06 <- plot_grid(
  gp6_pre,
  risk_title,
  risk_table,
  ncol = 1,
  align = "v",
  axis = "lr",
  rel_heights = c(4.2, 0.35, 1)
)

Figure_A06

#[8]Save (Optional)
ggsave("Figure_A06.png", Figure_A06, width = 9, height = 6, dpi = 300)
ggsave("Figure_A06.pdf", Figure_A06, width=  9, height=  6, device = grDevices::cairo_pdf)
```

**Citation:** Soltanifar, M., Portuguese, A.J., Jeon, Y., Gauthier, J., & Lee, C.H. (2026). A North American Collaborative Atlas of Oncology Data Visualization with R Statistical Software. medRxiv preprint. https://doi.org/xx.xxxxx/xxxx.xx.xx.xxxxxxxx, ©2026 Soltanifar, M. et al. (CC BY 4.0/MIT)

**Figure 5.1.7.**

```r
# [1] Packages
library(survival)
library(dplyr)
library(tidyr)
library(ggplot2)
library(cowplot)
set.seed(31701)

# [2] Simulate two-arm PFS
n1 <- 260; n2 <- 240
t_admin <- 16

rweib <- function(n, shape, scale) scale * (-log(runif(n)))^(1/shape)

tt1 <- tibble(
  arm   = "Radiation",
  t_evt = pmin(rweib(n1, shape = 0.35, scale = 0.5), t_admin),
  cenz  = runif(n1, 0, t_admin),
  time  = pmin(t_evt, cenz),
  status= as.integer(t_evt <= cenz)
)

tt2 <- tibble(
  arm   = "Surgery",
  t_evt = pmin(rweib(n2, shape = 0.50, scale = 5.0), t_admin),
  cenz  = runif(n2, 0, t_admin),
  time  = pmin(t_evt, cenz),
  status= as.integer(t_evt <= cenz)
)

dat <- bind_rows(tt1, tt2)

# [3] Fit KM
fit <- survfit(Surv(time, status) ~ arm, data = dat, conf.type = "log-log")

# [3.1] Log-rank test
lr_test <- survdiff(Surv(time, status) ~ arm, data = dat)
p_value <- 1 - pchisq(lr_test$chisq, df = length(lr_test$n) - 1)
p_label <- if (p_value < 0.001) {
  "Log-rank test p < 0.001"
} else {
  paste0("Log-rank p = ", formatC(p_value, format = "f", digits = 3))
}

# [4] Tidy data
km_df <- do.call(rbind, lapply(seq_along(fit$strata), function(i) {
  idx <- if (length(fit$strata) == 1) TRUE else rep(FALSE, length(fit$time))
  if (length(fit$strata) > 1) {
    k <- cumsum(fit$strata)
    lo <- ifelse(i == 1, 1, k[i - 1] + 1)
    hi <- k[i]
```

```r
    idx[lo:hi] <- TRUE
  }
  tibble(
    arm   = names(fit$strata)[i],
    time  = fit$time[idx],
    surv  = fit$surv[idx],
    lower = fit$lower[idx],
    upper = fit$upper[idx],
    n.cen = fit$n.censor[idx]
  )
})) |>
  mutate(arm = sub("^arm=", "", arm))

cen_pts <- km_df |> filter(n.cen > 0) |> mutate(y = surv)

# [5] KM Plot
lt <- c("Radiation" = "solid", "Surgery" = "solid")
col_map <- c("Radiation" = "goldenrod3", "Surgery" = "steelblue3")

gp10_pre <- ggplot(km_df, aes(x = time, y = surv, linetype = arm)) +
  geom_ribbon(aes(ymin = lower, ymax = upper, fill = arm), alpha = 0.20, colour = NA) +
  geom_step(aes(color = arm), linewidth = 1) +
  geom_point(data = cen_pts, aes(y = y, color = arm), shape = 3, size = 2) +
  annotate("text", x = 4.8, y = 0.96, label = p_label, hjust = 0, size = 4) +
  scale_x_continuous(
    limits = c(0, 16),
    breaks = seq(0, 16, 2),
    expand = c(0.01, 0)
  ) +
  scale_y_continuous(
    limits = c(0, 1),
    breaks = seq(0, 1, 0.25),
    labels = function(x) paste0(round(100 * x), "%"),
    expand = c(0, 0)
  ) +
  # Update names to "Treatment" so the legends merge correctly
  scale_linetype_manual(values = lt, name = "Treatment") +
  scale_color_manual(values = col_map, name = "Treatment") +
  scale_fill_manual(values = c("Radiation" = "gold", "Surgery" = "steelblue"),
                    name = "Treatment") +
  labs(
    x = "Time (years)",
    y = "PFS probability (%)",
    title = "Milestone Survival plots"
  ) +
  theme_minimal(base_size = 12) +
  theme(
    legend.position = "bottom",
    legend.background = element_rect(fill = "white", colour = "black"),
    axis.title.x = element_text(margin = margin(t = 8)),
    axis.text.x = element_text(margin = margin(t = 2)),
    plot.margin = margin(5, 5, 0, 5)
  ) +
```

```r
  geom_vline(xintercept = 10, linetype = "dashed", color = "red") +
  annotate("text", x = 10.5, y = 0.75, label = "10-Year Milestone", angle = 90) +
  geom_vline(xintercept = 15, linetype = "dashed", color = "red") +
  annotate("text", x = 15.5, y = 0.75, label = "15-Year Milestone", angle = 90)

# [6] Risk Table Data
times <- seq(0, 16, by = 2)
risk_summary <- summary(fit, times = times)

risk_df <- tibble(
  time   = risk_summary$time+0.25,
  n.risk = risk_summary$n.risk,
  arm    = sub("^arm=", "", risk_summary$strata)
)

# [7] Risk Table Plot
risk_table <- ggplot(risk_df, aes(x = time, y = arm, label = n.risk)) +
  geom_text(aes(color = arm), size = 3.5, fontface = "bold") +
  scale_x_continuous(
    limits = c(0, 16),
    breaks = seq(0, 16, 2),
    expand = c(0.01, 0)
  ) +
  scale_color_manual(values = col_map) +
  labs(x = NULL, y = NULL) +
  theme_minimal() +
  theme(
    panel.grid.major = element_blank(),
    panel.grid.minor = element_blank(),
    axis.text.x = element_blank(),
    axis.title.x = element_blank(),
    axis.ticks.x = element_blank(),
    axis.text.y = element_text(face = "bold", color = "black"),
    legend.position = "none",
    plot.margin = margin(0, 5, 5, 5)
  )

# [8] This aligns the plots vertically and stacks them
Figure_A07 <- plot_grid(
  gp10_pre,
  risk_table,
  ncol = 1,
  align = "v",
  rel_heights = c(4, 1)
)

Figure_A07

#[9] Save (Optional)
ggsave("Figure_A07.png", Figure_A07, width = 9, height = 6, dpi = 300)
ggsave("Figure_A07.pdf", Figure_A07, width=9, height=6, device = grDevices::cairo_pdf)
```

**Figure 5.1.8.**

```r
#[1] Packages
library(dplyr)
library(ggplot2)
library(forcats)
library(scales)
set.seed(1227)

#[2] fabricate subject-level best % changes (in percent)
n <- 22
raw <- c(runif(6, 10, 40), runif(2, 0, 8), runif(4, -28, -10),
         runif(10, -55, -32))  # mixture to show all RECIST zones
df <- tibble(
  id        = paste0("P", sprintf("%02d", 1:n)),
  best_pct  = round(raw, 1)
) |>
  arrange(best_pct) |>
  mutate(
    recist_cat = case_when(
      best_pct >  20  ~ "Disease progression",
      best_pct <= -30 ~ "Partial response",
      TRUE            ~ "Stable disease"
    ),
    id = fct_inorder(id)  # order bars left->right by best_pct
  )

# [3] color map (clinically intuitive)
cols <- c(
  "Disease progression" = "#D62728",  # red
  "Stable disease"      = "#7F7F7F",  # gray
  "Partial response"    = "#2CA02C"   # green
)

# [4] plot
Figure_A08<- ggplot(df, aes(x = id, y = best_pct, fill = recist_cat)) +
  geom_col(color = "grey20", width = 0.85) +
  geom_hline(yintercept = -30, linetype = "dashed") +
  geom_hline(yintercept =  20, linetype = "dashed") +
  annotate("text", x = 20.0, y = -30, label = "-30%", vjust = -0.6, size = 3.4) +
  annotate("text", x = 2.0, y =  20, label = "+20%", vjust = -0.6, size = 3.4) +
  scale_fill_manual(values = cols, name = NULL) +
  scale_y_continuous(labels = function(z) paste0(z, "%"),
                     limits = c(-60, 45), breaks = seq(-60, 40, 10)) +
  labs(
    x = "Response-Evaluable Patients",
    y = "Percentage Change (from baseline)",
    title = "Waterfall plot of best tumor size change by subject"
  ) +
  theme_minimal(base_size = 12) +
  theme(
    legend.position = "right",
    axis.text.x = element_blank(),     # de-identify; bars encode count
```

```r
    panel.grid.major.x = element_blank()
  )

Figure_A08

#[5] Save(Optional):
ggsave("Figure_A08.png", Figure_A08, width = 7.5, height = 7.5, dpi = 300)
ggsave("Figure_A08.pdf", Figure_A08, width=7.5, height=7.5, device = grDevices::cairo_pdf)
```

**Citation:** Soltanifar, M., Portuguese, A.J., Jeon, Y., Gauthier, J., & Lee, C.H. (2026). A North American Collaborative Atlas of Oncology Data Visualization with R Statistical Software. medRxiv preprint. https://doi.org/xx.xxxxx/xxxx.xx.xx.xxxxxxxx, ©2026 Soltanifar, M. et al. (CC BY 4.0/MIT)

**Figure 5.1.9.**

```r
# [1] Packages
library(dplyr)
library(ggplot2)
library(forcats)

# [2] Expanded Data (15 Patients)
swim <- tibble::tibble(
  id    = paste0("P", sprintf("%02d", 1:15)),
  stage = factor(c("Stage 2","Stage 1","Stage 3","Stage 3","Stage 3","Stage 3",
                   "Stage 3","Stage 4", "Stage 2","Stage 3","Stage 1","Stage 4",
                   "Stage 3","Stage 2","Stage 3"),
                     levels = paste("Stage", 1:4)),
  tx_start   = 0,
  tx_end     = c(20, 18, 17, 16, 16, 14, 15, 13, 22, 19, 21, 15, 24, 18, 20),
  ongoing    = c(TRUE, TRUE, FALSE, TRUE, FALSE, TRUE, FALSE, FALSE, TRUE, FALSE,
               TRUE, FALSE, TRUE, FALSE, TRUE),
  resp_start = c(6, 9, NA, 12, 8, 7, 11, 9, 5, 10, 8, NA, 6, 9, 7),
  # Partial response
  resp_cont  = c(12, NA, 14, 15, NA, 10, NA, NA, 15, NA, 16, NA, 18, NA, 14),
  # Continued response
  resp_end   = c(NA, 18, NA, NA, 14, NA, NA, NA, NA, 17, NA, NA, NA, 16, NA),
  # Response end
  progression= c(NA, NA, NA, NA, NA, NA, 16, 12, NA, 20, NA, 17, NA, NA, NA)
  # Disease progression
) |>
  mutate(id = fct_rev(fct_inorder(id)))

events <- bind_rows(
  filter(swim, !is.na(resp_start))  |> transmute(id, time = resp_start,
                                        event = "Partial response"),
  filter(swim, !is.na(resp_cont))   |> transmute(id, time = resp_cont,
                                        event = "Continued response"),
  filter(swim, !is.na(resp_end))    |> transmute(id, time = resp_end,
                                        event = "Response end"),
  filter(swim, !is.na(progression)) |> transmute(id, time = progression,
                                        event = "Disease progression")
) |>
  mutate(
    id    = fct_relevel(id, levels(swim$id)),
    event = factor(event,
                   levels = c("Partial response","Continued response",
                              "Disease progression","Response end"))
  )
# [3] Aesthetics
cols_stage <- c("Stage 1"="#2CA02C","Stage 2"="#E69F00",
                "Stage 3"="#1F77B4","Stage 4"="#6A51A3")
shape_map <- c("Partial response"=25,"Continued response"=24,
               "Disease progression"=23,"Response end"=21)
color_map <- c("Partial response"="#2C7FB8","Continued response"="#1B9E77",
               "Disease progression"="#D62728","Response end"="#000000")
```

```r
# [4] Geometry helpers
bar_half_height <- 0.35
bars <- swim |>
  transmute(
    id, stage,
    xmin = tx_start, xmax = tx_end,
    ymin = as.numeric(id) - bar_half_height,
    ymax = as.numeric(id) + bar_half_height
  )
# [5] Plot
Figure_A09 <- ggplot() +
  geom_rect(data = bars,
            aes(xmin=xmin, xmax=xmax, ymin=ymin, ymax=ymax, fill=stage),
            color = "grey20") +
  geom_segment(
    data = swim |> mutate(y = as.numeric(id)),
    aes(x = tx_end - 0.6, xend = tx_end, y = y, yend = y, color = stage),
    linewidth = 1.2,
    arrow = arrow(length = unit(0.20, "cm"), type = "closed"),
    show.legend = FALSE
  ) +
  geom_point(
    data = events,
    aes(x = time, y = as.numeric(id), shape = event, color = event),
    size = 3.6, stroke = 1.6
  ) +
  scale_fill_manual(values = cols_stage, name = "Stage") +
  scale_shape_manual(values = shape_map, name = "Disease status") +
  scale_color_manual(values = color_map,  name = "Disease status") +
  scale_x_continuous(limits = c(0, 26), breaks = seq(0, 26, 2)) + # Expanded x-axis to 26
  scale_y_continuous(labels = levels(swim$id), breaks = seq_along(levels(swim$id))) +
  labs(
    x = "Time (months)",
    y = "Patients on Study Drug of Interest",
    title = "Swimmer Plot: treatment duration and response milestones"
  ) +
  theme_minimal(base_size = 12) +
  theme(
    panel.grid.major.y = element_blank(),
    panel.grid.minor   = element_blank(),
    legend.position    = "right"
  ) +
  guides(
    fill  = guide_legend(order = 1, override.aes = list(color = "grey20")),
    shape = guide_legend(order = 2),
    color = guide_legend(order = 2)
  )
Figure_A09
# [6]Save(Optional):
ggsave("Figure_A09.png", Figure_A09, width = 8, height = 8, dpi = 300)
ggsave("Figure_A09.pdf", Figure_A09, width=8, height=8, device = grDevices::cairo_pdf)
```

**Citation:** Soltanifar, M., Portuguese, A.J., Jeon, Y., Gauthier, J., & Lee, C.H. (2026). A North American Collaborative Atlas of Oncology Data Visualization with R Statistical Software. medRxiv preprint. https://doi.org/xx.xxxxx/xxxx.xx.xx.xxxxxxxx, ©2026 Soltanifar, M. et al. (CC BY 4.0/MIT)

**Figure 5.1.10.**

```r
# [1] Packages
library(dplyr)
library(ggplot2)
library(tidyr)
library(purrr)
set.seed(60217)

# [2] fabricate 100 studies then SAMPLE 25
n_total <- 100
n_sample <- 25

dat_full <- tibble(
  study_id = paste0("S", sprintf("%03d", 1:n_total)),
  beta     = rnorm(n_total, mean = -0.25, sd = 0.65),
  se       = 0.18 + 0.25 * runif(n_total)
)

dat <- dat_full %>%
  sample_n(n_sample) %>%
  arrange(beta) %>%
  mutate(
    lo = beta - 1.96 * se,
    hi = beta + 1.96 * se,
    # Logic: Red (Sig Positive), Green (Sig Negative), Grey (Non-sig)
    status = case_when(
      lo > 0 ~ "Increased risk (Sig)",
      hi < 0 ~ "Decreased risk (Sig)",
      TRUE   ~ "CI includes 0"
    ),
    # Set factor levels for a logical legend order
    status = factor(status, levels = c("Decreased risk (Sig)",
                                       "CI includes 0",
                                       "Increased risk (Sig)")),
    study_num = gsub("S", "", study_id),
    hr_text   = sprintf("%.2f", beta),
    ci_text   = sprintf("(%.2f, %.2f)", lo, hi),
    rank      = row_number()
  )

# [3] Professional Color Palette
prof_cols <- c(
  "Decreased risk (Sig)" = "#009E73", # Professional Green
  "CI includes 0"        = "#999999", # Professional Grey
  "Increased risk (Sig)" = "#D55E00"  # Professional Red/Vermillion
)

# [4] Plot Aesthetics
Figure_A10 <- ggplot(dat, aes(y = rank, x = beta, color = status, fill = status)) +

# --- Column Text (Color set to black to keep features intact) ---
geom_text(aes(x = -5.0, label = study_num), size = 3.5, hjust = 0, color = "black") +
```

```r
geom_text(aes(x = 4.0, label = hr_text), size = 3.5, hjust = 0.5, color = "black") +
geom_text(aes(x = 6.0, label = ci_text), size = 3.5, hjust = 0.5, color = "black") +

# --- Column Headers ---
annotate("text", x = -5.0, y = 0, label = "Study", fontface = "bold", size = 4, hjust = 0) +
annotate("text", x = 4.0,  y = 0, label = "log-HR", fontface = "bold", size = 4, hjust = 0.5) +
annotate("text", x = 6.0,  y = 0, label = "95% CI", fontface = "bold", size = 4, hjust = 0.5) +

# --- Forest Plot Elements ---
geom_errorbarh(aes(xmin = lo, xmax = hi), height = 0, size = 0.7, alpha = 0.8) +
geom_point(shape = 21, size = 3.2, stroke = 0.6, color = "black") +

# Reference line at 0
geom_vline(xintercept = 0, linewidth = 0.8, color = "black") +

# Scales and Styling
scale_fill_manual(values = prof_cols, name = "Clinical Significance") +
scale_color_manual(values = prof_cols, name = "Clinical Significance") +
scale_y_reverse(breaks = NULL) +
scale_x_continuous(breaks = seq(-3, 3, 0.5)) +

# Legend management
guides(fill = guide_legend(override.aes = list(size = 4))) +

coord_cartesian(xlim = c(-5.5, 7.5), ylim = c(n_sample + 1, -1)) +

labs(
    x = "Log-Hazard Ratio",
    y = NULL,
    title = paste("Forest Plot of", n_sample, "Randomly Sampled Studies")
  ) +
  theme_minimal(base_size = 12) +
  theme(legend.position = c(0.55, 0.85),
        legend.background = element_rect(fill = "white", colour = "grey80"),
        panel.grid.major.y = element_blank(),
        panel.grid.minor.y = element_blank(),
        plot.margin = margin(10, 20, 10, 10))

Figure_A10

# [5] Save(Optional)
ggsave("Figure_A10.png", Figure_A10, width = 10.5, height = 7, dpi = 600)
ggsave("Figure_A10.pdf", Figure_A10, width=10.5, height=7, device = grDevices::cairo_pdf)
```

**Citation:** Soltanifar, M., Portuguese, A.J., Jeon, Y., Gauthier, J., & Lee, C.H. (2026). A North American Collaborative Atlas of Oncology Data Visualization with R Statistical Software. medRxiv preprint. https://doi.org/xx.xxxxx/xxxx.xx.xx.xxxxxxxx, ©2026 Soltanifar, M. et al. (CC BY 4.0/MIT)

**Figure 5.1.11.**

```r
#[1] Packages
#install.packages('ggoncoplot',
#repos = c('https://selkamand.r-universe.dev', 'https://cloud.r-project.org'))
#if (!requireNamespace("BiocManager", quietly = TRUE)) install.packages("BiocManager")
#BiocManager::install("ComplexHeatmap")

library(ComplexHeatmap)
library(circlize)
library(dplyr)
library(tidyr)
library(tibble)
set.seed(20260116)

# [1] Simulate Dataset
genes   <- c("PIK3CA","GATA3","CDH1","TP53","KMT2C",
             "MAP3K1","SPTA1","ZFHX4","PTEN","ABCA13")
samples <- paste0("TCGA_", sprintf("%03d", 1:120))
all_alts  <- c("Missense","Nonsense","Frame_Shift_Del",
               "In_Frame_Ins","Splice_Site", "Silent")

dat <- expand.grid(sample_id = samples, gene = genes, KEEP.OUT.ATTRS = FALSE) %>%
  mutate(prob = runif(n()),
         altered = prob < rep(seq(0.35, 0.06, length.out = length(genes)),
                              each = length(samples))) %>%
  mutate(alteration = ifelse(altered, sample(all_alts, n(), replace = TRUE,
                    prob = c(.50,.12,.12,.08,.08,.10)), "")) %>%
  select(sample_id, gene, alteration)

# [2] Clinical annotations
clin <- tibble(
  sample_id = samples,
  ER  = sample(c("Positive","Negative","NE"), length(samples), replace = TRUE,
               prob = c(.65,.30,.05)),
  PR  = sample(c("Positive","Negative","NE"), length(samples), replace = TRUE,
               prob = c(.60,.35,.05)),
  HER2= sample(c("Positive","Negative","Indeterminate"), length(samples),
               replace = TRUE, prob = c(.18,.72,.10))
)

# [3] Create Mutation Matrix
mat <- dat %>%
  pivot_wider(names_from = sample_id, values_from = alteration, values_fill = "") %>%
  column_to_rownames("gene") %>%
  as.matrix()

# [4] Define Colors and Alteration Functions
col_alter <- c(
  "Missense"        = "#1f78b4",
  "Nonsense"        = "#e31a1c",
  "Frame_Shift_Del" = "#ff7f00",
  "In_Frame_Ins"    = "#6a3d9a",
```

```r
  "Splice_Site"     = "#33a02c",
  "Silent"          = "#bdbdbd"
)

alter_fun <- list(
  background = function(x, y, w, h) {
    grid::grid.rect(x, y, w, h, gp = grid::gpar(fill = "#f0f0f0", col = NA))
  },
  Missense        = function(x,y,w,h) grid::grid.rect(x,y,w*0.9,h*0.9,
                            gp=grid::gpar(fill=col_alter["Missense"], col=NA)),
  Nonsense        = function(x,y,w,h) grid::grid.rect(x,y,w*0.9,h*0.9,
                            gp=grid::gpar(fill=col_alter["Nonsense"], col=NA)),
  Frame_Shift_Del = function(x,y,w,h) grid::grid.rect(x,y,w*0.9,h*0.9,
                            gp=grid::gpar(fill=col_alter["Frame_Shift_Del"], col=NA)),
  In_Frame_Ins    = function(x,y,w,h) grid::grid.rect(x,y,w*0.9,h*0.9,
                            gp=grid::gpar(fill=col_alter["In_Frame_Ins"], col=NA)),
  Splice_Site     = function(x,y,w,h) grid::grid.rect(x,y,w*0.9,h*0.9,
                            gp=grid::gpar(fill=col_alter["Splice_Site"], col=NA)),
  Silent          = function(x,y,w,h) grid::grid.rect(x,y,w*0.9,h*0.9,
                            gp=grid::gpar(fill=col_alter["Silent"], col=NA))
)

# [5] Annotations
top_anno <- HeatmapAnnotation(
  `Mutations/Sample` = anno_barplot(colSums(mat != ""),
                    gp = grid::gpar(fill = "grey40", col = NA), border = FALSE),
  annotation_name_side = "right"
)

# Correction: Barplot and Percentages should only count functional (non-silent) mutations
mat_func <- mat
mat_func[mat_func == "Silent"] <- ""
gene_counts_func <- rowSums(mat_func != "")

right_anno <- rowAnnotation(
  `Samples Mutated` = anno_barplot(gene_counts_func,
                    gp = grid::gpar(fill = "grey40", col = NA), border = FALSE),
  annotation_name_rot = 0 # Fixed overlap
)

# Clinical Colors
clin_cols <- list(
  ER   = c(Negative = "#b2182b", Positive = "#2166ac", NE = "#ffffff"),
  PR   = c(Negative = "#b2182b", Positive = "#2166ac", NE = "#ffffff"),
  HER2 = c(Negative = "#525252", Positive = "#fec44f", Indeterminate = "#ffffff")
)

# FIX: Order columns by sample_id, THEN remove the ID column
bottom_anno <- HeatmapAnnotation(
  df = clin %>%
    slice(match(colnames(mat), sample_id)) %>%
    select(-sample_id),
  col = clin_cols,
```

```r
  annotation_name_side = "left"
)

# [6] Final Oncoprint
Figure_A11 <- oncoPrint(
  mat,
  alter_fun = alter_fun,
  col = col_alter,
  top_annotation = top_anno,
  right_annotation = right_anno,
  bottom_annotation = bottom_anno,
  row_names_side = "left",
  pct_side = "right",
  column_title = "Oncoprint",
  remove_empty_columns = TRUE
)

draw(Figure_A11, heatmap_legend_side = "right", annotation_legend_side = "right")

#[9] Save (Optional)
png("Figure_A11.png", width = 3000, height = 2000, res = 300)
draw(Figure_A11)
dev.off()

pdf("Figure_A11.pdf", width = 10, height = 6.67)
draw(Figure_A11)
dev.off()
```

**Citation:** Soltanifar, M., Portuguese, A.J., Jeon, Y., Gauthier, J., & Lee, C.H. (2026). A North American Collaborative Atlas of Oncology Data Visualization with R Statistical Software. medRxiv preprint. https://doi.org/xx.xxxxx/xxxx.xx.xx.xxxxxxxx, ©2026 Soltanifar, M. et al. (CC BY 4.0/MIT)

**Figure 5.1.12.**

```r
#[1] Packages
library(dplyr)
library(tidyr)
library(ggplot2)
library(scales)
library(tidytext)
set.seed(20260116)

#[2]AE dictionary (12 terms for brevity)
ae_terms <- c(
  "Neutropenia","Fatigue/lethargy","Infection (any)",
  "Biliary event","Hypertension","Vomiting","Acute kidney injury",
  "Diarrhoea","Anaemia","Febrile neutropenia",
  "Anorexia","Nausea"
)

#[3] helper to simulate a decreasing profile for ASC+FOLFOX and smaller for ASC alone
sim_panel <- function(type) {
  base <- seq(20, 2, length.out = length(ae_terms)) + rnorm(length(ae_terms), 0, 1.0)
  cf   <- if (type == "Chemo-related grade 3-5") c(12,11,10, 2,2,2,2,2,2,2,1,1) else base
  ascf <- pmax(round(cf, 1), 0.5)                                   # ASC+FOLFOX
  asca <- pmax(round(pmax(cf - runif(length(cf), 2, 8), 0), 1), 0.0)  # ASC alone smaller
  tibble(
    ae   = ae_terms,
    `ASC+FOLFOX` = ascf,
    `ASC alone`  = asca,
    type = type
  )
}

dat <- bind_rows(
  sim_panel("All grade 3-5"),
  sim_panel("Chemo-related grade 3-5")
)

#[4] reshape to long and mirror ASC alone to the left (negative sign)
plt <- dat |>
  pivot_longer(cols = c(`ASC alone`, `ASC+FOLFOX`), names_to = "arm", values_to = "pct") |>
  mutate(value = ifelse(arm == "ASC alone", -pct, pct)) |>
  group_by(type) |>
  mutate(ae = reorder_within(ae, abs(value)*(arm=="ASC+FOLFOX"), type)) |>
  ungroup()

# colors
cols <- c("ASC alone" = "#9ecae1", "ASC+FOLFOX" = "#fdae6b")

#[5] mirrored plot with two panels
Figure_A12 <- ggplot(plt, aes(x = value, y = ae, fill = arm)) +
  geom_col(width = 0.7, color = "grey40") +
  geom_vline(xintercept = 0, linewidth = 0.6) +
  geom_vline(xintercept = c(-5, 5), linetype = "dashed", color = "grey50") +
```

```r
  scale_fill_manual(values = cols, name = NULL) +
  scale_x_continuous(
    breaks = seq(-25, 25, by = 5),
    labels = function(x) paste0(abs(x), "%"),
    limits = c(-25, 25),
    expand = expansion(mult = c(0.01, 0.02))
  ) +
  scale_y_discrete(labels = scales::label_wrap(25)) +
  facet_wrap(~ type, ncol = 2, scales = "free_y") +
  labs(
    x = "Patients (%)",
    y = NULL,
    title = "Grade 3-5 adverse events and chemotherapy-related toxicity"
  ) +
  theme_minimal(base_size = 11) +
  theme(
    panel.grid.major.y = element_blank(),
    legend.position = "top",
    strip.text = element_text(face = "bold")
  )

Figure_A12

# [6] Save(Optional):
ggsave("Figure_A12.png", Figure_A12, width = 9, height = 6, dpi = 300)
ggsave("Figure_A12.pdf", Figure_A12, width=9, height=6, device = grDevices::cairo_pdf)
```

**Citation:** Soltanifar, M., Portuguese, A.J., Jeon, Y., Gauthier, J., & Lee, C.H. (2026). A North American Collaborative Atlas of Oncology Data Visualization with R Statistical Software. medRxiv preprint. https://doi.org/xx.xxxxx/xxxx.xx.xx.xxxxxxxx, ©2026 Soltanifar, M. et al. (CC BY 4.0/MIT)

**Figure 5.1.13.**

```r
#[1] Packages
library(dplyr)
library(tidyr)
library(ggplot2)
library(scales)
set.seed(20260116)

# [2] Data Simulation
# Expanded list with 6 new distinctive oncology AE terms
terms <- c(
"Nausea", "Diarrhea", "Fatigue", "Neutropenia", "Anemia", "Peripheral neuropathy",
"Alopecia", "Stomatitis", "Rash", "AST increased", "Arthralgia", "Infusion related reaction"
)

trts <- c("Y", "Z", "A", "B")  # four study treatments

# Baseline rates for all 12 terms
base <- c(70, 45, 85, 55, 40, 30, 60, 25, 35, 20, 30, 15)

# Term-by-treatment effects (Updated tribble with all 12 terms)
eff <- tibble::tribble(
  ~term,                        ~Y,  ~Z,  ~A,  ~B,
  "Nausea",                     -5, +20, +10, +15,
  "Diarrhea",                   -3, +18,  +6, +12,
  "Fatigue",                    -4, +15,  +8, +10,
  "Neutropenia",                -2, +10, +18, +12,
  "Anemia",                     -2,  +8, +12, +10,
  "Peripheral neuropathy",      -1,  +6, +20, +14,
  "Alopecia",                   +5, +25,  -5, +10,
  "Stomatitis",                 +2, +10,  +5,  +8,
  "Rash",                      +15,  +5, +25, +10,
  "AST increased",              +8,  +4, +12,  +6,
  "Arthralgia",                 +5, +15,  +2, +20,
  "Infusion related reaction", +12, +2,  +8,  +5
)

# Simulate counts per (term, treatment)
df <- expand.grid(term = terms, treatment = trts, KEEP.OUT.ATTRS = FALSE) |>
  as_tibble() |>
  left_join(
    eff |>
      pivot_longer(cols = all_of(trts), names_to = "treatment", values_to = "delta"),
    by = c("term", "treatment")
  ) |>
  mutate(
    # Correctly mapping baseline by indexing the terms vector
    base_val = base[match(term, terms)],
    n = pmax(0L, round(base_val + delta + rnorm(n(), 0, 4)))
  ) |>
  select(term, treatment, n)
```

```r
# [3] Heatmap (counts with labels)
Figure_A13 <- ggplot(df, aes(x = treatment, y = term, fill = n)) +
  geom_tile(color = "white", linewidth = 0.6) +
  geom_text(aes(label = paste0("N=", n)), size = 3, color = "black") +
  scale_fill_gradientn(
    colours = c("#FFF7BC", "#FEC44F", "#FB9A29", "#E34A33", "#B30000"),
    values  = rescale(c(0, 20, 45, 70, 110)), # Adjusted scale for higher counts
    breaks  = c(0, 20, 45, 70, 110),           # <--- Add this line
    limits  = c(0, 110),
    name    = "Number of Events"
  ) +
  labs(
    title = "Heat map for number of adverse events per preferred term",
    subtitle = "Summarized Oncology Safety Profiles",
    x = "Treatment",
    y = "Adverse Event Term"
  ) +
  theme_minimal(base_size = 11) +
  theme(
    panel.grid = element_blank(),
    axis.title.y = element_text(margin = margin(r = 8)),
    axis.title.x = element_text(margin = margin(t = 6)),
    plot.title = element_text(face = "bold")
  )

Figure_A13

# [4] Save (Optional):
ggsave("Figure_A13.png", Figure_A13, width=7, height=7, dpi = 300)
ggsave("Figure_A13.pdf", Figure_A13, width=7, height=7, device = grDevices::cairo_pdf)
```

**Citation:** Soltanifar, M., Portuguese, A.J., Jeon, Y., Gauthier, J., & Lee, C.H. (2026). A North American Collaborative Atlas of Oncology Data Visualization with R Statistical Software. medRxiv preprint. https://doi.org/xx.xxxxx/xxxx.xx.xx.xxxxxxxx, ©2026 Soltanifar, M. et al. (CC BY 4.0/MIT)

**Figure 5.1.14.**

```r
# [1] Packages
library(dplyr)
library(tidyr)
library(ggplot2)
library(forcats)
set.seed(20260116)

# [2] Simulate patient-specific dose change schedules
n_pat <- 15
# Use sprintf to add leading zeros: Patient_01, Patient_02, ...
patients <- paste0("Patient_", sprintf("%02d", 1:n_pat))

# Expanded dose grid
dose_levels <- c(25,50, 75, 100, 125, 150, 175, 200)

sim_sched <- function(id, start_dose = 100) {
  n_chg <- sample(2:3, 1)
  days  <- sort(sample(c(0, seq(10, 80, by = 5)), n_chg, replace = FALSE))
  step  <- sample(c(-25, 0, +25), n_chg, replace = TRUE, prob = c(.35, .20, .45))
  path  <- pmin(pmax(start_dose + cumsum(step), min(dose_levels)), max(dose_levels))
  tibble(patient_id = id, day = days, dose = path)
}

# Assign starting doses:
# 1-7: original (~100)
# 8-11: low-dose (< 75)
# 12-15: high-dose (> 150)
starts <- c(rep(100, 7), rep(50, 4), rep(175, 2), rep(200, 2))

raw_list <- mapply(sim_sched, patients, starts, SIMPLIFY = FALSE)
raw <- bind_rows(raw_list)

# Close trajectories with admin end day 85
raw <- raw %>%
  group_by(patient_id) %>%
  arrange(day, .by_group = TRUE) %>%
  group_modify(~ add_row(.x, day = 85, dose = dplyr::last(.x$dose))) %>%
  ungroup()

# Apply vertical offsets based on the now naturally-sorting patient IDs
offset_tbl <- tibble(
  patient_id = patients,
  offset = seq(0, by = 2, length.out = n_pat)
)

dat <- raw %>%
  left_join(offset_tbl, by = "patient_id") %>%
  mutate(
    dose_plot = dose + offset,
    patient_id = fct_inorder(patient_id)
  )
```

```
# [3] Build step chart
Figure_A14 <- ggplot(dat, aes(x = day, y = dose_plot, color = patient_id, group = patient_id)) +
  geom_step(linewidth = 1.05) +
  scale_color_discrete(name = "Patient ID") +
  scale_x_continuous(breaks = seq(0, 85, 10)) +
  scale_y_continuous(limits = c(0, 250), breaks =seq(0,250,25), labels = seq(0,250,25)) +
  labs(
    title = "Oncology Dose Step Chart",
    subtitle = "",
    x = "Study Day",
    y = "Dose Level (slightly offset for visibility)"
  ) +
  theme_minimal(base_size = 11) +
  theme(
    legend.position = "right",
    panel.grid.minor = element_blank()
  )

Figure_A14

#[4] Save(Optional)  :
ggsave("Figure_A14.png", Figure_A14, width =6, height = 6, dpi = 300)
ggsave("Figure_A14.pdf", Figure_A14, width=6, height=6, device = grDevices::cairo_pdf)
```

**Citation:** Soltanifar, M., Portuguese, A.J., Jeon, Y., Gauthier, J., & Lee, C.H. (2026). A North American Collaborative Atlas of Oncology Data Visualization with R Statistical Software. medRxiv preprint. https://doi.org/xx.xxxxx/xxxx.xx.xx.xxxxxxxx, ©2026 Soltanifar, M. et al. (CC BY 4.0/MIT)

**Figure 5.1.15.**

```
# [1] Packages
library(dplyr)
library(tidyr)
library(ggplot2)
set.seed(20260121)

# [2] Simulate a simple 3-class setting (benign, borderline, invasive)
N <- 1200
x1 <- rnorm(N); x2 <- runif(N, -1, 1)

# Linear predictors -> softmax probabilities (sum to 1)
lp_ben <- +1.1 - 0.6*x1 + 0.8*x2
lp_bor <- -0.7 + 1.4*x1 + 0.4*x2
lp_inv <-  1.0 + 0.8*x1 + 0.2*x2
denom  <- exp(lp_ben) + exp(lp_bor) + exp(lp_inv)

p_ben <- exp(lp_ben)/denom
p_bor <- exp(lp_bor)/denom
p_inv <- exp(lp_inv)/denom

#[3] Draw observed outcomes from these probabilities
draw_one <- function(pb, pr, pi) sample(c("benign","borderline","invasive"), 1,
                                        prob = c(pb, pr, pi))
y <- mapply(draw_one, p_ben, p_bor, p_inv)

dat <- tibble(
  id = 1:N, y,
  p_benign = p_ben, p_borderline = p_bor, p_invasive = p_inv
)

# [4] Prepare per-class (one-vs-rest) calibration data (kept for completeness)
long <- dat |>
  pivot_longer(cols = starts_with("p_"),
               names_to = "class", values_to = "p_hat") |>
  mutate(class = sub("^p_", "", class),
         y_bin = as.integer(y == class))

#  [4] Replace LOESS with stylized curves on a common grid
grid <- tibble(p_hat = seq(0, 1, length.out = 201))

# Benign: upward parabola above 45-degree line
benign_curve <- grid %>%
  mutate(
    class = "benign",
    obs   = pmin(1, p_hat - 0.950 * (p_hat - 0.5)^2 + 0.25)
  )

# Invasive: upside-down parabola below 45-degree line
invasive_curve <- grid %>%
  mutate(
    class = "invasive",
```

```r
    obs   = pmax(0, p_hat + 0.950 * (p_hat - 0.5)^2 - 0.25)
  )

# Borderline: smooth wave between benign and invasive curves
borderline_curve <- grid %>%
  mutate(
    class = "borderline",
    ben = pmin(1, p_hat + 0.30 * (p_hat - 0.5)^2 + 0.02),
    inv = pmax(0, p_hat - 0.30 * (p_hat - 0.5)^2 - 0.02),
    mid = 0.5 * (ben + inv),
    wig = 0.5 * (ben - inv) * sin(5 * pi * 2.5 * p_hat),
    obs = pmin(ben, pmax(inv, mid + wig))
  ) %>%
  select(p_hat, class, obs)

smooth_curves <- bind_rows(benign_curve, borderline_curve, invasive_curve)

# [5] Plot: diagonal (perfect calibration) + three smooth class curves
cols <- c(
  invasive   = "#EC7014",  # orange
  borderline = "#2B6CB0",    # blue
  benign    =   "#238B45"   # green
)

Figure_A15 <- ggplot(smooth_curves, aes(x = p_hat, y = obs, color = class)) +
  geom_abline(slope = 1, intercept = 0,
              linetype = 2, linewidth = 0.6, color = "grey40") +
  geom_line(linewidth = 1.2) +
  scale_color_manual(
    values = cols,
    breaks = c("benign", "borderline", "invasive"),
    labels = c("Benign", "Borderline", "Invasive")
  ) +
  coord_equal(xlim = c(0, 1), ylim = c(0, 1), expand = FALSE) +
  labs(
    x = "Predicted Probability",
    y = "Observed Proportion",
    color = "Outcome",
    title = "Multinomial Calibration Plot (stylized curves)"
  ) +
  theme_minimal(base_size = 13) +
  theme(panel.grid.minor = element_blank())



Figure_A15

# [6] Save(Optional):

ggsave("Figure_A15.png", Figure_A15, width = 6, height = 6, dpi = 300)
ggsave("Figure_A15.pdf", Figure_A15, width=6, height=6, device = grDevices::cairo_pdf)
```

**Citation:** Soltanifar, M., Portuguese, A.J., Jeon, Y., Gauthier, J., & Lee, C.H. (2026). A North American Collaborative Atlas of Oncology Data Visualization with R Statistical Software. medRxiv preprint. https://doi.org/xx.xxxxx/xxxx.xx.xx.xxxxxxxx, ©2026 Soltanifar, M. et al. (CC BY 4.0/MIT)

**Figure 5.1.16.**

```r
# [1] Packages
library(survival)
library(survivalROC)
library(dplyr)
library(ggplot2)
set.seed(20260121)

# [2] Simulate data resembling a toxicity setting
n  <- 460
# Baseline marker (RILA-like, lower values -> higher risk)
rila <- pmax(0, rnorm(n, mean = 14, sd = 6))

# Link marker to hazard: lower marker => higher hazard
linpred <- scale(-rila)                  # negative so lower RILA => higher risk
lambda0 <- 0.07                                        # baseline event rate
hr      <- exp(0.6 * linpred)
time    <- rexp(n, rate = lambda0 * hr) * 60           # months
cens    <- runif(n, min = 12, max = 55)                # administrative censoring
status  <- as.integer(time <= cens)
ftime   <- pmin(time, cens)

dat <- data.frame(rila, ftime, status)

# ---- Helper: ROC(t), AUC(t) and Youden threshold ----
tdroc <- function(tm){
  roc <- survivalROC(
    Stime = dat$ftime,
    status = dat$status,
    marker = -dat$rila,           # higher marker => higher risk
    predict.time = tm,
    method = "NNE",
    span = 0.25                   # REQUIRED for NNE
  )

  youden <- which.max(roc$TP - roc$FP)

  list(
    time = tm,
    auc  = roc$AUC,
    fpr  = roc$FP,
    tpr  = roc$TP,
    thr  = roc$cut.values[youden],
    sen  = roc$TP[youden],
    spe  = 1 - roc$FP[youden]
  )
}

times <- c(12, 24, 36, 50)
res   <- lapply(times, tdroc)

# ---- Summary table (AUC + Youden cutpoint)
```

```r
summary_tbl <- do.call(rbind, lapply(res, function(x)
  data.frame(
    time        = x$time,
    AUC         = round(x$auc, 3),
    threshold   = round(x$thr, 2),
    sensitivity = round(x$sen, 3),
    specificity = round(x$spe, 3)
  )
))
summary_tbl

# [3]  Smooth ROC(t) curves (ggplot2) + AUC labels over each curve

# Long ROC-curve data
roc_long <- bind_rows(lapply(res, function(x){
  data.frame(time = x$time, FPR = x$fpr, TPR = x$tpr)
})) %>%
  mutate(time_f = factor(time, levels = times, labels = paste0(times, " month")))

# AUC per time (for labeling)
auc_tbl <- bind_rows(lapply(res, function(x){
  data.frame(time = x$time, AUC = x$auc)
})) %>%
  mutate(time_f = factor(time, levels = times, labels = paste0(times, " month")),
         label  = paste0("AUC=", sprintf("%.3f", AUC)))

# Pick a point ON each curve to place the AUC label
auc_pos <- roc_long %>%
  group_by(time_f) %>%
  slice(which.min(abs(FPR - 0.25))) %>%
  ungroup() %>%
  left_join(auc_tbl %>% select(time_f, label), by = "time_f") %>%
  mutate(x_lab =  c(0.8,0.4,0.2,0.6))   %>%
  mutate(y_lab =  c(0.8,0.4,0.2,0.6))

# [4] Plot
Figure_A16 <- ggplot(roc_long, aes(x = FPR, y = TPR, color = time_f, group = time_f)) +
  geom_abline(slope = 1, intercept = 0, linetype = 2, linewidth = 0.6, color = "grey50") +
  geom_line(linewidth = 1.2) +
  # AUC labels over curves
  geom_text(
    data = auc_pos,
    aes(x = x_lab , y = y_lab, label = label, color = time_f),
    inherit.aes = FALSE,
    size = 3.8,
    fontface = "bold",
    show.legend = FALSE
  ) +
  coord_equal(xlim = c(0, 1), ylim = c(0, 1), expand = FALSE) +
  labs(
    title = "Time-dependent ROC curves(ROC(t))",
    x = "1-Specificity",
    y = "Sensitivity",
```

```r
    color = "Prediction Time"
  ) +
  theme_minimal(base_size = 12) +
  theme(panel.grid.minor = element_blank(),
        legend.position = "right")

Figure_A16

# [5] Save(Optional):
ggsave("Figure_A16.png", Figure_A16, width = 9, height = 6, dpi = 300)
ggsave("Figure_A16.pdf", Figure_A16, width=9, height=6, device = grDevices::cairo_pdf)
```

**Citation:** Soltanifar, M., Portuguese, A.J., Jeon, Y., Gauthier, J., & Lee, C.H. (2026). A North American Collaborative Atlas of Oncology Data Visualization with R Statistical Software. medRxiv preprint. https://doi.org/xx.xxxxx/xxxx.xx.xx.xxxxxxxx, ©2026 Soltanifar, M. et al. (CC BY 4.0/MIT)

**Figure 5.1.17.**

```
# [1] Packages
library(dplyr)
library(ggplot2)
library(precrec)
set.seed(20260121)

#  [2] Simulate an imbalanced oncology-like dataset
n  <- 10000
pi <- 0.18                           # event prevalence ~6% (The Baseline)
y  <- rbinom(n, 1, pi)

eta_neg <- rnorm(n,  mean = -2.0, sd = 1.0)
eta_pos <- rnorm(n,  mean =  0.0, sd = 1.0)

score_A <- plogis(ifelse(y==1, eta_pos + 0.8, eta_neg - 0.2))
score_B <- plogis(ifelse(y==1, eta_pos + 0.2, eta_neg))
score_C <- plogis(ifelse(y==1, eta_pos - 0.2, eta_neg + 0.1))

# [3] Compute PR curves and AUPRC
mm  <- mmdata(scores = list(score_A, score_B, score_C),
              labels = y, modnames = c("Logistic", "RandomForest", "XGBoost"))
em  <- evalmod(mm, raw_curves = TRUE)

# Note: fortify uses singular "curvetype"
df  <- fortify(em) %>% filter(curvetype == "PRC")

# Note: auc() requires plural "curvetypes" and "modnames"
au <- auc(em) %>%
  filter(curvetypes == "PRC") %>%
  transmute(model = modnames, AUPRC = round(aucs, 3))

print(au)

#  [4] Plot PR curves

# Pick a point ON each curve to place the AUC label
auc_pos <- au %>% mutate(AUPRC = AUPRC + 0.001) %>%
  mutate(label_auc = paste0("AUC=", AUPRC )) %>%
  mutate(x_lab =  c(0.825,0.55,0.25))   %>%
  mutate(y_lab =  c(0.75,0.75,0.75))


cols <- c("Logistic"="#1f77b4", "RandomForest"="#d62728", "XGBoost"="#2ca02c")

Figure_A17 <- ggplot(df, aes(x = x, y = y, colour = modname)) +
  # ADDED: Baseline horizontal line at the prevalence (pi)
  geom_hline(yintercept = pi, linetype = "dashed", color = "grey",linewidth = 1.1) +
  geom_line(linewidth = 1.1) +
  # AUC labels over curves
  geom_text(
    data = auc_pos,
```

```r
    aes(x = x_lab , y = y_lab, label = label_auc, color = model),
    inherit.aes = FALSE,
    size = 3.8,
    fontface = "bold",
    show.legend = TRUE
  ) +
  coord_equal(xlim = c(0, 1), ylim = c(0, 1), expand = FALSE) +
    scale_colour_manual(values = cols, name = "Model") +
  labs(title = "Precision-Recall Curves (imbalanced outcome)",
       subtitle = paste0("Baseline Precision (prevalence) = ", pi),
       x = "Recall (Sensitivity)",
       y = "Precision (PPV)") +
  theme_minimal(base_size = 12) +
  theme(legend.position = "right")

Figure_A17

#[5] Save(Optional):
ggsave("Figure_A17.png", Figure_A17, width = 7.5, height = 5, dpi = 300)
ggsave("Figure_A17.pdf", Figure_A17, width=7.5, height=5, device = grDevices::cairo_pdf)
```

**Figure 5.1.18.**

```r
#[1] Packages
library(dplyr)
library(ggplot2)
set.seed(20260122)
# If preferred: install.packages("DecisionCurve")  # authors' package (JCO 2016)

#  [2] Simulate an oncology-like data set (rare-ish event)
n  <- 4000
P  <- 0.10                                     # event prevalence ~10%
y  <- rbinom(n, 1, P)

# Linear predictors (better separation for cases)
eta0 <- rnorm(n, mean = -2.2, sd = 1.0)       # controls
eta1 <- rnorm(n, mean = -0.3, sd = 1.0)       # cases
lp   <- ifelse(y == 1, eta1, eta0)
phat <- plogis(lp)                              # model-predicted risk in [0,1]

# ---- 2) Net Benefit function (per Kerr et al., JCO 2016) ----
nb_curve <- function(y, phat, thr) {
  P <- mean(y == 1)
  sapply(thr, function(R){
    pred_pos <- phat >= R
    TPR <- ifelse(sum(y==1)==0, 0, mean(pred_pos[y==1]))
    FPR <- ifelse(sum(y==0)==0, 0, mean(pred_pos[y==0]))
    NB  <- TPR*P - (R/(1-R))*FPR*(1-P)
    NB
  })
}

# Threshold grid (focus where decisions are plausible)
thr <- seq(0, 0.30, by = 0.005)

# Model NB
nb_model <- nb_curve(y, phat, thr)
# Treat-all NB: TPR = 1, FPR = 1
Pprev <- mean(y==1)
nb_all <- Pprev - (thr/(1-thr))*(1-Pprev)
# Treat-none NB: zero line
nb_none <- rep(0, length(thr))

dplot <- tibble(
  threshold = thr,
  `Risk model` = nb_model,
  `Treat all`  = nb_all,
  `Treat none` = nb_none
) |>
  tidyr::pivot_longer(-threshold, names_to = "strategy", values_to = "NB")

#  [3] Plot (smooth curves; no points)
cols <- c("Risk model"="#1f77b4", "Treat all"="#7f7f7f", "Treat none"="#000000")
lt   <- c("Risk model"="solid",  "Treat all"="solid",  "Treat none"="solid")
```

```r
Figure_A18 <- ggplot(dplot, aes(threshold, NB, colour = strategy, linetype = strategy)) +
  geom_line(linewidth = 1.5) +
  coord_equal(xlim = c(0, 0.32), ylim = c(-0.355, +0.155), expand = FALSE) +
  geom_vline(xintercept = Pprev, colour = "grey60", linetype = "dotted", linewidth = 1.0) +
  scale_colour_manual(values = cols, name = "Strategy") +
  scale_linetype_manual(values = lt,  name = "Strategy") +
  labs(
    title = "Decision Curve Analysis: \n(Net Benefit vs. Threshold Probability)",
    x = "Threshold Probability (risk cut-off)",
    y = "Net Benefit",
    caption = "Dotted vertical line marks prevalence;
                at R = prevalence, 'treat all' and 'treat none' intersect.") +
  theme_minimal(base_size = 12) +
  theme(
    legend.position = "right",
    plot.caption = element_text(hjust = 0.5)
  )

Figure_A18

# [4]Save (Optional):
ggsave("Figure_A18.png", Figure_A18, width=7.5, height=5, dpi=300)
ggsave("Figure_A18.pdf", Figure_A18, width=7.5, height=5, device = grDevices::cairo_pdf)
```

**Figure 5.1.19.**

```r
# [1] Packages
library(tidyverse)
library(binom)
library(patchwork)
set.seed(123)

# [2] Simulation logic
n <- 1000
dat <- tibble(
  size_cm = pmax(0, rnorm(n, 15, 10)),
  mitotic_high = rbinom(n, 1, 0.4),
  site = factor(sample(c("Stomach", "SmallIntestine", "Colon/Rectum"), n,
                       replace = TRUE),
                levels = c("Stomach", "SmallIntestine", "Colon/Rectum")))

# True coefficients for simulation (Higher values = Lower RFS probability)
lp_true <- 3.5 - 0.12 * dat$size_cm - 4.5 * dat$mitotic_high -
  1.5 * (dat$site == "SmallIntestine") - 3.5 * (dat$site == "Colon/Rectum")
dat$RFS5y <- rbinom(n, 1, plogis(lp_true))

# [3] Fit Model
fit <- glm(RFS5y ~ size_cm + mitotic_high + site, data = dat, family = binomial())
betas <- coef(fit)

# --- MATHEMATICAL CORRECTIONS START HERE ---

# 1. Scaling Factor (pt_per_logit)
# We find the variable with the largest absolute effect across its range to be
# our "100 point" anchor.
rng_size <- c(0, 45)
effect_size <- abs(betas["size_cm"] * diff(rng_size))
effect_mitosis <- abs(betas["mitotic_high"])
effect_site_si <- abs(betas["siteSmallIntestine"])
effect_site_cr <- abs(betas["siteColon/Rectum"])

# Determine which effect is the maximum to set the 100-point scale
max_effect <- max(effect_size, effect_mitosis, effect_site_si, effect_site_cr)
pt_per_logit <- 100 / max_effect

# 2. Map predictor values to points based strictly on model betas
# Points = |Beta * Value| * pt_per_logit
get_pts_size <- function(x) abs(betas["size_cm"] * x) * pt_per_logit
get_pts_mitosis <- function(m) abs(betas["mitotic_high"] * m) * pt_per_logit

# Mapping categorical levels (Stomach is reference/0 points)
site_pts_vals <- c(
  "Stomach" = 0,
  "SmallIntestine" = abs(betas["siteSmallIntestine"]) * pt_per_logit,
  "Colon/Rectum" = abs(betas["siteColon/Rectum"]) * pt_per_logit
)
```

```r
# 3. Probability Axis Mapping
# In a logistic model: logit(P) = Beta0 + Beta1*X1 + ...
# In our point system, Total Points (S) = Sum(|Beta_i * Xi| * pt_per_logit)
# Since all our betas (except intercept) are negative: LP = Beta0 - (S / pt_per_logit)
# Therefore, to find the Points (S) for a specific probability P:
# S = (Beta0 - logit(P)) * pt_per_logit
lp_intercept <- betas["(Intercept)"]
probs <- c(0.95, 0.9, 0.8, 0.7, 0.6, 0.5, 0.4, 0.3, 0.2, 0.1, 0.05)
prob_pts <- (lp_intercept - qlogis(probs)) * pt_per_logit

# --- END OF MATHEMATICAL CORRECTIONS ---

# [4A] Nomogram Plotting
draw_ruler <- function(y, x_range, df, title, tick_dir = 1) {
  df <- df %>% filter(!is.na(x) & is.finite(x) & x >= x_range[1] & x <= x_range[2])
  list(
    annotate("segment", x = x_range[1], xend = x_range[2],
             y = y, yend = y, linewidth = 0.7),
    annotate("segment", x = df$x, xend = df$x,
             y = y, yend = y + (0.25 * tick_dir), linewidth = 0.6),
    annotate("text", x = df$x, y = y + (0.6 * tick_dir), label = df$lab, size = 3,
             vjust = ifelse(tick_dir == 1, 0, 1)),
    annotate("text", x = x_range[1], y = y + 1.1, label = title,
             hjust = 0, fontface = "bold", size = 3.5)
  )
}

# Prepare dataframes for the plot
pts_df   <- data.frame(x = seq(0, 100, 10), lab = seq(0, 100, 10))
size_df  <- data.frame(x = get_pts_size(c(0, 5, 10, 15, 25, 35, 45)),
                       lab = c(0, 5, 10, 15, 25, 35, 45))
mit_df   <- data.frame(x = c(0, get_pts_mitosis(1)),
                       lab = c("<5/50 HPF", ">=5/50 HPF"))
site_df  <- data.frame(x = as.numeric(site_pts_vals),
                       lab = c("Stomach", "Small Intestine", "Colon/Rectum"))
total_pts_df <- data.frame(x = seq(0, 200, 20), lab = seq(0, 200, 20))
prob_df  <- data.frame(x = prob_pts, lab = paste0(probs * 100))

g_nom <- ggplot() +
  draw_ruler(12, c(0, 100), pts_df, "Points") +
  draw_ruler(10, c(0, 100), size_df, "Size (cm)") +
  draw_ruler(8, c(0, 100), mit_df, "Mitotic index") +
  draw_ruler(6, c(0, 100), site_df, "Site") +
  draw_ruler(3, c(0, 200), total_pts_df, "Total points") +
  draw_ruler(1, c(0, 200), prob_df, "Probability of 5-year RFS (%)") +
  coord_cartesian(xlim = c(-10, 210), ylim = c(0, 14)) +
  theme_void() +
  labs(title = "A. Corrected Simplified Nomogram")

g_nom <- g_nom +
  theme(plot.margin = margin(10, 10, 10, 10))
```

```r
# [4B] Calibration Plotting
dat$p_pred <- predict(fit, type = "response")
calib <- dat %>%
  mutate(dec = ntile(p_pred, 10)) %>%
  group_by(dec) %>%
  summarise(p_hat = mean(p_pred)*100, obs = mean(RFS5y)*100,
            lo = binom.confint(sum(RFS5y), n(), method="wilson")$lower*100,
            hi = binom.confint(sum(RFS5y), n(), method="wilson")$upper*100)

g_cal <- ggplot(calib, aes(x = p_hat, y = obs)) +
  geom_abline(slope = 1, intercept = 0, linetype = "dashed", color = "red", linewidth = 1) +
  geom_errorbar(aes(ymin = lo, ymax = hi), width = 2, color = "blue", linewidth = 1) +
  geom_line(color = "blue", linewidth = 1) +
  geom_point(shape = 21, fill = "gold", color = "blue", size = 3) +
  labs(x = "Nomogram-predicted RFS (%)", y = "Observed 5-year RFS (%)",
       title = "B. Calibration Plot") +
  theme_classic() +
  coord_cartesian(xlim = c(0, 100), ylim = c(0, 100))

g_cal <- g_cal +
  theme(plot.margin = margin(10, 10, 10, 10))

#[5A] Display Combined Plot-Horizontal
Figure_A19_1<- g_nom + g_cal + plot_layout(widths = c(1.5, 1.5))  #Horizontal
Figure_A19_1
#[5B] Save (Optional)
ggsave("Figure_A19_1.png", Figure_A19_1, width=15, height=7.5, dpi=300)
ggsave("Figure_A19_1.pdf", Figure_A19_1, width=10.5, height=10.5, device = grDevices::cairo_pdf)


#[5C] Display Combined Plot-Vertical
Figure_A19_2<- g_nom / g_cal + plot_layout(heights = c(1.2, 1))    #Vertical
Figure_A19_2

#[5D] Save (Optional)
ggsave("Figure_A19_2.png", Figure_A19_2, width=10.5, height=10.5, dpi=300)
ggsave("Figure_A19_2.pdf", Figure_A19_2, width=10.5, height=10.5, device = grDevices::cairo_pdf)
```

**Citation:** Soltanifar, M., Portuguese, A.J., Jeon, Y., Gauthier, J., & Lee, C.H. (2026). A North American Collaborative Atlas of Oncology Data Visualization with R Statistical Software. medRxiv preprint. https://doi.org/xx.xxxxx/xxxx.xx.xx.xxxxxxxx, ©2026 Soltanifar, M. et al. (CC BY 4.0/MIT)

**Figure 5.1.20.**

```
#[1] Packages
library(tidyverse)
library(ggplot2)
set.seed(20)

# [2] Simulate enrollment data
# To match the image, we simulate a process that accelerates slightly over time
T_end   <- 600
t_cens  <- 360  # Census time (vertical dashed line)
N_total <- 1150

# Generating times that match the curvature in the image
u <- sort(runif(N_total))
enroll_time <- T_end * (u^(1.3)) # Convex curve to match the image

dat <- tibble(
  patient_id = sprintf("P%04d", seq_len(N_total)),
  enroll_time = enroll_time
)

# [3] Calculate actual cumulative accrual (Step function)
# The black line in the image shows the discrete steps of actual accrual
df_actual <- dat %>%
  arrange(enroll_time) %>%
  mutate(accrual = row_number())

# [4] Poisson-Gamma (PG) model-based predictive mean + 95% bands
# We use data up to t_cens to estimate the model
dat_at_census <- dat %>% filter(enroll_time <= t_cens)
N_c <- nrow(dat_at_census)

# Model parameters (adjusted slightly to match the image's red curve slope)
alpha0 <- 2
beta0  <- 1/50
alpha_post <- alpha0 + N_c
beta_post  <- beta0  + t_cens

# Prediction grid
grid <- tibble(time = seq(0, T_end, length.out = 200)) %>%
  mutate(
    # Using a power-law style mean to match the non-linear red curve in the image
    # Note: A standard PG model is linear; the image shows an accelerating target.
    mean = (alpha_post / beta_post) * (time^1.15) / (t_cens^0.15),
    prob = beta_post / (beta_post + time),
    lo   = qnbinom(p = 0.025, size = alpha_post, prob = prob) * (time/t_cens)^0.5,
    hi   = qnbinom(p = 0.975, size = alpha_post, prob = prob) * (time/t_cens)^0.2
  )

# [5] Plotting to match the attached image
Figure_A20 <- gp20 <- ggplot() +
  # 1. Red Rug Plot
```

```r
  geom_point(data = dat, aes(x = enroll_time, y = 0),
             shape = "+", color = "red", size = 3) +

  # 2. Actual Accrual (Black Step Line) - Mapping to 'Actual' for legend
  geom_step(data = df_actual, aes(x = enroll_time, y = accrual, color = "Actual",
                                  linetype = "Actual"), linewidth = 0.6) +

  # 3. Predictive Bands (Red Dashed Lines) - Mapping to '95% Band' for legend
  geom_line(data = grid, aes(x = time, y = lo, color = "95% CI", linetype = "95% CI")) +
  geom_line(data = grid, aes(x = time, y = hi, color = "95% CI", linetype = "95% CI")) +

  # 4. Predictive Mean (Red Solid Line) - Mapping to 'Mean' for legend
  geom_line(data = grid, aes(x = time, y = mean, color = "Predictive Mean",
                             linetype = "Predictive Mean"), linewidth = 0.8) +

  # 5. Census Line (Vertical Dashed)
  geom_vline(xintercept = t_cens, linetype = "dashed", color = "black") +

  # Manual scales to define colors and line types for the legend
  scale_color_manual(name = "Accrual Status",
  values = c("Actual" = "black", "Predictive Mean" = "red", "95% CI" = "red")) +
  scale_linetype_manual(name = "Accrual Status",
  values = c("Actual" = "solid", "Predictive Mean" = "solid", "95% CI" = "dashed")) +

  # Formatting
  scale_x_continuous(limits = c(0, 600), breaks = seq(0, 600, 100), expand = c(0,0)) +
  scale_y_continuous(limits = c(0, 1200), breaks = seq(0, 1200, 200), expand = c(0,0)) +
  labs(
    title = "Accrual over Time with PG-model Forecast",
    x = "Time Since Activation (days)",
    y = "Accrual (cumulative enrolled)"
  ) +
  theme_classic(base_size = 12) +
  theme(
    panel.grid.major = element_blank(),
    panel.grid.minor = element_blank(),
    panel.border = element_rect(colour = "black", fill=NA, linewidth=1),
    axis.ticks.length = unit(0.2, "cm"),
    legend.position = "right" # Places legend on the right side
  )


Figure_A20

#[6] Save (Optional)
ggsave("Figure_A20.png", Figure_A20, width=9.0, height=6.0, dpi=300)
ggsave("Figure_A20.pdf", Figure_A20, width=9.0, height=6.0, device = grDevices::cairo_pdf)
```

**Citation:** Soltanifar, M., Portuguese, A.J., Jeon, Y., Gauthier, J., & Lee, C.H. (2026). A North American Collaborative Atlas of Oncology Data Visualization with R Statistical Software. medRxiv preprint. https://doi.org/xx.xxxxx/xxxx.xx.xx.xxxxxxxx, ©2026 Soltanifar, M. et al. (CC BY 4.0/MIT)

**Figure 5.1.21.**

```
#[1] Packages
library(rpact)
library(ggplot2)
library(patchwork)

# [2] Define shared parameters
info_rates <- c(0.25, 0.50, 0.738, 1.0)
alpha_val <- 0.025 # 1-sided

# [3] Create the 3 Designs
d_of <- getDesignGroupSequential(typeOfDesign = "asOF",
                                 informationRates = info_rates, alpha = alpha_val)
d_p  <- getDesignGroupSequential(typeOfDesign = "asP",
                                 informationRates = info_rates, alpha = alpha_val)
d_hp <- getDesignGroupSequential(typeOfDesign = "HP",
                                 informationRates = info_rates, alpha = alpha_val)

# [4] Extract data into a clean data frame for plotting
# We pull the Z-scores (criticalValues) and Alpha Spent (alphaSpent)
plot_data <- data.frame(
  InfRate = rep(info_rates, 3),
  Z_Score = c(d_of$criticalValues, d_p$criticalValues, d_hp$criticalValues),
  Alpha   = c(d_of$alphaSpent, d_p$alphaSpent, d_hp$alphaSpent),
  Design  = rep(c("O'Brien-Fleming (PROSPER)", "Pocock", "Haybittle-Peto"), each = 4)
)

# [5A] PLOT 1: BOUNDARY COMPARISON (Z-Scale)
gp21_1<-ggplot(plot_data, aes(x = InfRate, y = Z_Score, color = Design, group = Design)) +
  geom_line(linewidth = 1) +
  geom_point(size = 3) +
  scale_color_manual(values = c("O'Brien-Fleming (PROSPER)" = "black",
                                "Pocock" = "red",
                                "Haybittle-Peto" = "blue")) +
  labs(title = "A. Group-Sequential Boundary Comparison (Z-Scale)",
       x = "Information Fraction (Proportion of Events)",
       y = "Critical Value (Z-score)",
       color = "Boundary Type") +
  theme_minimal() +
  geom_hline(yintercept = 1.96, linetype = "dashed", alpha = 0.5) +
  annotate("text", x = 0.9, y = 1.8, label = "p = 0.05 (Z=1.96)", size = 3)

gp21_1 <-gp21_1 +  theme(plot.margin = margin(2, 2, 2, 2))

#  [5B] PLOT 2: ALPHA SPENDING COMPARISON
gp21_2 <- ggplot(plot_data, aes(x = InfRate, y = Alpha,
                                color = Design, group = Design)) +
  geom_line(linewidth= 1) +
  geom_point(size = 3) +
  scale_color_manual(values = c("O'Brien-Fleming (PROSPER)" = "black",
                                "Pocock" = "red",
                                "Haybittle-Peto" = "blue")) +
```

```r
  labs(title = "B. Alpha Spending Function Comparison",
       x = "Information Fraction (Proportion of Events)",
       y = "Cumulative Alpha Spent",
       color = "Design Type") +
  theme_minimal()

gp21_2 <-gp21_2 +  theme(plot.margin = margin(2, 2, 2, 2))

#[6A] Display Combined Plot-Vertical
Figure_A21<- gp21_1 / gp21_2 + plot_layout(heights = c(1, 1))    #Vertical
Figure_A21

#[6B] Save (Optional)
ggsave("Figure_A21.png", Figure_A21, width=8, height=8, dpi=300)
ggsave("Figure_A21.pdf", Figure_A21, width=8, height=8, device = grDevices::cairo_pdf)
```

**Figure 5.1.22.**

```r
#[1] Packages
library(ggplot2)
library(patchwork)
library(dplyr)
set.seed(123)

#[2] Simulation Setup

N_max <- 40
p_null <- 0.20
p_target <- 0.40

#[2A] Simulate a drug that actually has a 35% response rate
true_p <- 0.35
outcomes <- rbinom(N_max, 1, true_p)
cum_responses <- cumsum(outcomes)
n_obs <- 1:N_max

#[2B] Function to calculate Bayesian Predictive Probability (BPP)
# Using a Beta-Binomial conjugate prior (Alpha=1, Beta=1)
calc_bpp <- function(n, x, N_max, p_null) {
  rem_n <- N_max - n
  # Current Posterior: Beta(1 + x, 1 + n - x)
  # We want to know: at N_max, will we be > 95% sure p > p_null?
  # For simplicity, we calculate probability that final response rate > p_null

  # Simulate possible future outcomes (Beta-Binomial)
  possible_future_x <- 0:rem_n
  prob_future_x <- dbetabinom <- function(k, size, a, b) {
    choose(size, k) * beta(k + a, size - k + b) / beta(a, b)
  }

  probs <- prob_future_x(possible_future_x, rem_n, 1 + x, 1 + n - x)
  final_x <- x + possible_future_x

  # Success defined as: posterior prob(p > p_null) > 0.95 at the end of trial
  success_at_end <- sapply(final_x, function(fx) {
    pbeta(p_null, 1 + fx, 1 + N_max - fx, lower.tail = FALSE) > 0.95
  })

  return(sum(probs[success_at_end]))
}

#[2C] Calculate BPP for each step in our simulated trial
bpp_values <- sapply(1:N_max, function(i) calc_bpp(n_obs[i], cum_responses[i], N_max, p_null))

trial_data <- data.frame(
  n = n_obs,
  responses = cum_responses,
  bpp = bpp_values
)
```

```r
#[2D]   Calculate Decision Boundaries for Panel B
# Find min and max responders needed at each 'n' to trigger decisions
boundary_data <- expand.grid(n = 1:N_max, x = 0:N_max) %>%
  filter(x <= n) %>%
  rowwise() %>%
  mutate(bpp_val = calc_bpp(n, x, N_max, p_null)) %>%
  mutate(zone = case_when(
    bpp_val < 0.1 ~ "Futility",
    bpp_val > 0.9 ~ "Success",
    TRUE ~ "Continue"
  ))

# [3]  Plotting

#[3A] Panel A: Win Probability Plot
gp22_1 <- ggplot(trial_data, aes(x = n, y = bpp)) +
  geom_line(color = "#2c3e50", size = 1) +
  geom_point(color = "#2c3e50") +
  geom_hline(yintercept = c(0.1, 0.9), linetype = "dashed", color = "red") +
  annotate("text", x = 5, y = 0.95, label = "Efficacy Threshold (0.9)",
           color = "red", size = 3) +
  annotate("text", x = 5, y = 0.05, label = "Futility Threshold (0.1)",
           color = "red", size = 3) +
  scale_y_continuous(limits = c(0, 1.05), breaks = seq(0, 1, 0.2)) +
  labs(title = "A. Bayesian Predictive Prpobability (BPP) Over 'Information Time'",
       subtitle = "Win Probability Style Plot (Predictive Probability of Trial Success)",
       x = "Sample Size (n)", y = "Predictive Probability (PP)") +
  theme_minimal()

#[3B] Panel B: Decision Boundary Plot
gp22_2  <- ggplot() +
  geom_tile(data = boundary_data, aes(x = n, y = x, fill = zone), alpha = 0.3) +
  geom_step(data = trial_data, aes(x = n, y = responses), size = 1, color = "black") +
  scale_fill_manual(values = c("Futility" = "#e74c3c",
                               "Continue" = "#bdc3c7", "Success" = "#2ecc71")) +
  labs(title = "B. Decision Boundaries",
       subtitle = "Observed Responders vs. Theoretical Stopping Zones",
       x = "Sample Size (n)", y = "Number of Responders (x)",
       fill = "Action Zone") +
  theme_minimal() +
  theme(legend.position = "bottom")

#[4A] Display Combined Plot-Vertical
Figure_A22<- gp22_1 / gp22_2 + plot_layout(heights = c(1, 1))    #Vertical
Figure_A22

#[4B] Save (Optional)
ggsave("Figure_A22.png", Figure_A22, width=8, height=8, dpi=300)
ggsave("Figure_A22.pdf", Figure_A22, width=8, height=8, device = grDevices::cairo_pdf)
```

**Citation:** Soltanifar, M., Portuguese, A.J., Jeon, Y., Gauthier, J., & Lee, C.H. (2026). A North American Collaborative Atlas of Oncology Data Visualization with R Statistical Software. medRxiv preprint. https://doi.org/xx.xxxxx/xxxx.xx.xx.xxxxxxxx, ©2026 Soltanifar, M. et al. (CC BY 4.0/MIT)

**Figure 5.1.23.**

```r
#[1] Packages
library(ggplot2)
set.seed(23)

#[2] Simulated OC data (16 scenarios; two target DLT rates; 3 designs)
scenarios <- 1:16
targets <- c(0.25, 0.30)
designs <- c("3+3", "mTPI", "BOIN")

#[3] Helper to create "jagged but plausible" OC patterns with no extra points
#(only 16 per panel)
make_curve <- function(target, design) {
  s <- scenarios

  if (target == 0.25) {
    if (design == "3+3") {
      base <- c(36, 34, 28, 26, 29, 27, 23, 22, 26, 27, 24, 21, 23, 25, 27, 24)
    } else if (design == "mTPI") {
      base <- c(52, 45, 37, 34, 44, 40, 32, 33, 42, 40, 30, 36, 38, 41, 63, 56)
    } else { # BOIN
      base <- c(43, 38, 35, 33, 40, 34, 29, 30, 39, 33, 29, 31, 36, 39, 67, 63)
    }
  } else { # target == 0.30
    if (design == "3+3") {
      base <- c(32, 35, 23, 25, 25, 27, 20, 22, 26, 27, 20, 22, 24, 26, 16, 23)
    } else if (design == "mTPI") {
      base <- c(51, 61, 41, 48, 49, 46, 56, 39, 47, 46, 42, 49, 48, 57, 61, 71)
    } else { # BOIN
      base <- c(42, 55, 37, 47, 40, 44, 52, 37, 44, 41, 39, 47, 44, 53, 60, 70)
    }
  }

  # Add tiny noise but keep within [0, 100]; does NOT add points/markers.
  noise <- round(rnorm(length(base), mean = 0, sd = 1.0), 0)
  pmax(0, pmin(100, base + noise))
}

df <- do.call(
  rbind,
  lapply(targets, function(tar) {
    do.call(
      rbind,
      lapply(designs, function(des) {
        data.frame(
          scenario = scenarios,
          target_dlt = tar,
          design = des,
          pct_correct = make_curve(tar, des)
        )
      })
    )
```

```r
  })
)

# [4] Factor formatting for legend & facet labels
df$design <- factor(df$design, levels = c("3+3", "mTPI", "BOIN"))
df$target_dlt <- factor(df$target_dlt, levels = c(0.25, 0.30),
           labels = c("A. Target DLT rate = 25%", "B. Target DLT rate = 30%"))

# [5] Plot (Updated with professional colors)
Figure_A23 <- ggplot(df, aes(x = scenario, y = pct_correct, group = design, color = design)) +
  geom_line(aes(linetype = design)) +
  geom_point(aes(shape = design), size = 2) +
  facet_wrap(~ target_dlt, nrow = 2) +
  scale_x_continuous(breaks = 1:16) +
  scale_y_continuous(limits = c(0, 75), breaks = seq(0, 70, 10),
                     expand = expansion(mult = c(0.02, 0.05))) +
  # Adding professional color palette
  scale_color_manual(values = c("3+3" = "#000080",
                                "mTPI" = "#2CA02C", "BOIN" = "#D62728")) +
  labs(
    x = "Scenario",
    y = "Percentage of Correct Selection (%)",
    linetype = "Design",
    shape = "Design",
    color = "Design"
  ) +
  theme_bw() +
  theme(
    legend.position = "right",
    panel.grid.minor = element_blank(),
    strip.background = element_rect(fill = "white"),
    plot.caption = element_text(hjust = 0)
  )

Figure_A23

#[6] Save (Optional)
ggsave("Figure_A23.png", Figure_A23, width=6, height=9, dpi=300)
ggsave("Figure_A23.pdf", Figure_A23, width=6, height=9, device = grDevices::cairo_pdf)
```

**Citation:** Soltanifar, M., Portuguese, A.J., Jeon, Y., Gauthier, J., & Lee, C.H. (2026). A North American Collaborative Atlas of Oncology Data Visualization with R Statistical Software. medRxiv preprint. https://doi.org/xx.xxxxx/xxxx.xx.xx.xxxxxxxx, ©2026 Soltanifar, M. et al. (CC BY 4.0/MIT)

**Figure 5.1.24.**

```r
#[1] Packages
library(survival)
library(ggplot2)
library(dplyr)
library(tidyr)
library(patchwork)
set.seed(2401)


# [2] Simulate oncology-style TVC survival data

n <- 480
t_max <- 1500                # follow-up horizon (e.g., days)
p_switch <- 0.45             # probability subject ever switches XT: 0->1
hr_xt <- 0.45                # protective effect once XT=1 (HR < 1)
lambda0 <- 1/650             # baseline hazard rate (roughly median ~ 450 days)

#[3] switch times among switchers (late switches induce bias in naive KM)
switch_time <- ifelse(runif(n) < p_switch, runif(n, 80, 900), NA_real_)

#[4] simulate event time under piecewise exponential hazard:
# hazard = lambda0 for t < switch_time, lambda0 * hr_xt for t >= switch_time
#(if switch occurs)
rexp_piecewise <- function(rate0, hr, s, tmax) {
  u <- runif(1)
  if (is.na(s)) {
    # no switch
    t <- -log(u) / rate0
    return(min(t, tmax))
  }
  # cumulative hazard up to s: H(s)=rate0*s
  Hs <- rate0 * s
  # draw event time based on piecewise hazard
  # if event occurs before s:
  if (-log(u) <= Hs) {
    t <- (-log(u)) / rate0
    return(min(t, tmax))
  }
  # otherwise occurs after s with hazard rate0*hr
  # remaining cumulative hazard beyond s:
  u2 <- exp(-( -log(u) - Hs ))  # not used directly; keep exact approach
  # time after s:
  t_after <- ( (-log(u)) - Hs ) / (rate0 * hr)
  t <- s + t_after
  return(min(t, tmax))
}

true_event_time <- vapply(seq_len(n), function(i) {
  rexp_piecewise(rate0 = lambda0, hr = hr_xt, s = switch_time[i], tmax = t_max)
}, numeric(1))
```

```r
# independent censoring
cens_time <- runif(n, 300, t_max)
time <- pmin(true_event_time, cens_time)
status <- as.integer(true_event_time <= cens_time)

dat <- tibble(
  id = seq_len(n),
  time = time,
  status = status,
  xt_change_time = switch_time
)


#  [5A] Panel A: Naive KM (Naive Cox-style grouping)
#    Naive grouping treats XT as if known/fixed at baseline.

# "Naive" group indicator (ever switches at any time) -> classic immortal-time bias setup
dat <- dat %>%
  mutate(XT_value_naive = factor(ifelse(!is.na(xt_change_time), 1, 0),
                                 levels = c(0, 1),
                                 labels = c("XT_value=0", "XT_value=1")))

sf_naive <- survfit(Surv(time, status) ~ XT_value_naive, data = dat, conf.type = "log")


# helper to convert survfit to a ggplot-friendly dataframe
sf_to_df <- function(sf, group_name = "group") {
  s <- summary(sf)
  out <- tibble(
    time = s$time,
    surv = s$surv,
    lower = s$lower,
    upper = s$upper,
    group = factor(s$strata)
  )
  # FIXED: Use sub with a non-greedy match to only remove the first variable name prefix
  # This converts "XT_value_naive=XT_value=0" to "XT_value=0"
  out[[group_name]] <- sub("^.*?=", "", out$group)
  out <- out %>% select(-group)
  out[[group_name]] <- factor(out[[group_name]])
  out
}

df_naive <- sf_to_df(sf_naive, "XT_value")


# [5B] Panel B: Landmark KM (Landmark Cox-style)

landmark_time <- 300

lm_dat <- dat %>%
  mutate(alive_at_landmark = (time > landmark_time)) %>%
  filter(alive_at_landmark) %>%
```

```r
  mutate(
    # status and time re-defined from landmark onward
    time_lm = time - landmark_time,
    status_lm = status,
    # XT at landmark: has switched before landmark?
    XT_value_lm = factor(ifelse(!is.na(xt_change_time) & xt_change_time <= landmark_time, 1, 0),
                         levels = c(0, 1),
                         labels = c("XT_value=0", "XT_value=1"))
  )

sf_lm <- survfit(Surv(time_lm, status_lm) ~ XT_value_lm, data = lm_dat, conf.type = "log")
df_lm <- sf_to_df(sf_lm, "XT_value") %>%
  mutate(time = time + landmark_time)  # put on original time scale for plotting


# [5C] Panels C & D: Time-dependent Cox (TD Cox) + Smith-Zee + Extended KM

# Build counting-process (start, stop] data: XT(t)=0 until switch, then 1
# Two rows per subject if switch occurs before censor/event time
make_tdc <- function(d) {
  if (is.na(d$xt_change_time) || d$xt_change_time >= d$time) {
    return(tibble(id = d$id, tstart = 0, tstop = d$time, event = d$status, xt = 0))
  } else {
    rbind(
      tibble(id = d$id, tstart = 0, tstop = d$xt_change_time, event = 0, xt = 0),
      tibble(id = d$id, tstart = d$xt_change_time, tstop = d$time, event = d$status, xt = 1)
    )
  }
}
tdc <- dat %>% group_split(id) %>% purrr::map_dfr(~make_tdc(.x))

# TD Cox
cox_td <- coxph(Surv(tstart, tstop, event) ~ xt, data = tdc, ties = "efron")

# Baseline cumulative hazard for Smith-Zee path computations
bh <- basehaz(cox_td, centered = FALSE) %>% as_tibble() %>% rename(H0 = hazard, t = time)

# Helper: linear interpolation of H0(t)
H0_at <- function(tt) {
  approx(x = bh$t, y = bh$H0, xout = tt, method = "linear", rule = 2)$y
}

beta_hat <- unname(coef(cox_td)["xt"])
se_beta  <- sqrt(vcov(cox_td)["xt","xt"])

# Time grid for smooth-looking curves (model-based)
t_grid <- seq(0, t_max, by = 10)

# --- Extended KM (TD Cox): treat xt as fixed (0 vs 1) in newdata
# survfit.coxph gives model-based survival + CI
sf_ext0 <- survfit(cox_td, newdata = data.frame(xt = 0))
sf_ext1 <- survfit(cox_td, newdata = data.frame(xt = 1))
```

```r
ext_to_df <- function(sf_obj, label) {
  s <- summary(sf_obj, times = t_grid)
  tibble(time = s$time, surv = s$surv, lower = s$lower, upper = s$upper,
         XT_value = factor(label, levels = c("XT_value=0","XT_value=1")))
}

df_ext <- bind_rows(
  ext_to_df(sf_ext0, "XT_value=0"),
  ext_to_df(sf_ext1, "XT_value=1")
)

# --- Smith-Zee (TD Cox): marginalize over observed switch-time distribution
# Survival under a path with switch time s:
#   S(t|s) = exp( -[ H0(min(t,s)) + exp(beta) * (H0(t)-H0(s))_+ ] )
S_path <- function(t, s, beta) {
  if (is.na(s) || t <= s) {
    return(exp(-H0_at(t)))
  }
  Hs <- H0_at(s)
  Ht <- H0_at(t)
  exp(-(Hs + exp(beta) * (Ht - Hs)))
}

switchers <- dat %>% filter(!is.na(xt_change_time)) %>% pull(xt_change_time)

smith_curve <- function(beta) {
  # XT_value=0: never switch
  S0 <- exp(-H0_at(t_grid))
  # XT_value=1: average over observed switch times (no new markers; just curves)
  S1 <- vapply(t_grid,
function(tt) mean(vapply(switchers, function(s) S_path(tt, s, beta), numeric(1))),
               numeric(1))
  tibble(
    time = rep(t_grid, 2),
    surv = c(S0, S1),
    XT_value = factor(rep(c("XT_value=0", "XT_value=1"), each = length(t_grid)),
                      levels = c("XT_value=0","XT_value=1"))
  )
}

# point estimate + simple beta-based band (keeps curves smooth, avoids bootstrapped jitter)
df_sz_hat <- smith_curve(beta_hat)
df_sz_lo  <- smith_curve(beta_hat - 1.96*se_beta)
df_sz_hi  <- smith_curve(beta_hat + 1.96*se_beta)

# Get the correct CI for the baseline group (XT=0) from the Cox survfit object
df_baseline_ci <- ext_to_df(sf_ext0, "XT_value=0") %>%
  select(time, XT_value, lower, upper)

df_sz <- df_sz_hat %>%
  left_join(df_sz_lo %>% rename(lower = surv), by = c("time","XT_value")) %>%
  left_join(df_sz_hi %>% rename(upper = surv), by = c("time","XT_value")) %>%
  rows_update(df_baseline_ci, by = c("time", "XT_value"))
```

```r
# [6] Plotting (2x2)

palette_cols <- c("XT_value=0" = "#E76F51",  # red-ish: worse
                  "XT_value=1" = "#2A9D8F")  # teal: better

base_theme <- theme_minimal(base_size = 12) +
  theme(
    legend.position = "top",
    legend.title = element_blank(),
    plot.title = element_text(face = "bold", size = 12),
    panel.grid.minor = element_blank()
  )

p_naive <- ggplot(df_naive, aes(x = time, y = surv, color = XT_value, fill = XT_value)) +
  geom_ribbon(aes(ymin = lower, ymax = upper), alpha = 0.20, color = NA) +
  geom_step(linewidth = 0.9) +
  scale_color_manual(values = palette_cols) +
  scale_fill_manual(values = palette_cols) +
  coord_cartesian(ylim = c(0, 1)) +
  labs(title = "Naive KM (Naive Cox)", x = "Time", y = "Survival probability") +
  base_theme

p_lm <- ggplot(df_lm, aes(x = time, y = surv, color = XT_value, fill = XT_value)) +
  geom_ribbon(aes(ymin = lower, ymax = upper), alpha = 0.20, color = NA) +
  geom_step(linewidth = 0.9) +
  geom_vline(xintercept = landmark_time, linetype = "dashed", linewidth = 0.6) +
  annotate("text", x = landmark_time, y = 0.05, label = "Landmark",
           angle = 90, vjust = -0.5, size = 3) +
  scale_color_manual(values = palette_cols) +
  scale_fill_manual(values = palette_cols) +
  coord_cartesian(ylim = c(0, 1)) +
  labs(title = "Landmark KM (Landmark Cox)", x = "Time", y = "Survival probability") +
  base_theme

p_sz <- ggplot(df_sz, aes(x = time, y = surv, color = XT_value, fill = XT_value)) +
  geom_ribbon(aes(ymin = pmax(0, lower), ymax = pmin(1, upper)), alpha = 0.20, color = NA) +
  geom_line(linewidth = 0.9) +
  scale_color_manual(values = palette_cols) +
  scale_fill_manual(values = palette_cols) +
  coord_cartesian(ylim = c(0, 1)) +
  labs(title = "Smith-Zee (Time-dependent Cox)", x = "Time", y = "Survival probability") +
  base_theme

p_ext <- ggplot(df_ext, aes(x = time, y = surv, color = XT_value, fill = XT_value)) +
  geom_ribbon(aes(ymin = lower, ymax = upper), alpha = 0.20, color = NA) +
  geom_line(linewidth = 0.9) +
  scale_color_manual(values = palette_cols) +
  scale_fill_manual(values = palette_cols) +
  coord_cartesian(ylim = c(0, 1)) +
  labs(title = "Extended KM (Time-dependent Cox)", x = "Time", y = "Survival probability") +
  base_theme
```

```r
Figure_A24 <- (p_naive | p_lm) / (p_sz | p_ext) +
  plot_annotation(
    title = "KM plots of models",
    theme = theme(plot.title = element_text(face = "bold", size = 14))
  )

Figure_A24

#[7] Save (Optional)
ggsave("Figure_A24.png", Figure_A24, width=9, height=9, dpi=300)
ggsave("Figure_A24.pdf", Figure_A24, width=9, height=9, device = grDevices::cairo_pdf)
```

**Citation:** Soltanifar, M., Portuguese, A.J., Jeon, Y., Gauthier, J., & Lee, C.H. (2026). A North American Collaborative Atlas of Oncology Data Visualization with R Statistical Software. medRxiv preprint. https://doi.org/xx.xxxxx/xxxx.xx.xx.xxxxxxxx, ©2026 Soltanifar, M. et al. (CC BY 4.0/MIT)

#### Section B. Real World Evidence (RWE)

**Figure 5.2.1.**

```r
#[1] Packages
library(ggplot2)
set.seed(1234)

# [2] Simulated covariate list (style consistent with the example)
covariates <- c(
  "Propensity score (logit)",
  "Age (years)",
  "Race: White",
  "Race: Black",
  "Race: Asian",
  "Race: American Indian/Alaska Native",
  "Race: Native Hawaiian/Pacific Islander",
  "Race: Unknown",
  "Ethnicity: Non-Hispanic",
  "Charlson-Deyo comorbidity score (CDCC)",
  "Insurance: Uninsured",
  "Insurance: Private",
  "Insurance: Medicaid",
  "Insurance: Medicare",
  "Insurance: Other government",
  "Insurance: Unknown",
  "Urbanicity: Urban",
  "Urbanicity: Metro",
  "Urbanicity: Rural",
  "Urbanicity: Unknown",
  "Diagnosis year: 2014-2017",
  "Tumor grade: Low (I-II)",
  "Tumor grade: High (III-IV)",
  "Tumor grade: Unknown",
  "Lymphovascular invasion: Absent",
  "Lymphovascular invasion: Present",
  "Lymphovascular invasion: Unknown",
  "Age category (>=60 vs <60)",
  "Tumor size category (>=1 cm vs <1 cm)",
  "Estrogen receptor category"
)

# [3] Simulate ASMDs: larger pre-match, smaller post-match (typical love-plot pattern)
n_cov <- length(covariates)

asmd_unadj <- pmin(pmax(rnorm(n_cov, mean = 0.18, sd = 0.12), 0.00), 0.55)
asmd_adj   <- pmin(pmax(rnorm(n_cov, mean = 0.05, sd = 0.03), 0.00), 0.12)

df <- data.frame(
  covariate = covariates,
  Unadjusted = asmd_unadj,
  Adjusted   = asmd_adj,
  stringsAsFactors = FALSE
```

```r
)

# [4] Order covariates by Unadjusted ASMD (descending), as is common in love plots
df$covariate <- factor(df$covariate,
                levels = df$covariate[order(df$Unadjusted, decreasing = TRUE)])

# Long format without extra dependencies
df_long <- rbind(
  data.frame(covariate = df$covariate, Sample = "Adjusted", ASMD = df$Adjusted+0.05),
  data.frame(covariate = df$covariate, Sample = "Unadjusted", ASMD = df$Unadjusted)
)

# [5] Plot
Figure_B01 <- ggplot(df_long,
                  aes(x = ASMD, y = covariate, shape = Sample, color = Sample)) +
  geom_vline(xintercept = 0.10, linetype = "dashed") + # Balance threshold
  geom_point(size = 2.5) +          # Increased size slightly for visibility
  scale_x_continuous(limits = c(0, 0.55), breaks = seq(0, 0.5, 0.1)) +

  # --- UPDATED COLORS AND SHAPES WITH TITLE ---
  scale_color_manual(values = c("Adjusted" = "forestgreen", "Unadjusted" = "red"),
                     name = "Status") +
  scale_shape_manual(values = c("Adjusted" = 17, "Unadjusted" = 16),
                     name = "Status") +
  # -------------------------------------------

labs(
  title = "Arm 2 Covariate Balance (Unadjusted vs Adjusted (PSM))",
  x = "Absolute Standardized Mean Differences",
  y = NULL
) +
  theme_bw(base_size = 11) +
  theme(
    # Removed legend.title = element_blank() to allow the title to show
    legend.position = "right",
    plot.title = element_text(face = "bold"),
    panel.grid.minor = element_blank()
  )

Figure_B01

#[6] Save (Optional)
ggsave("Figure_B01.png", Figure_B01, width=8, height=8, dpi=300)
ggsave("Figure_B01.pdf", Figure_B01, width=8, height=8, device = grDevices::cairo_pdf)
```

**Figure 5.2.2.**

```r
# [1] Packages
library(ggplot2)
library(dplyr)

# [2] Data Preparation with Randomized/Modified Values
# We modify roughly 1/3 of the values to be > 1.0
raw_data <- tibble::tribble(
  ~subgroup, ~level,               ~events, ~patients, ~HR,   ~LCL,  ~UCL,   ~is_header,
  "Overall", NA,                    235,      616,      0.49,  0.38,  0.64,  TRUE,
  "Age",     NA,                    NA,       NA,       NA,    NA,    NA,    TRUE,
  "Age",     "<65 yr",              133,      312,      0.43,  0.31,  0.61,  FALSE,
  "Age",     ">=65 yr",              102,      304,      1.45,  1.10,  1.95,  FALSE,
  "Sex",     NA,                    NA,       NA,       NA,    NA,    NA,    TRUE,
  "Sex",     "Male",                143,      363,      0.70,  0.50,  0.99,  FALSE,
  "Sex",     "Female",               92,      253,      0.29,  0.19,  0.44,  FALSE,
  "ECOG performance-status score", NA, NA,    NA,       NA,    NA,    NA,    TRUE,
  "ECOG",    "0",                    74,      266,      0.44,  0.28,  0.71,  FALSE,
  "ECOG",    "1",                   159,      346,      1.32,  1.05,  1.65,  FALSE,
  "Smoking status", NA,             NA,       NA,       NA,    NA,    NA,    TRUE,
  "Smoking", "Current or former",   211,      543,      0.54,  0.41,  0.71,  FALSE,
  "Smoking", "Never",                24,       73,      0.23,  0.10,  0.54,  FALSE,
  "Brain metastases at baseline", NA, NA,     NA,       NA,    NA,    NA,    TRUE,
  "Brain mets", "Yes",               51,      108,      1.55,  1.20,  2.05,  FALSE,
  "Brain mets", "No",               184,      508,      0.53,  0.39,  0.71,  FALSE,
  "PD-L1 tumor proportion score", NA, NA,     NA,       NA,    NA,    NA,    TRUE,
  "PD-L1",   "<1%",                  84,      190,      0.59,  0.38,  0.92,  FALSE,
  "PD-L1",   ">=1%",                  135,      388,      0.47,  0.34,  0.66,  FALSE,
  "PD-L1",   "1-49%",                65,      186,      1.25,  1.02,  1.55,  FALSE,
  "PD-L1",   ">=50%",                 70,      202,      0.42,  0.26,  0.68,  FALSE,
  "Platinum-based drug", NA,        NA,       NA,       NA,    NA,    NA,    TRUE,
  "Platinum", "Carboplatin",        176,      445,      1.42,  1.15,  1.82,  FALSE,
  "Platinum", "Cisplatin",           59,      171,      0.41,  0.24,  0.69,  FALSE
)

# Process data for plotting and add Color Logic
plot_df <- raw_data %>%
  mutate(row_id = rev(row_number())) %>%
  mutate(
    # [LOGIC] Define colors based on HR and CI boundaries
    point_color = case_when(
      is.na(HR) ~ "black",
      UCL < 1.0 ~ "forestgreen", # Green if entire CI is below 1
      LCL > 1.0 ~ "red",          # Red if entire CI is above 1
      TRUE      ~ "black"        # Black if it crosses 1.0
    ),
    disp_subgroup = ifelse(is_header, subgroup, paste0("   ", level)),
    disp_events   = ifelse(is.na(events), "", paste0(events, "/", patients)),
    disp_hr       = ifelse(is.na(HR), "", sprintf("%.2f (%.2f-%.2f)", HR, LCL, UCL))
  )

# Define X-axis positions for Table columns
```

```r
col1_x <- 0.015
col2_x <- 0.08
col3_x <- 6.5

# [3] Build the Plot
Figure_B02 <- ggplot(plot_df, aes(y = row_id)) +
  # 1. Background Shading
  geom_rect(aes(xmin = 0.01, xmax = 20, ymin = row_id - 0.5, ymax = row_id + 0.5),
            fill = ifelse(plot_df$row_id %% 2 == 0, "#f7f7f7", "white"), alpha = 0.5) +

  # 2. Forest Plot Elements (with mapped colors)
  geom_vline(xintercept = 1.0, color = "darkgray", linewidth = 0.8) +
  geom_errorbarh(aes(xmin = LCL, xmax = UCL, color = point_color),
                 height = 0, linewidth = 0.8) +
  geom_point(aes(x = HR, color = point_color), shape = 15, size = 3) +

  # 3. Apply the literal colors from the data frame
  scale_color_identity() +

  # 4. Table Columns (Subgroup, Events, HR text)
  geom_text(aes(x = col1_x, label = disp_subgroup), hjust = 0, size = 3.8,
            fontface = ifelse(plot_df$is_header, "bold", "plain")) +
  geom_text(aes(x = col2_x, label = disp_events), hjust = 0.5, size = 3.5) +
  geom_text(aes(x = col3_x, label = disp_hr), hjust = 0, size = 3.5) +

  # 5. Header Labels
  annotate("text", x = col1_x, y = max(plot_df$row_id) + 1.2,
           label = "Subgroup", fontface = "bold", hjust = 0) +
  annotate("text", x = col2_x, y = max(plot_df$row_id) + 1.2,
           label = "No. of Events/ No. of Patients", fontface = "bold", vjust = 0.5) +
  annotate("text", x = col3_x-3.5, y = max(plot_df$row_id) + 1.2,
           label = "Hazard Ratio for Death (95% CI)", fontface = "bold", hjust = 0) +

  # 6. Bottom Arrows
  annotate("segment", x = 0.8, xend = 0.15, y = -0.5, yend = -0.5,
           arrow = arrow(length = unit(0.2, "cm")), color = "forestgreen") +
  annotate("segment", x = 1.2, xend = 4, y = -0.5, yend = -0.5,
           arrow = arrow(length = unit(0.2, "cm")), color = "red") +
  annotate("text", x = 0.4, y = -1.5, label = "Pembrolizumab Combination\nBetter",
           size = 3.2, fontface = "bold", color = "forestgreen") +
  annotate("text", x = 2.2, y = -1.5, label = "Placebo Combination\nBetter",
           size = 3.2, fontface = "bold", color = "red") +

  # Styling
  scale_x_log10(limits = c(0.01, 20), breaks = c(0.1, 0.5, 1.0, 2.0, 5.0),
                labels = c("0.1", "0.5", "1.0", "2.0", "5.0")) +
  scale_y_continuous(limits = c(-2, max(plot_df$row_id) + 2)) +
  labs(title = "Subgroup Analysis of Overall Survival") +
  theme_void() +
  theme(
    plot.title = element_text(face = "bold", size = 14,
                              margin = margin(b = 20), hjust = 0.05),
    plot.margin = margin(10, 10, 10, 10),
```

```r
    plot.background = element_rect(fill = "white", color = NA)
  )

Figure_B02

#[4] Save (Optional)
ggsave("Figure_B02.png", Figure_B02, width=12, height=8, dpi=300)
ggsave("Figure_B02.pdf", Figure_B02, width=12, height=8, device = grDevices::cairo_pdf)
```

**Citation:** Soltanifar, M., Portuguese, A.J., Jeon, Y., Gauthier, J., & Lee, C.H. (2026). A North American Collaborative Atlas of Oncology Data Visualization with R Statistical Software. medRxiv preprint. https://doi.org/xx.xxxxx/xxxx.xx.xx.xxxxxxxx, ©2026 Soltanifar, M. et al. (CC BY 4.0/MIT)

**Figure 5.2.3.**

```
#[1] Packages
library(dplyr)
library(tidyr)
library(ggplot2)
set.seed(20260201)


#[2] Simulated PSA dataset

n_sim <- 6000

psa <- tibble(
  sim_id = 1:n_sim,
  # QALYs (positive)
  qaly_chemo = pmax(rnorm(n_sim, mean = 0.90, sd = 0.22), 0.01),
  qaly_nivo  = pmax(rnorm(n_sim, mean = 1.80, sd = 0.28), 0.01),
  # Costs (positive, $)
  cost_chemo = pmax(rnorm(n_sim, mean =  60000, sd = 12000), 1000),
  cost_nivo  = pmax(rnorm(n_sim, mean = 180000, sd = 25000), 1000)
)

# WTP grid ($/QALY)
wtp_grid <- seq(0, 200000, by = 1000)


# [3] CEAC: P(strategy has highest NMB) for each WTP
# NMB = WTP * QALY - Cost

ceac_raw <- lapply(wtp_grid, function(wtp) {

  nmb_chemo <- wtp * psa$qaly_chemo - psa$cost_chemo
  nmb_nivo  <- wtp * psa$qaly_nivo  - psa$cost_nivo

  # Winner per simulation at this WTP (tie-break: chemo)
  win <- ifelse(nmb_nivo > nmb_chemo, "Nivolumab + ipilimumab", "Chemotherapy")

  tibble(
    wtp = wtp,
    strategy = c("Chemotherapy", "Nivolumab + ipilimumab"),
    prob_ce = c(mean(win == "Chemotherapy"), mean(win == "Nivolumab + ipilimumab"))
  )
}) %>%
  bind_rows()


# [4] Smooth curves (spline) on a fine WTP grid

wtp_fine <- seq(min(wtp_grid), max(wtp_grid), by = 250)

ceac_smooth <- ceac_raw %>%
  group_by(strategy) %>%
```

```r
  summarise(
    spline_fun = list(stats::splinefun(x = wtp, y = prob_ce, method = "monoH.FC")),
    .groups = "drop"
  ) %>%
  tidyr::uncount(length(wtp_fine)) %>%
  group_by(strategy) %>%
  mutate(
    wtp = wtp_fine,
    prob_ce = pmin(pmax(spline_fun[[1]](wtp), 0), 1)
  ) %>%
  ungroup() %>%
  select(wtp, strategy, prob_ce)


# [5] Plot

wtp_threshold <- 100000

# Create a thinned dataset for the points so they don't overlap too much
ceac_points <- ceac_smooth %>%
  filter(wtp %% 10000 == 0) # Only keep points every $10,000

Figure_B03 <- ggplot(ceac_smooth, aes(x = wtp, y = prob_ce, color = strategy)) +
  geom_line(linewidth = 1.1) +

  # --- ADDED POINTS WITH SPECIFIC SHAPES ---
  geom_point(data = ceac_points, aes(shape = strategy), size = 3) +
  scale_shape_manual(values = c("Chemotherapy" = 17, "Nivolumab + ipilimumab" = 16)) +
  # -----------------------------------------

geom_vline(xintercept = wtp_threshold, linetype = "dashed", linewidth = 0.8) +
  annotate("text",
           x = wtp_threshold, y = 0.98,
           label = "Threshold = $100,000/QALY",
           angle = 0, vjust = 1, hjust = -0.02, size = 3.5) +
  scale_y_continuous(limits = c(0, 1), breaks = seq(0, 1, 0.25)) +
  scale_x_continuous(labels = function(x) paste0("$", format(x, big.mark = ","))) +

  # Ensure the legend shows both color and shape together
  guides(color = guide_legend(override.aes = list(shape = c(17, 16)))) +

  labs(
    title = "Cost-Effectiveness Acceptability Curve (CEAC)",
    x = "Willingness-to-Pay Threshold ($/QALY)",
    y = "Probability of Cost-Effectiveness",
    color = "Strategy",
    shape = "Strategy"
  ) +
  theme_minimal(base_size = 12) +
  theme(
    legend.position = "right",
    panel.grid.minor = element_blank()
  )
```

```r
Figure_B03

#[6] Save (Optional)
ggsave("Figure_B03.png", Figure_B03, width=8, height=8, dpi=300)
ggsave("Figure_B03.pdf", Figure_B03, width=8, height=8, device = grDevices::cairo_pdf)
```

**Figure 5.2.4.**

```
#[1] Packages
library(ggplot2)
library(dplyr)
library(tidyr)
set.seed(20260130)

# [2] Simulate a simplified one-way DSA result set
# Impacts are centered around base-case (0).
# Negative = decreases outcome; Positive = increases outcome.
n_params <- 18

params <- c(
  "Drug acquisition cost",
  "Hospitalization cost (PD state)",
  "Outpatient visit cost",
  "Imaging / monitoring cost",
  "AE management cost",
  "Utility in PF state",
  "Utility in PD state",
  "Disutility (grade 3-4 AE)",
  "PF→PD transition rate",
  "PD mortality rate",
  "Discount rate",
  "Time horizon",
  "Treatment duration",
  "Dose intensity / adherence",
  "Post-progression therapy cost",
  "Lab test cost",
  "Travel / indirect cost",
  "Administration cost"
)

# Create asymmetric min/max swings (typical in HEOR models)
base <- tibble(
  parameter = params[seq_len(n_params)],
  min_impact = -runif(n_params, 1, 25),   # e.g., -25% to -1%
  max_impact =  runif(n_params, 5, 90)    # e.g., +5% to +90%
)

# Rank by total swing to create the "tornado" ordering
base <- base %>%
  mutate(swing = abs(min_impact) + abs(max_impact)) %>%
  arrange(desc(swing)) %>%
  mutate(parameter = factor(parameter, levels = rev(parameter)))
          # top at top after coord_flip()

# Long format for two bars (Minimum vs Maximum) with a single legend
plot_df <- base %>%
  select(parameter, min_impact, max_impact) %>%
  pivot_longer(cols = c(min_impact, max_impact),
               names_to = "bound",
```

```r
               values_to = "impact") %>%
  mutate(
    bound = recode(bound,
                   min_impact = "Minimum",
                   max_impact = "Maximum"),
    bound = factor(bound, levels = c("Maximum", "Minimum"))
  )

# [3] Build the Tornado Diagram
Figure_B04 <- ggplot(plot_df, aes(x = impact, y = parameter, fill = bound)) +
  # Draw bars
  geom_col(width = 0.70, color = NA) +

  # --- ADDED TEXT LABELS ---
  geom_text(aes(label = paste0(round(impact, 1), "%"),
                hjust = ifelse(impact >= 0, -0.2, 1.2)),
            size = 3.2, color = "black") +
  # -------------------------

# Center reference line at 0%
geom_vline(xintercept = 0, linetype = "dashed", linewidth = 0.5) +

  # Custom Colors
  scale_fill_manual(
    name = "Bound",
    values = c("Maximum" = "#D55E00",
               "Minimum" = "#0072B2")
  ) +

  # --- UPDATED X-AXIS FOR NEGATIVE TICKS ---
  scale_x_continuous(
    limits = c(-70, 120),   # Expanded slightly to prevent label clipping
    breaks = seq(-50, 100, by = 25),          # Ticks
    labels = function(x) paste0(x, "%")       # Appends % to the numbers
  ) +
  # ----------------------------------------

labs(
  title = "Tornado Diagram (One-way Deterministic Sensitivity Analysis)",
  x = "Impact on Outcome vs Base-Case (%)",
  y = NULL
) +

  theme_minimal(base_size = 12) +
  theme(
    legend.position = "right",
    plot.title = element_text(face = "bold", hjust = 0.5),
    axis.text.y = element_text(color = "black", size = 10),
    axis.text.x = element_text(color = "black"),
    panel.grid.major.y = element_blank(),
    panel.grid.major.x = element_line(color = "gray90"),
                                            # Vertical lines for the ticks
    panel.grid.minor = element_blank(),
```

```r
    plot.margin = margin(10, 20, 10, 10)
  )

Figure_B04

#[4] Save (Optional)
ggsave("Figure_B04.png", Figure_B04, width=12, height=8, dpi=300)
ggsave("Figure_B04.pdf", Figure_B04, width=12, height=8, device = grDevices::cairo_pdf)
```

**Citation:** Soltanifar, M., Portuguese, A.J., Jeon, Y., Gauthier, J., & Lee, C.H. (2026). A North American Collaborative Atlas of Oncology Data Visualization with R Statistical Software. medRxiv preprint. https://doi.org/xx.xxxxx/xxxx.xx.xx.xxxxxxxx, ©2026 Soltanifar, M. et al. (CC BY 4.0/MIT)

**Figure 5.2.5.**

```
# [1] Packages
library(dplyr)
library(tidyr)
library(networkD3)
library(htmlwidgets)
library(webshot2)
set.seed(20260202)

# [2] Simulate Patient Data
N_total <- 3995

# 1L: Replaced "Clinical Trial" with "BTK Inhibitor"
reg_1L_levels <- c("Pembrolizumab combo (1L)", "Chemotherapy (1L)",
                   "Pembrolizumab mono (1L)",
                   "Nivolumab + Ipi (1L)", "Targeted Therapy (1L)",
                   "BTK Inhibitor (1L)",
                   "Radiation + IO (1L)", "Other (1L)")
p_1L <- c(0.35, 0.15, 0.12, 0.10, 0.10, 0.08, 0.05, 0.05)

pat <- tibble(
  id = seq_len(N_total),
  col1 = sample(reg_1L_levels, size = N_total, replace = TRUE, prob = p_1L)
)

pat <- pat %>%
  mutate(
    col2 = case_when(
      col1 != "" ~ sample(c("Chemotherapy (2L)", "IO Mono (2L)", "ADCs (2L)",
                            "TKI (2L)", "Death/Censored (post-1L)"),
                          size = n(), replace = TRUE,
                          prob = c(0.12, 0.10, 0.05, 0.03, 0.70))
    ),
    col3 = case_when(
 col2 %in% c("Chemotherapy (2L)", "IO Mono (2L)", "ADCs (2L)", "TKI (2L)") ~
 sample(c("Other (3L)", "Best Supportive Care (3L)", "Death/Censored (post-2L)"),
              size = n(), replace = TRUE, prob = c(0.20, 0.15, 0.65)),
      TRUE ~ NA_character_
    )
  )

# [3] Create Links
links1 <- pat %>% count(source = col1, target = col2, name = "value")
links2 <- pat %>% filter(!is.na(col3)) %>% count(source = col2,
                                                 target = col3, name = "value")
links <- bind_rows(links1, links2)

# [4] Create Nodes and Add Percentages
nodes_names <- unique(c(links$source, links$target))
nodes <- tibble(name = nodes_names)

node_counts <- links %>%
```

```r
  group_by(source) %>% summarise(n = sum(value)) %>% rename(node = source) %>%
  bind_rows(links %>% group_by(target) %>% summarise(n = sum(value))
            %>% rename(node = target)) %>%
  group_by(node) %>% summarise(n = max(n))

nodes <- nodes %>%
  left_join(node_counts, by = c("name" = "node")) %>%
  mutate(
    pct = round(n / N_total * 100, 1),
    display_label = paste0(name, " (", pct, "%)")
  )

# --- COLOR MAPPING ---
nodes <- nodes %>%
  mutate(group = case_when(
    grepl("Death", name)        ~ "death",
    grepl("mono", name)         ~ "mono",
    grepl("combo", name)        ~ "combo",
    grepl("Chemotherapy", name) ~ "chemo",
    grepl("Targeted", name)     ~ "targeted",
    grepl("TKI", name)          ~ "tki",
    grepl("BTK", name)          ~ "btk", # New group for the replacement
    grepl("ADC", name)          ~ "adc",
    TRUE                        ~ "other"
  ))

links_idx <- links %>%
  mutate(
    source_idx = match(source, nodes$name) - 1,
    target_idx = match(target, nodes$name) - 1,
    group = nodes$group[match(source, nodes$name)]
  )

my_color_map <- c(
  death    = "#FF0000", # Red
  mono     = "#FFFF00", # Yellow
  combo    = "#2ca02c", # Green
  chemo    = "#ff7f0e", # Orange
  targeted = "#9467bd", # Purple
  tki      = "#17becf", # Teal
  btk      = "#D4AF37", # Gold (replacing trial)
  adc      = "#8c564b", # Brown
  other    = "#1f77b4"  # Blue
)

my_colors_js <- sprintf(
  'd3.scaleOrdinal().domain(["%s"]).range(["%s"])',
  paste(names(my_color_map), collapse = '", "'),
  paste(my_color_map, collapse = '", "')
)

# [6] Create the 3-Column Plot
Figure_B05 <- sankeyNetwork(
```

```r
  Links = links_idx,
  Nodes = nodes,
  Source = "source_idx",
  Target = "target_idx",
  Value = "value",
  NodeID = "display_label",
  NodeGroup = "group",
  LinkGroup = "group",
  fontSize = 12,
  nodeWidth = 50,
  nodePadding = 15,
  width = 1100,
  height = 700,
  colourScale = JS(my_colors_js),
  iterations = 0,
  sinksRight = FALSE
)

# Render
Figure_B05

# [4] Save  (Optional)

saveWidget(Figure_B05, "sankey_plot.html", selfcontained = TRUE)
webshot("sankey_plot.html", "Figure_B05.png", vwidth = 1200, vheight = 600)
webshot("sankey_plot.html", "Figure_B05.pdf", vwidth = 1200, vheight = 600)
```

**Figure 5.2.6.**

```
# [1] Packages
library(circlize)
library(dplyr)
set.seed(606)

# [2] Simulations
# [2-1] Data Simulation & Matrix Construction
# Define categories and colors
tx_levels <- c("IBR", "VEN", "BR", "FCR", "RCI", "IDER", "VENR", "OCI")
grid_col <- c(IBR="#E41A1C", VEN="#377EB8", BR="#4DAF4A", FCR="#984EA3",
              RCI="#FF7F00", IDER="#FFFF33", VENR="#A65628", OCI="#F781BF")

# [2-2] Simulate treatment-switching table
base_edges <- tribble(
  ~from,   ~to,     ~lambda,
  "BR",    "IBR",   70, "BR",    "VEN",   30, "FCR",   "IBR",   45,
  "FCR",   "VEN",   25, "RCI",   "IBR",   40, "RCI",   "VEN",   20,
  "OCI",   "IBR",   25, "OCI",   "VEN",   18, "IBR",   "VEN",   55,
  "IBR",   "VENR",  15, "VEN",   "VENR",  22, "VEN",   "IDER",  10,
  "IDER",  "VEN",   12, "VENR",  "VEN",    8
)

# [2-3] Add rare background switches
bg_pairs <- expand.grid(from = tx_levels, to = tx_levels,
                        stringsAsFactors = FALSE) %>%
  filter(from != to) %>%
  mutate(key = paste(from, to, sep = "->"))

base_keys <- paste(base_edges$from, base_edges$to, sep = "->")
bg_edges <- bg_pairs %>%
  filter(!(key %in% base_keys)) %>%
  slice_sample(n = 14) %>%
  transmute(from, to, lambda = sample(2:8, size = n(), replace = TRUE))

edges <- bind_rows(base_edges, bg_edges) %>%
  mutate(n = rpois(n(), lambda)) %>%
  filter(n > 0)

# [2-4] CREATE THE MISSING OBJECT 'M'
M <- matrix(0, nrow = length(tx_levels), ncol = length(tx_levels),
            dimnames = list(tx_levels, tx_levels))
for(i in seq_len(nrow(edges))) {
  M[edges$from[i], edges$to[i]] <- M[edges$from[i], edges$to[i]] + edges$n[i]
}
M[M < 6] <- 0 # Filter minor flows

# [3] Plot
gp_06 <- function(mat, levels, colors) {
  circos.clear()
  circos.par(
    start.degree = 90,
```

```r
    gap.after = rep(8, length(levels)),
    track.margin = c(0.01, 0.05),
    points.overflow.warning = FALSE
  )

  chordDiagram(
    x = mat,
    grid.col = colors,
    transparency = 0.15,       # High-quality, sharp colors
    directional = 1,
    direction.type = c("diffHeight", "arrows"),
    link.arr.type = "big.arrow",
    diffHeight = mm_h(2),
    annotationTrack = c("grid", "axis"),
    preAllocateTracks = list(track.height = 0.05)
  )

  circos.trackPlotRegion(
    track.index = 1,
    panel.fun = function(x, y) {
      circos.text(
        x = CELL_META$xcenter, y = CELL_META$ylim + 1.5,
        labels = CELL_META$sector.index,
        facing = "bending.inside", niceFacing = TRUE,
        cex = 0.9, font = 2
      )
    },
    bg.border = NA
  )

  title("CLL Treatment Switching Patterns", cex.main = 1.4)
}

# [5] Save (Optional)
# Export to Sharp PNG
png("Figure_B06.png", width = 8, height = 8, units = "in", res = 600)
gp_06(M, tx_levels, grid_col)
dev.off()

# Export to Vector PDF
pdf("Figure_B06.pdf", width = 8, height = 8)
gp_06(M, tx_levels, grid_col)
dev.off()

# Display in current R session
gp_06(M, tx_levels, grid_col)
```

**Figure 5.2.7.**

```r
#[1] Packages
library(ggplot2)
set.seed(707)

# [2] Simulated longitudinal SLD for 10 patients
patients <- paste0("", sprintf("%02d", 1:10))

# Include a pre-baseline assessment (week = -4) and baseline (week = 0),
# then follow-up visits roughly like the example figure.
weeks <- c(-4, 0, 8, 16, 24, 32, 40, 48)

# Patient-specific baseline heterogeneity (absolute SLD in mm)
baseline_sld <- round(runif(length(patients), min = 40, max = 180), 0)

# Patient-specific slope patterns to create mixed shrinkage/growth/stable courses
slopes <- rnorm(length(patients), mean = 0.4, sd = 1.2)# mm per week(can be negative)

dat <- do.call(rbind, lapply(seq_along(patients), function(i) {
  pid <- patients[i]
  b0  <- baseline_sld[i]
  m   <- slopes[i]

#Smooth-ish trajectory with mild curvature + noise (still continuous; no markers added)
  sld <- b0 - m * (weeks - 0) + 0.015 * (weeks - 0)^2 + rnorm(length(weeks), 0, 6)

  # Keep SLD non-negative
  sld <- pmax(sld, 5)

  data.frame(
    patient_id = pid,
    week       = weeks,
    sld_mm     = sld
  )
}))

# Ensure correct ordering for line drawing
dat <- dat[order(dat$patient_id, dat$week), ]

# [3] Plot (lines only; legend on the right)
Figure_B07 <- ggplot(dat, aes(x = week, y = sld_mm,
                              group = patient_id, color = patient_id)) +
  geom_line(linewidth = 1.0) +
  geom_point(size = 3) +
  geom_vline(xintercept = 0, linetype = "dashed", linewidth = 0.8) +
  scale_x_continuous(breaks = c(-4, 0, 8, 16, 24, 32, 40, 48)) +
  scale_y_continuous(breaks = c( 0, 50, 100, 150, 200, 250, 300,350)) +
  labs(
    title = "Spaghetti Plot of Absolute Tumor Burden (SLD) Over Time",
    x = "Weeks",
    y = "SLD (mm)",
    color = "Patient"
```

```r
  ) +
  theme_bw() +
  theme(
    legend.position = "right",
    panel.grid.minor = element_blank()
  )

Figure_B07

# [4] Save  (Optional)
ggsave("Figure_B07.png", Figure_B07, width=9, height=6, dpi=300)
ggsave("Figure_B07.pdf", Figure_B07, width=9, height=6,
                                                device = grDevices::cairo_pdf)
```

**Citation:** Soltanifar, M., Portuguese, A.J., Jeon, Y., Gauthier, J., & Lee, C.H. (2026). A North American Collaborative Atlas of Oncology Data Visualization with R Statistical Software. medRxiv preprint. https://doi.org/xx.xxxxx/xxxx.xx.xx.xxxxxxxx, ©2026 Soltanifar, M. et al. (CC BY 4.0/MIT)

**Figure 5.2.8.**

```r
#[1] Packages
library(ggplot2)
library(dplyr)

#  [2] Simulation of Data
#[2-1] Data for Treatment Segments (The Bars)
# Includes 'Worsened' states as segments to match the journey visualization
segments_data <- data.frame(
  id = c(1, 1, 2, 2, 3, 3, 4, 4, 4, 4, 5, 5, 5, 6, 6, 6, 7, 7, 7,
         8, 8, 8, 9, 9, 9, 9, 10, 10),
  start = c(0, 17, 0, 16, 0, 15, 0, 13, 16, 26, 0, 10, 15, 0, 8, 13,
            0, 6, 11, 0, 5, 10, 0, 4, 9, 11, 0, 4),
  end = c(17, 20, 16, 20, 15, 18, 13, 16, 26, 27, 10, 15, 17, 8, 13, 16,
          6, 11, 20, 5, 10, 15, 4, 9, 11, 15, 4, 10),
  Treatment = c("Surgery", "Chemotherapy", "Surgery", "Chemotherapy",
                "Surgery", "Chemotherapy",
                "Surgery", "Chemotherapy", "Immunotherapy", "Immunotherapy",
                "Surgery", "Chemotherapy",
                "Immunotherapy", "Surgery", "Chemotherapy", "Worsened_B",
                "Surgery", "Chemotherapy",
                "Immunotherapy", "Surgery", "Chemotherapy", "Immunotherapy",
                "Surgery", "Chemotherapy",
                "Worsened_G", "Worsened_B", "Surgery", "Immunotherapy")
)

# [2-2] Data for Status Markers (Circles)
status_data <- data.frame(
  id = c(6, 9, 6, 9), # Including 'Worsened' to force them into the legend
  day = c(16.5, 15.5, NA, NA),
  Status = factor(c("Improved", "Not changed", "Worsened ", "Worsened"),
                  levels = c("Improved", "Not changed", "Worsened ", "Worsened"))
)

# [2-3] Data for Events (Diamonds)
event_data <- data.frame(
  id = c(10, 10),
  day = c(11.5, NA),
  Event = factor(c("Death", "Discharge"), levels = c("Death", "Discharge"))
)


# Color Palettes
fill_colors <- c("Surgery" = "#A12B2B", "Chemotherapy" = "#D3433E",
                 "Immunotherapy" = "#FBE09C",
                 "Worsened_B" = "#C2D5DB", "Worsened_G" = "#D1D1D1")

status_colors <- c("Improved" = "#6BA1D7", "Not changed" = "#555555",
                   "Worsened " = "#C2D5DB", "Worsened" = "#D1D1D1")
#segments_data$id <- sprintf("%02d", segments_data$id)
```

```r
# [3] Plotting

Figure_B08 <- ggplot() +
  # Draw Bars
  geom_rect(data = segments_data, aes(xmin = start, xmax = end,
                                      ymin = id - 0.3, ymax = id + 0.3,
                                      fill = Treatment), color = NA) +
  # Vertical Ref Line
  geom_vline(xintercept = 9.8, linetype = "dashed", color = "black", size = 1) +
  # Status Points (Circles)
  geom_point(data = status_data, aes(x = day, y = id, color = Status), size = 4) +
  # Event Points (Diamonds)
  geom_point(data = event_data, aes(x = day, y = id, shape = Event), size = 4,
             fill = "black", color = "black") +
  # Scales & Legend Formatting
  scale_y_reverse(breaks = 1:10) +
  scale_x_continuous(breaks = seq(0, 30, 2), limits = c(-2, 31)) +
  scale_fill_manual(name = "Line Treatment", values = fill_colors,
                    breaks = c("Surgery", "Chemotherapy", "Immunotherapy")) +
  scale_color_manual(name = "Status", values = status_colors) +
  scale_shape_manual(name = "Event", values = c("Death" = 18, "Discharge" = 5)) +
  # Labs
  labs(
    title = "Patient-Journey Timeline (Gantt-style)",
    x = "Day",
    y = "Patient\nNo."
  ) +
  # Theme Customization to match the original layout
  theme_minimal() +
  theme(
    plot.title = element_text(hjust = 0.5, size = 22),
    axis.title.y = element_text(angle = 0, vjust = 1, hjust = 0.5, size = 10),
    axis.title.x = element_text(size = 12),
    axis.line.x = element_line(color = "black"),
    panel.grid = element_blank(),
    legend.position = "bottom",
    legend.box = "vertical",
    legend.title = element_text(face = "bold", size = 11),
    legend.margin = margin(t = 20)
  ) +
  # Layout the legends in horizontal rows
  guides(
    fill = guide_legend(nrow = 1, order = 1, title.position = "left"),
    color = guide_legend(nrow = 1, order = 2, title.position = "left"),
    shape = guide_legend(nrow = 1, order = 3, title.position = "left")
  ) +
  # Label for the vertical line
  annotate("text", x = 11.0, y = 10.8, label = "Last day of R", size = 4, vjust = 0)


Figure_B08

# [4] Save  (Optional)
```

```
ggsave("Figure_B08.png", Figure_B08, width=8, height=8, dpi=300)
ggsave("Figure_B08.pdf", Figure_B08, width=8, height=8,dpi=600,
                                                device = grDevices::cairo_pdf)
```

**Citation:** Soltanifar, M., Portuguese, A.J., Jeon, Y., Gauthier, J., & Lee, C.H. (2026). A North American Collaborative Atlas of Oncology Data Visualization with R Statistical Software. medRxiv preprint. https://doi.org/xx.xxxxx/xxxx.xx.xx.xxxxxxxx, ©2026 Soltanifar, M. et al. (CC BY 4.0/MIT)

**Figure 5.2.9.**

```r
#[1] Packages
library(ggplot2)
set.seed(2026)

#[2] Simulated PSA / bootstrap-style draws (100 points)
n <- 100
wtp <- 50000  # $/QALY

# Generate correlated data
mu_E <- -0.05
mu_C <-  4000
sd_E <-  0.06
sd_C <-  3500
rho  <-  0.85

z1 <- rnorm(n)
z2 <- rnorm(n)
dE_qaly <- mu_E + sd_E * z1
dC_cost <- mu_C + sd_C * (rho * z1 + sqrt(1 - rho^2) * z2)

dat <- data.frame(
  id = sprintf("Pt%03d", 1:n),
  dE_qaly = dE_qaly,
  dC_cost = dC_cost
)

#[3] Updated CEP plot
Figure_B09 <- ggplot(dat, aes(x = dE_qaly, y = dC_cost, color = id)) +
  # [1] Add dashed black axes at origin to define quadrants
  geom_hline(yintercept = 0, linetype = "dashed", color = "black") +
  geom_vline(xintercept = 0, linetype = "dashed", color = "black") +

  geom_point(size = 2, alpha = 0.9) +

  # WTP threshold line: Delta C = WTP * Delta E
  geom_abline(intercept = 0, slope = wtp, linewidth = 1) +
  # Add "WTP" label along the line
  # Coordinates chosen to be visible in the bottom-right area of your data range
  annotate(
    "text",
    x = -0.025,
    y = -250, # (wtp * -0.05)
    label = "WTP Threshold",
    angle = 45,        # Adjust angle based on your plot's final aspect ratio
    vjust = 1.5,       # Pushes text slightly below the line
    fontface = "italic",
    size = 3.5
  ) +
  labs(
    title = "Cost-Effectiveness Plane (QALY scale)",
    x = expression(paste("Incremental Effectiveness ", Delta, "E (QALY)")),
```

```r
    y = expression(paste("Incremental Cost ", Delta, "C ($)")),
    # [2] Professional legend title
    color = "Patient No."
  ) +
  theme_classic() +
  theme(
    legend.position = "right",
    legend.key.height = grid::unit(0.35, "cm"),
    legend.text = element_text(size = 7)
  )

Figure_B09

# [4] Save  (Optional)
ggsave("Figure_B09.png", Figure_B09, width=9, height=6, dpi=300)
ggsave("Figure_B09.pdf", Figure_B09, width=9, height=6,dpi=600,
                                              device = grDevices::cairo_pdf)
```

**Citation:** Soltanifar, M., Portuguese, A.J., Jeon, Y., Gauthier, J., & Lee, C.H. (2026). A North American Collaborative Atlas of Oncology Data Visualization with R Statistical Software. medRxiv preprint. https://doi.org/xx.xxxxx/xxxx.xx.xx.xxxxxxxx, ©2026 Soltanifar, M. et al. (CC BY 4.0/MIT)

**Figure 5.2.10.**

```
# [1] Packages
library(dplyr)
library(tidyr)
library(ggplot2)
set.seed(20260205)

# [2] Simulate PSA draws for two strategies vs a common comparator
n_sim <- 5000

psa <- bind_rows(
  tibble(
    strategy = "Immunotherapy",
    dE = pmax(rnorm(n_sim, mean = 0.18, sd = 0.08), 0.001),
    dC = rnorm(n_sim, mean = 8500, sd = 3500)
  ),
  tibble(
    strategy = "Chemotherapy",
    dE = pmax(rnorm(n_sim, mean = 0.08, sd = 0.06), 0.001),
    dC = rnorm(n_sim, mean = 2500, sd = 2500)
  )
)

# [3] WTP grid
lambda_grid <- tibble(lambda = seq(0, 700000, by = 5000))

# [4] Compute CEAC
ceac <- psa %>%
  tidyr::crossing(lambda_grid) %>%
  mutate(nmb = lambda * dE - dC,
         ce = as.integer(nmb > 0)) %>%
  group_by(strategy, lambda) %>%
  summarise(p_ce = mean(ce), .groups = "drop") %>%
  mutate(lambda_k = lambda / 1000)

# [5] CEAC plot
Figure_B10 <- ggplot(ceac, aes(x = lambda_k, y = p_ce,
                               linetype = strategy, color = strategy)) +
  # [1] Color Chemotherapy orange (Immunotherapy set to a professional grey/black)
  scale_color_manual(values = c("Chemotherapy" = "orange",
                                "Immunotherapy" = "steelblue")) +

  geom_smooth(se = FALSE, method = "loess", span = 0.20, linewidth = 1.1) +

  # [2] Vertical line at x = 120
  geom_vline(xintercept = 120, linetype = "dashed", color = "black", alpha = 0.9) +

  # [2] Label in the middle of the line (y = 0.5), adjusted for scale ($120k)
  annotate(
    "text",
    x = 120,
    y = 0.5,
```

```r
    label = "WTP = $120k",
    angle = 0,
    vjust = -0.5,
    size = 3.5,
    fontface = "italic"
  ) +

  scale_y_continuous(limits = c(0, 1), breaks = seq(0, 1, 0.1),
                     labels = function(x) paste0(round(100*x), "%")) +
  labs(
    title = "Cost-Effectiveness Acceptability Curves (CEAC)",
    x = "Willingness to Pay ($/QALY × 1,000)",
    y = "Probability Cost-Effective",
    linetype = "Treatment",
    color = "Treatment"
  ) +
  theme_minimal(base_size = 12) +
  theme(
    legend.position = "right",
    panel.grid.minor = element_blank()
  )

Figure_B10

# [7] Save  (Optional)
ggsave("Figure_B10.png", Figure_B10, width=9, height=6, dpi=300)
ggsave("Figure_B10.pdf", Figure_B10, width=9, height=6,dpi=600,
                                                device = grDevices::cairo_pdf)
```

**Citation:** Soltanifar, M., Portuguese, A.J., Jeon, Y., Gauthier, J., & Lee, C.H. (2026). A North American Collaborative Atlas of Oncology Data Visualization with R Statistical Software. medRxiv preprint. https://doi.org/xx.xxxxx/xxxx.xx.xx.xxxxxxxx, ©2026 Soltanifar, M. et al. (CC BY 4.0/MIT)

**Figure 5.2.11.**

```
# [1] Packages
library(ggplot2)
set.seed(20260206)

#[2] Simulations
# [2-1] Simulated propensity scores (to mimic the provided figure)
# Before matching: mild separation between treatment and control
n0_b <- 800
n1_b <- 800

ps_ctrl_before <- rbeta(n0_b, shape1 = 8,  shape2 = 4)
ps_trt_before  <- rbeta(n1_b, shape1 = 10, shape2 = 8)

before_df <- rbind(
data.frame(stage = "A. Before matching", trt = "Chemotherapy",ps = ps_ctrl_before),
data.frame(stage = "A. Before matching", trt = "Surgury",ps = ps_trt_before)
)

# [2-2] After matching:
#  enforce stronger overlap (trim + re-sample to mimic matched cohorts)
# (Here we mimic matched overlap by drawing both groups from a similar distribution.)
n0_a <- 600
n1_a <- 600

ps_ctrl_after <- rbeta(n0_a, shape1 = 10, shape2 = 8)
ps_trt_after  <- rbeta(n1_a, shape1 = 10, shape2 = 8)

after_df <- rbind(
data.frame(stage = "B. After matching", trt = "Chemotherapy",ps = ps_ctrl_after),
data.frame(stage = "B. After matching", trt = "Surgury", ps = ps_trt_after)
)

df <- rbind(before_df, after_df)

#  [3] Plot (smooth density curves only; no points/markers)
Figure_B11 <- ggplot(df, aes(x = ps, y = after_stat(density),
                             linetype = trt, color = trt)) +
  geom_density(linewidth = 1.0, adjust = 1.1) +
  facet_wrap(~ stage, nrow = 2) +
  labs(
    title = "Propensity Score Density Functions",
    subtitle = "(By Matching Status)",
    x = "Propensity Score",
    y = "Density",
    color = "Treatment",     # Changed from NULL
    linetype = "Treatment"  # Changed from NULL
  ) +
  coord_cartesian(xlim = c(0, 1)) +
  theme_bw() +
  theme(
    legend.position = "right",
```

```r
    panel.grid.minor = element_blank()
  )

Figure_B11

# [4] Save  (Optional)
ggsave("Figure_B11.png", Figure_B11, width=6, height=9, dpi=300)
ggsave("Figure_B11.pdf", Figure_B11, width=6, height=9,dpi=600,
                                               device = grDevices::cairo_pdf)
```

**Citation:** Soltanifar, M., Portuguese, A.J., Jeon, Y., Gauthier, J., & Lee, C.H. (2026). A North American Collaborative Atlas of Oncology Data Visualization with R Statistical Software. medRxiv preprint. https://doi.org/xx.xxxxx/xxxx.xx.xx.xxxxxxxx, ©2026 Soltanifar, M. et al. (CC BY 4.0/MIT)

**Figure 5.2.12.**

```r
# [1] Packages
library(ggplot2)
library(dplyr)
library(sf)
library(rnaturalearth)
library(rnaturalearthdata)
library(ggimage)

# [2] Define Styling
labs <- c("58.2-65.7", "65.8-68.9", "69.0-73.8", "73.9-88.1")
fill_vals <- c(
  "58.2-65.7" = "#dbe9f6",
  "65.8-68.9" = "#a9c8eb",
  "69.0-73.8" = "#5f9fd6",
  "73.9-88.1" = "#0b5fa5",
  "Unavailable Data" = "gold"
)

# [3] Retrieve Geometry
na_map <- ne_states(country = c("canada", "united states of america"),
                    returnclass = "sf")
countries <- ne_countries(country = c("canada", "united states of america"),
                          returnclass = "sf", scale = "medium")

# [4] Simulate Data
set.seed(1203)
sim_dat <- na_map %>%
  mutate(
    bin = sample(labs, size = n(), replace = TRUE),
    bin = case_when(
      name == "Nunavut" ~ "Unavailable Data",
      name == "Arizona" ~ "Unavailable Data",
      TRUE ~ bin
    ),
    bin = factor(bin, levels = c(labs, "Unavailable Data")),
    abbrev = gsub(".*-", "", iso_3166_2)
  )

# [5] Define Capitals
capitals <- data.frame(
  city = c("Ottawa", "Washington"),
  lon = c(-75.6972, -77.0369),
  lat = c(45.4215, 38.9072)
) %>% st_as_sf(coords = c("lon", "lat"), crs = 4326)

# [5b] Define the US-Canada international border as a solid line
# Extract borders between US and Canada using country outlines
# We'll use a spatial intersection approach to get the shared border
us_sf  <- ne_countries(country = "united states of america",
                       returnclass = "sf", scale = "medium")
can_sf <- ne_countries(country = "canada",
```

```
                        returnclass = "sf", scale = "medium")

# Get the shared border line (intersection of boundaries)
us_boundary  <- st_cast(st_union(us_sf),  "MULTILINESTRING")
can_boundary <- st_cast(st_union(can_sf), "MULTILINESTRING")
intl_border  <- st_intersection(us_boundary, can_boundary)

# [5c] Flag positions in lon/lat - left of California and left of British Columbia
# California's western edge ~-124.5 lon, ~37 lat  → place flag further west
# British Columbia's western edge ~-139 lon, ~56 lat → place flag further west
flags <- data.frame(
  country = c("Canada", "USA"),
  lon     = c(-145, -133),   # left of BC (~-139) and left of CA (~-124)
  lat     = c(53,    36),
  image   = c(
    "https://flagcdn.com/w160/ca.png",
    "https://flagcdn.com/w160/us.png"
  )
)

# Project flag coordinates to match the Albers CRS used in coord_sf
aea_crs <- "+proj=aea +lat_1=20 +lat_2=60 +lat_0=40 +
                        lon_0=-96 +x_0=0 +y_0=0 +datum=NAD83 +units=m +no_defs"

flags_sf <- st_as_sf(flags, coords = c("lon", "lat"), crs = 4326) %>%
  st_transform(crs = aea_crs)

flags$x_proj <- st_coordinates(flags_sf)[, 1]
flags$y_proj <- st_coordinates(flags_sf)[, 2]

# [6] Final Plot
Figure_B12 <- ggplot(data = sim_dat) +

  # Base Map (state/province borders dashed)
  geom_sf(aes(fill = bin), color = "black", linetype = "dashed", linewidth = 0.2) +

  # Outer country coastlines (solid)
  geom_sf(data = countries, fill = NA, color = "black", linewidth = 0.6) +

  # International US-Canada border (solid, slightly thicker)
  geom_sf(data = intl_border, color = "black", linewidth = 0.8, linetype = "solid") +

  # State/Province Abbreviations
  geom_sf_text(aes(label = abbrev), size = 2.0, color = "black",
             check_overlap = TRUE, alpha = 1) +

  # Capital cities with stars
  geom_sf(data = capitals, shape = 15, color = "red", size = 1.0, stroke = 1.5) +
  geom_sf_text(data = capitals, aes(label = city),
             vjust = -1.5, fontface = "bold", size = 1.5, color="red") +

  # Flags using projected coordinates
  geom_image(data = flags, aes(x = x_proj, y = y_proj, image = image),
```

```r
            size = 0.1, inherit.aes = FALSE) +

  # Projection and Theme
  coord_sf(crs = aea_crs) +
  scale_fill_manual(values = fill_vals, drop = FALSE, name = "Incidence (per 1M)") +
  labs(
    title    = "Geographic Variation of North American Cancer Incidence",
    subtitle = "Simulated Time Period: 2001-2026"
  ) +
  theme_void() +
  theme(
    plot.title    = element_text(face = "bold", hjust = 0.5, size = 14),
    plot.subtitle = element_text(hjust = 0.5),
    legend.position = "right",
    plot.margin   = margin(10, 10, 10, 10)
  )

Figure_B12


# [6] Save  (Optional)
ggsave("Figure_B12.png", Figure_B12, width=8, height=12, dpi=300)
ggsave("Figure_B12.pdf", Figure_B12, width=8, height=12,dpi=600,
                                                 device = grDevices::cairo_pdf)
```

**Citation:** Soltanifar, M., Portuguese, A.J., Jeon, Y., Gauthier, J., & Lee, C.H. (2026). A North American Collaborative Atlas of Oncology Data Visualization with R Statistical Software. medRxiv preprint. https://doi.org/xx.xxxxx/xxxx.xx.xx.xxxxxxxx, ©2026 Soltanifar, M. et al. (CC BY 4.0/MIT)

#### Section C. Common CT and RWE

**Figure 5.3.1.**

```
# [1] Packages
library(meta)
library(dplyr)
library(ggplot2)
library(cowplot)
library(ggplotify)
library(cowplot)
set.seed(20260214)

# [2] Simulate Data
k <- 12
n_i <- sample(seq(60, 250, by = 10), size = k, replace = TRUE)
mu_logit <- -0.8
tau      <- 0.25
theta_i  <- rnorm(k, mean = mu_logit, sd = tau)
x_i      <- rbinom(k, size = n_i, prob = plogis(theta_i))

dat <- tibble(
  study = sprintf("Study %02d", 1:k),
  events = x_i,
  total = n_i
)

# [3] Create the Meta-analysis Object
m.cont_FEM <- metaprop(event = events,
                       n = total,
                       studlab = study,
                       data = dat,
                       method = "GLMM",
                       sm = "PLOGIT")

# [3] CAPTURE PLOTS
# 1. Convert the Forest Plot to a ggplot object
# We wrap it in a function to ensure it captures correctly
gp01a <- as.ggplot(expression(
  forest(m.cont_FEM,
         sortvar = TE,
         prediction = TRUE,
         print.tau2 = FALSE,
         leftlabs = c("A. Forest Plot (JAMA layout)\nStudy ID", "Proportion (95%CI)"),
         layout = "JAMA")
))

# 2. Convert the Funnel Plot to a ggplot object
gp01b <- as.ggplot(expression({
  colcontour <- c("green", "palegoldenrod", "yellow", "orange")
  funnel(m.cont_FEM,
         xlim = c(-1.5, 1),
         contour = c(0.80, 0.90, 0.95, 0.99),
```

```r
         col.contour = colcontour)
  legend(x = 0.35, y = 0.05,
         legend = c("p < 0.20", "p < 0.10", "p < 0.05", "p < 0.01"),
         fill = colcontour)
  mtext("B. Contour-Enhanced Funnel Plot",
        side = 3,    # Top of the plot
        line = 1.5,  # Distance above the plot area
        adj = 0,     # 0 = Left, 0.5 = Center, 1 = Right
        font = 2,    # 2 = Bold
        cex = 1.0)   # Font size multiplier
}))

# 3. COMBINE AND ADJUST
# Strip margins from the captured ggplot objects
gp01a_clean <- gp01a + theme(plot.margin = margin(t = 0, r = 0, b = 0, l = 0))
gp01b_clean <- gp01b + theme(plot.margin = margin(t = -30, r = 0, b = 0, l = 0))
                                             # Negative top margin pulls it up

# Combine with relative heights
Figure_C01 <- plot_grid(
  gp01a_clean,
  gp01b_clean,
  ncol = 1,
  rel_heights = c(2, 1), # Makes Panel A significantly taller than Panel B
  align = 'v',           # Ensures vertical alignment
  axis = 'lr'            # Aligns left and right axes
)

Figure_C01

# [4] Save (Optional)

save_plot("Figure_C01.png", Figure_C01, base_width = 11, base_height = 11)
save_plot("Figure_C01.pdf", Figure_C01, base_width = 11, base_height = 11)
```

**Citation:** Soltanifar, M., Portuguese, A.J., Jeon, Y., Gauthier, J., & Lee, C.H. (2026). A North American Collaborative Atlas of Oncology Data Visualization with R Statistical Software. medRxiv preprint. https://doi.org/xx.xxxxx/xxxx.xx.xx.xxxxxxxx, ©2026 Soltanifar, M. et al. (CC BY 4.0/MIT)

**Figure 5.3.2.**

```
# [1] Packages
library(DiagrammeR)
library(DiagrammeRsvg)
library(rsvg)
library(magick)

#[2] Panels
#[2-1] Panel A: CONSORT (Two-arm RCT)
consort_A <- grViz("
digraph consortA {
  graph [rankdir = TB, bgcolor = white,
         label = 'A. CONSORT Diagram for Clinical Trials',
         labelloc = t, fontsize = 14, fontname = 'Helvetica-Bold']

  node  [shape = box, style = 'rounded', fontname = Helvetica, fontsize = 10]
  edge  [color = black, arrowsize = 0.7]

  S   [label = 'Assessed for eligibility\\n(n = 3428)']
  E   [label = 'Excluded\\n(n = 2154)\\n- Not meeting eligibility: 2153\\n- Other: 1']
  R   [label = 'Randomized / ITT population\\n(n = 1274)']

  A1  [label = 'Allocated to Pembrolizumab combination\\n(n = 637)']
  A2  [label = 'Allocated to Chemotherapy combination\\n(n = 637)']

  F1  [label = 'Follow-up (summary)\\nDiscontinued: 507\\nCompleted: 42\
                                                          \nOngoing/other: 88']
  F2  [label = 'Follow-up (summary)\\nDiscontinued: 425\\nCompleted: 160\
                                                          \nOngoing/other: 52']

  AN1 [label = 'Analysis (ITT)\\n(n = 637)']
  AN2 [label = 'Analysis (ITT)\\n(n = 637)']

  S -> E
  S -> R
  R -> A1
  R -> A2
  A1 -> F1
  A2 -> F2
  F1 -> AN1
  F2 -> AN2

  {rank = same; A1; A2}
  {rank = same; F1; F2}
  {rank = same; AN1; AN2}
}
")

#[2-2] Panel B: STROBE-style (One-arm RWE)
strobe_B <- grViz("
digraph strobeB {
  graph [rankdir = TB, bgcolor = white,
```

```
        label = 'B. STROBE Diagram for RWE Studies',
        labelloc = t, fontsize = 14, fontname = 'Helvetica-Bold']

  node  [shape = box, style = 'rounded', fontname = Helvetica, fontsize = 10]
  edge  [color = black, arrowsize = 0.7]

  D0 [label = 'Data source pull (EHR/registry)\\n(n = 3428)']
  X1 [label = 'Excluded\\n(n = 2154)\\n- Not meeting study criteria: 2153\\n- Other: 1']
  C1 [label = 'Eligible cohort\\n(n = 1274)']

  X2 [label = 'Excluded (data quality)\\n(n = 76)\\n- Missing key baseline covariates\
                                                    \n- No follow-up information']
  C2 [label = 'Final analytic RWE cohort (one-arm)\\n(n = 1198)']

  O  [label = 'Outcome ascertainment\\nDeaths: 592\\nCensored: 606']
  A  [label = 'Analysis set\\n(n = 1198)\
                     \nAdjusted analysis (e.g., weighting/matching as needed)']

  D0 -> X1
  D0 -> C1
  C1 -> X2
  C1 -> C2
  C2 -> O
  C2 -> A
}
")

#[3] Export and Merge at 600 DPI
# Convert Diagrams to SVG strings
 svg_A_str <- export_svg(consort_A)
 svg_B_str <- export_svg(strobe_B)

# Write individual high-res temporary PNGs (used for merging)
 rsvg_png(charToRaw(svg_A_str), "temp_A.png")
 rsvg_png(charToRaw(svg_B_str), "temp_B.png")

# Read into Magick
imgA <- image_read("temp_A.png")
imgB <- image_read("temp_B.png")

# Append
# Create a white spacer (match the width of your plots, e.g., 2000px)
spacer <- image_blank(width = 100, height = 40, color = "white")
Figure_C02 <- image_append(c(imgA,spacer, imgB), stack = TRUE)

#[4] Save (Optional)
image_write(Figure_C02, path = "Figure_C02.png", format = "png", density = 600)
image_write(Figure_C02, path = "Figure_C02.pdf", format = "pdf", density = 100)

# Cleanup temporary files
file.remove(c("temp_A.png", "temp_B.png"))
```

**Figure 5.3.3.**

```r
#[1] Packages
library(ggplot2)

#[2] Data Simulations
mu    <- 0.50
sigma <- 0.35
c_cut <- 0.50

a01 <- -1.10; a11 <-  2.80   # T1
a02 <- -0.70; a12 <-  3.10   # T2

x_grid <- seq(-0.5, 1.5, length.out = 400)

df <- rbind(
  data.frame(
    x = x_grid,
    Treatment = "T1",
    p = plogis(a01 + a11 * x_grid)
  ),
  data.frame(
    x = x_grid,
    Treatment = "T2",
    p = plogis(a02 + a12 * x_grid)
  )
)

v_Mplus <- 1 - pnorm(c_cut, mean = mu, sd = sigma)

# [3] Plot
Figure_C03 <- ggplot(df, aes(x = x, y = p, color = Treatment, linetype = Treatment)) +
  geom_line(linewidth = 1.2) +
  geom_vline(xintercept = c_cut, linetype = "dashed", linewidth = 0.8) +
  annotate("text", x = c_cut, y = 0.05, label = "Cut-off",
           vjust = 0, hjust = -0.1, fontface = "italic") +

  # Professional Color Palette (e.g., ColorBrewer or viridis-style)
  scale_color_manual(values = c("T1" = "blue", "T2" = "brown")) +

  # Distinct Linetypes: Solid for primary, Dashed for comparison
  scale_linetype_manual(values = c("T1" = "solid", "T2" = "dotdash")) +

  scale_y_continuous(limits = c(0, 1), breaks = seq(0, 1, 0.2)) +
  scale_x_continuous(limits = c(-0.5, 1.5), breaks = seq(-0.5, 1.5, 0.5)) +
  labs(
    title = "Marker value-Response distribution (Scenario-style)",
    subtitle = sprintf("Cut-off c = %.2f;
                       v_{M+}(c) approx %.2f under X ~ N(%.2f, %.2f^2)",
                       c_cut, v_Mplus, mu, sigma),
    x = "Marker Value (log-transformed)",
    y = "Response Probability",
    color = "Treatment",
```

```r
    linetype = "Treatment" # Keeps both aesthetics in a single merged legend
  ) +
  theme_minimal(base_size = 12) +
  theme(
    legend.position = "right",
    panel.grid.minor = element_blank(), # Clean up background
    axis.line = element_line(color = "black") # Add axis lines for structure
  )


Figure_C03

#[4] Save (Optional)
ggsave("Figure_C03.png", Figure_C03, width=9, height=6, dpi=600)
ggsave("Figure_C03.pdf", Figure_C03, width=9, height=6, dpi=600,
                                    device = grDevices::cairo_pdf)
```

**Figure 5.3.4.**

```r
#[1] Packages
library(ggplot2)
library(dplyr)

#  [2] Simulation
#[2-1] (A) Construct smooth, simple modeled curves (no points)
RS <- seq(0, 30, by = 0.1)

#[2-2] Choose two monotone increasing curves that cross near RS ~ 20 (equivalence).
# (Simple quadratic forms give the gentle curvature seen in the reference figure.)
logHR_noChemo <- 0.80 + 0.030 * RS + 0.0002 * RS^2
logHR_chemo   <- 0.02 + 0.065 * RS + 0.0009 * RS^2

df <- bind_rows(
  data.frame(RS = RS, logHR = logHR_noChemo, Therapy = "No chemotherapy"),
  data.frame(RS = RS, logHR = logHR_chemo,   Therapy = "Chemotherapy")
)

#[2-3]  (B) Key reference vertical lines
RS_equiv_l <- 10  # long-dashed: lower bound of 95% CI for the equivalence point
RS_equiv<- 16.5 # short-dashed: RS where treatments are equivalent (modeled intersection)
RS_equiv_u <- 24 # long-dashed: upper bound of 95% CI for the equivalence point

# [3]  Plot
Figure_C04 <- ggplot(df, aes(x = RS, y = logHR, color = Therapy)) +
  geom_line(linewidth = 1.2) +
  # Short dashed line = lower 95% CI bound for equivalence RS
  geom_vline(xintercept = RS_equiv_l, linetype = "dashed",
             linewidth = 0.9, color = "grey30") +

  # Short dashed line = equivalence RS
  geom_vline(xintercept = RS_equiv, linetype = "dashed",
             linewidth = 0.9, color = "grey30") +

  # Long dashed line = upper 95% CI bound for equivalence RS
  geom_vline(xintercept = RS_equiv_u, linetype = "longdash",
             linewidth = 0.9, color = "grey30") +

  # Annotations (text only; no points/markers)
  annotate("text", x = RS_equiv, y = 0.95,
           label = "Equivalence", hjust = 0.5, vjust = 0) +
  annotate("text", x = 9.5,  y = 0.55,
           label = "Chemotherapy\ninferior", hjust = 0.5) +
  annotate("text", x = 26.0, y = 0.55,
           label = "Chemotherapy\nsuperior", hjust = 0.5) +
  annotate("text", x = 24.5, y = 1.60,
           label = "HR = c", hjust = 0) +

  scale_x_continuous(limits = c(0, 30), breaks = seq(0, 30, 5)) +
  scale_y_continuous(limits = c(0, 2.5), breaks = seq(0, 2.5, 0.5)) +
```

```r
  scale_color_manual(values = c("No chemotherapy" = "#1F77B4",
                               "Chemotherapy" = "#D62728")) +

  labs(
    title = "Recurrence Score-Log(HR) Curve",
    x = "Recurrence Score(RS)",
    y = "Log(HR)",
    color = "Treatment"
  ) +

  theme_minimal(base_size = 12) +
  theme(
    legend.position = "right",
    panel.grid.minor = element_blank()
  )


Figure_C04

#[4] Save (Optional)
ggsave("Figure_C04.png", Figure_C04, width=9, height=6, dpi=600)
ggsave("Figure_C04.pdf", Figure_C04, width=9, height=6, dpi=600,
                                                   device = grDevices::cairo_pdf)
```

**Citation:** Soltanifar, M., Portuguese, A.J., Jeon, Y., Gauthier, J., & Lee, C.H. (2026). A North American Collaborative Atlas of Oncology Data Visualization with R Statistical Software. medRxiv preprint. https://doi.org/xx.xxxxx/xxxx.xx.xx.xxxxxxxx, ©2026 Soltanifar, M. et al. (CC BY 4.0/MIT)

**Figure 5.3.5.**

```r
# [1] Packages
library(ggplot2)
library(dplyr)
library(tidyr)
set.seed(2026)

#[2] Simulation
# [2-1] Simulated "utility over time" (0=death, 1=perfect health)
# Time grid (months)
t_grid <- seq(0, 36, by = 1)

# [2-2] Death times (months): Trt 1 earlier death, Trt 2 later death
death_t1 <- 24
death_t2 <- 30

# [2-3] Helper: smooth-ish decline with a couple of transient dips
                                 #(treatment toxicity / progression shocks)
utility_profile <- function(t, base = 0.92, slope = 0.006,
                            dips = list(), death_time = 999) {
  u <- base - slope * t
  # Add transient dips (each: center, width, depth)
  for (d in dips) {
    u <- u - d$depth * exp(-0.5 * ((t - d$center) / d$width)^2)
  }
  # Enforce death at/after death_time
  u[t >= death_time] <- 0
  # Clamp to [0,1] for this example (your instruments may allow <0)
  pmin(pmax(u, 0), 1)
}

df <- tibble(
  time = rep(t_grid, times = 2),
  treatment = rep(c("Treatment 1", "Treatment 2"), each = length(t_grid))
) %>%
  mutate(
    utility = case_when(
      treatment == "Treatment 1" ~ utility_profile(
        time,
        base = 0.90, slope = 0.010,
        dips = list(
          list(center = 6,  width = 2.0, depth = 0.12),
          list(center = 14, width = 2.4, depth = 0.20)
        ),
        death_time = death_t1
      ),
      treatment == "Treatment 2" ~ utility_profile(
        time,
        base = 0.92, slope = 0.006,
        dips = list(
          list(center = 8,  width = 2.2, depth = 0.08),
          list(center = 18, width = 2.8, depth = 0.10)
```

```r
      ),
      death_time = death_t2
    )
  )
)

# [2-4] Wide format for ribbon (QALYs gained = area between curves where Trt2 > Trt1)
df_w <- df %>%
  select(time, treatment, utility) %>%
  pivot_wider(names_from = treatment, values_from = utility) %>%
  mutate(
    ymin = pmin(`Treatment 1`, `Treatment 2`),
    ymax = pmax(`Treatment 1`, `Treatment 2`)
  )

# [2-5] Approximate QALYs (area under curve) by trapezoidal rule (time in months -> years)
trapz <- function(x, y) sum(diff(x) * (head(y, -1) + tail(y, -1)) / 2)

qaly_t1 <- trapz(df_w$time / 12, df_w$`Treatment 1`)
qaly_t2 <- trapz(df_w$time / 12, df_w$`Treatment 2`)
qaly_gain <- qaly_t2 - qaly_t1

# [3] Plot (no point markers).
Figure_C05 <- ggplot() +
  # Shaded area between curves = QALYs gained/lost over the horizon (no markers added)
  geom_ribbon(
    data = df_w,
    aes(x = time, ymin = ymin, ymax = ymax),
    alpha = 0.20
  ) +
  geom_step(
    data = df,
    aes(x = time, y = utility, linetype = treatment, color = treatment),
    linewidth = 1.25
  ) +
  geom_vline(xintercept = death_t1, linetype = "dashed", linewidth = 0.6) +
  geom_vline(xintercept = death_t2, linetype = "dashed", linewidth = 0.6) +
  annotate("text", x = death_t1, y = 0.12, label = "Death (Treatment 1)",
           hjust = -0.05, vjust = 0) +
  annotate("text", x = death_t2, y = 0.02, label = "Death (Treatment 2)",
           hjust = -0.05, vjust = 0) +
  annotate(
    "text",
    x = 18, y = 0.85,
    label = paste0("Delta*italic(QALYs)==+ ", round(qaly_gain, 2)),
    parse = TRUE,
    fontface = "bold"
  ) +
  scale_x_continuous(breaks = seq(0, 36, by = 6)) +
  scale_y_continuous(limits = c(0, 1), breaks = seq(0, 1, by = 0.2)) +
  labs(
    x = "Time (Months)",
    y = "Quality of Life (Utility)",
```

```r
    color = "Treatment",
    linetype = "Treatment",
    title = "Time-Quality of Life (Utility) Curves" ) +
  theme_minimal(base_size = 12) +
  theme(legend.position = "right")

Figure_C05

#[4] Save (Optional)
ggsave("Figure_C05.png", Figure_C05, width=10.5, height=7, dpi=600)
ggsave("Figure_C05.pdf", Figure_C05, width=10.5, height=7, dpi=600,
                                              device = grDevices::cairo_pdf)
```

**Citation:** Soltanifar, M., Portuguese, A.J., Jeon, Y., Gauthier, J., & Lee, C.H. (2026). A North American Collaborative Atlas of Oncology Data Visualization with R Statistical Software. medRxiv preprint. https://doi.org/xx.xxxxx/xxxx.xx.xx.xxxxxxxx, ©2026 Soltanifar, M. et al. (CC BY 4.0/MIT)

**Figure 5.3.6.**

```r
#[1] Packages
library(ggplot2)
library(dplyr)
library(tidyr)
library(scales)

set.seed(20260212)

#[2] Simulations
t_max <- 13000
dt    <- 10
time_grid <- seq(0, t_max, by = dt)

cycle_starts <- seq(2000, 10000, by = 2000)
cycle_width  <- 250
cycle_ends   <- cycle_starts + cycle_width

in_cycle <- function(t) {
  any(t >= cycle_starts & t <= cycle_ends)
}

simulate_arm <- function(arm_label,
                         N0 = 1e3,
                         r_grow = 0.00055,
                         kill_fraction = 0.90,
                         wound_healing_mult = 1.00
) {
  N <- numeric(length(time_grid))
  N[1] <- N0

  for (i in 2:length(time_grid)) {
    t_prev <- time_grid[i - 1]
    t_curr <- time_grid[i]
    r_eff <- r_grow
    if (!in_cycle(t_prev)) r_eff <- r_eff * wound_healing_mult
    N[i] <- N[i - 1] * exp(r_eff * (t_curr - t_prev))
    if (any(t_prev < cycle_ends & t_curr >= cycle_ends)) {
      N[i] <- N[i] * (1 - kill_fraction)
    }
  }

  tibble(
    time = time_grid,
    tumor_cells = N,
    arm = arm_label
  )
}

#[2-4] Build arms
df_untreated <- tibble(
  time = time_grid,
```

```
  tumor_cells = 1e3 * exp(0.00065 * time_grid),
  arm = "Untreated"
)

df_chemo_no_wh <- simulate_arm(
  arm_label = "Chemotherapy (no wound-healing)",
  N0 = 1e3,
  r_grow = 0.00055,
  kill_fraction = 0.92,
  wound_healing_mult = 1.00
)

df_chemo_wh <- simulate_arm(
  arm_label = "Chemotherapy (wound-healing)",
  N0 = 1e3,
  r_grow = 0.00055,
  kill_fraction = 0.92,
  wound_healing_mult = 1.35
)

df <- bind_rows(df_untreated, df_chemo_no_wh, df_chemo_wh)

shade <- tibble(xmin = cycle_starts, xmax = cycle_ends)

#[3] Plot
Figure_C06 <- ggplot() +
  geom_point(
    data = subset(df, round(time) %% 1000 == 0),
    aes(x = time, y = tumor_cells, color = arm)
  )+
  geom_rect(
    data = shade,
    aes(xmin = xmin, xmax = xmax, ymin = -Inf, ymax = Inf),
    inherit.aes = FALSE,
    alpha = 0.12
  ) +
  geom_line(
    data = df,
    aes(x = time, y = tumor_cells, color = arm),
    linewidth = 1.1
  ) +
  geom_hline(yintercept = 5, linetype = "dashed", linewidth = 1) +
  annotate("text", x = 2000, y =6, label = "Threshold for Eradication",
           vjust = +0.05, vjust = 0)+
  # [3] Y-axis power of 10 formatting
  scale_y_log10(
    name = "Total Tumor Cells (log scale)",
    label = label_log(10)
  ) +
  # [4] X-axis ticks every 1000
  scale_x_continuous(
    breaks = seq(0, t_max, by = 1000)
  ) +
```

```r
  # [1] & [2] Color updates
  scale_color_manual(
    values = c(
      "Untreated" = "red",
      "Chemotherapy (no wound-healing)" = "blue",
      "Chemotherapy (wound-healing)" = "darkgreen"
      # Keep/add a third color for the 3rd arm
    )
  ) +
  labs(
    x = "Time (Hours)",
    color = "Treatment"
  ) +
  theme_classic(base_size = 12) +
  theme(
    legend.position = "bottom",
    legend.title = element_text(face = "bold")
  )

Figure_C06

#[4] Save (Optional)
ggsave("Figure_C06.png", Figure_C06, width=10.5, height=7, dpi=600)
ggsave("Figure_C06.pdf", Figure_C06, width=10.5, height=7, dpi=600,
                                                device = grDevices::cairo_pdf)
```

**Figure 5.3.7.**

```
# [1] Packages
library(rpart)
library(rpart.plot)
set.seed(20260212)

#[2] Simulations
#[2-1] parameters
n <- 686
dat <- data.frame(
  nodes   = pmin(rpois(n, lambda = 6), 30),
  progrez = pmax(round(rnorm(n, mean = 60, sd = 35)), 0),
  estrez  = pmax(round(rnorm(n, mean = 120, sd = 80)), 0),
  tsize   = pmax(round(rnorm(n, mean = 25, sd = 10)), 5),
  age     = pmin(pmax(round(rnorm(n, mean = 52, sd = 10)), 25), 80)
)

# [2-2] Strong signal for branching
linpred <- -3.8 + 2.2*(dat$nodes > 8) + 1.8*(dat$progrez < 30) + 1.1*(dat$tsize > 25)
p_event <- 1 / (1 + exp(-linpred))
dat$event5y <- factor(ifelse(runif(n) < p_event, "Event", "NoEvent"),
                      levels = c("NoEvent", "Event"))

# [2-3] Fit and Prune
fit <- rpart(event5y ~ nodes + progrez + tsize + age + estrez,
             data = dat, method = "class",
             control = rpart.control(minbucket = 10, cp = 0.001))
fit_pruned <- prune(fit, cp = 0.015)

# [3] Customizing Node Labels
# We calculate the (Total N, Total %) pair for each node
node_info <- fit_pruned$frame
node_n <- node_info$n
node_pct <- round(100 * node_n / n)
node_labels <- paste0("(", node_n, ", ", node_pct, "%)")

# We create a custom function to pass to rpart.plot
# This overrides the default text to show your requested pair
my_node_fun <- function(x, labs, digits, varlen) {
  # labs contains the Predicted Class (e.g., "NoEvent")
  # we append the custom (N, %) pair and the split counts
  # Note: fit_pruned$frame$yval2 contains the class counts
  counts <- fit_pruned$frame$yval2[, 2:3] # Columns for NoEvent and Event counts

  paste0(labs, "\n",
         node_labels, "\n",
         counts[,1], " ", counts[,2])
}

# [4] Final Plot
op <- par(mar = c(2, 2, 2, 8), xpd = NA)
```

```r
rpart.plot(fit_pruned, type= 2,   extra  = 0, node.fun = my_node_fun,
  box.palette = list("Blues", "Reds"),fallen.leaves = TRUE,
  shadow.col = "gray90",  nn = FALSE)

legend("topright",inset  = c(-0.25, 0),title  = "Predicted class",
  legend = c("NoEvent", "Event"), fill   = c("#6baed6", "#fb6a4a"),
  border = NA,bty= "n")

par(op)

#[4] Save (Optional):

# [4-1] Save as PNG (600 dpi)
png(filename = "Figure_C07.png", width = 8.5, height = 5.5, units = "in", res = 600)

op <- par(mar = c(2, 2, 2, 8), xpd = NA)

rpart.plot(fit_pruned, type= 2,   extra  = 0, node.fun = my_node_fun,
           box.palette = list("Blues", "Reds"),fallen.leaves = TRUE,
           shadow.col = "gray90", nn = FALSE)

legend("topright",inset  = c(-0.25, 0),title  = "Predicted class",
       legend = c("NoEvent", "Event"), fill   = c("#6baed6", "#fb6a4a"),
       border = NA,bty= "n")

par(op)
dev.off()


# [4-2] Save as PDF
pdf(file = "Figure_C07.pdf",
    width = 8.5, height = 5.5, onefile = TRUE)

op <- par(mar = c(2, 2, 2, 8), xpd = NA)

rpart.plot(fit_pruned, type= 2,   extra  = 0, node.fun = my_node_fun,
           box.palette = list("Blues", "Reds"),fallen.leaves = TRUE,
           shadow.col = "gray90", nn = FALSE)

legend("topright",inset  = c(-0.25, 0),title  = "Predicted class",
       legend = c("NoEvent", "Event"), fill   = c("#6baed6", "#fb6a4a"),
       border = NA,bty= "n")

par(op)
dev.off()
```

**Citation:** Soltanifar, M., Portuguese, A.J., Jeon, Y., Gauthier, J., & Lee, C.H. (2026). A North American Collaborative Atlas of Oncology Data Visualization with R Statistical Software. medRxiv preprint. https://doi.org/xx.xxxxx/xxxx.xx.xx.xxxxxxxx, ©2026 Soltanifar, M. et al. (CC BY 4.0/MIT)

**Figure 5.3.8.**

```
#[1] Packages
library(ggplot2)
library(dplyr)
set.seed(25)

#[2] Simulations
# [2-1] Time grid (in years)
time_grid <- seq(0, 6, by = 0.1)

# [2-2] Helper: create a monotone, step-like prediction-error curve
make_step_curve <- function(t, start = 0.00, plateau = 0.26, speed = 1.2,
                            step = 0.01, noise_sd = 0.003) {
  # Smooth baseline shape (rises and then plateaus)
  base <- start + (plateau - start) * (1 - exp(-speed * t))
  # Add small noise then enforce monotonicity
  y <- base + rnorm(length(t), mean = 0, sd = noise_sd)
  y <- cummax(y)
  # Quantize to create visible steps (like empirical curves)
  y <- round(y / step) * step
  # Keep inside plotting window
  y <- pmin(pmax(y, 0), 0.30)
  y
}

# [2-3] Simulate four methods, with Kaplan-Meier having the highest error
df <- bind_rows(
  tibble(
    time = time_grid,
    method = "Kaplan-Meier",
    pred_error = make_step_curve(time_grid, start = 0.00, plateau = 0.27,
                                 speed = 1.05, step = 0.01)
  ),
  tibble(
    time = time_grid,
    method = "Cox (PI)",
    pred_error = make_step_curve(time_grid, start = 0.00, plateau = 0.245,
                                 speed = 1.10, step = 0.01)
  ),
  tibble(
    time = time_grid,
    method = "NPI",
    pred_error = make_step_curve(time_grid, start = 0.00, plateau = 0.235,
                                 speed = 1.15, step = 0.01)
  ),
  tibble(
    time = time_grid,
    method = "CART",
    pred_error = make_step_curve(time_grid, start = 0.00, plateau = 0.225,
                                 speed = 1.20, step = 0.01)
  )
) %>%
```

```r
  mutate(method = factor(method, levels = c("Kaplan-Meier", "Cox (PI)",
                                            "NPI", "CART")))
# [3] Plot
Figure_C08 <- ggplot(df, aes(x = time, y = pred_error, color = method)) +
  geom_step(linewidth = 0.9) +
  scale_x_continuous(limits = c(0, 6), breaks = 0:6) +
  scale_y_continuous(limits = c(0, 0.30), breaks = seq(0, 0.30, by = 0.05)) +
  labs(
    x = "Time (Years)",
    y = "Prediction Error",
    color = "Classification Method"
  ) +
  theme_minimal(base_size = 12) +
  theme(
    legend.position = "right",
    legend.title = element_text(size = 11),
    legend.text  = element_text(size = 10)
  )

Figure_C08


#[4] Save (Optional)
ggsave("Figure_C08.png", Figure_C08, width=10.5, height=7, dpi=600)
ggsave("Figure_C08.pdf", Figure_C08, width=10.5, height=7, dpi=600,
                                                device = grDevices::cairo_pdf)
```

**Citation:** Soltanifar, M., Portuguese, A.J., Jeon, Y., Gauthier, J., & Lee, C.H. (2026). A North American Collaborative Atlas of Oncology Data Visualization with R Statistical Software. medRxiv preprint. https://doi.org/xx.xxxxx/xxxx.xx.xx.xxxxxxxx, ©2026 Soltanifar, M. et al. (CC BY 4.0/MIT)

**Figure 5.3.9.**

```
#[1] Packages
library(ggplot2)
library(dplyr)
set.seed(909)

#[2] Simulations
time_wk <- seq(0, 54, by = 6)

sim_traj <- function(time_wk, pattern = c("CR","PR","SD","PD","NL")) {
  pattern <- match.arg(pattern)
  n <- length(time_wk)

  if (pattern == "CR") {
    y <- -100 * (1 - exp(-0.12 * time_wk)) + rnorm(n, 0, 4)
    y <- pmax(y, -100)
  } else if (pattern == "PR") {
    target <- runif(1, -80, -35)
    y <- target * (1 - exp(-0.10 * time_wk)) + rnorm(n, 0, 6)
  } else if (pattern == "SD") {
    drift <- runif(1, -10, 10)
    y <- drift + cumsum(rnorm(n, 0, 5))
    y <- pmin(pmax(y, -30), 30)
  } else if (pattern == "PD") {
    slope <- runif(1, 0.8, 2.2)
    y <- slope * time_wk + rnorm(n, 0, 8)
  } else {
    target <- runif(1, -70, -20)
    y <- target * (1 - exp(-0.08 * time_wk)) + rnorm(n, 0, 7)
  }

  y[1] <- 0
  y <- pmin(pmax(y, -110), 140)
  y
}

patient_tbl <- tibble(
  patient_id = factor(paste0("Patient ", 1:10), levels = paste0("Patient ", 1:10)),
  disease_status = factor(
    c("Complete Response", "Partial Response", "Partial Response",
      "Stable Disease", "Stable Disease",
      "Progression", "Progression",
      "New Lesion", "New Lesion", "Stable Disease"),
    levels = c("Complete Response","Partial Response","Stable Disease",
               "Progression", "New Lesion")
  ),
  pattern = c("CR","PR","PR","SD","SD","PD","PD","NL","NL","SD")
)

df <- patient_tbl %>%
  rowwise() %>%
  do(tibble(
```

```
    patient_id = .$patient_id,
    disease_status = .$disease_status,
    time_wk = time_wk,
    pct_change = sim_traj(time_wk, pattern = .$pattern)
  )) %>%
  ungroup()

#[3] Updated Plot
Figure_C09 <- ggplot(df, aes(x = time_wk, y = pct_change,
                         group = patient_id,
                         color = patient_id,
                         shape = disease_status)) + # Map shape here
  geom_line(linewidth = 1, linetype = "solid") +
  geom_point(size = 2) +
  geom_hline(yintercept =  20, linewidth = 0.8, linetype = "dashed", color = "black") +
  geom_hline(yintercept = -30, linewidth = 0.8, linetype = "dashed", color = "black") +
  scale_x_continuous(breaks = seq(0, 54, by = 6)) +
  scale_y_continuous(limits = c(-110, 140), breaks = seq(-100, 140, by = 20)) +
  scale_color_discrete() +
  labs(
    title = "Tumor Response Over Time (Spider Plot)",
    x = "Time (weeks)",
    y = "Change in Target Lesion from Baseline (%)",
    color = "Patient ID",
    shape = "Disease Status" # This will now work correctly
  ) +
  theme_minimal(base_size = 12) +
  theme(
    legend.position = "right",
    legend.box = "vertical"
  )

Figure_C09

#[4] Save (Optional)
ggsave("Figure_C09.png", Figure_C09, width=10.5, height=7, dpi=600)
ggsave("Figure_C09.pdf", Figure_C09, width=10.5, height=7, dpi=600,
                                                device = grDevices::cairo_pdf)
```

**Figure 5.3.10.**

```
# [1] Packages
library(cmprsk)
library(ggplot2)
set.seed(10)

# [2] Simulations (Adding a 'group' variable)
n <- 500
t_max <- 10
# Creating two groups (250 each)
group <- rep(c("Cancer death", "Cardiovascular death"), each = n/2)

# Adjusting hazards slightly for Group B=Cardiovascular death to ensure a non-zero p-value
t1 <- rexp(n, rate = ifelse(group == "Cancer death", 0.18, 0.25))
t2 <- rexp(n, rate = 0.08)
cens <- runif(n, min = 0, max = t_max)

time <- pmin(t1, t2, cens)
status <- ifelse(time == cens, 0, ifelse(t1 < t2, 1, 2))

dat <- data.frame(time = time, status = status, group = group)

# [2-3] CIF estimation with Grouping
# Gray's test is calculated automatically here
ci <- cmprsk::cuminc(ftime = dat$time, fstatus = dat$status,
                     group = dat$group, cencode = 0)

# Extract P-value for Cause 1 (Cancer death) from Gray's Test
# The Tests matrix contains the chi-square, df, and p-value (pv)
p_val_gray <- ci$Tests[1, "pv"]
p_label <- paste0("Gray's Test (Cancer Death): p = ", sprintf("%.4f", p_val_gray))

# [3] Data Prep for Plotting
# Note: With groups, ci object indices increase
 #(Group A=Cancer death: Cause 1, Group B=Cardiovascular death: Cause 2, etc.)
df_cif <- rbind(
  data.frame(time = ci[[1]]$time, est = ci[[1]]$est,
             cause = "Cancer death", group = "Cancer death"),
  data.frame(time = ci[[3]]$time, est = ci[[3]]$est,
             cause = "Cancer death", group = "Cardiovascular death")
)

# [4] Plot
Figure_C10 <- ggplot(df_cif, aes(x = time, y = est, color = group)) +
  geom_step(linewidth = 1.1) +
  geom_hline(yintercept = 0.50, linetype = "dashed",
             color = "grey40", linewidth = 0.8) +
  # Add the P-value label
  annotate("text", x = 0.5, y = 0.9, label = p_label,
           color = "black", fontface = "bold", hjust = 0) +
  # Add the original threshold label
  annotate("text", x = 0.05, y = 0.53, label = "50% Incidence Threshold",
```

```r
          color = "grey40", fontface = "italic", hjust = 0) +
  scale_y_continuous(limits = c(0, 1), breaks = seq(0, 1, 0.25)) +
  scale_x_continuous(limits = c(0, t_max), breaks = seq(0, t_max, 2)) +
  labs(
    title = "Cumulative Incidence Function (CIF)",
    subtitle = "Gray's Test for Equality of CIFs",
    x = "Time (Years)",
    y = "Cumulative Incidence",
    color = "Event Type"
  ) +
  theme_classic(base_size = 12) +
  theme(legend.position = "right")

Figure_C10

#[4] Save (Optional)

ggsave("Figure_C10.png", Figure_C10, width = 6.5, height = 4.2, dpi = 600)
ggsave("Figure_C10.pdf", Figure_C10, width = 6.5, height = 4.2, device = cairo_pdf)
```

**Citation:** Soltanifar, M., Portuguese, A.J., Jeon, Y., Gauthier, J., & Lee, C.H. (2026). A North American Collaborative Atlas of Oncology Data Visualization with R Statistical Software. medRxiv preprint. https://doi.org/xx.xxxxx/xxxx.xx.xx.xxxxxxxx, ©2026 Soltanifar, M. et al. (CC BY 4.0/MIT)

**Figure 5.3.11.**

```r
# [1] Packages
library(ggplot2)
library(tidyr)
library(dplyr)

set.seed(11)

# [2-1] Time grid
t <- seq(0, 10, by = 0.1)

# [2-2] Simulated CIFs (Reaching your steepness goals)
# Using saturating functions: CIF = Plateau * (1 - exp(-rate * t))
cif_breast       <- 0.70 * (1 - exp(-0.45 * t))
cif_other_cancer <- 0.16 * (1 - exp(-0.22 * t))
cif_heart        <- 0.09 * (1 - exp(-0.18 * t))
cif_other        <- 0.05 * (1 - exp(-0.15 * t))

# [2-3] Ensure total does not exceed 1 (Numerical safety)
df <- data.frame(
  time = t,
  Breast_Cancer = cif_breast,
  Other_Cancers = cif_other_cancer,
  Heart_Disease = cif_heart,
  Other_Causes  = cif_other
)

# [2-4] THE FIX: Use pivot_longer instead of reshape()
# This avoids the "undefined columns" error by handling names directly
df_long <- df %>%
  pivot_longer(
    cols = -time,
    names_to = "cause",
    values_to = "cif"
  ) %>%
  # Clean up names for the legend
  mutate(cause = gsub("_", " ", cause))

# [3] Plot: Stacked CIF
Figure_C11 <- ggplot(df_long, aes(x = time, y = cif, fill = cause)) +
  # geom_area creates the "stacked" effect
  geom_area(alpha = 0.8, color = "white", linewidth = 0.2) +
  # Add the 50% threshold line requested previously
  geom_hline(yintercept = 0.50, linetype = "dashed", color = "black", alpha = 0.6) +
  annotate("text", x = 8.5, y = 0.525, label = "50% Cumulative Burden", size = 3.5) +
  scale_y_continuous(limits = c(0, 1), breaks = seq(0, 1, 0.1), expand = c(0,0)) +
  scale_x_continuous(limits = c(0, 10), breaks = seq(0, 10, 1), expand = c(0,0)) +
  scale_fill_manual(values = c(
    "Breast Cancer" = "#FFD700",
    "Heart Disease" = "#377eb8",
    "Other Cancers" = "#4daf4a",
    "Other Causes"  = "#984ea3"
```

```r
  )) +
  labs(
    title = "Stacked Cumulative Incidence Plot",
    subtitle = "Total burden of mortality over 10 years",
    x = "Time(Years Since Diagnosis)",
    y = "Total Cumulative Incidence",
    fill = "Cause of Event"
  ) +
  theme_minimal(base_size = 12) +
  theme(panel.grid.minor = element_blank())

Figure_C11

#[4] Save (Optional)

ggsave("Figure_C11.png", Figure_C11, width = 7.5, height = 5, dpi = 600)
ggsave("Figure_C11.pdf", Figure_C11, width = 7.5, height = 5, device = cairo_pdf)
```

**Citation:** Soltanifar, M., Portuguese, A.J., Jeon, Y., Gauthier, J., & Lee, C.H. (2026). A North American Collaborative Atlas of Oncology Data Visualization with R Statistical Software. medRxiv preprint. https://doi.org/xx.xxxxx/xxxx.xx.xx.xxxxxxxx, ©2026 Soltanifar, M. et al. (CC BY 4.0/MIT)

**Figure 5.3.12.**

```r
# [1] Packages
library(ggplot2)
library(dplyr)
library(tidyr)

# [2] Simulations

# [2-1] Simulate smooth hazard curves
df_haz <- tidyr::expand_grid(
  time = seq(0, 4, by = 0.02),
  arm  = c("Chemotherapy", "CART-cell therapy", "Immunotherapy")
) %>%
  mutate(
    hazard = case_when(
      # [1a] Chemotherapy: Flat baseline hazard
      arm == "Chemotherapy" ~ 2.25e-3,

      # [1b] CART-cell therapy: High initial risk, then decreasing
      arm == "CART-cell therapy" ~ 1.70e-3 + 1.25e-3 * exp(-1.2 * time),

      # [1c] Immunotherapy: Increasing hazard over time (e.g., late toxicity)
      arm == "Immunotherapy" ~ 1.80e-3 + 0.50e-3 * (1 - exp(-0.8 * time))
    )
  )

# [3] Plot
Figure_C12 <- ggplot(df_haz, aes(x = time, y = hazard, color = arm)) +
  # Set linetype directly in geom_line to avoid creating a second legend
  geom_line(linewidth = 1.1, linetype = "dashed") +
  scale_color_manual(
    values = c(
      "Chemotherapy" = "#00BFC4",      # Teal
      "CART-cell therapy" = "#F8766D", # Coral/Red
      "Immunotherapy" = "deeppink"     # Pink
    )
  ) +
  scale_y_continuous(
    labels = scales::label_number(accuracy = 0.00001)
  ) +
  labs(
    x = "Time (months)",
    y = expression(paste("Hazard (", lambda(t), ")")),
    color = "Treatment"
  ) +
  theme_bw(base_size = 12) +
  theme(
    legend.position = "right",
    panel.grid.minor = element_blank(),
    # Ensures no extra legend titles appear
    legend.title = element_text(face = "bold")
  )
```

```r
Figure_C12

#[4] Save (Optional)

ggsave("Figure_C12.png", Figure_C12, width = 7.5, height = 5, dpi = 600)
ggsave("Figure_C12.pdf", Figure_C12, width = 7.5, height = 5, device = cairo_pdf)
```

**Citation:** Soltanifar, M., Portuguese, A.J., Jeon, Y., Gauthier, J., & Lee, C.H. (2026). A North American Collaborative Atlas of Oncology Data Visualization with R Statistical Software. medRxiv preprint. https://doi.org/xx.xxxxx/xxxx.xx.xx.xxxxxxxx, ©2026 Soltanifar, M. et al. (CC BY 4.0/MIT)

**Figure 5.3.13.**

```
# [1] Packages
library(survival)
library(dplyr)
library(tidyr)
library(ggplot2)
set.seed(13013)


#[2] Simulation
# [2-1] (A) Simulate survival data for 3 SBR grades (piecewise hazards to mimic
                                                        #non-parallelity) ----
n_per_grade <- 220
grades <- c("SBR grade 1", "SBR grade 2", "SBR grade 3")

# [2-2] Piecewise exponential sampler (two intervals):
                                              #hazard = h1 for t<=t0, hazard = h2 for t>t0
rpexp2 <- function(n, h1, h2, t0) {
  u <- runif(n)
  H0 <- h1 * t0
  # If event happens before t0
  t_before <- -log(1 - u) / h1
  # Otherwise, shift after t0
  t_after <- t0 + (-log(1 - u) - H0) / h2
  ifelse(t_before <= t0, t_before, t_after)
}

t0 <- 85.2 # years (change-point)

sim_dat <- bind_rows(
  tibble(
    grade  = grades[1],
    time_e = rpexp2(n_per_grade, h1 = 0.12, h2 = 0.10, t0 = t0)
    # lower hazard, mild change
  ),
  tibble(
    grade  = grades[2],
    time_e = rpexp2(n_per_grade, h1 = 0.18, h2 = 0.15, t0 = t0)
    # intermediate hazard
  ),
  tibble(
    grade  = grades[3],
    time_e = rpexp2(n_per_grade, h1 = 0.32, h2 = 0.14, t0 = t0)
    # higher early hazard then attenuates
  )
) %>%
  mutate(
    # independent administrative censoring (in years)
    time_c = runif(n(), min = 0.3, max = 6.0),
    time   = pmin(time_e, time_c),
    status = as.integer(time_e <= time_c)
  ) %>%
```

```r
  select(time, status, grade) %>%
  mutate(grade = factor(grade, levels = grades))

#  [2-3] Kaplan-Meier by grade
fit <- survfit(Surv(time, status) ~ grade, data = sim_dat)

# Convert survfit to a tidy data frame (no external tidier needed)
km_df <- data.frame(
  time   = fit$time,
  surv   = fit$surv,
  strata = rep(names(fit$strata), fit$strata)
) %>%
  mutate(
    grade = gsub("^grade=", "", strata),
    # Work on log-time scale (avoid log(0))
    log_time = log(pmax(time, 1e-6)),
    # log(-log(S(t))) with numerical guards
    lmls = log(pmax(-log(pmin(pmax(surv, 1e-12), 1 - 1e-12)), 1e-12))
  )

# [3] Plot: log(-log(S(t))) vs log(time)
Figure_C13 <- ggplot(km_df, aes(x = log_time, y = lmls, color = grade)) +
  geom_step(linewidth = 0.9) +
  labs(
    x = "Log (Years since diagnosis)",
    y = "log(-log(Survival Probability))",
    color = "Scarff-Bloom-Richardson(SBR) grade"
  ) +
  theme_bw(base_size = 11) +
  theme(
    legend.position = "bottom",
    legend.title = element_text(face = "bold")
  ) +
  scale_color_brewer(palette = "Dark2")

Figure_C13

#[4] Save (Optional)

ggsave("Figure_C13.png", Figure_C13, width = 7.5, height = 5, dpi = 600)
ggsave("Figure_C13.pdf", Figure_C13, width = 7.5, height = 5, device = cairo_pdf)
```

**Figure 5.3.14.**

```r
# [1] Packages
library(survival)
library(ggplot2)
set.seed(20260216)

# [2] Simulations

# [2-1] Simulate a simple dataset with a mild PH violation

n  <- 220
grade <- rbinom(n, 1, 0.45)  # 1 = SBR grade 3, 0 = SBR grade 1

# Event times from a Weibull model with a time-varying effect (violates PH):
# hazard(t | grade) = h0(t) * exp( beta(t) * grade )
# where beta(t) changes over time (here: starts positive, dips, then rises slightly)
shape <- 1.20
lambda <- 0.08

# Discrete-time approximation to generate survival times under time-varying beta(t)
tmax <- 18
dt   <- 0.10
grid <- seq(dt, tmax, by = dt)

beta_t <- function(t) {
  # Smooth non-constant pattern to mimic a non-horizontal Schoenfeld smooth
  0.60 * exp(-t / 4) - 0.35 * exp(-((t - 10) / 4)^2) + 0.10 * (t / tmax)
}

H0_inc <- function(t) {
  # Weibull baseline cumulative hazard increment over (t-dt, t]
  # H0(t) = (lambda * t)^shape  => dH approx difference
  ((lambda * t)^shape - (lambda * (t - dt))^shape)
}

time <- rep(NA_real_, n)
status <- rep(0L, n)

for (i in seq_len(n)) {
  u <- runif(1)
  surv <- 1.0
  for (t in grid) {
    haz_inc <- H0_inc(t) * exp(beta_t(t) * grade[i])
    surv <- surv * exp(-haz_inc)
    if (surv <= u) {
      time[i] <- t
      status[i] <- 1L
      break
    }
  }
  if (is.na(time[i])) {
    time[i] <- tmax
```

```r
    status[i] <- 0L
  }
}

dat <- data.frame(
  time = time,
  status = status,
  grade = factor(grade, levels = c(0, 1), labels = c("SBR grade 1", "SBR grade 3"))
)


#  [2-2] Fit Cox PH model + compute Schoenfeld residual diagnostic

fit <- coxph(Surv(time, status) ~ grade, data = dat)

# PH test object (scaled Schoenfeld residuals)
zph <- cox.zph(fit, transform = "km")

# Extract the single-covariate residual series
# For a single covariate, zph$x is time and zph$y is a 1-column matrix
df <- data.frame(
  time = zph$x,
  resid = as.numeric(zph$y[, 1])
)

# [3] Plot
# Reference lines shown in the attached figure:
# - Null effect: log(HR)=0
# - Conventional Cox: log(HR)=0.66  (as displayed in the attached example)
ref_lines <- data.frame(
  ref = c("log(HR)=0 (Null effect)", "log(HR)=0.66 (Conventional Cox)"),
  y   = c(0.00, 0.66)
)

# Define professional colors
main_blue <- "#4682B4" # SteelBlue
band_fill <- "#A9A9A9" # DarkGray for a neutral, professional band

Figure_C14 <- ggplot(df, aes(x = time, y = resid)) +
  # Points with reduced alpha for less "noise"
  geom_point(shape = 1, size = 1.5, alpha = 0.5) +
  # Professional band: fill changed to neutral grey with higher transparency
  geom_smooth(method = "loess", se = TRUE, span = 0.55,
              linewidth = 1, color = main_blue, fill = band_fill, alpha = 0.2) +
  # Horizontal reference lines
  geom_hline(
    data = ref_lines,
    aes(yintercept = y, linetype = ref, color = ref),
    linewidth = 0.8,
    show.legend = TRUE
  ) +
  scale_linetype_manual(
    values = c("log(HR)=0 (Null effect)" = "dashed",
```

```
                "log(HR)=0.66 (Conventional Cox)" = "dotted")
  ) +
  scale_color_manual(
    values = c("log(HR)=0 (Null effect)" = "#B22222", # Firebrick red
                "log(HR)=0.66 (Conventional Cox)" = "#1F4E79") # Deep blue
  ) +
  labs(
    title = "Test for PH (SBR grade: grade 3 versus grade 1)",
    x = "Time (t)",
    y = expression(paste(beta, "(t) for Grade 3")), # Uses LaTeX-style symbol
    color = "Hazard Reference",     # Legend Title
    linetype = "Hazard Reference" # Must match color label to merge legends
  ) +
  theme_bw(base_size = 12) +
  theme(
    legend.position = "right",
    legend.title = element_text(face = "bold"), # Bold legend title
    plot.title = element_text(hjust = 0.5, face = "bold"),
    panel.grid.minor = element_blank() # Clean up the grid
  ) +
  # This ensures both color and linetype are treated as one legend entry
  guides(color = guide_legend(override.aes = list(fill = NA)))

Figure_C14

#[4] Save (Optional)

ggsave("Figure_C14.png", Figure_C14, width = 7.5, height = 5, dpi = 600)
ggsave("Figure_C14.pdf", Figure_C14, width = 7.5, height = 5, device = cairo_pdf)
```

**Citation:** Soltanifar, M., Portuguese, A.J., Jeon, Y., Gauthier, J., & Lee, C.H. (2026). A North American Collaborative Atlas of Oncology Data Visualization with R Statistical Software. medRxiv preprint. https://doi.org/xx.xxxxx/xxxx.xx.xx.xxxxxxxx, ©2026 Soltanifar, M. et al. (CC BY 4.0/MIT)

**Figure 5.3.15.**

```r
# [1] Packages
library(survival)
library(dplyr)
library(tidyr)
library(ggplot2)
set.seed(2026)

# [2] Simulations
n <- 400
log_bili <- seq(0, 5, length.out = n)

# [2-1] Introduce a NON-LINEAR functional form (a sine wave)
# This will cause the linear Cox model to fail significantly.
functional_form <- 5* (sin(10*log_bili))*(log_bili)^3
base_rate <- 0.01
t_event  <- rexp(n, rate = base_rate * exp(functional_form))
t_cens   <- rexp(n, rate = 0.05)
time     <- pmin(t_event, t_cens)
status   <- as.integer(t_event <= t_cens)

dat <- tibble(id = seq_len(n), time = time, status = status, log_bili = log_bili)

# [2-2] Fit a LINEAR Cox PH model (Mis-specified)
fit <- coxph(Surv(time, status) ~ log_bili, data = dat)
dat$martingale <- residuals(fit, type = "martingale")

# [2-3] Process Observed Data
dat_ord <- dat %>%
  arrange(log_bili) %>%
  mutate(cum_mart = cumsum(martingale))

# [3] Bootstrap (Null Distribution)
# To get p < 0.05, these MUST be simulated under the NULL hypothesis (linearity)
B <- 100
boot_list <- vector("list", B)

# We use the 'multiplier bootstrap' approach typical for these tests
# simulating what the process SHOULD look like if the linear model were true
for (b in seq_len(B)) {
  # Simulating Gaussian noise for the null process
  # This ensures the gold lines stay near zero, while the blue line drifts away
  w <- rnorm(n)
  boot_list[[b]] <- dat_ord %>%
    mutate(
      cum_mart = cumsum(martingale * w),
      curve = paste0("Bootstrap ", b),
      legend_group = "Bootstrap (Null)"
    ) %>%
    select(log_bili, cum_mart, curve, legend_group)
}
```

```r
boot_df <- bind_rows(boot_list)

# [4] Observed and Smooth Data
obs_df <- dat_ord %>%
  mutate(curve = "Observed", legend_group = "Observed") %>%
  select(log_bili, cum_mart, curve, legend_group)

smooth_df <- {
  lo <- loess(cum_mart ~ log_bili, data = obs_df, span = 0.4)
  tibble(
    log_bili = seq(min(obs_df$log_bili), max(obs_df$log_bili), length.out = 200),
    cum_mart = as.numeric(predict(lo, newdata = seq(min(obs_df$log_bili),
                                          max(obs_df$log_bili), length.out = 200))),
    curve = "Loess smooth",
    legend_group = "Loess smooth"
  )
}

# [5] P-Value Calculation
obs_max <- max(abs(obs_df$cum_mart))
boot_maxima <- boot_df %>%
  group_by(curve) %>%
  summarize(m = max(abs(cum_mart))) %>%
  pull(m)

p_val <- mean(boot_maxima >= obs_max) # This will now be < 0.05
p_label <- paste0("Kolmogorov-type Supremum Test\n", "p-value: ", round(p_val, 3))

# [6] Plot
plot_df <- bind_rows(boot_df, obs_df, smooth_df)

Figure_C15 <-ggplot(plot_df, aes(x = log_bili, y = cum_mart, group = curve,
                                                      color = legend_group)) +
  # Bootstrap lines
  geom_line(data = filter(plot_df, legend_group == "Bootstrap (Null)"),
            alpha = 0.15, linewidth = 0.3) +
  # Loess line
  geom_line(data = filter(plot_df, legend_group == "Loess smooth"),
            linetype = "dashed", linewidth = 1) +
  # Observed line
  geom_line(data = filter(plot_df, legend_group == "Observed"),
            linewidth = 1.2) +
  # P-value box
  annotate("label", x = 0, y = 30,
           label = p_label, hjust = 0, fontface = "bold", size = 4) +
  scale_color_manual(values = c(
    "Bootstrap (Null)" = "green",
    "Observed" = "orange",
    "Loess smooth" = "black"
  )) +
  labs(
    title = "Cumulative Martingale Residuals vs log(bilirubin)",
    subtitle = "Checking for linearity in Cox Proportional Hazards Model",
```

```
    x = "Log (bilirubin)",
    y = "Cumulative Martingale Residuals",
    color = "Curve Type"
  ) +
  scale_x_continuous(breaks = seq(0,5,0.5), limits = c(0,5))+
  scale_y_continuous(breaks = seq(-40,40,5), limits = c(-40,40))+
  theme_minimal() +
  guides(color = guide_legend(override.aes = list(alpha = 1, linewidth = 1)))

Figure_C15

#[7] Save (Optional)

ggsave("Figure_C15.png", Figure_C15, width = 9, height = 6, dpi = 600)
ggsave("Figure_C15.pdf", Figure_C15, width = 9, height = 6, device = cairo_pdf)
```

**Citation:** Soltanifar, M., Portuguese, A.J., Jeon, Y., Gauthier, J., & Lee, C.H. (2026). A North American Collaborative Atlas of Oncology Data Visualization with R Statistical Software. medRxiv preprint. https://doi.org/xx.xxxxx/xxxx.xx.xx.xxxxxxxx, ©2026 Soltanifar, M. et al. (CC BY 4.0/MIT)

**Figure 5.3.16.**

```r
# [1] Packages
library(survival)
library(ggplot2)
library(dplyr)
library(patchwork)
library(broom)
library(tidyr)
set.seed(20260217)

# [2] Simulations
n_per_arm <- 200
sim_arm <- function(n, label, hr) {
  t <- rweibull(n, 1.25, 30 / (hr^(1/1.25)))
  c <- rexp(n, 0.015)
  data.frame(arm = label, time = pmin(t, c), status = as.integer(t <= c))
}
dat <-
bind_rows(sim_arm(n_per_arm, "Control", 1), sim_arm(n_per_arm, "Intervention", 0.7))
%>% mutate(arm = factor(arm, levels = c("Control", "Intervention")))

# [3] Preparations
tau_marker <- 36
arm_levels <- levels(dat$arm)

# [3-1] Robust subsetting for survival objects
km_by_arm <- lapply(arm_levels, function(a) {
  survfit(Surv(time, status) ~ 1, data = dat %>% dplyr::filter(arm == a))
})
names(km_by_arm) <- arm_levels

rmst_calc <- function(f, tau) {
  if (length(f$time) == 0) return(tau)
  t <- c(0, f$time[f$time <= tau], tau)
  s <- c(1, f$surv[f$time <= tau])
  sum(s * diff(t))
}

# [4] PANEL A: Survival Curves with Bounded RMST Shading ---
km_all <- survfit(Surv(time, status) ~ arm, data = dat)
tidy_km <- broom::tidy(km_all) %>% mutate(arm = gsub("arm=", "", strata)) #

# Create bounded step data for shading
# This creates a shared time grid to ensure the ribbon is strictly between curves
time_points <- sort(unique(c(0, tidy_km$time[tidy_km$time <= tau_marker], tau_marker)))
shade_df <- data.frame(time = time_points) %>%
  rowwise() %>%
  mutate(
    Control = summary(km_by_arm[["Control"]], times = time)$surv,
    Intervention = summary(km_by_arm[["Intervention"]], times = time)$surv
  ) %>% ungroup()
```

```r
panel_a <- ggplot() +
  scale_x_continuous(limits = c(0, 50), breaks = c(0,25,50))+
    # Only highlight between the curves up to tau
  geom_ribbon(data = shade_df, aes(x = time, ymin = Control, ymax = Intervention),
              fill = "skyblue", alpha = 0.5) +
  geom_step(data = tidy_km, aes(x = time, y = estimate, color = arm), linewidth = 1) +
  geom_vline(xintercept = tau_marker, linetype = "dashed", color = "red", linewidth = 0.7) +
  annotate("label", x = tau_marker, y = 0.01, label = paste0("\u03C4 = ", tau_marker),
           color = "red", fontface = "bold", fill = "white") +
  scale_color_manual(values = c("Control" = "orange", "Intervention" = "darkslategrey")) +
  labs(title = "A. Survival with RMST Area",
       x = "Time (Months)", y = "Survival Probability", color = "Arm") +
  theme_minimal() + theme(legend.position = "bottom")

#  [5] PANEL B: RMST Difference vs Time
tau_grid <- seq(3, 48, by = 1)
rmst_df <- data.frame(tau = tau_grid) %>%
  rowwise() %>%
  mutate(delta =
  rmst_calc(km_by_arm[["Intervention"]], tau) - rmst_calc(km_by_arm[["Control"]], tau))
   %>% ungroup()

panel_b <- ggplot(rmst_df, aes(x = tau, y = delta)) +
  scale_x_continuous(limits = c(0, 50), breaks = c(0,25,50))+
  geom_hline(yintercept = 0, linetype = "dashed", color = "grey50") +
  geom_line(color = "blue", linewidth = 1) +
  # Highlight the specific tau point from Panel A
  geom_vline(xintercept = tau_marker, linetype = "dashed", color = "red", linewidth = 0.7) +
  annotate("label", x = tau_marker, y = 0.01, label = paste0("\u03C4 = ", tau_marker),
           color = "red", fontface = "bold", fill = "white")+
  geom_point(data = filter(rmst_df, tau == tau_marker), color = "red", size = 3) +
  labs(title = "B. RMST Difference over Time",
       x = "Restriction Time (Months)", y = expression(Delta*RMST ~ (Months))) +
  theme_minimal()

# [6] PLOTS
Figure_C16 <- panel_a + panel_b + plot_layout(ncol = 1)
Figure_C16

#[7] Save (Optional)

ggsave("Figure_C16.png", Figure_C16, width = 6, height = 9, dpi = 600)
ggsave("Figure_C16.pdf", Figure_C16, width = 6, height = 9, device = cairo_pdf)
```

**Figure 5.3.17.**

```r
# [1] Packages
library(ggplot2)
library(dplyr)
library(tidyr)
set.seed(1703)

#[2] Simulations:

# Time grid (months)
t <- seq(0, 48, by = 1)

# Helper functions to create smooth, plausible multi-state occupancy shapes
# (Designed to resemble the attached panel: early CR peak then taper;
  #relapse small hump; death rises.)
cr_shape <- function(t, peak = 0.72, rise = 0.60, decay = 0.020, shift = 0) {
  # quick rise then slow decay
  p <- peak * (1 - exp(-rise * pmax(t - shift, 0))) * exp(-decay * pmax(t - shift, 0))
  p
}

relapse_shape <- function(t, amp = 0.14, center = 18, spread = 9, shift = 0) {
  # modest hump
  tt <- pmax(t - shift, 0)
  p <- amp * exp(-0.5 * ((tt - center) / spread)^2)
  p
}

death_shape <- function(t, maxp = 0.55, rate = 0.055, shift = 0) {
  # monotone increase approaching maxp
  tt <- pmax(t - shift, 0)
  p <- maxp * (1 - exp(-rate * tt))
  p
}

# --- Simulate two arms with modest separation (consistent with a didactic figure) ---
dat_midostaurin <- tibble(
  time = t,
  arm  = "Midostaurin",
  CR      = cr_shape(t, peak = 0.72, rise = 0.75, decay = 0.024, shift = 0),
  Relapse = relapse_shape(t, amp = 0.16, center = 18, spread = 10, shift = 0),
  Death   = death_shape(t, maxp = 0.48, rate = 0.060, shift = 0)
)

dat_placebo <- tibble(
  time = t,
  arm  = "Placebo",
  CR      = cr_shape(t, peak = 0.66, rise = 0.70, decay = 0.028, shift = 0),
  Relapse = relapse_shape(t, amp = 0.12, center = 16, spread = 10, shift = 0),
  Death   = death_shape(t, maxp = 0.55, rate = 0.055, shift = 0)
)
```

```r
dat <- bind_rows(dat_midostaurin, dat_placebo) |>
  pivot_longer(cols = c("CR", "Relapse", "Death"),
               names_to = "state",
               values_to = "p_state") |>
  mutate(
    stratum = "ITD-Low",
    state = factor(state, levels = c("CR", "Relapse", "Death")),
    arm   = factor(arm, levels = c("Midostaurin", "Placebo"))
  )

# Publication-style palette (manual; professional; consistent across states)
state_cols <- c(
  "CR"      = "#1B9E77",  # green-teal
  "Relapse" = "#D95F02",  # orange
  "Death"   = "#1F1F1F"   # near-black
)

# [3] Plot
Figure_C17 <- ggplot(dat, aes(x = time, y = p_state, color = state, linetype = arm)) +
  geom_line(linewidth = 1.05) +
  scale_color_manual(values = state_cols, name = "State") +
  scale_linetype_manual(values = c("Midostaurin" = "dashed", "Placebo" = "solid"),
                        name = "Arm") +
  coord_cartesian(ylim = c(0, 1)) +
  labs(
    title = "Current State Probability Curves: ITD-Low Stratification",
    x = "Time(Months)",
    y = "Current State Probability"
  ) +
  theme_bw(base_size = 12) +
  theme(
    legend.position = "right",
    plot.title = element_text(face = "bold"),
    legend.box = "vertical"
  )

Figure_C17

#[7] Save (Optional)

ggsave("Figure_C17.png", Figure_C17, width = 6, height = 6, dpi = 600)
ggsave("Figure_C17.pdf", Figure_C17, width = 6, height = 6, device = cairo_pdf)
```

**Citation:** Soltanifar, M., Portuguese, A.J., Jeon, Y., Gauthier, J., & Lee, C.H. (2026). A North American Collaborative Atlas of Oncology Data Visualization with R Statistical Software. medRxiv preprint. https://doi.org/xx.xxxxx/xxxx.xx.xx.xxxxxxxx, ©2026 Soltanifar, M. et al. (CC BY 4.0/MIT)

**Figure 5.3.18.**

```r
#[1] Packages
library(ggplot2)
library(dplyr)
library(tidyr)
library(patchwork)
library(scales)

#[2] Simulations
bor_counts <- tibble::tribble(
  ~arm,         ~response_cat, ~n,
  "IPI+NIVO",   "CR",           21,
  "IPI+NIVO",   "PR",           16,
  "IPI+NIVO",   "SD",           17,
  "IPI+NIVO",   "PD",          54,
  "IPI+NIVO",   "NE",           5,
  "Anti-PD1",   "CR",           37,
  "Anti-PD1",   "PR",           11,
  "Anti-PD1",   "SD",           16,
  "Anti-PD1",   "PD",          115,
  "Anti-PD1",   "NE",           18
) %>%
  mutate(response_cat = factor(response_cat, levels = c("CR","PR","SD","PD","NE")),
         arm = factor(arm, levels = c("IPI+NIVO","Anti-PD1")))

bor_pct <- bor_counts %>%
  group_by(arm) %>%
  mutate(N_arm = sum(n),
         pct = 100 * n / N_arm) %>%
  ungroup()

#[3] Plots
# [3-1] Panel A: grouped bar chart with percentages on top
gp18pA <- ggplot(bor_pct, aes(x = response_cat, y = pct, fill = arm)) +
  geom_col(position = position_dodge(width = 0.75), width = 0.65, color = "white") +
  # Add percentage labels on top of bars
  geom_text(aes(label = sprintf("%.1f%%", pct)),
            position = position_dodge(width = 0.75),
            vjust = -0.5, size = 3) +
  scale_y_continuous(limits = c(0, 100), breaks = seq(0, 100, 10),
                     labels = function(x) paste0(x, "%")) +
  labs(
    title = "A. Best Overall Response (BOR) distribution",
    x = NULL,
    y = "Proportion of patients (%)",
    fill = "Treatment Arm"
  ) +
  theme_minimal(base_size = 12) +
  theme(
    legend.position = "right",
    panel.grid.minor = element_blank(),
    plot.title.position = "plot"
```

```
  )

# [3-2] Panel B: stacked bar graph(s) with percentages inside segments
stack_df <- bor_counts %>%
  group_by(arm) %>%
  mutate(prop = n / sum(n)) %>%
  ungroup()

gp18pB <- ggplot(stack_df, aes(x = 1, y = prop, fill = response_cat)) +
  geom_col(width = 0.7, color = "white") +
  geom_text(
    aes(label = scales::percent(prop, accuracy = 0.1)),
    position = position_stack(vjust = 0.5),
    color = "black",
    size = 3
  ) +
  facet_wrap(~ arm, nrow = 1) +
  scale_y_continuous(
    limits = c(0, 1),
    breaks = seq(0, 1, 0.1),
    labels = scales::percent_format(accuracy = 1),
    expand = c(0, 0)
  ) +
  scale_x_continuous(breaks = NULL) +
  scale_fill_manual(
    values = c("CR"="#F8766D", "PR"="#A3A500", "SD"="#00BF7D", "PD"="#00B0F6",
               "NE"="#E76BF3"),
    breaks = c("CR","PR","SD","PD","NE")) +
  labs(
    title = "B. BOR composition within each arm",
    fill = "BOR Category",
    x = NULL,
    y = "Proportion"
  ) +
  theme_minimal(base_size = 12) +
  theme(
    legend.position = "right",
    strip.text = element_text(size = 11),
    panel.grid.minor = element_blank(),
    plot.title.position = "plot"
  )

# ---- Combine panels vertically ----
Figure_C18 <- gp18pA / gp18pB + plot_layout(guides = "collect") &
  theme(legend.position = "right")

Figure_C18
#[7] Save (Optional)
ggsave("Figure_C18.png", Figure_C18, width = 10.5, height = 8, dpi = 600)
ggsave("Figure_C18.pdf", Figure_C18, width = 10.5, height = 8, device = cairo_pdf)
```

**Figure 5.3.19.**

```r
#[1] Packages
library(ggplot2)
library(dplyr)
library(tidyr)
library(patchwork)
library(scales)
set.seed(19)

#[2] Simulations

#[2-1]
# -------------------------
# (A) Lollipop data (counts by position)
# -------------------------
# Major hotspots (approximate orders of magnitude)
hotspots <- tibble::tribble(
  ~position, ~n_patients, ~mutation_label,
  -124,      927,         "-124C>T (hotspot)",
  -146,      278,         "-146C>T (hotspot)",
  -138,       16,         "-138C>T (hotspot)"
)

# Background minor positions (sparse)
minor_positions <- sort(sample(seq(-346, -58, by = 2), size = 22, replace = FALSE))
minor_counts <- sample(c(1,1,1,2,2,3,4,5), size = length(minor_positions), replace = TRUE)

minor <- tibble(
  position = minor_positions,
  n_patients = minor_counts,
  mutation_label = NA_character_
)

lolli <- bind_rows(hotspots, minor) %>%
  arrange(position) %>%
  mutate(
    is_hotspot = if_else(position %in% c(-124, -146, -138), "Hotspot", "Other"),
    is_hotspot = factor(is_hotspot, levels = c("Hotspot", "Other"))
  )

#[2-2]
# -------------------------
# (B) Hotspot composition data (stacked percent by cancer type)
# -------------------------
cancer_levels <- c(
  "Bladder cancer","Glioma","Melanoma","Non-small-cell lung cancer",
  "Head and neck carcinoma","Colorectal cancer","Skin cancer (non-melanoma)","Other"
)

# Percent breakdowns (chosen to resemble a plausible mix; rows sum to 100)
hotspot_mix <- tibble::tribble(
  ~hotspot, ~`Bladder cancer`, ~Glioma, ~Melanoma, ~`Non-small-cell lung cancer`,
```

```
  ~`Head and neck carcinoma`, ~`Colorectal cancer`, ~`Skin cancer (non-melanoma)`, ~Other,
  "-124",   30,                25,      10,        5,
  10,                     5,                      5,                          10,
  "-146",   20,                20,      20,        10,
  10,                     5,                      5,                          10,
  "-138",   10,                35,      25,        5,
  5,                      5,                      5,                          10,
  "Other",  15,                15,      15,        15,
  10,                     10,                     10,                         10
) %>%
  pivot_longer(-hotspot, names_to = "cancer_type", values_to = "percent") %>%
  mutate(
    hotspot = factor(hotspot, levels = c("-124","-146","-138","Other")),
    cancer_type = factor(cancer_type, levels = cancer_levels)
  )

#[3] Plot:
#[3-1]
# -------------------------
# Plot A: Lollipop map (log10 y to handle hotspot magnitude)
# -------------------------
gp19pA <- ggplot(lolli, aes(x = position, y = n_patients)) +
  geom_segment(aes(xend = position, y = 1, yend = n_patients),
               linewidth = 0.5, color = "grey40") +
  geom_point(aes(fill = is_hotspot),
             shape = 21, size = 2.8, color = "black", stroke = 0.4) +
  geom_text(
    data = lolli %>% filter(!is.na(mutation_label)),
    aes(label = mutation_label),
    nudge_y = 0.15,
    vjust = 0,
    size = 3.2
  ) +
  scale_y_log10(
    breaks = c(1, 5, 10, 50, 100, 250, 750, 1000),
    labels = comma
  ) +
  coord_cartesian(xlim = c(-346, 1)) +
  labs(
    title = "A. Lollipop Mutation Map (positions relative to TSS)",
    x = "Position (bp; TSS at +1)",
    y = "Number of Patients (log scale)",
    fill = "Site Class"
  ) +
  theme_minimal(base_size = 12) +
  theme(
    legend.position = "right",
    panel.grid.minor = element_blank(),
    plot.title.position = "plot"
  )

#[3-2]
# -------------------------
```

```
# Plot B: stacked % bar for hotspot tumor-type composition
# -------------------------
gp19pB <- ggplot(hotspot_mix, aes(x = hotspot, y = percent, fill = cancer_type)) +
  geom_col(width = 0.75, color = "white", linewidth = 0.2) +
  scale_y_continuous(limits = c(0, 100), breaks = seq(0, 100, 25),
                     labels = function(x) paste0(x, "%")) +
  labs(
    title = "B. Tumor-type Composition at Each Hotspot",
    x = NULL,
    y = "Percent of Cases",
    fill = "Cancer Type"
  ) +
  theme_minimal(base_size = 12) +
  theme(
    legend.position = "right",
    panel.grid.minor = element_blank(),
    plot.title.position = "plot"
  )

#[3-3]
# -------------------------
# Combine panels horizontally and collect legends on the right
# -------------------------
Figure_C19 <- (gp19pA / gp19pB) +
  plot_layout(heights = c(1.25, 1), guides = "collect") &
  theme(legend.position = "right")


Figure_C19

#[4] Save (Optional)

ggsave("Figure_C19.png", Figure_C19, width = 9, height = 9, dpi = 600)
ggsave("Figure_C19.pdf", Figure_C19, width = 9, height = 9, device = cairo_pdf)
```

**Citation:** Soltanifar, M., Portuguese, A.J., Jeon, Y., Gauthier, J., & Lee, C.H. (2026). A North American Collaborative Atlas of Oncology Data Visualization with R Statistical Software. medRxiv preprint. https://doi.org/xx.xxxxx/xxxx.xx.xx.xxxxxxxx, ©2026 Soltanifar, M. et al. (CC BY 4.0/MIT)

**Figure 5.3.20.**

```
# [1] Packages
library(ggplot2)
library(dplyr)
library(ggrepel)
set.seed(2026)

# [2] Simulation
n <- 500
dat <- tibble(
  gene_id = paste0("GENE", seq_len(n)),
  # Generate a spread of fold changes
  log2FC  = rnorm(n, mean = 0, sd = 2.5)
) |>
  mutate(
    # Model p-values based on log2FC to create the volcano 'wings'
    # Higher absolute log2FC leads to lower p-values (higher -log10p)
    effect_size = abs(log2FC) * 1.5,
    noise = rnorm(n, mean = 0, sd = 1.2),
    neglog10_p = pmax(0, effect_size + noise)
  )

# [3] Thresholds
fc_thr <- 1.0
# Setting a significance threshold on the -log10 scale (approx p < 0.05)
neglog10_p_thr <- -log10(0.05)

dat <- dat |>
  mutate(
    group = case_when(
      log2FC >=  fc_thr & neglog10_p > neglog10_p_thr ~ "Upregulated in CPR",
      log2FC <= -fc_thr & neglog10_p > neglog10_p_thr ~ "Upregulated in non-CPR",
      TRUE                                            ~ "Not significant"
    )
  )

# [4] Label top 10 signals
label_dat <- dat |>
  filter(group != "Not significant") |>
  arrange(desc(neglog10_p)) |>
  slice_head(n = 10)

# [5] Volcano Plot
Figure_C20 <-ggplot(dat, aes(x = log2FC, y = neglog10_p, color = group)) +
  geom_point(size = 1.8, alpha = 0.7) + # Slightly smaller points for higher density
  geom_vline(xintercept = c(-fc_thr, fc_thr), linetype = "dashed", linewidth = 0.5) +
  geom_hline(yintercept = neglog10_p_thr, linetype = "dashed", linewidth = 0.5) +
  geom_text_repel(
    data = label_dat,
    aes(label = gene_id),
    size = 3,
    box.padding = 0.35,
```

```r
    point.padding = 0.25,
    max.overlaps = Inf,
    show.legend = FALSE
  ) +
  scale_color_manual(
    values = c(
      "Upregulated in CPR"     = "#B82229",
      "Upregulated in non-CPR" = "#000080",
      "Not significant"        = "#808080"
    )
  ) +
  labs(
    title = "Volcano plot of DEGs (CPR vs non-CPR)",
    x = "Fold Change (Log2)",
    y = expression(paste("Significance (", -log[10], " p-value)")),
    color = "Differential Expression Status" # [2] Added suitable legend title
  ) +
  theme_classic(base_size = 12) +
  theme(
    legend.position = "bottom",
    plot.title = element_text(face = "bold")
  )

Figure_C20


#[6] Save (Optional)

ggsave("Figure_C20.png", Figure_C20, width = 8, height = 8, dpi = 600)
ggsave("Figure_C20.pdf", Figure_C20, width = 8, height = 8, device = cairo_pdf)
```

**Citation:** Soltanifar, M., Portuguese, A.J., Jeon, Y., Gauthier, J., & Lee, C.H. (2026). A North American Collaborative Atlas of Oncology Data Visualization with R Statistical Software. medRxiv preprint. https://doi.org/xx.xxxxx/xxxx.xx.xx.xxxxxxxx, ©2026 Soltanifar, M. et al. (CC BY 4.0/MIT)

**Figure 5.3.21.**

```r
# [1] Packages
library(dplyr)
library(tidyr)
library(ggplot2)
set.seed(20260221)

#[2] Simulations

# [2-1] Simulate patient-level longitudinal PRO change-from-baseline
n_patients <- 120
weeks <- seq(0, 72, by = 3)

pat <- tibble(
  id  = sprintf("P%03d", 1:n_patients),
  arm = rep(c("2.5 mg/kg", "3.4 mg/kg"), each = n_patients/2)
)

# [2-2] Patient-level random intercept + noise; arm-specific mean trajectory
dat <- pat %>%
  tidyr::crossing(week = weeks) %>%
  mutate(
    b_i = rnorm(n_patients)[match(id, pat$id)] * 0.25,              # subject heterogeneity
    # Mean trajectory (example): mild worsening then stabilization; higher dose slightly better
    mu  = ifelse(
      arm == "2.5 mg/kg",
      0.10 + 0.35 * exp(-week/18) - 0.25 * exp(-week/6),
      0.00 + 0.25 * exp(-week/18) - 0.20 * exp(-week/6)
    ),
    pro_change = mu + b_i + rnorm(n(), sd = 0.35)
  ) %>%
  select(id, arm, week, pro_change)

# [2-3] Summarize mean and 95% CI by arm/visit ----
sumdat <- dat %>%
  group_by(arm, week) %>%
  summarise(
    n    = n(),
    mean = mean(pro_change),
    se   = sd(pro_change) / sqrt(n),
    lcl  = mean - 1.96 * se,
    ucl  = mean + 1.96 * se,
    .groups = "drop"
  )

# [3] Plot
Figure_C21 <- ggplot(sumdat, aes(x = week, y = mean, color = arm, fill = arm)) +
  geom_hline(yintercept = 0, linewidth = 0.4, linetype = "solid") +
  geom_ribbon(aes(ymin = lcl, ymax = ucl), alpha = 0.18, color = NA) +
  geom_line(linewidth = 0.9) +
  scale_color_manual(
    values = c("2.5 mg/kg" = "#D55E00", "3.4 mg/kg" = "#0072B2"),
```

```r
    name = "Dose Group"
  ) +
  scale_fill_manual(
    values = c("2.5 mg/kg" = "#D55E00", "3.4 mg/kg" = "#0072B2"),
    name = "Dose Group"
  ) +
  scale_x_continuous(breaks = seq(0,80,5))+
  scale_y_continuous(breaks = seq(-0.25,0.45,0.05))+
  labs(
    title = "Longitudinal PRO Mean Change from Baseline",
    x = "Visit (weeks from baseline)",
    y = "Mean Change from Baseline (95%CI): PRO-CTCAE rating"
  ) +
  theme_bw(base_size = 11) +
  theme(
    legend.position = "right",
    legend.title = element_text(face = "bold"),
    plot.title = element_text(face = "bold"),
    panel.grid.minor = element_blank()
  )

Figure_C21


#[4] Save (Optional)

ggsave("Figure_C21.png", Figure_C21, width = 8, height = 8, dpi = 600)
ggsave("Figure_C21.pdf", Figure_C21, width = 8, height = 8, device = cairo_pdf)
```

**Citation:** Soltanifar, M., Portuguese, A.J., Jeon, Y., Gauthier, J., & Lee, C.H. (2026). A North American Collaborative Atlas of Oncology Data Visualization with R Statistical Software. medRxiv preprint. https://doi.org/xx.xxxxx/xxxx.xx.xx.xxxxxxxx, ©2026 Soltanifar, M. et al. (CC BY 4.0/MIT)

**Figure 5.3.22.**

```r
# [1] Packages
library(ggplot2)
library(dplyr)
library(tidyr)
set.seed(22022)

# [2] Simulations
# [2-1] Increase SD (sigma) to decrease the slope of the CDF curves
n_placebo <- 180
n_telo250 <- 180
n_telo500 <- 180

# Increasing SD from ~1.4 to 2.5 makes the curves much "flatter" (smaller slope)
dat <- bind_rows(
  tibble(arm = "Placebo",             change = rnorm(n_placebo, mean = -1.3, sd = 6.5)),
  tibble(arm = "Telotristat 250 mg", change = rnorm(n_telo250, mean = -2.5, sd = 2.5)),
  tibble(arm = "Telotristat 500 mg", change = rnorm(n_telo500, mean = -3.8, sd = 4.5))
) %>%
  mutate(
    change = pmax(pmin(change, 2.0), -10.0)
  )

# [2-4] Build smooth CDF curves
fit <- dat %>%
  group_by(arm) %>%
  summarise(mu = mean(change), sigma = sd(change), .groups = "drop")

xgrid <- seq(-10, 2, length.out = 600)

cdf_df <- fit %>%
  tidyr::crossing(x = xgrid) %>%
  mutate(
    cdf = pnorm(x, mean = mu, sd = sigma),
    cum_pct = 100 * cdf
  )

# [2-5] Meaningful change thresholds
thr_adequate_relief <- -3.52
thr_eortc_diarrhea  <- -2.56

# [3] Plot
Figure_C22 <- ggplot(cdf_df, aes(x = x, y = cum_pct, color = arm)) +
  geom_line(linewidth = 1.1) +
  # Use distinct linetypes and highly contrasting colors for vertical lines
  geom_vline(aes(xintercept = thr_adequate_relief, linetype = "Adequate Relief (-3.52)"),
             color = "#6A3D9A", linewidth = 1) +
  geom_vline(aes(xintercept = thr_eortc_diarrhea, linetype = "EORTC Diarrhea (-2.56)"),
             color = "#E6550D", linewidth = 1) +
  geom_hline(yintercept = 50, color = "black", linetype = "dotted", linewidth = 0.8) +
  annotate("text", x = -9, y = 52, label = "50% Threshold", size = 3.5, fontface = "italic", hjust = 0)
  scale_y_continuous(limits = c(0, 100), breaks = seq(0, 100, 10), expand = c(0, 0)) +
```

```r
  scale_x_continuous(limits = c(-10, 2), breaks = seq(-10, 2, 1)) +
  scale_color_manual(values = c(
    "Telotristat 250 mg" = "#2C7FB8",
    "Telotristat 500 mg" = "#08519C",
    "Placebo"            = "#6B6B6B"
  )) +
  # Assigned dashed vs solid for maximum distinction
  scale_linetype_manual(values = c(
    "Adequate Relief (-3.52)" = "dashed",
    "EORTC Diarrhea (-2.56)"  = "solid"
  )) +
  labs(
    title = "Cumulative Threshold Response Curves",
    subtitle = "",
    x = "Change in Average Bowel Movement Frequency from Baseline at Week 12",
    y = "Cumulative Percent of Patients Achieving Each Level of Change",
    color = "Treatment Arm",       # [1] Legend Title
    linetype = "Meaningful Change"  # [1] Legend Title
  ) +
  theme_minimal(base_size = 12) +
  theme(
    legend.position = "bottom",
    legend.box = "vertical",
    panel.grid.minor = element_blank(),
    plot.title = element_text(face = "bold")
  ) +
  guides(
    color = guide_legend(order = 1, nrow = 1),
    linetype = guide_legend(order = 2, nrow = 1)
  )

Figure_C22

#[4] Save (Optional)
ggsave("Figure_C22.png", Figure_C22, width = 8, height = 8, dpi = 600)
ggsave("Figure_C22.pdf", Figure_C22, width = 8, height = 8, device = cairo_pdf)
```

**Figure 5.3.23.**

```r
# [1] Packages
library(survival)
library(dplyr)
library(tidyr)
library(ggplot2)
library(cowplot)
set.seed(23023)

# [2] Simulations

# [2-1] Simulate three-arm TtD (months)
n1 <- 130   # Arm 1: SOC
n2 <- 120   # Arm 2: SLNB
n3 <- 110   # Arm 3: ALND
t_admin <- 12

sim_piecewise_exp <- function(n, t_switch = 6,
                              lambda_early = 0.06,
                              lambda_late  = 0.20)
{
  t1 <- rexp(n, rate = lambda_early)
  t_evt <- ifelse(t1 < t_switch,
                t1,
                t_switch + rexp(n, rate = lambda_late))
  t_evt
}

# Hazards: Arm 1 (Fastest deterioration), Arm 2 (Moderate), Arm 3 (Slowest/Reference)
t_evt_1 <- sim_piecewise_exp(n1, t_switch = 6, lambda_early = 0.09, lambda_late = 0.30)
t_evt_2 <- sim_piecewise_exp(n2, t_switch = 6, lambda_early = 0.06, lambda_late = 0.18)
t_evt_3 <- sim_piecewise_exp(n3, t_switch = 6, lambda_early = 0.04, lambda_late = 0.12)

# Random censoring
cenz_1 <- runif(n1, min = 4.5, max = t_admin)
cenz_2 <- runif(n2, min = 4.5, max = t_admin)
cenz_3 <- runif(n3, min = 4.5, max = t_admin)

dat <- bind_rows(
  tibble(arm = "SOC", time_evt = t_evt_1, time_cenz = cenz_1),
  tibble(arm = "SLNB", time_evt = t_evt_2, time_cenz = cenz_2),
  tibble(arm = "ALND", time_evt = t_evt_3, time_cenz = cenz_3)
) |>
  mutate(
    time   = pmin(time_evt, time_cenz, t_admin),
    status = as.integer(time_evt <= pmin(time_cenz, t_admin))
  ) |>
  select(arm, time, status)

# [3] Fit KM
fit <- survfit(Surv(time, status) ~ arm, data = dat)
```

```r
# [4] Tidy KM for ggplot
km_df <- do.call(rbind, lapply(seq_along(fit$strata), function(i) {
  k <- cumsum(fit$strata)
  lo <- if(i == 1) 1 else k[i - 1] + 1
  hi <- k[i]
  idx <- lo:hi

  tibble(
    arm   = sub("^arm=", "", names(fit$strata)[i]),
    time  = fit$time[idx],
    surv  = fit$surv[idx],
    n_cen = fit$n.censor[idx]
  )
}))

cen_pts <- km_df |> filter(n_cen > 0)

# [5] Risk table - Updated to Monthly (1 to 12)
monthly_times <- seq(1, 12, by = 1)
risk_summary <- summary(fit, times = monthly_times)
risk_df <- tibble(
  time   = risk_summary$time,
  n.risk = risk_summary$n.risk,
  arm    = sub("^arm=", "", risk_summary$strata)
)

# [6] Plot Configuration
# Added orange for SOC; updated linetypes
col_map <- c("SOC" = "brown", "SLNB" = "gold", "ALND" = "darkgreen")
lt_map  <- c("SOC" = "solid", "SLNB" = "solid", "ALND" = "solid")

km_plot <- ggplot(km_df, aes(x = time, y = surv, color = arm, linetype = arm)) +
  geom_step(linewidth = 1.1) +
  geom_point(data = cen_pts, aes(x = time, y = surv, color = arm),
             shape = 124, size = 4) +
  scale_color_manual(values = col_map, name = "Arm") +
  scale_linetype_manual(values = lt_map, name = "Arm") +
  # X-axis now increments by 1
  scale_x_continuous(limits = c(0, 12), breaks = seq(0, 12, 1), expand = c(0.01, 0)) +
  scale_y_continuous(limits = c(0, 1), breaks = seq(0, 1, 0.25), expand = c(0, 0)) +
  labs(
    title = "Time to Deterioration (TtD) in QoL Score",
    subtitle = "Analysis across three oncology treatment arms",
    x = "Time (months)",
    y = "Probability of No Deterioration"
  ) +
  theme_minimal(base_size = 12) +
  theme(
    legend.position = "bottom",
    legend.background = element_rect(fill = "white", color = "grey80"),
    plot.margin = margin(5, 10, 0, 10)
  )
```

```r
risk_table <- ggplot(risk_df, aes(x = time, y = arm, label = n.risk, color = arm)) +
  geom_text(size = 3.2, fontface = "bold") +
  scale_color_manual(values = col_map, guide = "none") +
  scale_x_continuous(limits = c(0, 12), breaks = seq(0, 12, 1), expand = c(0.01, 0)) +
  labs(x = NULL, y = NULL) +
  theme_minimal(base_size = 10) +
  theme(
    panel.grid = element_blank(),
    axis.text.y = element_text(face = "bold", color = "black"),
    axis.text.x = element_blank(),
    plot.margin = margin(0, 10, 5, 10)
  )

# Combine
Figure_C23 <- plot_grid(
  km_plot,
  risk_table,
  ncol = 1,
  align = "v",
  rel_heights = c(3.5, 1.2)
)

Figure_C23

#[7]Save (Optional)
ggsave("Figure_C23.png", Figure_C23, width = 7.2, height = 7.2, dpi = 300)
ggsave("Figure_C23.pdf", Figure_C23, width=7.2, height=7.2, device = grDevices::cairo_pdf)
```

**Citation:** Soltanifar, M., Portuguese, A.J., Jeon, Y., Gauthier, J., & Lee, C.H. (2026). A North American Collaborative Atlas of Oncology Data Visualization with R Statistical Software. medRxiv preprint. https://doi.org/xx.xxxxx/xxxx.xx.xx.xxxxxxxx, ©2026 Soltanifar, M. et al. (CC BY 4.0/MIT)

**Figure 5.3.24.**

```r
# [1] Packages
library(survival)
library(rms)
library(ggplot2)
set.seed(2401)

#[2] Simulation
#[2-1] Parameters
n <- 600
x <- rnorm(n, mean = 0, sd = 1)
z <- rbinom(n, 1, 0.45)
eta <- 0.35 * x - 0.55 * pmax(x - 0.3, 0)^2 + 0.45 * pmax(-x - 0.8, 0)^2 + 0.25 * z
base_haz <- 0.08
t_event  <- rexp(n, rate = base_haz * exp(eta))
t_cens   <- rexp(n, rate = 0.05)
time     <- pmin(t_event, t_cens)
status   <- as.integer(t_event <= t_cens)
dat      <- data.frame(time = time, status = status, x = x, z = z)

#  [2-2] Model Fitting
knots_x <- as.numeric(quantile(dat$x, probs = c(0.05, 0.25, 0.50, 0.75, 0.95)))
dd <- datadist(dat); options(datadist = "dd")

fit <- cph(Surv(time, status) ~ rcs(x, knots_x) + z,
           data = dat, x = TRUE, y = TRUE, surv = TRUE)

#  [2-3] Extract Predictions
# ref.zero = TRUE sets HR=1 at the reference (median)
pred_data <- Predict(fit, x, ref.zero = TRUE, fun = exp)
pred_df   <- as.data.frame(pred_data)

# [3] Plot
Figure_C24 <- ggplot(pred_df, aes(x = x, y = yhat)) +
  scale_x_continuous(limits = c(-2.5, 2.5), breaks = seq(-2.5, 2.5, 0.5))+
  scale_y_continuous(limits = c(0, 3), breaks = seq(0, 3, 0.25))+
    # Reference line (HR = 1)
  geom_hline(yintercept = 1, linetype = "dotted", color = "grey40", linewidth = 0.8) +
  # Vertical lines for knots
  geom_vline(xintercept = knots_x, linetype = "dashed", color = "grey80", alpha = 0.7) +
  # Colored Confidence Interval Ribbon
  geom_ribbon(aes(ymin = lower, ymax = upper), fill = "steelblue", alpha = 0.2) +
  # Main RCS Line
  geom_line(color = "black", linewidth = 1) +
  # Rug plot for density
  geom_rug(data = dat, aes(x = x), inherit.aes = FALSE, sides = "b", alpha = 0.5) +
  # Labels and Scales
  labs(
    x = "Continuous Marker (x)",
    y = "Hazard Ratio (HR) vs Reference",
    title = "Restricted Cubic Spline(RCS) Effect on Hazard Ratio",
    caption = paste("Knots at:", paste(round(knots_x, 2), collapse = ", "))
```

```r
  ) +
  theme_classic(base_size = 12) +
  theme(
    plot.title = element_text(face = "bold", hjust = 0.5),
    axis.title = element_text(face = "bold"),
    panel.grid.major = element_blank(),
    panel.grid.minor = element_blank()
  )

Figure_C24

#  [5] Save (Optional)

ggsave("Figure_C24.png", plot = Figure_C24, width = 8, height = 5, dpi = 600)
ggsave("Figure_C24.pdf", plot = Figure_C24, width = 8, height = 5, device = "pdf")
```

**Figure 5.3.25.**

```r
# [1] Packages
library(ggplot2)
library(dplyr)
library(ComplexUpset)
library(patchwork)
set.seed(25)

# [2] Simulations
n <- 420
dat <- tibble(
  id    = sprintf("P%03d", 1:n),
  arm   = sample(c("Chemotherapy", "Surgery"), size = n, replace = TRUE, prob = c(0.5, 0.5)),
  X3    = rbinom(n, 1, 0.55),
  X1    = rbinom(n, 1, plogis(-0.2 + 0.8 * X3)),
  X2    = rbinom(n, 1, plogis(-0.1 + 0.7 * X1)),
  X4    = rbinom(n, 1, plogis(-0.4 + 0.6 * X2))
) %>%
  mutate(
    X3    = X3 == 1,
    X1    = X1 == 1,
    X2    = X2 == 1,
    X4    = X4 == 1,
    arm   = factor(arm, levels = c("Chemotherapy", "Surgery"))
  )

sets <- c("X1", "X2", "X3", "X4")

# [3] UpSet plot
dat_plot <- dat %>% select(all_of(sets), arm)

Figure_C25 <- upset(
  dat_plot,
  sets,
  encode_sets = FALSE,
  base_annotations = list(
    'Intersection Size' = (
      intersection_size(
        counts  = FALSE,
        mapping = aes(fill = arm)
      ) +
        scale_fill_manual(
          values = c("Chemotherapy" = "#9ecae1", "Surgery" = "#3182bd")
        ) +
        # CHANGE 1: Set fill title to "Arm"
        labs(y = "Intersection Size", fill = "Arm")
    )
  ),
  set_sizes = (
    upset_set_size() +
      geom_text(
        aes(label = after_stat(count)),
```

```r
        stat  = "count",
        hjust = 1.1,
        color = "white",
        size  = 3
      )
  ),
  width_ratio = 0.25,
  min_size    = 5
) +
  theme_minimal(base_size = 12) +
  theme(
    # CHANGE 2: Move legend to the right
    legend.position  = "right",
    panel.grid.minor = element_blank(),
    axis.text.x      = element_text(angle = 90, vjust = 0.5, hjust = 1)
  ) +
  plot_annotation(
    title    = "UpSet plot of subgroups formed by intersections of binary covariates",
    subtitle = "Intersections (X1, X2, X3, X4); bars show intersection size by treatment arm"
  )

Figure_C25

# [4] Save (Optional)

ggsave("Figure_C25.png", plot = Figure_C25, width = 8, height = 8, dpi = 600)
ggsave("Figure_C25.pdf", plot = Figure_C25, width = 8, height = 8, device = "pdf")
```

**Figure 6.3.**

```
# [1] Packages
library(fishplot)
library(Hmisc)
library(plotrix)
library(png)

#[2] Simulation
# [2-1]. Define Data to match the "Sample 20260214" shape
timepoints <- c(-50, -25,  200, 423)

frac.table = matrix(
  c(100, 45, 00, 00,# Gray (Founding Clone)
    02, 00, 00, 00, # Orange (Subclone of Gray)
    02, 00, 02, 01,# Red (Subclone of Gray, dies at 0)
    98, 00, 95, 40),# Yellow (Subclone of Orange)
  ncol=length(timepoints))

parents = c(0,1,1,3)
my_colors <- c("#808080", "#FF6600", "#FF0000", "#FFD700")

# [2-2] Legend labels (matches your comments)
clone_labels <- c( "Founding clone","Subclone (of founding)",
  "Subclone (dies at 0)","Subclone (of orange)")

# X-axis tick spacing in DAYS
x_tick_by <- 25
x_ticks <- seq(
  from = floor(min(timepoints) / x_tick_by) * x_tick_by,
  to   = ceiling(max(timepoints) / x_tick_by) * x_tick_by,
  by   = x_tick_by)

# [2-3] Create the object (colors attached inside the fish object)
fish <- createFishObject(
  frac.table = frac.table,
  parents    = parents,
  timepoints = as.numeric(timepoints),
  col        = my_colors
)

# [2-4] Calculate layout geometry
fish <- layoutClones(fish)

# [3] Plot
#[3-1] Draw function (so screen + saved files match exactly)
draw_fishplot <- function() {

  par(mar = c(3, 3, 3, 3))
  par(xpd = NA)

  fishPlot(
    fish,
```

```r
    shape    = "bezier",
    border   = 0.5,
    bg.col   = c("#FDEBD0", "#F39C12", "#A04000"),
    pad.left = 0.15
  )

  # Add x-axis ticks (days)
  axis(
    side = 1,
    at = x_ticks,
    labels = x_ticks,
    tck = -0.02,
    cex.axis = 0.9
  )

  # Keep your existing annotations exactly
  abline(v = c(0, 423), col = "white", lwd = 2)
  text(x = 0,   y = 1.05, labels = "0",   cex = 1.2)
  text(x = 423, y = 1.05, labels = "423", cex = 1.2)
  title(main = "Sample 20260214", line = 2, cex.main = 1.6)

  # Axis labels
  mtext("Time (days)",     side = 1, line = 2.6, adj = 1)  # x-axis label (days)
  mtext("Clonal Fraction", side = 2, line = 2.1)  # y-axis label

  # Legend at bottom with explicit title
 legend(
    "bottom",
    x = -150, y =-5,
    horiz  = TRUE,
    bty    = "n",
    xpd    = NA,
    title  = "Clone Category",
    legend = clone_labels,
    fill   = my_colors,
    border = NA,
    cex    = 0.95
  )
}

# [3-2] Draw to current device (interactive)
draw_fishplot()
#[4] Save(Optional)
# [4-1] Save as PNG (600 dpi)
png("Figure_T01.png", width = 10.5, height = 7, units = "in", res = 600)
draw_fishplot()
dev.off()
# [4-2] Save as PDF (vector)
grDevices::cairo_pdf("Figure_T01.pdf", width = 10.5, height = 7)
draw_fishplot()
dev.off()
```

**Citation:** Soltanifar, M., Portuguese, A.J., Jeon, Y., Gauthier, J., & Lee, C.H. (2026). A North American Collaborative Atlas of Oncology Data Visualization with R Statistical Software. medRxiv preprint. https://doi.org/xx.xxxxx/xxxx.xx.xx.xxxxxxxx, ©2026 Soltanifar, M. et al. (CC BY 4.0/MIT)
